# Prioritization of putatively detrimental variants in euploid miscarriages

**DOI:** 10.1101/2021.01.02.20248961

**Authors:** Silvia Buonaiuto, Immacolata Di Biase, Valentina Aleotti, Amin Ravaei, Adriano De Marino, Gianluca Damaggio, Marco Chierici, Madhuri Pulijala, Palmira D’Ambrosio, Gabriella Esposito, Qasim Ayub, Cesare Furlanello, Pantaleo Greco, Antonio Capalbo, Michele Rubini, Sebastiano Di Biase, Vincenza Colonna

## Abstract

Miscarriage is the spontaneous termination of a pregnancy before 24 weeks of gestation. We studied the genome of euploid miscarried embryos from mothers in the range of healthy adult individuals to understand genetic susceptibility to miscarriage not caused by chromosomal aneuploidies. We developed GP, a pipeline that we used to prioritize 439 unique variants in 399 genes, including genes known to be associated with miscarriages. Among the prioritized genes we found *STAG2* coding for the cohesin complex subunit, for which inactivation in mouse is lethal, and *TLE4* a target of Notch and Wnt, physically interacting with a region on chromosome 9 associated to miscarriages.

## Introduction

Miscarriage, the spontaneous termination of a pregnancy before 24 weeks of gestation, occurs in 10-15% of all pregnancies^1–3^ and has both environmental and genetic causes^1^. Miscarriages are often the result of chromosomal aneuploidies of the gametes but they can also have non random genetic causes like small mutations (SNPs and indels), both de-novo or inherited from parents. Miscarriages are mostly studied using parental genetic information^4,5^and at a resolution that leaves the vast majority of the genome unexplored. Comparative genomic hybridization detects variants of several thousand base pairs^6–8^, while targeted resequencing resolves point mutations. Both are currently the most accurate methods for the genetic analysis of parental DNA of miscarriages but are not sensitive to small variants, or target only a few coding regions. Using a different approach, the only study so far that tests for genome-wide genetic association in a large cohort of miscarriages is also based on maternal information^9^. Depending on the mode of inheritance, the study of parental genome might be ineffective as there is uncertainty about which parts of the parental genomes are actually inherited by the embryo, and it provides no way to identify *de novo* mutations. Therefore, extending the analysis to fetal genomes is the necessary next step to fully understand the genetics of miscarriages.

DNA sequence information of miscarried fetuses has been already used to determine the genetic component of miscarriages^10,11^. Most studies adopt a family-based approach integrating pedigree and parental genomic data, often with focus on a reduced range of fetal phenotype^12–15^. Very often the focus is on candidates genes. Examples are the identification of a mutation in the X-linked gene *FOXP3* in siblings male miscarriages^16^, and the identification of a truncating *TCTN3* mutations in unrelated embryos^17^. A number of studies analyze instead on exome sequences with different approaches^18–22^. Among them, one study selects only variants transmitted to both sibling miscarriages^19^, others limit to autozygous variants^17,18^, while some concentrate on delivering accurate diagnosis^21^. All these studies consider number of cases in the order of the tens and in most cases are motivated by phenotypic information mostly deriving from ultrasound scans. Two other studies adopt a cohort-based approach analyzing up to thousands of embryonic genomes with a range of phenotypes^23,24^. One of them focuses on searching causative variants, demonstrating that exome sequencing effectively informs genetic diagnosis in about one third of the 102 cases considered^24^. The other one focuses on conserved genes in copy number variable (CNV) regions in 1810 cases to identify 275 genes, often in clusters, located in the CNVs and potentially implicated essential embryonic developmental processes^23^ Because the number of embryos they analyze is too small for genetic association analysis to be effective, all studies mentioned so far perform sequencing followed by variant annotation and prioritization. All investigate apparently euploid embryos and focus on rare variation, but they use different criteria to select the variants and never release code to fully reproduce the variant prioritization.

In this study, we analyzed whole-genome sequencing on euploid embryos from idiopathic spontaneous pregnancy losses (both first and recurrent) and developed GP, a pipeline to prioritize putatively causative variants in coding regions. GP performs filtering of high-quality genomic variants based on prediction of the functional effect of the variants and using a set of parameters that can be specified by the user. This first selection is completed by filters for technical artifacts (e.g. mapping errors, read depth) and for false positives through resampling in a control cohort. Our pipeline can incorporate prior information on candidate genes, but is also robust to the discovery of novel genes. We prioritize on average 49 variants per embryo with high and moderate impact in genes relevant for embryonic development and mitochondrial metabolism, some of which were previously identified for having a role in miscarriages. We demonstrate that variant prioritization can be effective also when dealing with a limited number of samples and develop an approach that can be applied to a larger-scale project. Results from this study can be used to inform molecular diagnosis of pregnancy loss.

## Results

To understand genetic susceptibility to miscarriage we studied the genome of forty-six spontaneously miscarried embryos. The embryos’ gestational age at pregnancy termination, calculated as the interval between the pregnancy termination date and the last menstruation date, ranges from 7.14 to 19.43 weeks (median is 10.3 weeks). Twenty-one embryos classifies as the product of recurrent miscarriages^25^. The mothers of the embryos are mostly of European origin (87%) and their median age at the date of collection was 36.7±5.9 years, with slightly significant higher age in recurrent cases compared to first ones (Figure **??**, Mann–Whitney p-value=0.02). Medical records of the mothers of the embryos report no major comorbidities. Folic acid was taken by 71% of the mothers with no difference between first and recurrent cases (Figure **??**, Chi-square p-value=0.96). Median body mass index and menarche age are comparable between first and recurrent cases, as well as comparable to a group of control women (Figure **??**). Altogether, from the available medical records, we suppose that the recruited mothers of the embryos were in the range of healthy adult individuals.

It is known from literature that about half of the miscarriages in the first trimester are due to large chromosomal aneuploidies, such as trisomies or deletions of large chromosomal chunks^26^. In this study we want to focus on cases in which the genome is euploid, therefore the forty-six embryos were screened for chromosomal aneuploidies prior to whole-genome sequencing. We find that 15 (32.6%) of samples were euploid while 56.6% of the embryos presented aneuploidies (Figure 2). The most common aneuploidy in our data set is the trisomy of chromosome 22 (26.9%), followed by trisomy of chromosome 16 (15.4%). In particular, a first round of detection of aneuploidies on chromosomes 13, 15, 16, 18, 21, 22, X, and Y through Short Tandem Repeats analysis discarded 45.7% of samples, and a subsequent analysis through comparative genomic hybridization and copy number variation detection form low-coverage sequencing discarded another 10.9% of the samples. Finally, a number of embryos (10.9%) dropped off the analysis due to low-quality DNA or maternal contamination.

**Figure 1.**
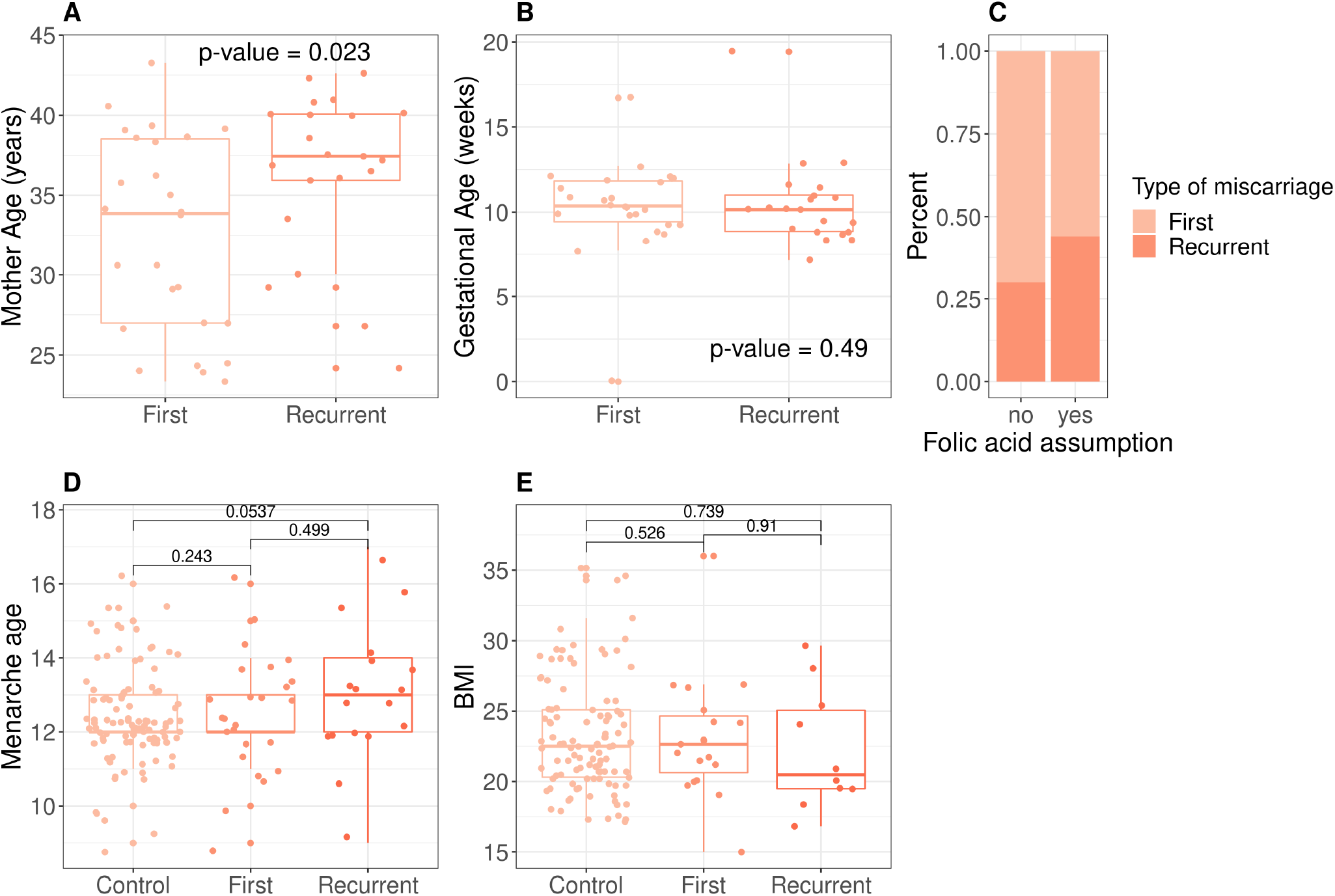
Features of the embryo’s mothers. **(A)** Median age of the mother at the event is 33.9±6.16 and 37.5± 5.22 for first and recurrent miscarriages, with no significant difference. **(B)** Gestational age at the time of the pregnancy termination range from 7.14 to 19.4 weeks with no significant difference between first and recurrent cases. **(C)** Folic acid intake. Range of values of menarche age **(D)** and Body Mass Index **(E)** in embryo’s mothers are not significantly different from a control set of mothers undergoing voluntary termination of pregnancy.

**Figure 2.**
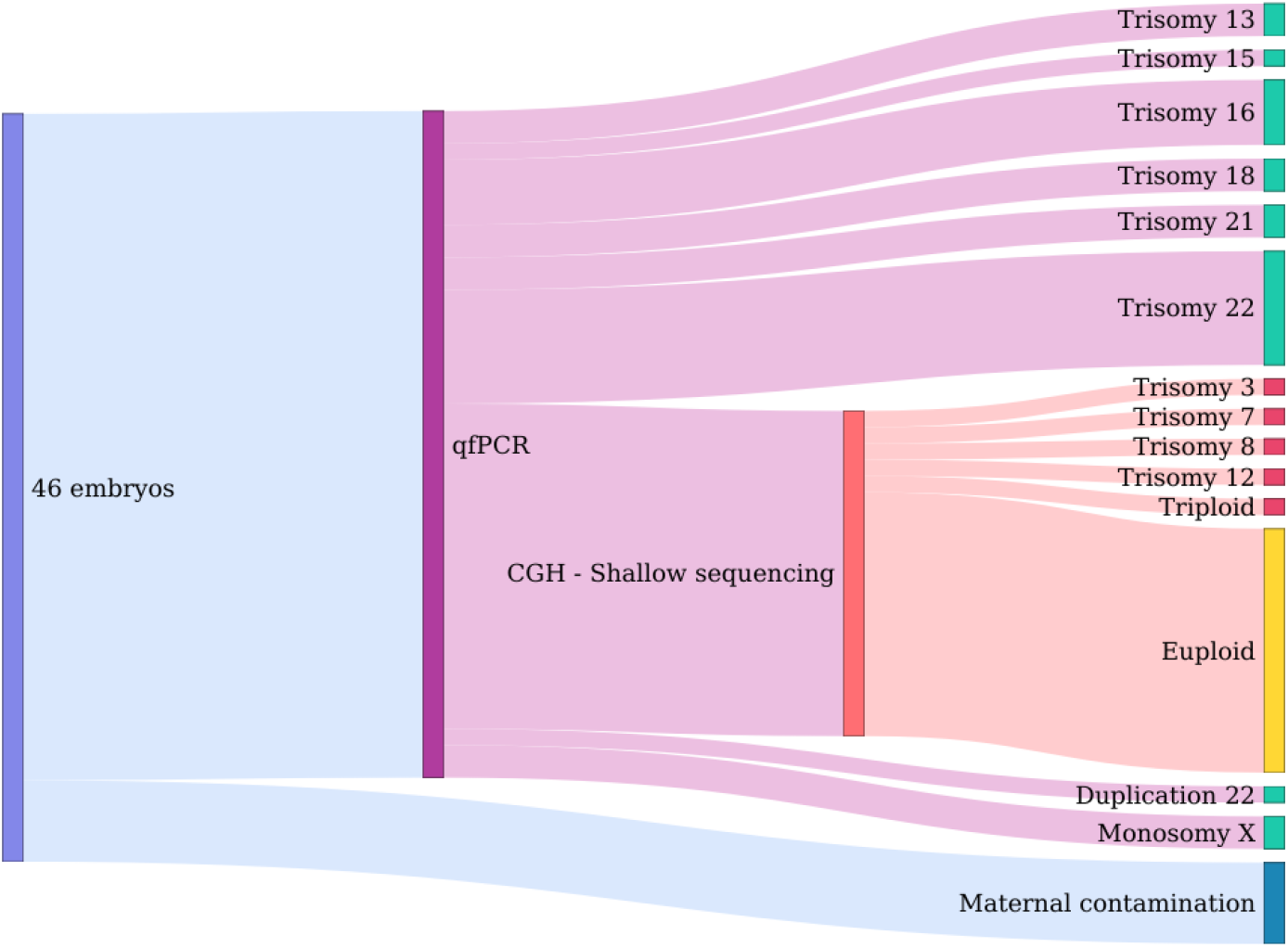
Outcome of the screening for aneuploidies in the embryos. Forty-six embryos were screened by quantitative PCR to determine aneuploidies of chromosomes 13, 15, 16, 18, 21, 22, X, and Y, as well as to determine maternal contamination. DNA of embryos with no anuploidies in these chromosomes were further analyzed by comparative hybridization and shallow sequencing. Overall we found aneuploidies in 56.6% of the embryos, the most common being the trisomy of chromosome 22. In yellow the fraction of euploid embryos.

Due to resources constrains, the whole genome of ten out of 15 euploid embryos was sequenced using Illumina short-reads at 30X coverage. In the set of embryos genomes, we identified 11M single-nucleotide polymorphisms (SNPs) and 2M small insertions or deletions (indels).

### Prioritization of genetic variants in coding genomic regions

Genomic variations was analyzed by prioritizing variants at embryo-level, in the hypothesis of genetic heterogeneity among causative variants and phenotypes and considering the limited sample size that would limit conclusions form aggregate analyses. We developed the GP pipeline to prioritize putatively damaging genetic variants from sequencing data. GP takes as input genomic variants information from cases and controls (including the per-individual allelic counts) in form of a vcf file and outputs a table of variants prioritized according to user-defined parameters. GP uses functional annotations of genomic variants, information from publicly available sequence data of presumably healthy individuals, and, if available, knowledge of genes involved in the trait under study. GP currently analyze coding regions and performs four filtering steps (Figure 3). The first filter (Filter I) retains variants based on: (i) an overall impact on the gene product classified as moderate or high^27^; (ii) a user-defined threshold of allele frequency in control populations; (iii) the combined property of being putatively damaging (quantified by the CADD score^28^) and located in genes intolerant to loss of function (determined by the pLI score^29^). In addition it is possible to incorporate one or more user-defined lists of genes relevant to the trait under study. Variants retained by Filter I (hits) are further filtered to control for false positives with Filters II and III. In particular Filter II removes variants in genes with too many hits, while Filter III determines the chance for genes to be selected in a control population based on criteria specified in Filter I. In practice, a number of control individuals are sampled a number of times and their genetic data filtered using Filter I to obtain a list of genes selected by chance (Figure 3C). Finally, Filter IV excludes private variants with read depth outside the range found in non-private ones.

**Figure 3.**
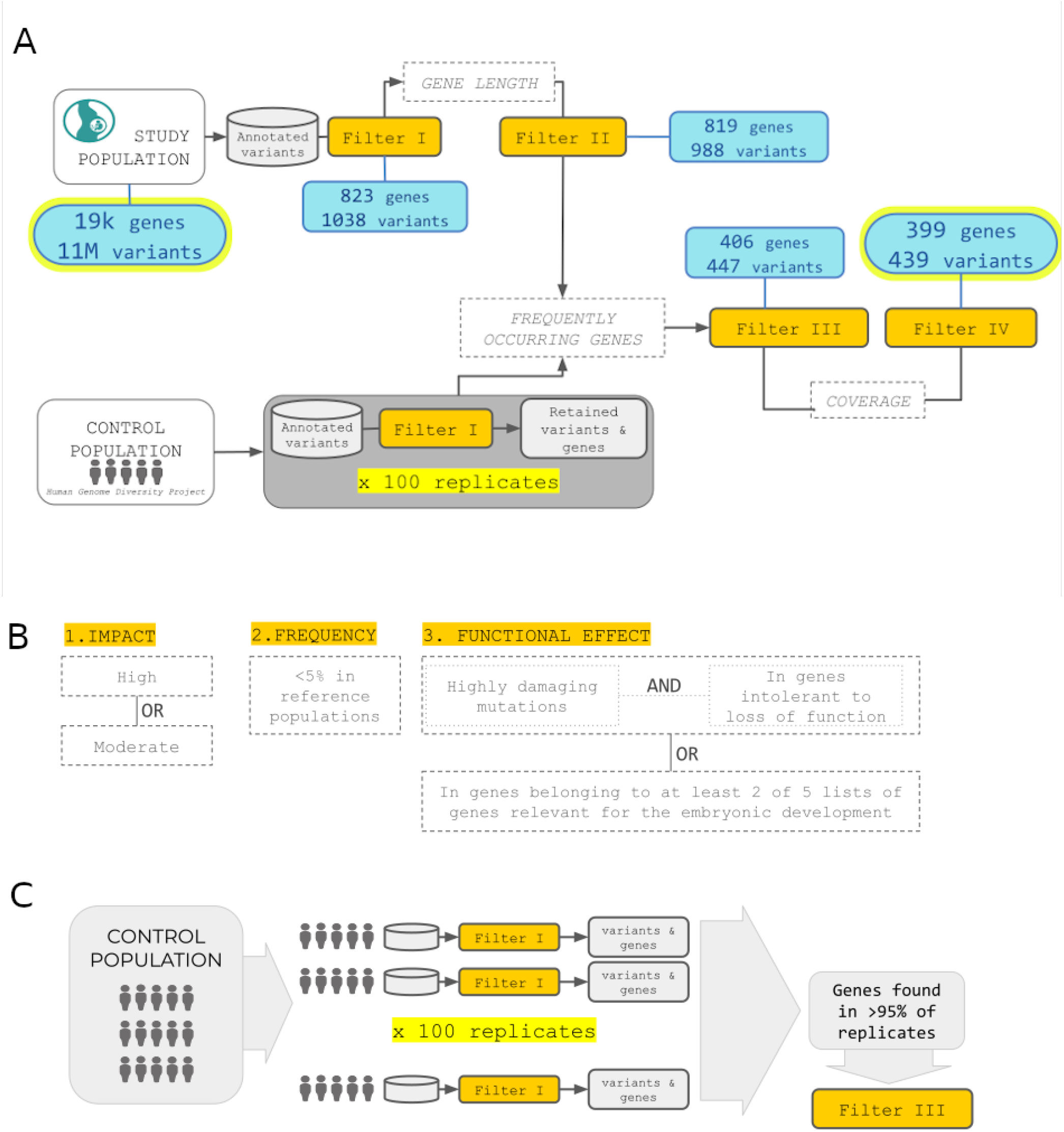
Overview of the pipeline for prioritization of the genetic variants. **(A)** GP takes as input genomic variants information from cases and controls and outputs a subset of variants prioritized according to user-defined parameters GP currently analyzes coding regions and performs four filtering steps. **(B)** Filter I retains variants based on three criteria: overall impact on the gene product moderate or high, allele frequency in control populations, the combined property of being putatively damaging (quantified bythe CADD score) and located in genes intolerant to loss of function (determined by the pLIscore). It is also possible to incorporate one or more user-defined lists of genes relevant to the trait under study. **(C)** Filter III determines the chance for genes to be selected in a control population based on criteria specified in Filter I. In practice, a number of control individuals are sampled a number of times and their genetic data filtered using Filter I to obtain a list of genes selected by chance.

We applied the GP pipeline to data from the high-coverage whole-genome sequences of genomic DNA of the embryos. For Filter I we set allele frequency <0.05% in the 1000 Genomes^30^ and gnomAD^29^ reference populations, while the functional effect of the variant within the gene context was taken into account in two ways: either selecting for putatively deleterious variants (CADD score >90th percentile) in genes highly intolerant to loss of function (pLI score >0.9), or selecting for variants in genes known to be involved in early embryonic development. In particular for this last option we included five lists of genes, namely genes involved in embryo development (Gene Ontology GO:0009790), genes lethal during embryonic stages^31^, essential for embryo development^31^, genes discovered through the Deciphering Developmental Disorders project^32^, and a manually curated list of candidate genes known to be involved in miscarriages. We requested the variant to satisfy one or both these criteria: (i) be in a gene present in at least two of the five lists or (ii) have CADD score above the 90th percentile and be in a gene with pLI>0.9. Overall, filter I retained 1,038 variants (hits) in embryos.

Filter II removed variants in genes with >5 hits, under the assumption that variants found in these genes are likely to be sequencing and alignment artifacts. With few exceptions, we observed that the number of hits per gene at the 99th percentile was five, even if there is no significant correlation between number of hits and gene length (Spearman r^2^=0.05 p-value=0.124), and that hits in genes with >5 hits are enriched for private variants (Figure 4A).

**Figure 4.**
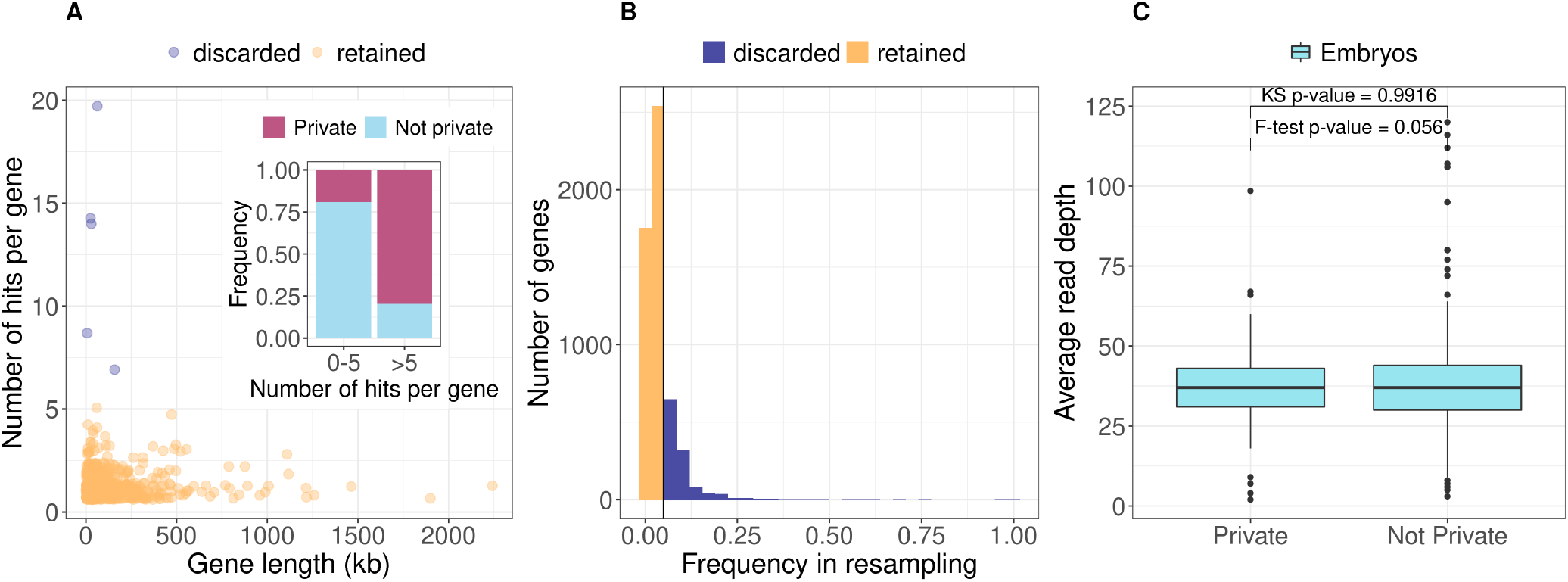
Features of the filtering steps. **(A)** Number of hits per gene after Filter II. The majority of genes have less than five hits and there is no significant correlation between number of hits and gene length (Spearman r=0.05 p-value=0.124). In the insert: the genes with >5 hits are enriched for private variants. (B) Frequency across 100 replicates of genes that pass Filter I in resampling form a control population. Most genes are retained *<*5% of times (yellow) therefore are retained if found in the embryos. 1,531 genes are instead retained in >5% of replicates (blue) and therefore discarded if found in the embryos, under the assumption that they can be filtered by chance in healthy controls. **(C)** Despite comparable read depth between private and non-private variants After Filter III to control for possible artifacts due to scanty coverage, we further filter to remove hits that are private and with read depth outside the range found in non-private ones.

For Filter III we used as control population 929 individuals from the Human Genome Diversity Project^33^ from which we resampled 100 times ten individuals after checking for population stratification (Figure S2). On each resampled set we performed Filter I analysis and recorded the genes that were retained. Overall 5,488 unique genes were retained in controls with different frequencies in samples across replicates (Figure 4B). When considering the 95th percentile, 1,531 genes are found >5% of times across replicates, therefore hits within these genes were removed by Filter III.

Filter II and III, retained 447 hits of which 21% are private with respect to 1000 Genomes and gnomAD data sets. Despite comparable read depth between private and non-privates variants (Figure 4C, KS test p-value=0.99, F-test p-value=0.06), to control for possible artifacts due to scanty coverage, we applied a further filter that removes hits which are private variants with read depth outside the range found in non-private ones.

#### Properties and biological significance of the prioritized variants and genes

After all filters, GP prioritize 439 unique variants in 399 genes that code for 980 transcripts (Supplementary Table 2). Almost all the prioritized genes (n=378) have an OMIM accession number and 18.8% of them were not in the lists of candidate genes used by GP as input during the prioritization, demonstrating that GP is robust to detection of genes never investigated before in relationship to the phenotype under study.

Nine genes are involved in the pathway of mitochondrial translation (Reactome identifier R-HSA-5368287) and this number represent a significant 4.9 fold enrichment over random expectations (Supplementary Table 3, p-value=1.45E-04, FDR=0.03). Similarly, we observe over overrepresentation of genes involved in cell cycle checkpoints (R-HSA-69620) and signaling by Rho GTPases (R-HSA-194315). With reference to the cellular compartments where the gene product are expressed, we observe a 7.7 fold significant enrichment (p-value 7.82E-04, FDR=0.04) of protein expressed in the mitotic spindle pole or in associated complexes (Supplementary Table 4), among which the product of *STAG2* for which we observe an high-impact mutation in one embryo from this study. Finally, seven genes (*BHLHE40,DBN1, FOXA1, HSPD1, PLXNA3, SLC35A2, SRF*) were previously identified as essential genes in copy-number variable regions from the analysis of hundreds of miscarried fetuses^23^

In the embryos 4.1% of the prioritized variants are stop gains/loss, frameshift indels, and variants that disrupt splicing sites, all classified as having high impact on the gene products, while missense mutations prevail among the variant with moderate effect (Figure 5A). Averages per embryos are 48.9±8.0 genomic variants in 47.8±7.7 genes coding for 113.5±24.6 transcripts (Figure 5B). In almost all prioritized genes, GP retains only one variant per embryo, with few exception (five cases with two e and one with three variants per gene), as shown in Figure 5B, where the allele dosage and impact are also shown.

**Figure 5.**
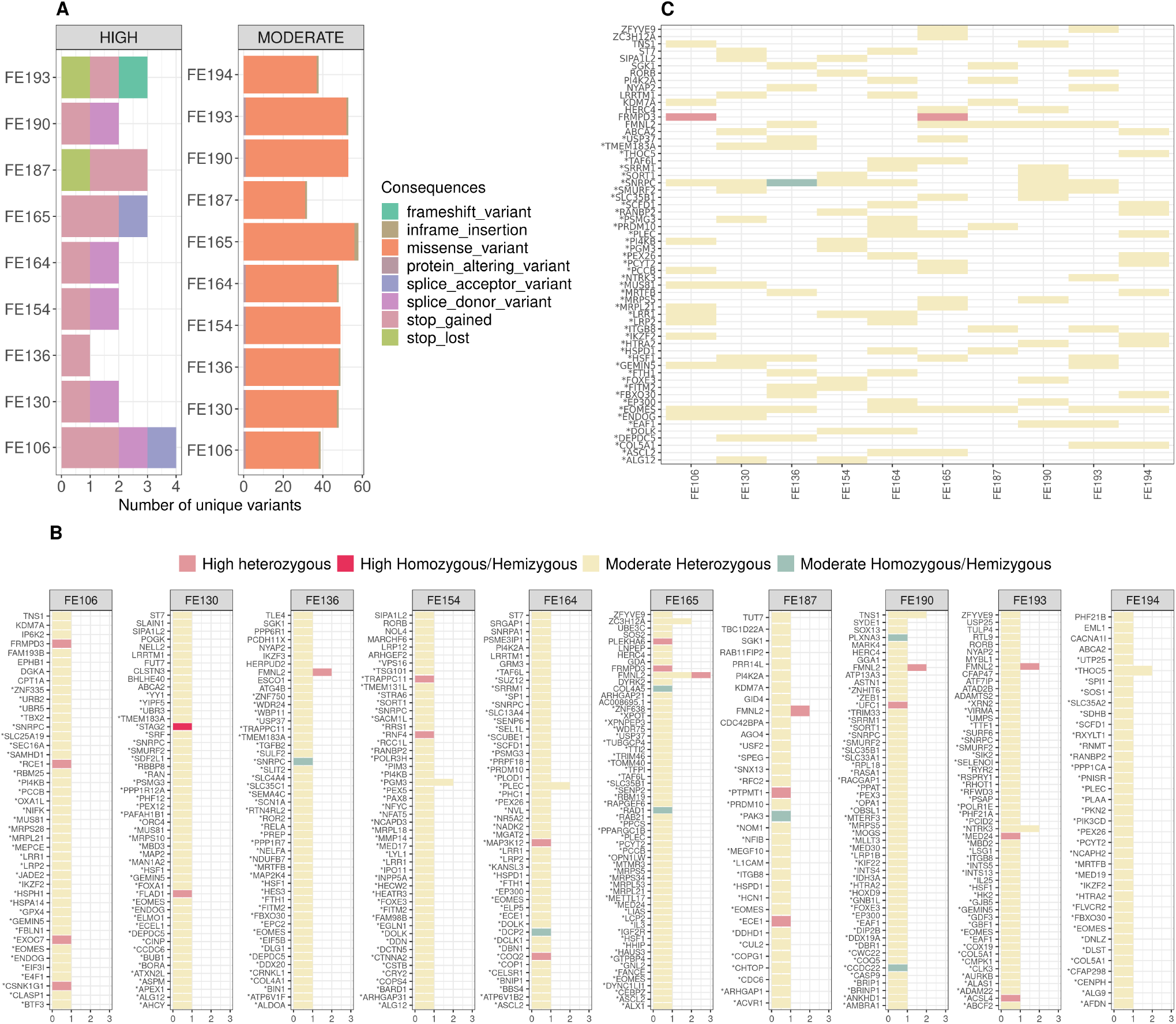
Results of the prioritization pipeline. **(A)** Number of variants per embryo stratified by impact. Overall 4.1% of the prioritized variants are classified as having high impact on the gene products. **(B)** Results per embryo. On the y-axis the prioritized genes while on the x-axis the number of mutations per gene. Colors indicate the allele count and the class of severity. **(C)** Selection of prioritized variants/genes shared by embryos.

#### Mutations in STAG2, FLAD1, TLE4, FRMPD3, and FMNL2 in the embryos

Among the selected mutations through variant prioritization we would like to highlight a few cases for their relevance. The male embryo FE130 carries two high-impact mutations in single copy. The first is a one extremely rare T>G transversion (rs913664484, G frequency is 4.7e-05 in 42.7k individuals from gnomeAD) at the 5’ end of the first intron of the Stromal antigen 2 (*STAG2*) gene. The mutation disrupts a splicing site, therefore having an high impact on the gene product. *STAG2* is located on the X chromosome and its inactivation is the cause of severe congenital and developmental defects in embryos and infants^32,34–36^ as well as chromosomal aneuploidies in several types of human cancers^37^. Interestingly, only mildly-deleterious mutations have been found in alive human males, while females can carry highly deleterious mutations in heterozygosis^35^. *STAG2* codes for the cohesin subunit SA-2^38^. Cohesins are ring-shaped protein complexes that bring into close proximity two different DNA molecules or two distant parts of the same DNA molecule and are responsible for the cohesion of sister chromatids^39^. In mouse, inactivation *Stag2* causes early embryo lethality^40^.

The second high-impact mutation of FE130 is a stop gain in the Flavin Adenine Dinucleotide Synthetase 1 (*FLAD1*) gene that is expressed in the mitochondrial DNA where it catalyzes the adenylation of flavin mononucleotide (FMN) to form flavin adenine dinucleotide (FAD) coenzyme^41^. The FAD synthase is an essential protein as the products of its activity, the flavocoenzymes play a vital role in many metabolic processes and in fact FAD synthase deficiencies (OMIM 255100) associated with homozygous severe mutations cause death in the first months of life^42^. In FE130 the stop mutation p.Q159* affects one of the five isoforms (Uniprot identifier Q8NFF5-5) at the second last residue, therefore we can speculate that it might not seriously compromise the function of the protein.

The embryo FE136 carries an heterozygous missense mutation (rs41307447) in the Transducin-like enhancer protein 4 gene (*TLE4*, synonym *GRG-4*) that causes a substitution of a polar amino acid with another polar amino acid (Ser>Tre) in the seventh exon of the gene, corresponding to a low complexity domain of the protein. The rs41307447 polymorphism is tolerated (SIFT score 0.18) and supposed to be benign (PolyPhen score 0.003), nevertheless the *TLE4* gene is classified as highly intolerant to loss of function (pLI score 0.999) and the CADD score associated to rs41307447 is in the 99.8th percentile. *TLE4* is a trascriptional repressor of the Groucho-family expressed in the embryonic stem cells where it represses naive pluripotency gene^43^ and it is a direct transcriptional target of Notch^44^. *TLE4* is also expressed in the extravillous trophoblasts^45^ where it is part of the Wnt signaling pathway that promotes implantation, trophoblast invasion, and endometrial function^46^. Finally, a study in a cohort of 750 women finds significant association between the A allele of rs7859844 on chromosome 9 and recurrent miscarriages, further showing that rs7859844 physically interact with *TLE4*^9^. In our study among all embryos only FE106 carries the intergenic variant rs7859844.

Among prioritized variants shared by more than one embryo, the male FE165 and female FE106 embryos share a stop gain mutation (p.Q1758*) in the X-linked FERM and PDZ domain containing 3 (*FRMPD3*) gene, which is highly intolerant to loss of function (pLI = 0.91). The mutation falls at the protein C-terminal in a polyQ stretch (27 residues). While little is known in humans about this gene, a study in lion head goose finds significant association between high expression of *FRMPD3* and low production of eggs^47^.

Five embryos, among which the carrier of the missense mutation in *TLE4*, share one copy of an haplotype composed of two T alleles 4bp apart causing stop-gain (rs750755379) and missense (rs866373641) substitutions in the Formin-like protein 2 gene (*FMNL2*, Figure 5C). The two alleles exist at moderate-to-high frequency in human populations (Figure 6A) and are in perfect linkage disequilibrium (r^2^=1) in the embryos. In addition to the two mutations described above, the embryo FE165 has a deleterious and probably damaging missense mutation in phase with the two others (rs189416564, SIFT=0, PolyPhen =0.969). *FMNL2* codes for a formin-related protein expressed in multiple human tissues and in particular in gastrointestinal and mammary epithelia, lymphatic tissues, placenta, and in the reproductive tract^48^. In the fetus *FMNL2* is expressed in the cytoplasm of brain, spinal cord, and rectum^49^. *FMNL2* is an elongation factor of actin filaments that drives cell migration by increasing the efficiency of lamellipodia protrusion^50,51^, and its overexpression is associated with cancer^52^. The stop-gain mutation we find in the five embryos is located in the first domain of the protein, a Rho GTPase-binding/formin homology 3 (GBD/FH3) domain involved in subcellular localization and regulation of activation (Figure 6B). The stop codon produces a truncated protein that lacks the Formin Homology-2 (FH2) domain, which directly binds to the actin filament catalyzing its nucleation and elongation.

**Figure 6.**
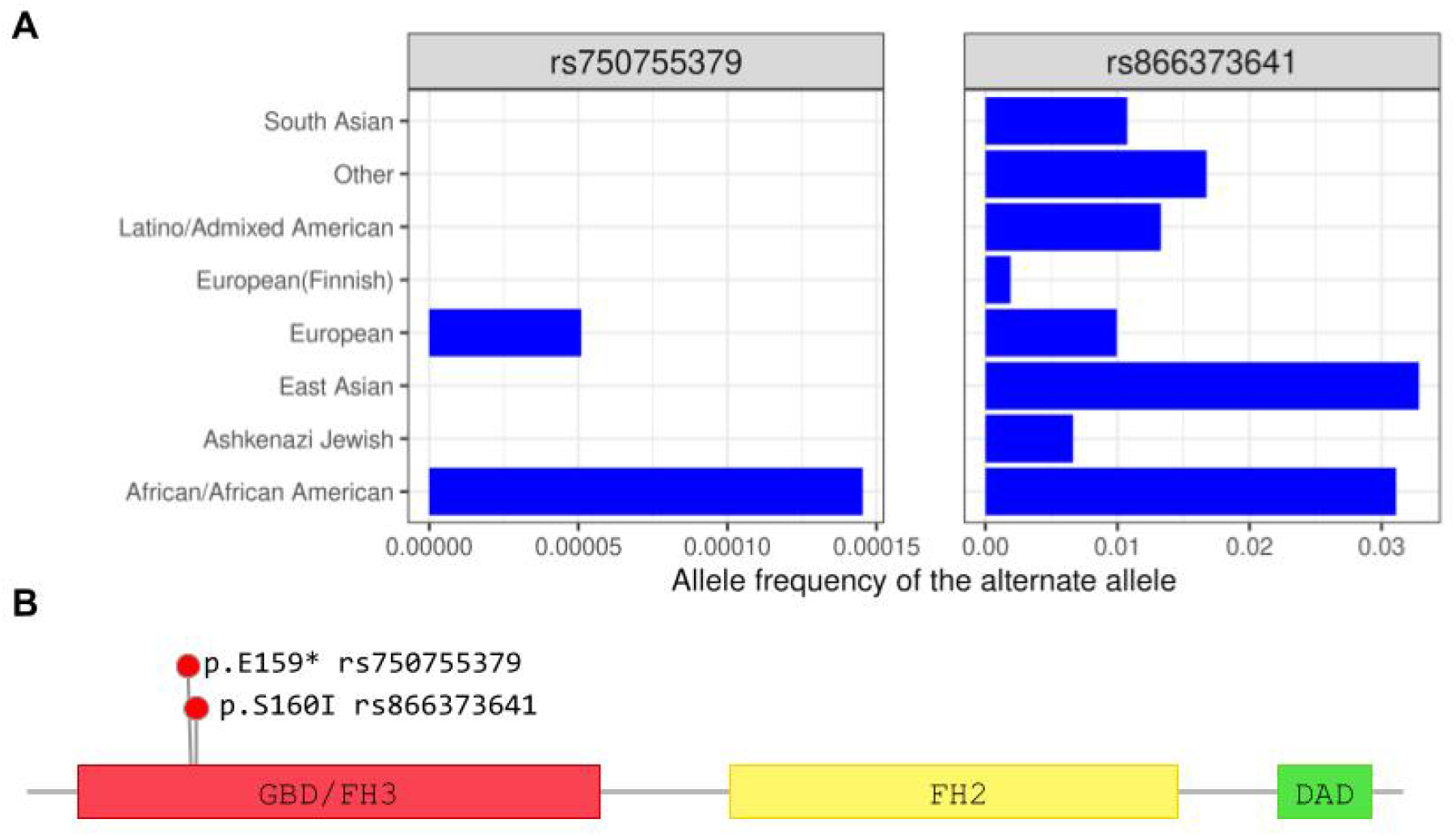
Two-bases haplotype in *FMNL2* prioritized in five embryos. **(A)** Alelle frequencies from the gnomAD database of the aternate alleles at rs750755379 and rs866373641. The two alleles exist at low frequency in human populations. **(B)** Position in the protein of rs750755379 (stop gain) and rs866373641 (missense). The stop-gained mutation is located in the Rho GTPase-binding/formin homology 3 (GBD/FH3) domain involved in subcellular localization and regulation of activation. The resulting truncated protein lacks the Formin Homology-2 (FH2) domain, which directly binds to the actin filament catalyzing its nucleation and elongation.

## Discussion

Miscarriages are frequent events with a complex aetiology whose genetic components have not been completely understood. We developed a scalable pipeline that investigates small genetic variation which has rarely been considered in the context of miscarriages. We use our pipeline to analyze coding regions of the genome of ten miscarried euploid embryos to prioritize putatively detrimental variants in genes that are relevant for embryonic development.

Our pipeline prioritized 439 putatively causative single nucleotide polymorphisms among 11M variants discovered in the ten embryos. Through systematic investigation of all coding regions GP selected about 47 genes per embryo and by manual curation of the selected genes we highlight a few cases. Among them, we find three examples relevant to embryonic development. An hemizygous splice site mutation in one male embryo on *STAG2*, known in literature for its role in congenital and developmental disorders as well as in cancer^32,34–37^. A missense mutation in *TLE4*, a gene that interacts with the genomic region on chromosome 9 genetically associated with miscarriages in a genome-wide study on mothers^9^. *TLE4* appears to be a key gene in embryonic development, as it is expressed in both embryonic and extraembryonic tissues where it participates in the Wnt and Notch signalling pathways^43–45^. Finally, a 4-bp haplotype in five embryos, containing a stop gain and a missense mutations in *FMNL2*, a gene involved in cell motility with a major role in driving cell migration^50,51^. The stop gain mutation truncates the protein well before the main functional domain of *FMNL2*, i.e. the domain that binds the actin filaments, therefore causing a complete loss of function of the protein product. In this study we focus on single nucleotide variants. GP combines functional information on variants and genes with population genomics and literature information to sift millions of variants in search for the relevant ones. This approach closes a gap as genetic analyses of miscarriages mostly focused on detecting chromosomal aneuploidies and large chromosomal aberrations (which explain less than half of the cases) leaving unexplored small size genetic variants, the most abundant type of genomic variation. To some extent small genetic variants have been considered in a number of cases that performed target resequencing of candidate genes^16,17^, a valid but still not systematic approach because it does not fully exploit genomic information. GP filter both variants in genes known from literature to be associated with miscarriages, and variants never described before in this context but potentially highly damaging and in genes intolerant to loss of function. As a result, our approach is robust to both discovery of novel association and investigation of genes with known association to miscarriages, overcoming the major limitation of candidate genes studies.

Variant prioritization is done at an individual level. While we expect that the same gene might be the cause of multiple miscarriages, given our limited sample size we do not expect that the same exact mutation to cause the gene’s loss of function. Therefore, by filtering at the individual level GP accounts for inter-individual variation, i.e. the larger fraction of genomic variability, as well as for the high chance of occurrence of *de novo* mutations. Nevertheless, in five embryos GP selected the exact same combination of two linked alelles in *FMNL2*, showing that while it is individual-based GP is still capable of finding variants shared by more than one case.

Our pipeline is reproducible and easy to scale to larger studies and different phenotypes. To improve robustness, it includes a control population to filter out genes that can be prioritized by chance. GP is suitable for cases where it is not possible to rely on an adequate number of samples to perform association analysis. The future integration of genomic information on parents (not available in this collection) will allow us to infer inheritance mechanisms and distinguish between *de novo* and recessive mutations, with implications for clinical applications in the case of causative recessive mutations in the parents. Collecting genomic information from larger families, with several miscarriages/live births from the same couple will also further increase the strength of mendelian segregation analysis and the true discovery rate.

In conclusion, this exploratory study demonstrates that filtering and prioritizing is effective in identifying genomic variants putatively responsible for miscarriages and provides indications and tools for developing a larger study. Compared to previous similar studies our work focuses on a systematic exploration of the genome that combines previous knowledge with hypothesis-free prioritization, making it robust not only to the discovery of mutations in genes known to be associated with miscarriage, but also in the identification of novel genes. Our findings have wide clinical implications. While only providing a proof of concept study, have already produced information about genes that can be used to test genetic predisposition to miscarriages in parents that are planning to conceive or particularly for recurrent miscarriage patients. In a wider context, the results of this study might be relevant for genetic counseling and risk management in miscarriages. Future development will include the extension of the analysis to non-coding regions and to structural variants, as well as the enrollment of trios to fully exploit parental information.

## Methods

### Embryo data and samples collection

The study protocol was examined by the Comitato Etico di Area Vasta Emilia Centro (CE-AVEC) of the Azienda Ospedaliero - Universitaria di Bologna Policlinico S. Orsola-Malpighi. The committee gave the ethical approval of the study (reference CE/FE 170475). All participants provided written informed consents before entering the study. Cases were recruited at the Unit of Obstetrics and Gynecology of the Sant’Anna University Hospital in Ferrara, Italy, from 2017 to 2018. The inclusion criteria were: age between 18 and 42 years and gestational age up to 12 weeks. Exclusion criterion was any clinical condition that could prevent full-term pregnancies. Known causes of pregnancy losses were excluded by standard diagnostic protocol including hysteroscopy, laparoscopy, ultrasound, karyotype analysis, detection of immunological risk factors (anticardiolipin, lupus anticoagulant, antinuclear antibodies) and hormonal status (gonadotrophins, FSH, LH, prolactin, thyroid hormones, thyroperoxidase) before inclusion in the study. Gestational weeks were calculated from the last menstrual period. Demographic, antropometric and clinical data of cases, including obstetric history, family history of malformations, and periconceptional supplementation with folic acid, were anonymized and linked to biological samples by coding.

### DNA preparation and sequencing

Retained product of conception was removed from uterus using a suction curette, and chorionic villi (CV) were carefully dissected from decidual tissue. We used dry homogenization after exploring a range of possibilities (Figure S1A). Genomic DNA was extracted from CV samples using QIAamp DNA Mini Kit (Ref: 51304, Qiagen) according to manufacturer’s protocol. This kit was chosen after considering the yield of two types of resin and one membrane (Figure S1B). DNA was titrated using Qubit 2.0 Fluorometer (Life Technologies). Whole-genome sequencing of the genomic DNA extracted from chorionic villi was done through a service provider (Macrogen). In particular, libraries for sequencing were prepared using the Illumina TruSeq DNA PCR-free Library (insert size 350bp) and samples were sequenced at 30X mapped (110Gb) 150bp PE on HiSeqX.

### Detection of chromosomal aneuploidies in embryos

A rapid screening of sex and anuploidies for chromosomes 13, 15, 16, 18, 21, 22 and X was carried out on geomic DNA extracted from the chorionic villi performing five multiplex Quantitative Fluorescent PCR (QFPCR) assays. QFPCR assays were performed in a total volume of 25l containing 40–100ng of genomic DNA, 10mM dNTP (Roche), 6-30 pmol final concentration of each primer, 1Fast taq polymerase buffer (15mmol/l MgCl 2) (Roche), and 2.5 U of Fasta taq polymerase (Roche). QFPCR conditions were as follows: denaturation at 95C for 10 min followed by 10 cycles consisting of melting at 95C for 1 min, annealing at 65C (−1C / cycle) for 1 minutes, and then extension at 72C for 40 seconds, then for 23 cycles at 95C for 1 min, 55C for 1 min, and 72C for 1 min. Final extension was for 10min at 72C and at 60C for 60 min. Fluorescence-labelled QFPCR products were electrophoresed in an CEQ 8000 Backman by combining 40 l of Hi-Di Formamide and 0.5 l of DNA size standard 400 (Backman); QFPCR products were visualized and quantified as peak areas of each respective repeat lengths. In normal heterozygous subjects, the QFPCR product of each STR should show two peaks with similar fluorescent activities and thus a ratio of peak areas close to 1:1 (ranging from 0.8 to 1.4:1). A trisomy is suspected when the ratio is above or below this range (peak area ratios 0.6 and 1.8, trisomic diallelic pattern), otherwise there are three alleles of equal peak area with a ratio of 1:1:1 (trisomic triallelic). The presence of trisomic triallelic or diallelic patterns for at least two different STRs on the same chromosome is considered as evidence of trisomy. Trisomic patterns observed for all chromosome-specific STRs are indicative of triploidy. Therefore accurate X chromosome dosage, to perform diagnosis of X monosomy, can be assessed by TAF9L marker allowing This gene has a high degree of sequence identity between chromosome 3 and chromosome X; primers on this gene amplify a 3 b.p. deletion generating a chromosome X specific product of 141 b.p. and a chromosome 3 specific product of 144 b.p. Maternal contamination was also checked by QFPCR comparing the alleles found in miscarriages with those found in maternal blood.

Comparative Genomic Hybridization was carried out using the Agilent SurePrint G3 Human CGH Microarray. Samples underwent DNA quantification and quality analysis prior to be labeled and hybridized on the microarray. Following hybridization samples were washed and the chip was scanned at 3 microns using the Agilent SureScan Microarray Scanner. The LogRatio from the arrays were segmented into regions of estimated equal copy number using both the method implemented in theAgilent CytoGenomics V3.0.4 software, and the Penalized least square implemented in the R package Copynumber (PLS,^53^). Classification as copy number of gains or losses (copy number variants) was done using as criteria at least five probes and Zscore *<*0.0016 (SD*4)^54^.

### Statistical and sequence analyses

Data cleaning, refining, and analysis (summary statistics, hypothesis testing) were performed using R^55^. Reads in the FASTQ file sequence data were aligned against the reference genome GRChg38.p12 using BWA^56^ and SAMTOOLS^57^. Variant calling was done using FREEBAYES^58^. The resulting VCF files were refined in further steps: VCFFILTER^59^ was used to filter variants for quality score>20, leaving only variants with estimated 99% probability of a polymorphic genotype call; VT^60^ was used to normalize variants and deconstruct multiallelic variants. Refined VCF files were compressed and indexed using samtools^57^. Variants were annotated for functional effects and allele frequency in other populations using Variant Effect Predictor^27^. Phasing was done using Beagle 5.1^61^ under standard parameters.

Principal component analysis was done with PLINK^62^ using 1,2M autosomal SNPs.

The GP pipeline for variant prioritization is written in Python and R and the code is publicly available (https://github.com/SilviaBuonaiuto/gpPipeline). The manually curated list of genes associated with miscarriages (recurrent and spontaneous) was obtained through a comprehensive search of the published literature. We considered seven studies highlighting the association of genes with miscarriages^4,5,9,19,20,63,64^. This compendium was further supplemented by genes from curated repositories such as Human Phenotype Ontology (HPO) [Robinson et al., 2008; URL: https://hpo.jax.org/app/browse/term/HP:0200067 last accessed: 1/12/2020 11:01:00 PM] and DisGeNET [Piñero et al., 2015; URL: http://www.disgenet.org/search last accessed: 1/12/2020 11:12:00 PM]. The search terms used were “recurrent miscarriages”, “abortion”, “spontaneous abortion”, and “recurrent spontaneous abortion”. After filtering by removing the duplicates, combining the gene sets obtained from the literature and databases yielded a total of 608 unique genes (Supplementary Table 1). Additional information of genes such as HGNC symbol, HGNC ID, Gene Stable ID, Chromosomal coordinates (GRChg38), karyotype band, transcript count, protein stable ID were extracted from Ensembl Biomart^65^.

Overrepresentation tests and protein classification were performed using the R package ReactomePA^66^.

## Data Availability

Data is available through collaboration

## Acknowledgements

We are thankful to all volunteers that were enrolled in the study, as well as all the medical personnel that contributed to the collection of samples. We are also thankful to Prof. Nicole Soranzo and Prof. Erik Garrison for help in the early stage of the project.

## Author contributions

Q.A., A.C., S.D.B., and V.C. conceived and designed the study. S.B., A.D.M., G.D.M., M.P., and V.C. wrote the code and performed the bioinformatics analyses. M.C. and C.F. provided support to bioinformatics analyses. V.A., A.R., I.D.B., P.D.A., and G.E. performed the experiments. P.G., and M.R. contributed to the clinical samples and collected clinical data. S.B. and V.C. wrote the manuscript. All authors critically reviewed the manuscript and approved the final version.

## Materials Correspondence

Correspondence should be addressed to

## Supplementary Information

### Supplementary Figures

**Figure S1.**
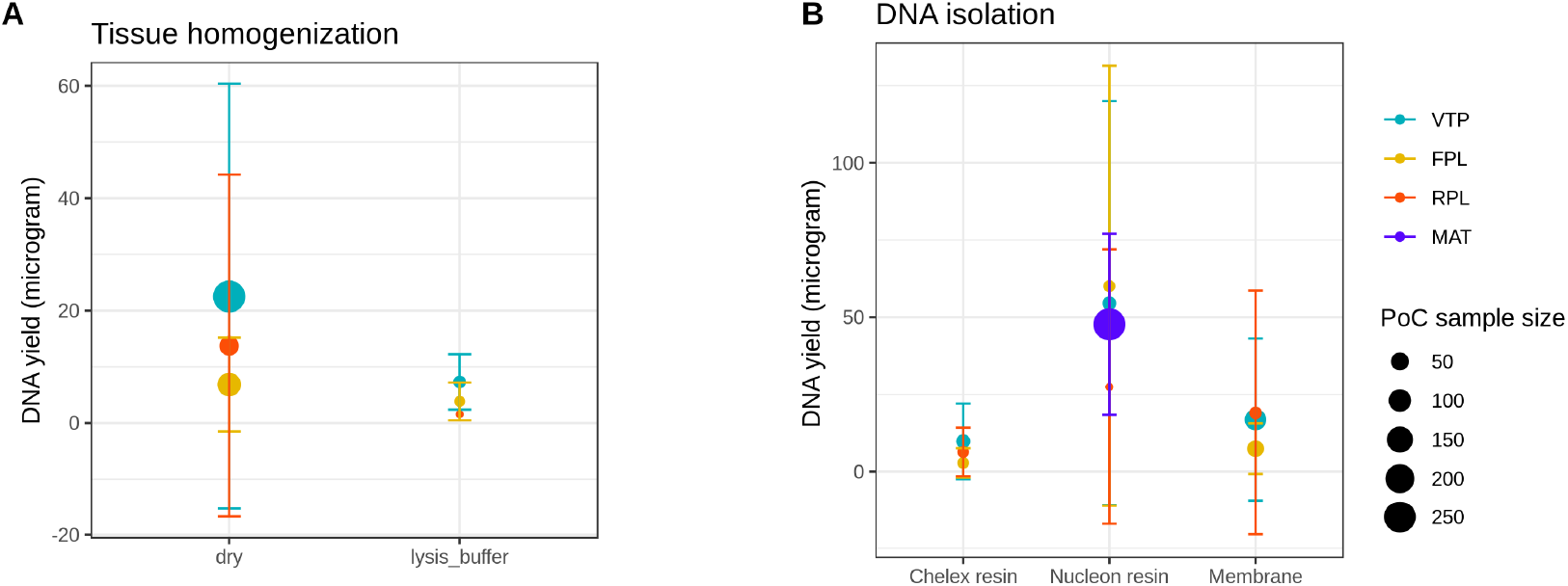
Optimization of tissue homogenization and DNA extraction. We do not observe significant difference between two methods of tissue homogenization (**A**), and three methods of DNA isolation (**b**) apart form a slightly higher range of yield for one type of resin. VTP: voluntary pregnancy termination; FPL: first pregnancy loss; RPL:recurrent pregnancy loss; MAT: maternal bllod; PoC: product of conception.

**Figure S2.**
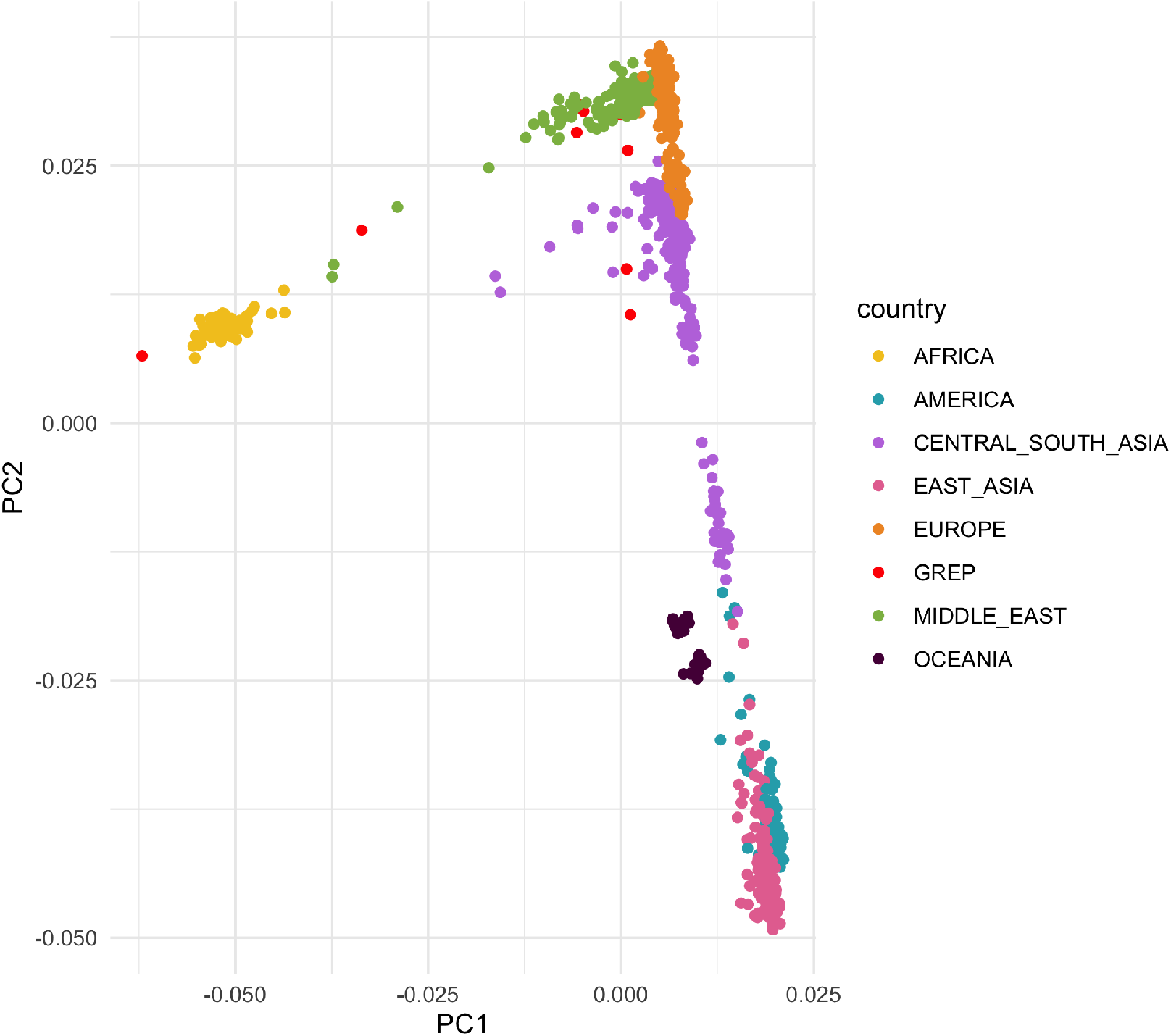
**Principal Component Analysis** of embryos from this study (GREP) and individuals in the HGDP panel used as controls

