## Supplementary Tables for "Prioritization of putatively detrimental variants in euploid miscarriages"

**Supplementary Table 1 – Manually curated list of genes involved in miscarriages – coordinates refer to GRC-hg38**

| HGNC symbol | HGNC ID | Gene stable ID | Chromosome / scaffold name | Gene start (bp) | Gene end (bp) | Karyotype band | Transcript count | Protein stable ID | Protein Coding | miRNA / RNA / Psuedogene |
|---|---|---|---|---|---|---|---|---|---|---|
| ABCB1 | HGNC:40 | ENSG00000085563 | 7 | 87503633 | 87713323 | q21.12 | 11 | ENSP00000265724 | Yes | |
| ACE | HGNC:2707 | ENSG00000159640 | 17 | 63477061 | 63498380 | q23.3 | 22 | ENSP00000290866 | Yes | |
| ACE2 | HGNC:13557 | ENSG00000130234 | X | 15561033 | 15602148 | p22.2 | 5 | ENSP00000252519 | Yes | |
| ACHE | HGNC:108 | ENSG00000087085 | 7 | 100889994 | 100896974 | q22.1 | 14 | ENSP00000403474 | Yes | |
| ACKR4 | HGNC:1611 | ENSG00000129048 | 3 | 132597270 | 132618967 | q22.1 | 2 | ENSP00000249887 | Yes | |
| ACP1 | HGNC:122 | ENSG00000143727 | 2 | 264140 | 278283 | p25.3 | 13 | ENSP00000272067 | Yes | |
| ACTA2 | HGNC:130 | ENSG00000107796 | 10 | 88935074 | 88991339 | q23.31 | 6 | ENSP00000224784 | Yes | |
| ACVR1 | HGNC:171 | ENSG00000115170 | 2 | 157736444 | 157875862 | q24.1 | 12 | ENSP00000263640 | Yes | |
| ADA | HGNC:186 | ENSG00000196839 | 20 | 44619522 | 44652233 | q13.12 | 9 | ENSP00000361965 | Yes | |
| ADAMTS1 | HGNC:217 | ENSG00000154734 | 21 | 26835755 | 26845409 | q21.3 | 6 | ENSP00000284984 | yes | |
| ADCYAP1 | HGNC:241 | ENSG00000141433 | 18 | 904871 | 912172 | p11.32 | 4 | ENSP00000411658 | yes | |
| ADH1B | HGNC:250 | ENSG00000196616 | 4 | 99304971 | 99352760 | q23 | 7 | ENSP00000306606 | yes | |
| ADIPOQ | HGNC:13633 | ENSG00000181092 | 3 | 186842704 | 186858463 | q27.3 | 2 | ENSP00000320709 | yes | |
| ADRA2B | HGNC:282 | ENSG00000274286 | 2 | 96112876 | 96116571 | q11.2 | 1 | ENSP00000480573 | yes | |
| ADRB3 | HGNC:288 | ENSG00000188778 | 8 | 37962990 | 37966599 | p11.23 | 2 | ENSP00000343782 | yes | |
| AGT | HGNC:333 | ENSG00000135744 | 1 | 230702523 | 230714122 | q42.2 | 1 | ENSP00000355627 | yes | |
| AGTR1 | HGNC:336 | ENSG00000144891 | 3 | 148697784 | 148743008 | q24 | 9 | ENSP00000419422 | yes | |
| AHR | HGNC:348 | ENSG00000106546 | 7 | 16916359 | 17346152 | p21.1 | 7 | ENSP00000242057 | yes | |
| AKT1 | HGNC:391 | ENSG00000142208 | 14 | 104769349 | 104795751 | q32.33 | 20 | ENSP00000451828 | yes | |
| ALDH2 | HGNC:404 | ENSG00000111275 | 12 | 111766887 | 111817532 | q24.12 | 5 | ENSP00000403349 | yes | |
| ALOX15 | HGNC:433 | ENSG00000161905 | 17 | 4630902 | 4642294 | p13.2 | 7 | ENSP00000458832 | yes | |
| ALPG | HGNC:441 | ENSG00000163286 | 2 | 232406844 | 232410714 | q37.1 | 1 | ENSP00000295453 | yes | |
| ALPP | HGNC:439 | ENSG00000163283 | 2 | 232378724 | 232382889 | q37.1 | 3 | ENSP00000375881 | yes | |
| AMHR2 | HGNC:465 | ENSG00000135409 | 12 | 53423855 | 53431672 | q13.13 | 7 | ENSP00000257863 | yes | |
| AMN | HGNC:14604 | ENSG00000166126 | 14 | 102922656 | 102933596 | q32.32 | 7 | ENSP00000299155 | yes | |
| ANGPT2 | HGNC:485 | ENSG00000091879 | 8 | 6499651 | 6563409 | p23.1 | 4 | ENSP00000314897 | yes | |
| ANTXR2 | HGNC:21732 | ENSG00000163297 | 4 | 79901146 | 80125454 | q21.21 | 9 | ENSP00000385575 | yes | |
| ANXA5 | HGNC:543 | ENSG00000164111 | 4 | 121667946 | 121696995 | q27 | 11 | ENSP00000296511 | yes | |
| APLF | HGNC:28724 | ENSG00000169621 | 2 | 68467572 | 68655862 | p13.3 | 7 | ENSP00000307004 | yes | |

| | | | | | | | | | | |
|---|---|---|---|---|---|---|---|---|---|---|
| APOB | HGNC:603 | ENSG00000084674 | 2 | 21001429 | 21044073 | p24.1 | 2 | ENSP00000233242 | yes | |
| APOE | HGNC:613 | ENSG00000130203 | 19 | 44905791 | 44909393 | q13.32 | 5 | ENSP00000252486 | yes | |
| AQP9 | HGNC:643 | ENSG00000103569 | 15 | 58138169 | 58185911 | q21.3 | 4 | ENSP00000452673 | yes | |
| AR | HGNC:644 | ENSG00000169083 | X | 67544021 | 67730619 | q12 | 9 | ENSP00000363822 | yes | |
| AREG | HGNC:651 | ENSG00000109321 | 4 | 74445136 | 74455005 | q13.3 | 3 | ENSP00000379097 | yes | |
| ARNT | HGNC:700 | ENSG00000143437 | 1 | 150809713 | 150876708 | q21.3 | 12 | ENSP00000351407 | yes | |
| ASPH | HGNC:757 | ENSG00000198363 | 8 | 61500556 | 61714640 | q12.3 | 30 | ENSP00000368767 | yes | |
| ATG9B | HGNC:21899 | ENSG00000181652 | 7 | 151012209 | 151024499 | q36.1 | 12 | ENSP00000491504 | yes | long non coding RNA |
| AURKB | HGNC:11390 | ENSG00000178999 | 17 | 8204733 | 8210600 | p13.1 | 14 | ENSP00000313950 | yes | |
| B9D2 | HGNC:28636 | ENSG00000123810 | 19 | 41354417 | 41364165 | q13.2 | 3 | ENSP00000243578 | yes | |
| BAX | HGNC:959 | ENSG00000087088 | 19 | 48954815 | 48961798 | q13.33 | 13 | ENSP00000293288 | yes | |
| BCL2 | HGNC:990 | ENSG00000171791 | 18 | 63123346 | 63320128 | q21.33 | 4 | ENSP00000381185 | yes | |
| BCL9L | HGNC:23688 | ENSG00000186174 | 11 | 118893875 | 118925608 | q23.3 | 5 | ENSP00000335320 | | |
| BIN1 | HGNC:1052 | ENSG00000136717 | 2 | 127048027 | 127107288 | q14.3 | 14 | ENSP00000365281 | yes | |
| BMP2 | HGNC:1069 | ENSG00000125845 | 20 | 6767686 | 6780246 | p12.3 | 1 | ENSP00000368104 | yes | |
| BMP4 | HGNC:1071 | ENSG00000125378 | 14 | 53949736 | 53958761 | q22.2 | 9 | ENSP00000245451 | yes | |
| BMP7 | HGNC:1074 | ENSG00000101144 | 20 | 57168753 | 57266641 | q13.31 | 9 | ENSP00000379204 | yes | |
| BMP8A | HGNC:21650 | ENSG00000183682 | 1 | 39491636 | 39529869 | p34.3 | 1 | ENSP00000327440 | yes | |
| BNC2 | HGNC:30988 | ENSG00000173068 | 9 | 16409503 | 16870843 | p22.2 | 14 | ENSP00000370047 | yes | |
| BOK | HGNC:1087 | ENSG00000176720 | 2 | 241551424 | 241574131 | q37.3 | 2 | ENSP00000314132 | yes | |
| BSG | HGNC:1116 | ENSG00000172270 | 19 | 571277 | 583493 | p13.3 | 16 | ENSP00000473664 | yes | |
| BTC | HGNC:1121 | ENSG00000174808 | 4 | 74744759 | 74794523 | q13.3 | 2 | ENSP00000379092 | yes | |
| BTF3 | HGNC:1125 | ENSG00000145741 | 5 | 73498408 | 73505635 | q13.2 | 9 | ENSP00000338516 | yes | |
| BUB1 | HGNC:1148 | ENSG00000169679 | 2 | 110637528 | 110678063 | q13 | 14 | ENSP00000302530 | yes | |
| BUB1B | HGNC:1149 | ENSG00000156970 | 15 | 40161023 | 40221136 | q15.1 | 11 | ENSP00000287598 | yes | |
| BUB3 | HGNC:1151 | ENSG00000154473 | 10 | 123154402 | 123170467 | q26.13 | 5 | ENSP00000357858 | yes | |
| C10orf55 | HGNC:31008 | ENSG00000222047 | 10 | 73909969 | 73922777 | q22.2 | 2 | ENSP00000409225 | yes | |
| C1orf167 | HGNC:25262 | ENSG00000215910 | 1 | 11761787 | 11789585 | p36.22 | 7 | ENSP00000414909 | yes | |
| C3 | HGNC:1318 | ENSG00000125730 | 19 | 6677704 | 6730562 | p13.3 | 18 | ENSP00000245907 | yes | |
| C4BPA | HGNC:1325 | ENSG00000123838 | 1 | 207104233 | 207144972 | q32.2 | 3 | ENSP00000356037 | yes | |
| C4BPB | HGNC:1328 | ENSG00000123843 | 1 | 207088860 | 207099993 | q32.1 | 8 | ENSP00000243611 | Yes | |
| C5AR1 | HGNC:1338 | ENSG00000197405 | 19 | 47290023 | 47322066 | q13.32 | 3 | ENSP00000347197 | yes | |
| C6orf221 | | | 4 | 162984795 | 162985416 | | 1 | | | Processed Pseudogene |

| | | | | | | | | | | |
|---|---|---|---|---|---|---|---|---|---|---|
| CALCA | HGNC:1437 | ENSG00000110680 | 11 | 14966668 | 14972354 | p15.2 | 6 | ENSP00000417833 | yes | |
| CAMK1 | HGNC:1459 | ENSG00000134072 | 3 | 9757347 | 9769992 | p25.3 | 6 | ENSP00000256460 | yes | |
| CASC15 | HGNC:28245 | ENSG00000272168 | 6 | 21664185 | 22654455 | p22.3 | 105 | | | RNA gene |
| CASP10 | HGNC:1500 | ENSG00000003400 | 2 | 201182881 | 201229406 | q33.1 | 13 | ENSP00000286186 | yes | |
| CASP12 | HGNC:19004 | ENSG00000204403 | 11 | 104885718 | 104898670 | q22.3 | 12 | ENSP00000482745 | yes | Pseudogene |
| CASP3 | HGNC:1504 | ENSG00000164305 | 4 | 184627696 | 184649509 | q35.1 | 6 | ENSP00000377210 | yes | |
| CASP6 | HGNC:1507 | ENSG00000138794 | 4 | 109688622 | 109703583 | q25 | 8 | ENSP00000285333 | yes | |
| CASP8 | HGNC:1509 | ENSG00000064012 | 2 | 201233443 | 201287711 | q33.1 | 20 | ENSP00000376091 | yes | |
| CASP9 | HGNC:1511 | ENSG00000132906 | 1 | 15490832 | 15526534 | p36.21 | 11 | ENSP00000449584 | yes | |
| CBS | HGNC:1550 | ENSG00000160200 | 21 | 43053191 | 43076943 | q22.3 | 17 | ENSP00000381231 | yes | |
| CC2D2A | HGNC:29253 | ENSG00000048342 | 4 | 15469865 | 15601557 | p15.32 | 20 | ENSP00000403465 | | |
| CCDC22 | HGNC:28909 | ENSG00000101997 | X | 49235470 | 49250520 | p11.23 | 3 | ENSP00000365401 | yes | |
| CCR5 | HGNC:1606 | ENSG00000160791 | 3 | 46370854 | 46376206 | p21.31 | 2 | ENSP00000292303 | yes | |
| CD14 | HGNC:1628 | ENSG00000170458 | 5 | 140631728 | 140633701 | q31.3 | 5 | ENSP00000304236 | yes | |
| CD163 | HGNC:1631 | ENSG00000177575 | 12 | 7470813 | 7503893 | p13.31 | 11 | ENSP00000352071 | yes | |
| CD164 | HGNC:1632 | ENSG00000135535 | 6 | 109366514 | 109382467 | q21 | 10 | ENSP00000309376 | yes | |
| CD226 | HGNC:16961 | ENSG00000150637 | 18 | 69831158 | 69961803 | q22.2 | 8 | ENSP00000463152 | yes | |
| CD320 | HGNC:16692 | ENSG00000167775 | 19 | 8302127 | 8308358 | p13.2 | 6 | ENSP00000471773 | yes | |
| CD44 | HGNC:1681 | ENSG00000026508 | 11 | 35138870 | 35232402 | p13 | 39 | ENSP00000263398 | yes | |
| CD46 | HGNC:6953 | ENSG00000117335 | 1 | 207752057 | 207795513 | q32.2 | 19 | ENSP00000350893 | yes | |
| CD55 | HGNC:2665 | ENSG00000196352 | 1 | 207321376 | 207386804 | q32.2 | 14 | ENSP00000356031 | yes | |
| CD68 | HGNC:1693 | ENSG00000129226 | 17 | 7579491 | 7582111 | p13.1 | 4 | ENSP00000250092 | yes | |
| CD69 | HGNC:1694 | ENSG00000110848 | 12 | 9752486 | 9760901 | p13.31 | 4 | ENSP00000228434 | yes | |
| CD7 | HGNC:1695 | ENSG00000173762 | 17 | 82314868 | 82317608 | q25.3 | 7 | ENSP00000463612 | yes | |
| CD82 | HGNC:6210 | ENSG00000085117 | 11 | 44564427 | 44620358 | p11.2 | 14 | ENSP00000435682 | yes | |
| CD84 | HGNC:1704 | ENSG00000066294 | 1 | 160541095 | 160579516 | q23.3 | 8 | ENSP00000357033 | yes | |
| CD8A | HGNC:1706 | ENSG00000153563 | 2 | 86784610 | 86808396 | p11.2 | 4 | ENSP00000321631 | yes | |
| CDH1 | HGNC:1748 | ENSG00000039068 | 16 | 68737292 | 68835541 | q22.1 | 14 | ENSP00000261769 | yes | |
| CDH11 | HGNC:1750 | ENSG00000140937 | 16 | 64943753 | 65126112 | q21 | 19 | ENSP00000268603 | yes | |
| CDKN1C | HGNC:1786 | ENSG00000273707 | CHR_HSCHR11_1_CTG7 | 2883205 | 2885904 | | 5 | ENSP00000488761 | yes | |
| CDKN2B | HGNC:1788 | ENSG00000147883 | 9 | 22002903 | 22009305 | p21.3 | 3 | ENSP00000276925 | yes | |
| CEACAM1 | HGNC:1814 | ENSG00000079385 | 19 | 42507304 | 42561234 | q13.2 | 14 | ENSP00000244291 | yes | |
| CEACAM5 | HGNC:1817 | ENSG00000105388 | 19 | 41708585 | 41729798 | q13.2 | 10 | ENSP00000381600 | yes | |

| | | | | | | | | | |
|---|---|---|---|---|---|---|---|---|---|
| CEACAM6 | HGNC:1818 | ENSG00000086548 | 19 | 41750977 | 41772211 | q13.2 | 2 | ENSP00000469752 | yes |
| CEP128 | HGNC:20359 | ENSG00000100629 | 14 | 80476983 | 80959517 | q31.1 | 16 | ENSP00000451319 | yes |
| CEP250 | HGNC:1859 | ENSG00000126001 | 20 | 35455164 | 35519280 | q11.22 | 14 | ENSP00000398747 | yes |
| CEP295NL | HGNC:44659 | ENSG00000178404 | 17 | 78870910 | 78903217 | q25.3 | 5 | ENSP00000465968 | yes |
| CEP55 | HGNC:1161 | ENSG00000138180 | 10 | 93496612 | 93529092 | q23.33 | 3 | ENSP00000360540 | Yes |
| CFI | HGNC:5394 | ENSG00000205403 | 4 | 109740694 | 109802179 | q25 | 8 | ENSP00000378130 | yes |
| CGB1 | HGNC:16721 | ENSG00000267631 | 19 | 49035610 | 49036895 | q13.33 | 2 | ENSP00000301407 | yes |
| CGB5 | HGNC:16452 | ENSG00000189052 | 19 | 49043848 | 49045311 | q13.33 | 1 | ENSP00000301408 | yes |
| CGB8 | HGNC:16453 | ENSG00000213030 | 19 | 49047638 | 49049106 | q13.33 | 1 | ENSP00000403649 | yes |
| CHIA | HGNC:17432 | ENSG00000134216 | 1 | 111290851 | 111320566 | p13.2 | 10 | ENSP00000387671 | yes |
| CHRNA1 | HGNC:1955 | ENSG00000138435 | 2 | 174747592 | 174787935 | q31.1 | 9 | ENSP00000500507 | yes |
| CKAP5 | HGNC:28959 | ENSG00000175216 | 11 | 46743048 | 46846308 | p11.2 | 12 | ENSP00000432768 | yes |
| CLDN4 | HGNC:2046 | ENSG00000189143 | 7 | 73799542 | 73832693 | q11.23 | 5 | ENSP00000409544 | yes |
| COL13A1 | HGNC:2190 | ENSG00000197467 | 10 | 69801867 | 69964275 | q22.1 | 15 | ENSP00000496051 | yes |
| COL1A1 | HGNC:2197 | ENSG00000108821 | 17 | 50183289 | 50201632 | q21.33 | 13 | ENSP00000225964 | yes |
| COL1A2 | HGNC:2198 | ENSG00000164692 | 7 | 94394561 | 94431232 | q21.3 | 13 | ENSP00000297268 | yes |
| COL2A1 | HGNC:2200 | ENSG00000139219 | 12 | 47972967 | 48004554 | q13.11 | 9 | ENSP00000369889 | yes |
| COL4A6 | HGNC:2208 | ENSG00000197565 | X | 108155607 | 108439497 | q22.3 | 10 | ENSP00000334733 | yes |
| COL5A1 | HGNC:2209 | ENSG00000130635 | 9 | 134641774 | 134844843 | q34.3 | 8 | ENSP00000360882 | yes |
| COL5A2 | HGNC:2210 | ENSG00000204262 | 2 | 189031898 | 189225312 | q32.2 | 4 | ENSP00000364000 | yes |
| COL6A1 | HGNC:2211 | ENSG00000142156 | 21 | 45981737 | 46005050 | q22.3 | 7 | ENSP00000355180 | yes |
| COL6A3 | HGNC:2213 | ENSG00000163359 | 2 | 237324003 | 237414375 | q37.3 | 14 | ENSP00000295550 | yes |
| COL9A2 | HGNC:2218 | ENSG00000049089 | 1 | 40300489 | 40317813 | p34.2 | 12 | ENSP00000361834 | yes |
| COMT | HGNC:2228 | ENSG00000093010 | 22 | 19941607 | 19969975 | q11.21 | 11 | ENSP00000354511 | yes |
| CPB2 | HGNC:2300 | ENSG00000080618 | 13 | 46053186 | 46105033 | q14.13 | 2 | ENSP00000181383 | yes |
| CR1 | HGNC:2334 | ENSG00000203710 | 1 | 207496147 | 207641765 | q32.2 | 12 | ENSP00000383744 | yes |
| CRISP3 | HGNC:16904 | ENSG00000096006 | 6 | 49727384 | 49744437 | p12.3 | 4 | ENSP00000263045 | yes |
| CSF1 | HGNC:2432 | ENSG00000184371 | 1 | 109910242 | 109930992 | p13.3 | 9 | ENSP00000434527 | yes |
| CSF1R | HGNC:2433 | ENSG00000182578 | 5 | 150053291 | 150113372 | q32 | 9 | ENSP00000422212 | yes |
| CSF3 | HGNC:2438 | ENSG00000108342 | 17 | 40015361 | 40017813 | q21.1 | 9 | ENSP00000464421 | yes |
| CSRNP3 | HGNC:30729 | ENSG00000178662 | 2 | 165469647 | 165689407 | q24.3 | 7 | ENSP00000412081 | yes |
| CTLA4 | HGNC:2505 | ENSG00000163599 | 2 | 203867771 | 203873965 | q33.2 | 6 | ENSP00000497102 | yes |
| CTNNA3 | HGNC:2511 | ENSG00000183230 | 10 | 65912523 | 67696195 | q21.3 | 5 | ENSP00000389714 | yes |
| CX3CR1 | HGNC:2558 | ENSG00000168329 | 3 | 39263494 | 39281735 | p22.2 | 6 | ENSP00000382166 | yes |

| | | | | | | | | | | |
|---|---|---|---|---|---|---|---|---|---|---|
| CXCL10 | HGNC:10637 | ENSG00000169245 | 4 | 76021118 | 76023497 | q21.1 | 1 | ENSP00000305651 | yes | |
| CXCL8 | HGNC:6025 | ENSG00000169429 | 4 | 73740541 | 73743716 | q13.3 | 3 | ENSP00000306512 | yes | |
| CYBB | HGNC:2578 | ENSG00000165168 | X | 37780011 | 37813461 | p21.1 | 2 | ENSP00000367851 | yes | |
| CYP17A1 | HGNC:2593 | ENSG00000148795 | 10 | 102830531 | 102837472 | q24.32 | 8 | ENSP00000492313 | yes | |
| CYP19A1 | HGNC:2594 | ENSG00000137869 | 15 | 51208057 | 51338601 | q21.2 | 20 | ENSP00000379683 | yes | |
| CYP1A1 | HGNC:2595 | ENSG00000140465 | 15 | 74719542 | 74725536 | q24.1 | 10 | ENSP00000369050 | yes | |
| CYP1A2 | HGNC:2596 | ENSG00000140505 | 15 | 74748845 | 74756607 | q24.1 | 1 | ENSP00000342007 | yes | |
| CYP1B1 | HGNC:2597 | ENSG00000138061 | 2 | 38066973 | 38109902 | p22.2 | 7 | ENSP00000478561 | yes | |
| CYP1B1-AS1 | HGNC:28543 | ENSG00000232973 | 2 | 38073447 | 38231651 | p22.2 | 40 | | | RNA gene |
| CYP24A1 | HGNC:2602 | ENSG00000019186 | 20 | 54153446 | 54173986 | q13.2 | 6 | ENSP00000216862 | yes | |
| CYP2D6 | HGNC:2625 | ENSG00000272532 | CHR_HSCHR22_2_CTG1 | 42126481 | 42130888 | | 2 | ENSP00000476021 | yes | |
| DHFR | HGNC:2861 | ENSG00000228716 | 5 | 80626226 | 80654983 | q14.1 | 7 | ENSP00000396308 | yes | |
| DIAPH2-AS1 | HGNC:16972 | ENSG00000236256 | X | 97431286 | 97642589 | q21.33 | 4 | | | RNA gene |
| DICER1 | HGNC:17098 | ENSG00000100697 | 14 | 95086228 | 95158010 | q32.13 | 15 | ENSP00000343745 | yes | |
| DLC1 | HGNC:2897 | ENSG00000164741 | 8 | 13083361 | 13604610 | p22 | 18 | ENSP00000276297 | yes | |
| DLL4 | HGNC:2910 | ENSG00000128917 | 15 | 40929340 | 40939073 | q15.1 | 3 | ENSP00000249749 | | |
| DNM2 | HGNC:2974 | ENSG00000079805 | 19 | 10718079 | 10833488 | p13.2 | 22 | ENSP00000468734 | yes | |
| DNMT3L | HGNC:2980 | ENSG00000142182 | 21 | 44246339 | 44262216 | q22.3 | 4 | ENSP00000270172 | Yes | |
| DROSHA | HGNC:17904 | ENSG00000113360 | 5 | 31400497 | 31532196 | p13.3 | 21 | ENSP00000425979 | yes | |
| DYNC2H1 | HGNC:2962 | ENSG00000187240 | 11 | 103109410 | 103479863 | q22.3 | 11 | ENSP00000364887 | Yes | |
| ECEL1 | HGNC:3147 | ENSG00000171551 | 2 | 232479827 | 232487834 | q37.1 | 4 | ENSP00000302051 | Yes | |
| ECM1 | HGNC:3153 | ENSG00000143369 | 1 | 150508062 | 150513789 | q21.2 | 7 | ENSP00000358045 | yes | |
| ECM2 | HGNC:3154 | ENSG00000106823 | 9 | 92493554 | 92536655 | q22.31 | 4 | ENSP00000344758 | yes | |
| EGFR | HGNC:3236 | ENSG00000146648 | 7 | 55019017 | 55211628 | p11.2 | 11 | ENSP00000415559 | yes | |
| EMP1 | HGNC:3333 | ENSG00000134531 | 12 | 13196723 | 13219941 | p13.1 | 12 | ENSP00000256951 | yes | |
| EOMES | HGNC:3372 | ENSG00000163508 | 3 | 27715949 | 27722711 | p24.1 | 3 | ENSP00000295743 | yes | |
| EPAS1 | HGNC:3374 | ENSG00000116016 | 2 | 46293667 | 46386697 | p21 | 10 | ENSP00000263734 | yes | |
| ERBB4 | HGNC:3432 | ENSG00000178568 | 2 | 211375717 | 212538841 | q34 | 9 | ENSP00000342235 | yes | |
| EREG | HGNC:3443 | ENSG00000124882 | 4 | 74365145 | 74388749 | q13.3 | 3 | ENSP00000244869 | yes | |
| ERICH2 | HGNC:44395 | ENSG00000204334 | 2 | 170783786 | 170798971 | q31.1 | 1 | ENSP00000387298 | | |
| ESR1 | HGNC:3467 | ENSG00000091831 | 6 | 151656691 | 152129619 | q25.1 | 15 | ENSP00000342630 | yes | |
| ESR2 | HGNC:3468 | ENSG00000140009 | 14 | 64084232 | 64338112 | q23.3 | 13 | ENSP00000452485 | yes | |
| F12 | HGNC:3530 | ENSG00000131187 | 5 | 177402140 | 177409576 | q35.3 | 6 | ENSP00000253496 | yes | |

| | | | | | | | | | |
|---|---|---|---|---|---|---|---|---|---|
| F13A1 | HGNC:3531 | ENSG00000124491 | 6 | 6144085 | 6321013 | p25.1 | 6 | ENSP00000264870 | yes |
| F2 | HGNC:3535 | ENSG00000180210 | 11 | 46719180 | 46739506 | p11.2 | 5 | ENSP00000308541 | yes |
| F2R | HGNC:3537 | ENSG00000181104 | 5 | 76716126 | 76735770 | q13.3 | 2 | ENSP00000321326 | yes |
| F5 | HGNC:3542 | ENSG00000198734 | 1 | 169511953 | 169586588 | q24.2 | 3 | ENSP00000356771 | yes |
| F7 | HGNC:3544 | ENSG00000057593 | 13 | 113105788 | 113120681 | q34 | 6 | ENSP00000329546 | yes |
| FAS | HGNC:11920 | ENSG00000026103 | 10 | 88990531 | 89017059 | q23.31 | 21 | ENSP00000477997 | yes |
| FASLG | HGNC:11936 | ENSG00000117560 | 1 | 172659103 | 172666876 | q24.3 | 2 | ENSP00000356694 | yes |
| FASTKD3 | HGNC:28758 | ENSG00000124279 | 5 | 7859159 | 7869037 | p15.31 | 8 | ENSP00000264669 | yes |
| FBLN1 | HGNC:3600 | ENSG00000077942 | 22 | 45502238 | 45601135 | q13.31 | 18 | ENSP00000415289 | yes |
| FETUB | HGNC:3658 | ENSG00000090512 | 3 | 186635969 | 186653141 | q27.3 | 8 | ENSP00000396581 | yes |
| FGA | HGNC:3661 | ENSG00000171560 | 4 | 154583128 | 154590745 | q31.3 | 3 | ENSP00000498441 | yes |
| FGB | HGNC:3662 | ENSG00000171564 | 4 | 154562956 | 154571086 | q31.3 | 7 | ENSP00000306099 | yes |
| FGF7 | HGNC:3685 | ENSG00000140285 | 15 | 49423178 | 49488775 | q21.2 | 6 | ENSP00000267843 | yes |
| FGF9 | HGNC:3687 | ENSG00000102678 | 13 | 21671073 | 21704498 | q12.11 | 3 | ENSP00000371790 | yes |
| FGFR2 | HGNC:3689 | ENSG00000066468 | 10 | 121478334 | 121598458 | q26.13 | 27 | ENSP00000351276 | yes |
| FGFR3 | HGNC:3690 | ENSG00000068078 | 4 | 1793293 | 1808872 | p16.3 | 11 | ENSP00000414914 | yes |
| FGG | HGNC:3694 | ENSG00000171557 | 4 | 154604134 | 154612967 | q32.1 | 12 | ENSP00000384860 | yes |
| FHIT | HGNC:3701 | ENSG00000189283 | 3 | 59747277 | 61251459 | p14.2 | 8 | ENSP00000417557 | yes |
| FKBP4 | HGNC:3720 | ENSG00000004478 | 12 | 2794970 | 2805423 | p13.33 | 8 | ENSP00000001008 | yes |
| FLRT3 | HGNC:3762 | ENSG00000125848 | 20 | 14322985 | 14337614 | p12.1 | 3 | ENSP00000367292 | yes |
| FLT1 | HGNC:3763 | ENSG00000102755 | 13 | 28300346 | 28495145 | q12.3 | 9 | ENSP00000282397 | yes |
| FLVCR2 | HGNC:20105 | ENSG00000119686 | 14 | 75578620 | 75663214 | q24.3 | 13 | ENSP00000238667 | yes |
| FMR1 | HGNC:3775 | ENSG00000102081 | X | 147911951 | 147951125 | q27.3 | 21 | ENSP00000218200 | Yes |
| FN1 | HGNC:3778 | ENSG00000115414 | 2 | 215360440 | 215436073 | q35 | 27 | ENSP00000394423 | yes |
| FOXA2 | HGNC:5022 | ENSG00000125798 | 20 | 22581005 | 22585455 | p11.21 | 2 | ENSP00000400341 | yes |
| FOXD1 | HGNC:3802 | ENSG00000251493 | 5 | 73444827 | 73448777 | q13.2 | 2 | ENSP00000481581 | yes |
| FOXP3 | HGNC:6106 | ENSG00000049768 | X | 49250436 | 49270477 | p11.23 | 9 | ENSP00000365380 | yes |
| FRAS1 | HGNC:19185 | ENSG00000138759 | 4 | 78057323 | 78544269 | q21.21 | 7 | ENSP00000326330 | yes |
| FREM3 | HGNC:25172 | ENSG00000183090 | 4 | 143577302 | 143700675 | q31.21 | 2 | ENSP00000332886 | |
| FSHR | HGNC:3969 | ENSG00000170820 | 2 | 48962157 | 49154537 | p16.3 | 5 | ENSP00000384708 | yes |
| FST | HGNC:3971 | ENSG00000134363 | 5 | 53480626 | 53487134 | q11.2 | 5 | ENSP00000256759 | yes |
| FZD4 | HGNC:4042 | ENSG00000174804 | 11 | 86945679 | 86955395 | q14.2 | 1 | ENSP00000434034 | yes |
| FZD6 | HGNC:4044 | ENSG00000164930 | 8 | 103298433 | 103332866 | q22.3 | 7 | ENSP00000351605 | Yes |
| GBE1 | HGNC:4180 | ENSG00000114480 | 3 | 81489703 | 81761645 | p12.2 | 6 | ENSP00000410833 | yes |

| | | | | | | | | | | |
|---|---|---|---|---|---|---|---|---|---|---|
| GCLC | HGNC:4311 | ENSG00000001084 | 6 | 53497341 | 53616970 | p12.1 | 14 | ENSP00000495056 | yes | |
| GDF15 | HGNC:30142 | ENSG00000130513 | 19 | 18374731 | 18389176 | p13.11 | 5 | ENSP00000469819 | yes | |
| GHR | HGNC:4263 | ENSG00000112964 | 5 | 42423439 | 42721878 | p13.1 | 13 | ENSP00000230882 | yes | |
| GLE1 | HGNC:4315 | ENSG00000119392 | 9 | 128504700 | 128542288 | q34.11 | 3 | ENSP00000308622 | Yes | |
| GLS | HGNC:4331 | ENSG00000115419 | 2 | 190880827 | 190965552 | q32.2 | 15 | ENSP00000317379 | yes | |
| GNLY | HGNC:4414 | ENSG00000115523 | 2 | 85685175 | 85698854 | p11.2 | 14 | ENSP00000387116 | yes | |
| GOLPH3 | HGNC:15452 | ENSG00000113384 | 5 | 32124716 | 32174319 | p13.3 | 3 | ENSP00000265070 | yes | |
| GP6 | HGNC:14388 | ENSG00000277439 | CHR_HSCHR19_4_CTG3_1 | 55021820 | 55046379 | | 5 | ENSP00000479873 | yes | |
| GPC1 | HGNC:4449 | ENSG00000063660 | 2 | 240435663 | 240468076 | q37.3 | 9 | ENSP00000264039 | yes | |
| GPHN | HGNC:15465 | ENSG00000171723 | 14 | 66507407 | 67181803 | q23.3 | 16 | ENSP00000312771 | yes | |
| GPX3 | HGNC:4555 | ENSG00000211445 | 5 | 151020438 | 151028992 | q33.1 | 11 | ENSP00000373477 | yes | |
| GPX4 | HGNC:4556 | ENSG00000167468 | 19 | 1103926 | 1106791 | p13.3 | 13 | ENSP00000346103 | yes | |
| GRIK2 | HGNC:4580 | ENSG00000164418 | 6 | 101181257 | 102070083 | q16.3 | 11 | ENSP00000397026 | yes | |
| GRIN3A | HGNC:16767 | ENSG00000198785 | 9 | 101569353 | 101738580 | q31.1 | 2 | ENSP00000355155 | | |
| GRK2 | HGNC:289 | ENSG00000173020 | 11 | 67266473 | 67286556 | q13.2 | 14 | ENSP00000312262 | yes | |
| GRK6 | HGNC:4545 | ENSG00000198055 | 5 | 177403204 | 177442901 | q35.3 | 11 | ENSP00000422873 | yes | |
| GRP | HGNC:4605 | ENSG00000134443 | 18 | 59220158 | 59230774 | q21.32 | 5 | ENSP00000256857 | yes | |
| GSTA1 | HGNC:4626 | ENSG00000243955 | 6 | 52791371 | 52803860 | p12.2 | 3 | ENSP00000335620 | yes | |
| GSTA6P | HGNC:4630 | ENSG00000223622 | 6 | 52805613 | 52813460 | p12.2 | 1 | | | Unprocessed Psuedogene |
| GSTM1 | HGNC:4632 | ENSG00000134184 | 1 | 109687814 | 109709039 | p13.3 | 8 | ENSP00000311469 | Yes | |
| GSTO1 | HGNC:13312 | ENSG00000148834 | 10 | 104235356 | 104267459 | q25.1 | 7 | ENSP00000358727 | Yes | |
| GSTO2 | HGNC:23064 | ENSG00000065621 | 10 | 104268873 | 104304950 | q25.1 | 7 | ENSP00000345023 | Yes | |
| GSTP1 | HGNC:4638 | ENSG00000084207 | 11 | 67583595 | 67586656 | q13.2 | 8 | ENSP00000381607 | Yes | |
| GSTT1 | HGNC:4641 | ENSG00000277656 | CHR_HSCHR22_1_CTG7 | 24033952 | 24042493 | | 15 | ENSP00000485976 | Yes | |
| GTF2A1L | HGNC:30727 | ENSG00000242441 | 2 | 48617798 | 48733148 | p16.3 | 7 | ENSP00000412645 | Yes | |
| H19 | HGNC:4713 | ENSG00000130600 | 11 | 1995176 | 2001470 | p15.5 | 12 | | | RNA gene |
| HABP2 | HGNC:4798 | ENSG00000148702 | 10 | 113550837 | 113589602 | q25.3 | 3 | ENSP00000277903 | Yes | |
| HAVCR2 | HGNC:18437 | ENSG00000135077 | 5 | 157085832 | 157142869 | q33.3 | 6 | ENSP00000312002 | Yes | |
| HCP5 | HGNC:21659 | ENSG00000227429 | CHR_HSCHR6_MHC_MCF_CTG1 | 31540726 | 31542189 | | 2 | | | RNA gene |
| HDC | HGNC:4855 | ENSG00000140287 | 15 | 50241947 | 50265965 | q21.2 | 7 | ENSP00000267845 | Yes | |
| HESX1 | HGNC:4877 | ENSG00000163666 | 3 | 57197838 | 57227606 | p14.3 | 4 | ENSP00000498190 | Yes | |

| | | | | | | | | | | |
|---|---|---|---|---|---|---|---|---|---|---|
| HLA-A | HGNC:4931 | ENSG00000206503 | 6 | 29941260 | 29945884 | p22.1 | 9 | ENSP00000366005 | Yes | |
| HLA-C | HGNC:4933 | ENSG00000204525 | 6 | 31268749 | 31272130 | p21.33 | 10 | ENSP00000365402 | Yes | |
| HLA-DPB1 | HGNC:4940 | ENSG00000226826 | CHR_HSCHR6_MHC_SSTO_CTG1 | 33216374 | 33227649 | | 9 | ENSP00000407674 | Yes | Pseudogene |
| HLA-DQA1 | HGNC:4942 | ENSG00000196735 | 6 | 32628179 | 32647062 | p21.32 | 7 | ENSP00000405797 | Yes | |
| HLA-DQB1 | HGNC:4944 | ENSG00000206237 | CHR_HSCHR6_MHC_COX_CTG1 | 32582490 | 32590101 | | 1 | ENSP00000372602 | Yes | |
| HLA-DRB1 | HGNC:4948 | ENSG00000196126 | 6 | 32578769 | 32589848 | p21.32 | 1 | ENSP00000353099 | Yes | |
| HLA-DRB5 | HGNC:4953 | ENSG00000198502 | 6 | 32517353 | 32530287 | p21.32 | 1 | ENSP00000364114 | Yes | |
| HLA-E | HGNC:4962 | ENSG00000204592 | 6 | 30489509 | 30494194 | p22.1 | 3 | ENSP00000365817 | Yes | |
| HLA-G | HGNC:4964 | ENSG00000204632 | 6 | 29826967 | 29831125 | p22.1 | 7 | ENSP00000412927 | Yes | |
| HMOX1 | HGNC:5013 | ENSG00000100292 | 22 | 35380361 | 35394207 | q12.3 | 4 | ENSP00000413316 | Yes | |
| HMX3 | HGNC:5019 | ENSG00000188620 | 10 | 123135970 | 123139423 | q26.13 | 1 | ENSP00000350549 | Yes | |
| HOXA10 | HGNC:5100 | ENSG00000253293 | 7 | 27170592 | 27180261 | p15.2 | 6 | ENSP00000283921 | Yes | |
| HOXA11 | HGNC:5101 | ENSG00000005073 | 7 | 27181510 | 27185223 | p15.2 | 2 | ENSP00000448962 | Yes | |
| HOXA13 | HGNC:5102 | ENSG00000106031 | 7 | 27193503 | 27200091 | p15.2 | 2 | ENSP00000497112 | Yes | |
| HOXC10 | HGNC:5122 | ENSG00000180818 | 12 | 53985065 | 53990279 | q13.13 | 5 | ENSP00000421946 | Yes | |
| HOXC6 | HGNC:5128 | ENSG00000197757 | 12 | 53990624 | 54030823 | q13.13 | 4 | ENSP00000424124 | Yes | |
| HOXC9 | HGNC:5130 | ENSG00000180806 | 12 | 53994895 | 54003337 | q13.13 | 3 | ENSP00000302836 | Yes | |
| HOXC-AS2 | HGNC:43750 | ENSG00000250133 | 12 | 53993810 | 53996785 | q13.13 | 2 | | | Retained Intron/processed transcript/RNA gene |
| HRG | HGNC:5181 | ENSG00000113905 | 3 | 186660216 | 186678240 | q27.3 | 3 | ENSP00000232003 | Yes | |
| HRH1 | HGNC:5182 | ENSG00000196639 | 3 | 11137093 | 11263557 | p25.3 | 4 | ENSP00000406705 | Yes | |
| HRH2 | HGNC:5183 | ENSG00000113749 | 5 | 175658030 | 175710756 | q35.2 | 4 | ENSP00000366506 | Yes | |
| HSD17B1 | HGNC:5210 | ENSG00000108786 | 17 | 42549214 | 42555213 | q21.2 | 5 | ENSP00000466799 | Yes | |
| HSD3B1 | HGNC:5217 | ENSG00000203857 | 1 | 119507198 | 119515054 | p12 | 5 | ENSP00000435999 | Yes | |
| HTR1A | HGNC:5286 | ENSG00000178394 | 5 | 63960356 | 63962507 | q12.3 | 2 | ENSP00000316244 | Yes | |
| ICAM1 | HGNC:5344 | ENSG00000090339 | 19 | 10271093 | 10286615 | p13.2 | 5 | ENSP00000264832 | Yes | |
| IDO1 | HGNC:6059 | ENSG00000131203 | 8 | 39902275 | 39928790 | p11.21 | 9 | ENSP00000429297 | Yes | |
| IDO2 | HGNC:27269 | ENSG00000188676 | 8 | 39934614 | 40016391 | p11.21 | 4 | ENSP00000443432 | Yes | |
| IDUA | HGNC:5391 | ENSG00000127415 | 4 | 986997 | 1004564 | p16.3 | 13 | ENSP00000247933 | Yes | |
| IFI35 | HGNC:5399 | ENSG00000068079 | 17 | 43006725 | 43014456 | q21.31 | 8 | ENSP00000394579 | Yes | |
| IFI44 | HGNC:16938 | ENSG00000137965 | 1 | 78649796 | 78664078 | p31.1 | 9 | ENSP00000359783 | Yes | |
| IFI6 | HGNC:4054 | ENSG00000126709 | 1 | 27666064 | 27672198 | p35.3 | 3 | ENSP00000354736 | Yes | |

| | | | | | | | | | |
|---|---|---|---|---|---|---|---|---|---|
| IFNA1 | HGNC:5417 | ENSG00000197919 | 9 | 21440439 | 21441316 | p21.3 | 1 | ENSP00000276927 | Yes |
| IFNA10 | HGNC:5418 | ENSG00000186803 | 9 | 21206181 | 21207143 | p21.3 | 1 | ENSP00000369566 | Yes |
| IFNG | HGNC:5438 | ENSG00000111537 | 12 | 68154768 | 68159740 | q15 | 1 | ENSP00000229135 | Yes |
| IFNGR1 | HGNC:5439 | ENSG00000027697 | 6 | 137197484 | 137219449 | q23.3 | 12 | ENSP00000356713 | Yes |
| IFNW1 | HGNC:5448 | ENSG00000177047 | 9 | 21140214 | 21142145 | p21.3 | 1 | ENSP00000369578 | Yes |
| IFT122 | HGNC:13556 | ENSG00000163913 | 3 | 129440036 | 129520507 | q21.3 | 32 | ENSP00000323973 | Yes |
| IGF1 | HGNC:5464 | ENSG00000017427 | 12 | 102395874 | 102481744 | q23.2 | 7 | ENSP00000337612 | Yes |
| IGF2 | HGNC:5466 | ENSG00000167244 | 11 | 2129112 | 2141238 | p15.5 | 7 | ENSP00000370802 | Yes |
| IGF2R | HGNC:5467 | ENSG00000197081 | 6 | 159969099 | 160113507 | q25.3 | 6 | ENSP00000349437 | Yes |
| IGFBP1 | HGNC:5469 | ENSG00000146678 | 7 | 45888360 | 45893660 | p12.3 | 3 | ENSP00000275525 | Yes |
| IGFBP3 | HGNC:5472 | ENSG00000146674 | 7 | 45912245 | 45921874 | p12.3 | 11 | ENSP00000370476 | Yes |
| IGFBP6 | HGNC:5475 | ENSG00000167779 | 12 | 53097436 | 53102345 | q13.13 | 5 | ENSP00000448953 | Yes |
| IHH | HGNC:5956 | ENSG00000163501 | 2 | 219054424 | 219060921 | q35 | 1 | ENSP00000295731 | Yes |
| IL10 | HGNC:5962 | ENSG00000136634 | 1 | 206767602 | 206774541 | q32.1 | 7 | ENSP00000412237 | Yes |
| IL10RA | HGNC:5964 | ENSG00000110324 | 11 | 117986348 | 118003037 | q23.3 | 11 | ENSP00000227752 | Yes |
| IL11 | HGNC:5966 | ENSG00000095752 | 19 | 55364382 | 55370463 | q13.42 | 4 | ENSP00000264563 | Yes |
| IL11RA | HGNC:5967 | ENSG00000137070 | 9 | 34650702 | 34661892 | p13.3 | 15 | ENSP00000450565 | Yes |
| IL12A | HGNC:5969 | ENSG00000168811 | 3 | 159988835 | 159996019 | q25.33 | 6 | ENSP00000303231 | Yes |
| IL12B | HGNC:5970 | ENSG00000113302 | 5 | 159314783 | 159330887 | q33.3 | 1 | ENSP00000231228 | Yes |
| IL13 | HGNC:5973 | ENSG00000169194 | 5 | 132656263 | 132661110 | q31.1 | 6 | ENSP00000304915 | Yes |
| IL15 | HGNC:5977 | ENSG00000164136 | 4 | 141636583 | 141733987 | q31.21 | 8 | ENSP00000323505 | Yes |
| IL16 | HGNC:5980 | ENSG00000172349 | 15 | 81159575 | 81314058 | q25.1 | 13 | ENSP00000378155 | Yes |
| IL17A | HGNC:5981 | ENSG00000112115 | 6 | 52186375 | 52190638 | p12.2 | 1 | ENSP00000497968 | Yes |
| IL17F | HGNC:16404 | ENSG00000112116 | 6 | 52236681 | 52244537 | p12.2 | 2 | ENSP00000337432 | Yes |
| IL17RB | HGNC:18015 | ENSG00000056736 | 3 | 53846568 | 53865794 | p21.1 | 3 | ENSP00000288167 | Yes |
| IL18 | HGNC:5986 | ENSG00000150782 | 11 | 112143253 | 112164096 | q23.1 | 6 | ENSP00000280357 | Yes |
| IL1A | HGNC:5991 | ENSG00000115008 | 2 | 112773915 | 112784590 | q14.1 | 1 | ENSP00000263339 | Yes |
| IL1B | HGNC:5992 | ENSG00000125538 | 2 | 112829751 | 112836816 | q14.1 | 8 | ENSP00000263341 | Yes |
| IL1R1 | HGNC:5993 | ENSG00000115594 | 2 | 102064544 | 102179874 | q11.2 | 13 | ENSP00000401646 | Yes |
| IL1R2 | HGNC:5994 | ENSG00000115590 | 2 | 101991960 | 102028544 | q11.2 | 9 | ENSP00000330959 | Yes |
| IL1RL1 | HGNC:5998 | ENSG00000115602 | 2 | 102311502 | 102352037 | q12.1 | 9 | ENSP00000233954 | Yes |
| IL1RN | HGNC:6000 | ENSG00000136689 | 2 | 113107214 | 113134016 | q14.1 | 9 | ENSP00000354816 | Yes |
| IL2 | HGNC:6001 | ENSG00000109471 | 4 | 122451470 | 122456725 | q27 | 2 | ENSP00000226730 | Yes |
| IL20RA | HGNC:6003 | ENSG00000016402 | 6 | 136999971 | 137045180 | q23.3 | 8 | ENSP00000314976 | Yes |

| | | | | | | | | | | |
|---|---|---|---|---|---|---|---|---|---|---|
| IL21 | HGNC:6005 | ENSG00000138684 | 4 | 122610108 | 122621066 | q27 | 3 | ENSP00000497915 | Yes | |
| IL21-AS1 | HGNC:40299 | ENSG00000227145 | 4 | 122618983 | 122689156 | q27 | 4 | | | RNA gene |
| IL24 | HGNC:11346 | ENSG00000162892 | 1 | 206897443 | 206904139 | q32.1 | 7 | ENSP00000375795 | Yes | |
| IL25 | HGNC:13765 | ENSG00000166090 | 14 | 23372809 | 23376403 | q11.2 | 2 | ENSP00000380417 | Yes | |
| IL2RA | HGNC:6008 | ENSG00000134460 | 10 | 6010689 | 6062370 | p15.1 | 6 | ENSP00000369293 | Yes | |
| IL33 | HGNC:16028 | ENSG00000137033 | 9 | 6215786 | 6257983 | p24.1 | 5 | ENSP00000370842 | Yes | |
| IL4 | HGNC:6014 | ENSG00000113520 | 5 | 132673986 | 132682678 | q31.1 | 4 | ENSP00000231449 | Yes | |
| IL4R | HGNC:6015 | ENSG00000077238 | 16 | 27313668 | 27364778 | p12.1 | 19 | ENSP00000455632 | Yes | |
| IL5 | HGNC:6016 | ENSG00000113525 | 5 | 132541445 | 132556838 | q31.1 | 3 | ENSP00000231454 | Yes | |
| IL5RA | HGNC:6017 | ENSG00000091181 | 3 | 3066326 | 3126613 | p26.2 | 11 | ENSP00000412209 | Yes | |
| IL6 | HGNC:6018 | ENSG00000136244 | 7 | 22725884 | 22732002 | p15.3 | 9 | ENSP00000385675 | Yes | |
| IL6R | HGNC:6019 | ENSG00000160712 | 1 | 154405193 | 154469450 | q21.3 | 8 | ENSP00000357470 | Yes | |
| IL6ST | HGNC:6021 | ENSG00000134352 | 5 | 55935095 | 55995022 | q11.2 | 16 | ENSP00000370698 | Yes | |
| IL9 | HGNC:6029 | ENSG00000145839 | 5 | 135892246 | 135895841 | q31.1 | 1 | ENSP00000274520 | Yes | |
| INHA | HGNC:6065 | ENSG00000123999 | 2 | 219569162 | 219575711 | q35 | 2 | ENSP00000243786 | Yes | |
| INHBA | HGNC:6066 | ENSG00000122641 | 7 | 41667168 | 41705834 | p14.1 | 5 | ENSP00000242208 | Yes | |
| INS-IGF2 | HGNC:33527 | ENSG00000129965 | 11 | 2132538 | 2161209 | p15.5 | 2 | ENSP00000348986 | Yes | |
| IRF1 | HGNC:6116 | ENSG00000125347 | 5 | 132481609 | 132490777 | q31.1 | 11 | ENSP00000245414 | Yes | |
| ITGA2 | HGNC:6137 | ENSG00000164171 | 5 | 52989340 | 53094779 | q11.2 | 6 | ENSP00000296585 | Yes | |
| ITGA5 | HGNC:6141 | ENSG00000161638 | 12 | 54395261 | 54419266 | q13.13 | 13 | ENSP00000293379 | Yes | |
| ITGB3 | HGNC:6156 | ENSG00000259207 | 17 | 47253846 | 47311816 | q21.32 | 3 | ENSP00000452786 | Yes | |
| ITGB4 | HGNC:6158 | ENSG00000132470 | 17 | 75721328 | 75757818 | q25.1 | 13 | ENSP00000463651 | Yes | |
| ITGB6 | HGNC:6161 | ENSG00000115221 | 2 | 160099667 | 160200313 | q24.2 | 8 | ENSP00000283249 | Yes | |
| JAK2 | HGNC:6192 | ENSG00000096968 | 9 | 4984390 | 5128183 | p24.1 | 4 | ENSP00000489812 | Yes | |
| KCNH2 | HGNC:6251 | ENSG00000055118 | 7 | 150944961 | 150978321 | q36.1 | 6 | ENSP00000328531 | Yes | |
| KCNQ1OT1 | HGNC:6295 | ENSG00000269821 | 11 | 2608328 | 2699994 | p15.5 | 1 | | | long non coding RNA |
| KDR | HGNC:6307 | ENSG00000128052 | 4 | 55078481 | 55125595 | q12 | 4 | ENSP00000263923 | Yes | |
| KHDC3L | HGNC:33699 | ENSG00000203908 | 6 | 73362658 | 73364171 | q13 | 1 | ENSP00000359392 | Yes | |
| KIF14 | HGNC:19181 | ENSG00000118193 | 1 | 200551497 | 200620751 | q32.1 | 2 | ENSP00000356319 | Yes | |
| KIF1BP | HGNC:23419 | ENSG00000198954 | 10 | 68988721 | 69043544 | q22.1 | 14 | ENSP00000490704 | Yes | |
| KIF26A | HGNC:20226 | ENSG00000066735 | 14 | 104138723 | 104180894 | q32.33 | 2 | ENSP00000388241 | | |
| KIF7 | HGNC:30497 | ENSG00000166813 | 15 | 89608789 | 89655467 | q26.1 | 3 | ENSP00000377934 | Yes | |

| | | | | | | | | | | |
|---|---|---|---|---|---|---|---|---|---|---|
| KIR2DL1 | HGNC:6329 | ENSG00000277356 | CHR_HSCHR19KIR_FH06_A_HAP_CTG3_1 | 54769746 | 54783981 | | 1 | ENSP00000480247 | Yes | |
| KIR2DL2 | HGNC:6330 | ENSG00000277725 | CHR_HSCHR19KIR_FH06_BA1_HAP_CTG3_1 | 54754632 | 54769000 | | 1 | ENSP00000484319 | Yes | |
| KIR2DL3 | HGNC:6331 | ENSG00000274108 | CHR_HSCHR19KIR_FH06_A_HAP_CTG3_1 | 54738481 | 54752986 | | 1 | ENSP00000483192 | Yes | |
| KIR2DL4 | HGNC:6332 | ENSG00000277750 | CHR_HSCHR19KIR_FH06_A_HAP_CTG3_1 | 54803572 | 54814476 | | 1 | ENSP00000478362 | Yes | |
| KIR2DS1 | HGNC:6333 | ENSG00000278120 | CHR_HSCHR19LRC_COX2_CTG3_1 | 54504993 | 54518921 | | 1 | ENSP00000483434 | Yes | |
| KLF9 | HGNC:1123 | ENSG00000119138 | 9 | 70384597 | 70414624 | q21.12 | 1 | ENSP00000366330 | Yes | |
| KLK10 | HGNC:6358 | ENSG00000129451 | 19 | 51012739 | 51020175 | q13.41 | 6 | ENSP00000311746 | Yes | |
| KLRC1 | HGNC:6374 | ENSG00000134545 | 12 | 10442264 | 10454685 | p13.2 | 7 | ENSP00000441432 | Yes | |
| KLRC3 | HGNC:6376 | ENSG00000205810 | 12 | 10412312 | 10420595 | p13.2 | 2 | ENSP00000379716 | Yes | |
| KLRC4 | HGNC:6377 | ENSG00000183542 | 12 | 10407382 | 10409757 | p13.2 | 1 | ENSP00000310216 | Yes | |
| KLRC4-KLRK1 | HGNC:48357 | ENSG00000255819 | 12 | 10372353 | 10410146 | p13.2 | 13 | ENSP00000456286 | Yes | Nonsense mediated decay/processed transcript |
| LAMA4 | HGNC:6484 | ENSG00000112769 | 6 | 112107931 | 112254939 | q21 | 24 | ENSP00000374114 | Yes | |
| LAMA5 | HGNC:6485 | ENSG00000130702 | 20 | 62307955 | 62367312 | q13.33 | 14 | ENSP00000252999 | | |
| LCMT1 | HGNC:17557 | ENSG00000205629 | 16 | 25111731 | 25178231 | p12.1 | 13 | ENSP00000382021 | Yes | |
| LEFTY2 | HGNC:3122 | ENSG00000143768 | 1 | 225936598 | 225941383 | q42.12 | 3 | ENSP00000355785 | Yes | |
| LEP | HGNC:6553 | ENSG00000174697 | 7 | 128241278 | 128257629 | q32.1 | 1 | ENSP00000312652 | Yes | |
| LEPR | HGNC:6554 | ENSG00000116678 | 1 | 65420652 | 65641559 | p31.3 | 8 | ENSP00000360098 | Yes | |
| LGALS14 | HGNC:30054 | ENSG00000006659 | 19 | 39704481 | 39709444 | q13.2 | 3 | ENSP00000375905 | Yes | |
| LGALS3 | HGNC:6563 | ENSG00000131981 | 14 | 55124110 | 55145423 | q22.3 | 7 | ENSP00000451526 | Yes | |
| LHCGR | HGNC:6585 | ENSG00000138039 | 2 | 48686774 | 48755730 | p16.3 | 6 | ENSP00000294954 | Yes | |
| LIF | HGNC:6596 | ENSG00000128342 | 22 | 30240453 | 30246759 | q12.2 | 2 | ENSP00000249075 | Yes | |
| LIFR | HGNC:6597 | ENSG00000113594 | 5 | 38474668 | 38608354 | p13.1 | 9 | ENSP00000263409 | Yes | |
| LPAR3 | HGNC:14298 | ENSG00000171517 | 1 | 84811602 | 84893206 | p22.3 | 3 | ENSP00000395389 | Yes | |
| LTA | HGNC:6709 | ENSG00000226979 | 6 | 31572054 | 31574324 | p21.33 | 4 | ENSP00000403495 | Yes | |
| MACF1 | HGNC:13664 | ENSG00000127603 | 1 | 39081316 | 39487177 | p34.3 | 36 | ENSP00000434859 | Yes | |

| | | | | | | | | | | |
|---|---|---|---|---|---|---|---|---|---|---|
| MAD1L1 | HGNC:6762 | ENSG00000002822 | 7 | 1815793 | 2233243 | p22.3 | 23 | ENSP00000385334 | Yes | |
| MAD2L1 | HGNC:6763 | ENSG00000164109 | 4 | 120055623 | 120066858 | q27 | 5 | ENSP00000296509 | Yes | |
| MAP2 | HGNC:6839 | ENSG00000078018 | 2 | 209424058 | 209734118 | q34 | 16 | ENSP00000199940 | Yes | |
| MAPRE3 | HGNC:6892 | ENSG00000084764 | 2 | 26970637 | 27027219 | p23.3 | 10 | ENSP00000233121 | Yes | |
| MBD4 | HGNC:6919 | ENSG00000129071 | 3 | 129430944 | 129440179 | q21.3 | 10 | ENSP00000394080 | Yes | |
| MBL2 | HGNC:6922 | ENSG00000165471 | 10 | 52765380 | 52771700 | q21.1 | 1 | ENSP00000363079 | Yes | |
| MCAM | HGNC:6934 | ENSG00000076706 | 11 | 119308529 | 119321521 | q23.3 | 17 | ENSP00000264036 | Yes | |
| MCL1 | HGNC:6943 | ENSG00000143384 | 1 | 150574551 | 150579738 | q21.2 | 4 | ENSP00000358022 | Yes | |
| MDM2 | HGNC:6973 | ENSG00000135679 | 12 | 68808177 | 68850686 | q15 | 37 | ENSP00000258149 | Yes | |
| MGAT2 | HGNC:7045 | ENSG00000168282 | 14 | 49620799 | 49623481 | q21.3 | 1 | ENSP00000307423 | Yes | |
| MGP | HGNC:7060 | ENSG00000111341 | 12 | 14880864 | 14885857 | p12.3 | 4 | ENSP00000445907 | Yes | |
| MICA | HGNC:7090 | ENSG00000183214 | CHR_HSCHR6_MHC_QBL_CTG1 | 31389818 | 31406373 | | 7 | ENSP00000383176 | Yes | |
| MIF | HGNC:7097 | ENSG00000240972 | 22 | 23894383 | 23895227 | q11.23 | 3 | ENSP00000215754 | Yes | |
| MIR125A | HGNC:31505 | ENSG00000208008 | 19 | 51693254 | 51693339 | q13.41 | 1 | | | miRNA |
| MIR146A | HGNC:31533 | ENSG00000283733 | 5 | 160485352 | 160485450 | q33.3 | 1 | | | miRNA |
| MIR149 | HGNC:31536 | ENSG00000207611 | 2 | 240456001 | 240456089 | q37.3 | 1 | | | miRNA |
| MIR196A2 | HGNC:31568 | ENSG00000207924 | 12 | 53991738 | 53991847 | q13.13 | 1 | | | miRNA |
| MIR3142HG | HGNC:51944 | ENSG00000253522 | 5 | 160438594 | 160487426 | q33.3 | 2 | | | miRNA |
| MIR4482 | HGNC:41792 | ENSG00000266852 | 10 | 104268336 | 104268405 | q25.1 | 1 | | | miRNA |
| MIR4761 | HGNC:41591 | ENSG00000284031 | 22 | 19963753 | 19963834 | q11.21 | 1 | | | miRNA |
| MIR4769 | HGNC:41694 | ENSG00000263858 | X | 47587429 | 47587505 | p11.3 | 1 | | | miRNA |
| MIR483 | HGNC:32340 | ENSG00000207805 | 11 | 2134134 | 2134209 | p15.5 | 1 | | | miRNA |
| MIR499A | HGNC:32133 | ENSG00000207635 | 20 | 34990376 | 34990497 | q11.22 | 1 | | | miRNA |
| MIR99B | HGNC:31651 | ENSG00000207550 | 19 | 51692612 | 51692681 | q13.41 | 1 | | | miRNA |
| MIRLET7E | HGNC:31482 | ENSG00000198972 | 19 | 51692786 | 51692864 | q13.41 | 1 | | | miRNA |
| MMP1 | HGNC:7155 | ENSG00000196611 | 11 | 102789919 | 102798160 | q22.2 | 1 | ENSP00000322788 | Yes | |
| MMP10 | HGNC:7156 | ENSG00000166670 | 11 | 102770502 | 102780628 | q22.2 | 2 | ENSP00000279441 | Yes | |
| MMP11 | HGNC:7157 | ENSG00000275365 | CHR_HSCHR22_1_CTG7 | 23768226 | 23784316 | | 13 | ENSP00000215743 | Yes | |
| MMP12 | HGNC:7158 | ENSG00000262406 | 11 | 102862736 | 102874982 | q22.2 | 1 | ENSP00000458585 | Yes | |
| MMP15 | HGNC:7161 | ENSG00000102996 | 16 | 58025754 | 58046901 | q21 | 2 | ENSP00000219271 | Yes | |
| MMP19 | HGNC:7165 | ENSG00000123342 | 12 | 55835433 | 55842966 | q13.2 | 9 | ENSP00000447363 | Yes | |
| MMP2 | HGNC:7166 | ENSG00000087245 | 16 | 55389700 | 55506691 | q12.2 | 8 | ENSP00000461421 | Yes | |

| | | | | | | | | | | |
|---|---|---|---|---|---|---|---|---|---|---|
| MMP26 | HGNC:14249 | ENSG00000167346 | 11 | 4704927 | 4992429 | p15.4 | 3 | ENSP00000369753 | Yes | |
| MMP7 | HGNC:7174 | ENSG00000137673 | 11 | 102520508 | 102530750 | q22.2 | 3 | ENSP00000260227 | Yes | |
| MMP9 | HGNC:7176 | ENSG00000100985 | 20 | 46008908 | 46016561 | q13.12 | 1 | ENSP00000361405 | Yes | |
| MPL | HGNC:7217 | ENSG00000117400 | 1 | 43337849 | 43352772 | p34.2 | 4 | ENSP00000361548 | Yes | |
| MRTFB | HGNC:29819 | ENSG00000186260 | 16 | 14071321 | 14266773 | p13.12 | 13 | ENSP00000458340 | Yes | |
| MSH4 | HGNC:7327 | ENSG00000057468 | 1 | 75796882 | 75913242 | p31.1 | 1 | ENSP00000263187 | Yes | |
| MSX1 | HGNC:7391 | ENSG00000163132 | 4 | 4859665 | 4863936 | p16.2 | 2 | ENSP00000372170 | Yes | |
| MTHFD1 | HGNC:7432 | ENSG00000100714 | 14 | 64388031 | 64463457 | q23.3 | 22 | ENSP00000438588 | Yes | |
| MTHFR | HGNC:7436 | ENSG00000177000 | 1 | 11785723 | 11806920 | p36.22 | 19 | ENSP00000365777 | Yes | |
| MTMR14 | HGNC:26190 | ENSG00000163719 | 3 | 9649433 | 9702393 | p25.3 | 13 | ENSP00000323462 | Yes | |
| MTR | HGNC:7468 | ENSG00000116984 | 1 | 236795292 | 236921278 | q43 | 9 | ENSP00000355536 | Yes | |
| MTRR | HGNC:7473 | ENSG00000124275 | 5 | 7851186 | 7906025 | p15.31 | 25 | ENSP00000402510 | Yes | |
| MUC1 | HGNC:7508 | ENSG00000185499 | 1 | 155185824 | 155192916 | q22 | 29 | ENSP00000483482 | Yes | |
| MUC4 | HGNC:7514 | ENSG00000278303 | CHR_HSCHR3_5_CTG3 | 195744993 | 195789727 | | 24 | ENSP00000487233 | Yes | |
| MUSK | HGNC:7525 | ENSG00000030304 | 9 | 110668779 | 110806558 | q31.3 | 7 | ENSP00000363571 | | long non coding RNA |
| MYBPC1 | HGNC:7549 | ENSG00000196091 | 12 | 101568353 | 101686028 | q23.2 | 23 | ENSP00000447404 | Yes | |
| MYH7B | HGNC:15906 | ENSG00000078814 | 20 | 34975403 | 35002437 | q11.22 | 9 | ENSP00000262873 | Yes | |
| MYOM1 | HGNC:7613 | ENSG00000101605 | 18 | 3066807 | 3220108 | p11.31 | 7 | ENSP00000348821 | Yes | |
| NAT2 | HGNC:7646 | ENSG00000156006 | 8 | 18391282 | 18401218 | p22 | 2 | ENSP00000286479 | Yes | |
| NAV2 | HGNC:15997 | ENSG00000166833 | 11 | 19350724 | 20121601 | p15.1 | 17 | ENSP00000353871 | Yes | |
| NBN | HGNC:7652 | ENSG00000104320 | 8 | 89933336 | 90003228 | q21.3 | 11 | ENSP00000265433 | | |
| NCAM1 | HGNC:7656 | ENSG00000149294 | 11 | 112961247 | 113278436 | q23.2 | 30 | ENSP00000480132 | Yes | |
| NCOA1 | HGNC:7668 | ENSG00000084676 | 2 | 24491914 | 24770702 | p23.3 | 11 | ENSP00000385097 | Yes | |
| NCOA2 | HGNC:7669 | ENSG00000140396 | 8 | 70109782 | 70403808 | q13.3 | 8 | ENSP00000399968 | Yes | |
| NCR1 | HGNC:6731 | ENSG00000275156 | CHR_HSCHR19_4_CTG3_1 | 54914255 | 54924255 | | 8 | ENSP00000481971 | Yes | |
| NCR2 | HGNC:6732 | ENSG00000096264 | 6 | 41335608 | 41350889 | p21.1 | 3 | ENSP00000362175 | Yes | |
| NCR3 | HGNC:19077 | ENSG00000204475 | 6 | 31588895 | 31593006 | p21.33 | 6 | ENSP00000365241 | Yes | |
| NDP | HGNC:7678 | ENSG00000124479 | X | 43948776 | 43973395 | p11.3 | 3 | ENSP00000495972 | Yes | |
| NDUFA13 | HGNC:17194 | ENSG00000186010 | 19 | 19515736 | 19529054 | p13.1 | 7 | ENSP00000423673 | Yes | |
| NDUFA6-DT | HGNC:45273 | ENSG00000237037 | 22 | 42090931 | 42137742 | q13.2 | 16 | | | RNA gene |
| NEU1 | HGNC:7758 | ENSG00000234343 | CHR_HSCHR6_MHC_DBB_CTG1 | 31839844 | 31845091 | | 5 | ENSP00000364782 | Yes | |

| | | | | | | | | | | |
|---|---|---|---|---|---|---|---|---|---|---|
| NF1 | HGNC:7765 | ENSG00000196712 | 17 | 31094927 | 31382116 | q11.2 | 23 | ENSP00000491589 | Yes | |
| NFE2L2 | HGNC:7782 | ENSG00000116044 | 2 | 177227595 | 177392697 | q31.2 | 14 | ENSP00000416308 | Yes | |
| NHLRC2 | HGNC:24731 | ENSG00000196865 | 10 | 113854661 | 113917194 | q25.3 | 2 | ENSP00000358307 | Yes | |
| NLRP10 | HGNC:21464 | ENSG00000182261 | 11 | 7957537 | 7965426 | p15.4 | 2 | ENSP00000327763 | Yes | |
| NLRP2 | HGNC:22948 | ENSG00000275399 | CHR_HSCHR19LRC_LRC_S_CTG3_1 | 54972989 | 55008845 | | 4 | ENSP00000479352 | Yes | |
| NLRP5 | HGNC:21269 | ENSG00000171487 | 19 | 55999726 | 56061810 | q13.43 | 3 | ENSP00000375063 | Yes | |
| NLRP7 | HGNC:22947 | ENSG00000167634 | 19 | 54923509 | 54966312 | q13.42 | 9 | ENSP00000467123 | Yes | |
| NOP14 | HGNC:16821 | ENSG00000087269 | 4 | 2937933 | 2963406 | p16.3 | 5 | ENSP00000405068 | Yes | |
| NOS3 | HGNC:7876 | ENSG00000164867 | 7 | 150991017 | 151014588 | q36.1 | 10 | ENSP00000297494 | Yes | |
| NR2F2 | HGNC:7976 | ENSG00000185551 | 15 | 96325938 | 96340263 | q26.2 | 5 | ENSP00000401674 | Yes | |
| NR3C1 | HGNC:7978 | ENSG00000113580 | 5 | 143277931 | 143435512 | q31.3 | 17 | ENSP00000377977 | Yes | |
| NTF6B | HGNC:8026 | ENSG00000267685 | 19 | 49038479 | 49039038 | q13.33 | 1 | | | Processed Pseudogene |
| OBSL1 | HGNC:29092 | ENSG00000124006 | 2 | 219550728 | 219571859 | q35 | 15 | ENSP00000385636 | Yes | |
| OFD1 | HGNC:2567 | ENSG00000046651 | X | 13734745 | 13769357 | p22.2 | 9 | ENSP00000344314 | Yes | |
| OGG1 | HGNC:8125 | ENSG00000114026 | 3 | 9749944 | 9788219 | p25.3 | 18 | ENSP00000305584 | Yes | |
| OSBPL5 | HGNC:16392 | ENSG00000021762 | 11 | 3087107 | 3166739 | p15.4 | 19 | ENSP00000437141 | Yes | |
| PADI6 | HGNC:20449 | ENSG00000276747 | 1 | 17372196 | 17401699 | p36.13 | 1 | ENSP00000483125 | Yes | |
| PAEP | HGNC:8573 | ENSG00000122133 | 9 | 135561756 | 135566955 | q34.3 | 9 | ENSP00000431712 | Yes | |
| PAPPA | HGNC:8602 | ENSG00000182752 | 9 | 116153791 | 116402321 | q33.1 | 3 | ENSP00000330658 | Yes | |
| PARG | HGNC:8605 | ENSG00000227345 | 10 | 49818279 | 49970203 | q11.23 | 7 | ENSP00000384408 | Yes | |
| PCCB | HGNC:8654 | ENSG00000114054 | 3 | 136250340 | 136337896 | q22.3 | 19 | ENSP00000251654 | Yes | |
| PCDHA3 | HGNC:8669 | ENSG00000255408 | 5 | 140801028 | 141012347 | q31.3 | 2 | ENSP00000434086 | Yes | |
| PCYT1A | HGNC:8754 | ENSG00000161217 | 3 | 196214222 | 196287957 | q29 | 16 | ENSP00000392397 | Yes | |
| PDE2A | HGNC:8777 | ENSG00000186642 | 11 | 72576141 | 72674591 | q13.4 | 27 | ENSP00000334910 | Yes | |
| PDE8B | HGNC:8794 | ENSG00000113231 | 5 | 77210449 | 77429807 | q13.3 | 9 | ENSP00000425720 | Yes | |
| PDSS1P1 | HGNC:49740 | ENSG00000182347 | 9 | 5084999 | 5086112 | p24.1 | 1 | | | Processed Pseudogene |
| PDZD2 | HGNC:18486 | ENSG00000133401 | 5 | 31639131 | 32110932 | p13.3 | 12 | ENSP00000402033 | Yes | |
| PER1 | HGNC:8845 | ENSG00000179094 | 17 | 8140472 | 8156506 | p13.1 | 19 | ENSP00000314420 | Yes | |
| PES1 | HGNC:8848 | ENSG00000100029 | 22 | 30576625 | 30607083 | q12.2 | 13 | ENSP00000346725 | Yes | |
| PGF | HGNC:8893 | ENSG00000119630 | 14 | 74941834 | 74955626 | q24.3 | 8 | ENSP00000451040 | Yes | |
| PGM1 | HGNC:8905 | ENSG00000079739 | 1 | 63593411 | 63660245 | p31.3 | 6 | ENSP00000360125 | Yes | |
| PGR | HGNC:8910 | ENSG00000082175 | 11 | 101029624 | 101129813 | q22.1 | 11 | ENSP00000325120 | Yes | |

| | | | | | | | | | | |
|---|---|---|---|---|---|---|---|---|---|---|
| PGR-AS1 | HGNC:52650 | ENSG00000282728 | 11 | 101129077 | 101209591 | q22.1 | 2 | | | RNA gene |
| PLA2G4A | HGNC:9035 | ENSG00000116711 | 1 | 186828949 | 186988981 | q31.1 | 2 | ENSP00000356436 | Yes | |
| PLAU | HGNC:9052 | ENSG00000122861 | 10 | 73909177 | 73917496 | q22.2 | 5 | ENSP00000474318 | Yes | |
| PLAUR | HGNC:9053 | ENSG00000011422 | 19 | 43646095 | 43670547 | q13.31 | 16 | ENSP00000342049 | Yes | |
| PLCD4 | HGNC:9062 | ENSG00000115556 | 2 | 218607855 | 218637184 | q35 | 15 | ENSP00000388631 | Yes | |
| PLG | HGNC:9071 | ENSG00000122194 | 6 | 160702238 | 160753315 | q26 | 12 | ENSP00000355891 | Yes | |
| PNPT1 | HGNC:23166 | ENSG00000138035 | 2 | 55634061 | 55693863 | p16.1 | 7 | ENSP00000400646 | | |
| POLR1C | HGNC:20194 | ENSG00000171453 | 6 | 43509702 | 43562419 | p21.1 | 16 | ENSP00000395401 | Yes | |
| POMC | HGNC:9201 | ENSG00000115138 | 2 | 25160853 | 25168903 | p23.3 | 5 | ENSP00000384092 | Yes | |
| PPARG | HGNC:9236 | ENSG00000132170 | 3 | 12287368 | 12434356 | p25.2 | 25 | ENSP00000380205 | Yes | |
| PRKDC | HGNC:9413 | ENSG00000253729 | 8 | 47773108 | 47960183 | q11.21 | 11 | ENSP00000313420 | Yes | |
| PRL | HGNC:9445 | ENSG00000172179 | 6 | 22287244 | 22302826 | p22.3 | 5 | ENSP00000302150 | Yes | |
| PRLR | HGNC:9446 | ENSG00000113494 | 5 | 35048756 | 35230487 | p13.2 | 20 | ENSP00000231423 | Yes | |
| PROC | HGNC:9451 | ENSG00000115718 | 2 | 127418427 | 127429246 | q14.3 | 10 | ENSP00000234071 | Yes | |
| PROCR | HGNC:9452 | ENSG00000101000 | 20 | 35172072 | 35216240 | q11.22 | 3 | ENSP00000216968 | Yes | |
| PROK1 | HGNC:18454 | ENSG00000143125 | 1 | 110451149 | 110457358 | p13.3 | 1 | ENSP00000271331 | Yes | |
| PROK1 | HGNC:18454 | ENSG00000143125 | 1 | 110451149 | 110457358 | p13.3 | 1 | ENSP00000271331 | Yes | |
| PROKR1 | HGNC:4524 | ENSG00000169618 | 2 | 68643589 | 68658247 | p13.3 | 1 | ENSP00000303775 | Yes | |
| PROKR2 | HGNC:15836 | ENSG00000101292 | 20 | 5302040 | 5314369 | p12.3 | 1 | ENSP00000217270 | Yes | |
| PROS1 | HGNC:9456 | ENSG00000184500 | 3 | 93873033 | 93980003 | q11.1 | 12 | ENSP00000377783 | Yes | |
| PROZ | HGNC:9460 | ENSG00000126231 | 13 | 113158648 | 113172386 | q34 | 3 | ENSP00000364697 | Yes | |
| PTEN | HGNC:9588 | ENSG00000171862 | 10 | 87863625 | 87971930 | q23.31 | 6 | ENSP00000361021 | Yes | |
| PTGFR | HGNC:9600 | ENSG00000122420 | 1 | 78303884 | 78540701 | p31.1 | 4 | ENSP00000359794 | Yes | |
| PTGIS | HGNC:9603 | ENSG00000124212 | 20 | 49503874 | 49568137 | q13.13 | 2 | ENSP00000244043 | Yes | |
| PTGS1 | HGNC:9604 | ENSG00000095303 | 9 | 122370530 | 122395703 | q33.2 | 10 | ENSP00000494717 | Yes | |
| PTGS2 | HGNC:9605 | ENSG00000073756 | 1 | 186671791 | 186680423 | q31.1 | 5 | ENSP00000356438 | Yes | |
| PTHLH | HGNC:9607 | ENSG00000087494 | 12 | 27958084 | 27972733 | p11.22 | 9 | ENSP00000441765 | Yes | |
| PTPN11 | HGNC:9644 | ENSG00000179295 | 12 | 112418351 | 112509913 | q24.13 | 7 | ENSP00000376376 | Yes | |
| PTX3 | HGNC:9692 | ENSG00000163661 | 3 | 157436850 | 157443633 | q25.32 | 1 | ENSP00000295927 | Yes | |
| PZP | HGNC:9750 | ENSG00000126838 | 12 | 9148840 | 9208395 | p13.31 | 7 | ENSP00000261336 | Yes | |
| RAD9B | HGNC:21700 | ENSG00000151164 | 12 | 110501655 | 110532086 | q24.11 | 9 | ENSP00000387329 | | |
| RAG2 | HGNC:9832 | ENSG00000175097 | 11 | 36575574 | 36598279 | p12 | 9 | ENSP00000308620 | Yes | |
| RAMP1 | HGNC:9843 | ENSG00000132329 | 2 | 237858893 | 237912106 | q37.3 | 4 | ENSP00000384688 | Yes | |
| RAN | HGNC:9846 | ENSG00000132341 | 12 | 130872037 | 130877678 | q24.33 | 12 | ENSP00000446215 | Yes | |

| | | | | | | | | | | |
|---|---|---|---|---|---|---|---|---|---|---|
| RBP4 | HGNC:9922 | ENSG00000138207 | 10 | 93591687 | 93601744 | q23.33 | 4 | ENSP00000360519 | Yes | |
| RERE | HGNC:9965 | ENSG00000142599 | 1 | 8352397 | 8848921 | p36.23 | 20 | ENSP00000338629 | Yes | |
| RETN | HGNC:20389 | ENSG00000104918 | 19 | 7669049 | 7670455 | p13.2 | 3 | ENSP00000221515 | Yes | |
| RGS2 | HGNC:9998 | ENSG00000116741 | 1 | 192809039 | 192812275 | q31.2 | 4 | ENSP00000235382 | Yes | |
| RHOB | HGNC:668 | ENSG00000143878 | 2 | 20447074 | 20449440 | p24.1 | 1 | ENSP00000272233 | Yes | |
| RMDN2 | HGNC:26567 | ENSG00000115841 | 2 | 37923187 | 38067142 | p22.2 | 12 | ENSP00000393705 | Yes | |
| RMDN2-AS1 | HGNC:41150 | ENSG00000235848 | 2 | 37949911 | 38067041 | p22.2 | 14 | | | RNA gene |
| RNA5SP67 | HGNC:42844 | ENSG00000252231 | 1 | 173921070 | 173921200 | q25.1 | 1 | | | rRNA Pseudogene |
| RPS27AP19 | HGNC:52298 | ENSG00000269228 | 19 | 7665717 | 7666476 | p13.2 | 1 | | | Processed Pseudogene |
| RPS6KA3 | HGNC:10432 | ENSG00000177189 | X | 20149911 | 20267519 | p22.12 | 16 | ENSP00000368884 | Yes | |
| RYK | HGNC:10481 | ENSG00000163785 | 3 | 134065303 | 134250744 | q22.2 | 9 | ENSP00000478721 | | |
| RYR1 | HGNC:10483 | ENSG00000196218 | 19 | 38433699 | 38587564 | q13.2 | 10 | ENSP00000347667 | Yes | |
| S1PR3 | HGNC:3167 | ENSG00000213694 | 9 | 88990863 | 89005155 | q22.1 | 7 | ENSP00000365006 | Yes | |
| SAT2 | HGNC:23160 | ENSG00000141504 | 17 | 7626234 | 7627876 | p13.1 | 14 | ENSP00000269298 | Yes | |
| SBNO2 | HGNC:29158 | ENSG00000278788 | CHR_HSCHR19_4_CTG2 | 1107635 | 1156979 | | 11 | ENSP00000488808 | Yes | |
| SCN10A | HGNC:10582 | ENSG00000185313 | 3 | 38696802 | 38816286 | p22.2 | 5 | ENSP00000390600 | | |
| SDC2 | HGNC:10659 | ENSG00000169439 | 8 | 96493813 | 96611790 | q22.1 | 8 | ENSP00000307046 | Yes | |
| SDF2L1 | HGNC:10676 | ENSG00000128228 | 22 | 21642302 | 21644299 | q11.21 | 2 | ENSP00000248958 | Yes | |
| SDHB | HGNC:10681 | ENSG00000117118 | 1 | 17018722 | 17054170 | p36.13 | 8 | ENSP00000364649 | Yes | |
| SELL | HGNC:10720 | ENSG00000188404 | 1 | 169690665 | 169711702 | q24.2 | 7 | ENSP00000236147 | Yes | |
| SELP | HGNC:10721 | ENSG00000174175 | 1 | 169588849 | 169630193 | q24.2 | 7 | ENSP00000391694 | Yes | |
| SERPINA10 | HGNC:15996 | ENSG00000278767 | CHR_HSCHR14_7_CTG1 | 94280460 | 94293271 | | 2 | ENSP00000484632 | Yes | |
| SERPINA6 | HGNC:1540 | ENSG00000277405 | CHR_HSCHR14_7_CTG1 | 94304248 | 94323394 | | 3 | ENSP00000486588 | Yes | |
| SERPINB10 | HGNC:8942 | ENSG00000242550 | 18 | 63897174 | 63936111 | q21.33 | 4 | ENSP00000381082 | Yes | |
| SERPINB2 | HGNC:8584 | ENSG00000197632 | 18 | 63871692 | 63903888 | q21.33 | 6 | ENSP00000385397 | Yes | |
| SERPINB3 | HGNC:10569 | ENSG00000057149 | 18 | 63655197 | 63661893 | q21.33 | 2 | ENSP00000283752 | Yes | |
| SERPINB4 | HGNC:10570 | ENSG00000206073 | 18 | 63637259 | 63644256 | q21.33 | 4 | ENSP00000343445 | Yes | |
| SERPINC1 | HGNC:775 | ENSG00000117601 | 1 | 173903804 | 173917378 | q25.1 | 4 | ENSP00000356671 | Yes | |
| SERPINE1 | HGNC:8583 | ENSG00000106366 | 7 | 101127089 | 101139266 | q22.1 | 1 | ENSP00000223095 | Yes | |
| SHBG | HGNC:10839 | ENSG00000129214 | 17 | 7613946 | 7633382 | p13.1 | 21 | ENSP00000345675 | Yes | |
| SIK1 | HGNC:11142 | ENSG00000142178 | 21 | 43414483 | 43427131 | q22.3 | 6 | ENSP00000270162 | Yes | |

| | | | | | | | | | | |
|---|---|---|---|---|---|---|---|---|---|---|
| SLC13A1 | HGNC:10916 | ENSG00000081800 | 7 | 123113531 | 123199972 | q31.32 | 4 | ENSP00000194130 | Yes | |
| SLC19A1 | HGNC:10937 | ENSG00000173638 | 21 | 45493572 | 45573365 | q22.3 | 14 | ENSP00000393988 | Yes | |
| SLC2A1 | HGNC:11005 | ENSG00000117394 | 1 | 42925375 | 42958868 | p34.2 | 10 | ENSP00000486694 | Yes | |
| SLC31A1 | HGNC:11016 | ENSG00000136868 | 9 | 113221544 | 113264492 | q32 | 2 | ENSP00000363329 | Yes | |
| SLCO2A1 | HGNC:10955 | ENSG00000174640 | 3 | 133928145 | 134052184 | q22.2 | 9 | ENSP00000311291 | Yes | |
| SMAD2 | HGNC:6768 | ENSG00000175387 | 18 | 47808957 | 47931146 | q21.1 | 12 | ENSP00000262160 | Yes | |
| SMAD4 | HGNC:6770 | ENSG00000141646 | 18 | 51028394 | 51085045 | q21.2 | 17 | ENSP00000465878 | Yes | |
| SMARCC2 | HGNC:11105 | ENSG00000139613 | 12 | 56162359 | 56189567 | q13.2 | 15 | ENSP00000377591 | Yes | |
| SNED1 | HGNC:24696 | ENSG00000162804 | 2 | 240998618 | 241095568 | q37.3 | 12 | ENSP00000384871 | Yes | |
| SNRPN | HGNC:11164 | ENSG00000128739 | 15 | 24823637 | 24978723 | q11.2 | 12 | ENSP00000494831 | Yes | |
| SOD2 | HGNC:11180 | ENSG00000112096 | 6 | 159669069 | 159745186 | q25.3 | 14 | ENSP00000446252 | Yes | |
| SOX4 | HGNC:11200 | ENSG00000124766 | 6 | 21593751 | 21598619 | p22.3 | 1 | ENSP00000244745 | | |
| SPACA6 | HGNC:27113 | ENSG00000182310 | 19 | 51689128 | 51712387 | q13.41 | 14 | ENSP00000496692 | Yes | |
| SPAG5 | HGNC:13452 | ENSG00000076382 | 17 | 28577565 | 28599025 | q11.2 | 16 | ENSP00000323300 | Yes | |
| SPP1 | HGNC:11255 | ENSG00000118785 | 4 | 87975650 | 87983426 | q22.1 | 11 | ENSP00000237623 | Yes | |
| SRC | HGNC:11283 | ENSG00000197122 | 20 | 37344685 | 37406050 | q11.23 | 11 | ENSP00000362680 | Yes | |
| SSFA2 | | ENSG00000260742 | 2 | 181887851 | 181891663 | | 1 | | | RNA gene/Long non coding RNA gene |
| SST | HGNC:11329 | ENSG00000157005 | 3 | 187668912 | 187670394 | q27.3 | 1 | ENSP00000287641 | Yes | |
| STAG3 | HGNC:11356 | ENSG00000066923 | 7 | 100177563 | 100221488 | q22.1 | 19 | ENSP00000400359 | Yes | |
| STAT3 | HGNC:11364 | ENSG00000168610 | 17 | 42313324 | 42388568 | q21.2 | 14 | ENSP00000264657 | Yes | |
| STAT5A | HGNC:11366 | ENSG00000126561 | 17 | 42287547 | 42311943 | q21.2 | 12 | ENSP00000341208 | Yes | |
| STAT5B | HGNC:11367 | ENSG00000173757 | 17 | 42199168 | 42276707 | q21.2 | 7 | ENSP00000293328 | Yes | |
| STIL | HGNC:10879 | ENSG00000123473 | 1 | 47250139 | 47314147 | p33 | 9 | ENSP00000353544 | Yes | |
| STON1-GTF2A1L | HGNC:30651 | ENSG00000068781 | 2 | 48529925 | 48776517 | p16.3 | 6 | ENSP00000385701 | Yes | |
| SULF1 | HGNC:20391 | ENSG00000137573 | 8 | 69466624 | 69660915 | q13.2 | 23 | ENSP00000436608 | Yes | |
| SYCP3 | HGNC:18130 | ENSG00000139351 | 12 | 101728648 | 101739472 | q23.2 | 5 | ENSP00000376655 | Yes | |
| SYN1 | HGNC:11494 | ENSG00000008056 | X | 47571901 | 47619857 | p11.23 | 5 | ENSP00000295987 | Yes | |
| SYN2 | HGNC:11495 | ENSG00000157152 | 3 | 12004388 | 12192032 | p25.2 | 6 | ENSP00000480050 | Yes | |
| SYN3 | HGNC:11496 | ENSG00000185666 | 22 | 32512552 | 33058372 | q12.3 | 13 | ENSP00000351614 | Yes | |
| TCN2 | HGNC:11653 | ENSG00000185339 | 22 | 30607003 | 30627271 | q12.2 | 7 | ENSP00000215838 | Yes | |
| TCTN3 | HGNC:24519 | ENSG00000119977 | 10 | 95663396 | 95694143 | q24.1 | 7 | ENSP00000360261 | Yes | |
| TEK | HGNC:11724 | ENSG00000120156 | 9 | 27109141 | 27230174 | p21.2 | 5 | ENSP00000430686 | Yes | |
| TEX12 | HGNC:11734 | ENSG00000150783 | 11 | 112167372 | 112172556 | q23.1 | 2 | ENSP00000436893 | Yes | |

| | | | | | | | | | | |
|---|---|---|---|---|---|---|---|---|---|---|
| TFPI | HGNC:11760 | ENSG00000003436 | 2 | 187464230 | 187565760 | q32.1 | 12 | ENSP00000376172 | Yes | |
| TFRC | HGNC:11763 | ENSG00000072274 | 3 | 196027183 | 196082096 | q29 | 15 | ENSP00000414015 | Yes | |
| TGFB1 | HGNC:11766 | ENSG00000105329 | 19 | 41301587 | 41353922 | q13.2 | 4 | ENSP00000472767 | Yes | |
| TGFBI | HGNC:11771 | ENSG00000120708 | 5 | 136028988 | 136063818 | q31.1 | 15 | ENSP00000416330 | Yes | |
| TGFBR1 | HGNC:11772 | ENSG00000106799 | 9 | 99104038 | 99154192 | q22.33 | 10 | ENSP00000449934 | Yes | |
| THBD | HGNC:11784 | ENSG00000178726 | 20 | 23045633 | 23049741 | p11.21 | 1 | ENSP00000366307 | Yes | |
| THPO | HGNC:11795 | ENSG00000090534 | 3 | 184371935 | 184381968 | q27.1 | 8 | ENSP00000494504 | Yes | |
| THRB | HGNC:11799 | ENSG00000151090 | 3 | 24117153 | 24495756 | p24.2 | 19 | ENSP00000379904 | Yes | |
| TIMELESS | HGNC:11813 | ENSG00000111602 | 12 | 56416363 | 56449426 | q13.3 | 5 | ENSP00000450607 | | |
| TIMP1 | HGNC:11820 | ENSG00000102265 | X | 47582408 | 47586789 | p11.3 | 5 | ENSP00000218388 | Yes | |
| TIMP2 | HGNC:11821 | ENSG00000035862 | 17 | 78852977 | 78925387 | q25.3 | 5 | ENSP00000467584 | Yes | |
| TIMP3 | HGNC:11822 | ENSG00000100234 | 22 | 32801701 | 32863043 | q12.3 | 1 | ENSP00000266085 | Yes | |
| TIMP4 | HGNC:11823 | ENSG00000157150 | 3 | 12153068 | 12158912 | p25.2 | 1 | ENSP00000287814 | Yes | |
| TLE6 | HGNC:30788 | ENSG00000104953 | 19 | 2977538 | 2995179 | p13.3 | 10 | ENSP00000246112 | Yes | |
| TLR3 | HGNC:11849 | ENSG00000164342 | 4 | 186069152 | 186088069 | q35.1 | 5 | ENSP00000296795 | Yes | |
| TMEM91 | HGNC:32393 | ENSG00000142046 | 19 | 41350911 | 41384083 | q13.2 | 14 | ENSP00000441900 | Yes | |
| TNC | HGNC:5318 | ENSG00000041982 | 9 | 115019575 | 115118257 | q33.1 | 14 | ENSP00000265131 | Yes | |
| TNF | HGNC:11892 | ENSG00000232810 | 6 | 31575565 | 31578336 | p21.33 | 1 | ENSP00000398698 | Yes | |
| TNFRSF1A | HGNC:11916 | ENSG00000067182 | 12 | 6328757 | 6342114 | p13.31 | 16 | ENSP00000441803 | Yes | |
| TNFSF10 | HGNC:11925 | ENSG00000121858 | 3 | 172505508 | 172523475 | q26.31 | 6 | ENSP00000241261 | Yes | |
| TNFSF13 | HGNC:11928 | ENSG00000161955 | 17 | 7558292 | 7561608 | p13.1 | 9 | ENSP00000464998 | Yes | |
| TNFSF15 | HGNC:11931 | ENSG00000181634 | 9 | 114784652 | 114806039 | q32 | 2 | ENSP00000363157 | Yes | |
| TNFSF8 | HGNC:11938 | ENSG00000106952 | 9 | 114893343 | 114930595 | q33.1 | 3 | ENSP00000223795 | Yes | |
| TNR | HGNC:11953 | ENSG00000116147 | 1 | 175315194 | 175743616 | q25.1 | 3 | ENSP00000356646 | Yes | |
| TP53 | HGNC:11998 | ENSG00000141510 | 17 | 7661779 | 7687550 | p13.1 | 29 | ENSP00000410739 | Yes | |
| TP73 | HGNC:12003 | ENSG00000078900 | 1 | 3652516 | 3736201 | p36.32 | 14 | ENSP00000367545 | Yes | |
| TP73-AS1 | HGNC:29052 | ENSG00000227372 | 1 | 3735511 | 3747373 | p36.32 | 29 | | | RNA gene |
| TPH1 | HGNC:12008 | ENSG00000129167 | 11 | 18017564 | 18042426 | p15.1 | 4 | ENSP00000250018 | Yes | |
| TRAF1 | HGNC:12031 | ENSG00000056558 | 9 | 120902393 | 120929173 | q33.2 | 3 | ENSP00000362994 | Yes | |
| TRAF3IP1 | HGNC:17861 | ENSG00000204104 | 2 | 238320441 | 238400897 | q37.3 | 5 | ENSP00000375851 | Yes | |
| TRO | HGNC:12326 | ENSG00000067445 | X | 54920462 | 54931431 | p11.21 | 25 | ENSP00000388947 | Yes | |
| TSHR | HGNC:12373 | ENSG00000165409 | 14 | 80954989 | 81146302 | q31.1 | 12 | ENSP00000450549 | Yes | |
| TYMP | HGNC:3148 | ENSG00000025708 | 22 | 50525752 | 50530032 | q13.33 | 17 | ENSP00000498433 | Yes | |
| TYMS | HGNC:12441 | ENSG00000176890 | 18 | 657653 | 673578 | p11.32 | 6 | ENSP00000315644 | Yes | |

| | | | | | | | | | | |
|---|---|---|---|---|---|---|---|---|---|---|
| TYROBP | HGNC:12449 | ENSG00000011600 | 19 | 35904401 | 35908295 | q13.12 | 9 | ENSP00000465081 | Yes | |
| UBE2N | HGNC:12492 | ENSG00000177889 | 12 | 93405673 | 93441947 | q22 | 6 | ENSP00000316176 | Yes | |
| UBN1 | HGNC:12506 | ENSG00000118900 | 16 | 4846665 | 4882401 | p13.3 | 9 | ENSP00000262376 | Yes | |
| ULBP1 | HGNC:14893 | ENSG00000111981 | 6 | 149963943 | 149973715 | q25.1 | 1 | ENSP00000229708 | Yes | |
| UTRN | HGNC:12635 | ENSG00000152818 | 6 | 144285701 | 144853034 | q24.2 | 13 | ENSP00000356515 | Yes | |
| VCAM1 | HGNC:12663 | ENSG00000162692 | 1 | 100719742 | 100739045 | p21.2 | 6 | ENSP00000359137 | Yes | |
| VEGFA | HGNC:12680 | ENSG00000112715 | 6 | 43770184 | 43786487 | p21.1 | 33 | ENSP00000361137 | Yes | |
| WDR60 | HGNC:12681 | ENSG00000126870 | 7 | 158856558 | 158956747 | q36.3 | 5 | ENSP00000384290 | Yes | |
| WNT4 | HGNC:12682 | ENSG00000162552 | 1 | 22117313 | 22143969 | p36.12 | 3 | ENSP00000290167 | Yes | |
| WNT5A | HGNC:12683 | ENSG00000114251 | 3 | 55465715 | 55490539 | p14.3 | 7 | ENSP00000417310 | Yes | |
| WNT6 | HGNC:12684 | ENSG00000115596 | 2 | 218859805 | 218874233 | q35 | 2 | ENSP00000233948 | Yes | |
| WNT7A | HGNC:12685 | ENSG00000154764 | 3 | 13816258 | 13880071 | p25.1 | 3 | ENSP00000285018 | Yes | |
| WRN | HGNC:12686 | ENSG00000165392 | 8 | 31033788 | 31175916 | p12 | 6 | ENSP00000498593 | Yes | |
| WTAPP1 | HGNC:12687 | ENSG00000255282 | 11 | 102746968 | 102836766 | q22.2 | 5 | | | Pseudogene |
| XIST | HGNC:12688 | ENSG00000229807 | X | 73820649 | 73852723 | q13.2 | 30 | | | RNA gene |
| XPO5 | HGNC:12689 | ENSG00000124571 | 6 | 43522334 | 43576038 | p21.1 | 12 | ENSP00000265351 | Yes | |
| YES1 | HGNC:12690 | ENSG00000176105 | 18 | 721588 | 812546 | p11.32 | 5 | ENSP00000464380 | Yes | |
| ZC3H13 | HGNC:12691 | ENSG00000123200 | 13 | 45954465 | 46052759 | q14.13 | 5 | ENSP00000242848 | Yes | |
| ZEB1 | HGNC:12692 | ENSG00000148516 | 10 | 31318495 | 31529814 | p11.22 | 25 | ENSP00000452787 | Yes | |
| ZNF628 | HGNC:12693 | ENSG00000197483 | 19 | 55476617 | 55484487 | q13.42 | 3 | ENSP00000469591 | Yes | |

### Supplementary Table 2 – Main results, list of prioritized variants

| SYMBOL | OMIM | Uploaded_variation | Gene | Feature | Consequence | CDNA position | CDS position | Protein position | Amino acids | Codons | Existing variation | IMPACT | SIFT | PolyPhen | ID | ALT count |
|---|---|---|---|---|---|---|---|---|---|---|---|---|---|---|---|---|
| ABCA2 | 600047 | chr9_137014012_C/A | ENSG00000107331 | ENST00000341511 | missense_variant | 4364 | 4267 | 1423 | V/F | Gtc/Ttc | rs147917446,CM1515218 | MODERATE | tolerated (0.15) | benign (0.163) | FE130 | 1 |
| ABCA2 | 600047 | chr9_137014012_C/A | ENSG00000107331 | ENST00000341511 | missense_variant | 4364 | 4267 | 1423 | V/F | Gtc/Ttc | rs147917446,CM1515218 | MODERATE | tolerated (0.15) | benign (0.163) | FE194 | 1 |
| ABCF2 | 612510 | chr7_151214934_T/C | ENSG00000285292 | ENST00000222388 | missense_variant | 1725 | 1679 | 560 | N/S | aAt/aGt | rs55866446 | MODERATE | deleterious (0.01) | benign (0.391) | FE193 | 1 |
| AC008695.1 | NA | chr5_131431342_G/C | ENSG00000273217 | ENST00000514667 | missense_variant | 4194 | 4132 | 1378 | L/V | Ctc/Gtc | rs34896346 | MODERATE | tolerated_low_confidence (0.48) | benign (0) | FE165 | 1 |
| ACSL4 | 300157 | chrX_109659421_A/G | ENSG00000068366 | ENST00000514500 | stop_lost,NMD_transcript_variant | 468 | 469 | 157 | */R | Tga/Cga | rs780359191 | HIGH | - | - | FE193 | 1 |
| ACVR1 | 102576 | chr2_157799450_G/C | ENSG00000115170 | ENST00000539637 | missense_variant | 390 | 44 | 15 | A/G | gCt/gGt | rs13406336 | MODERATE | deleterious_low_confidence (0.04) | benign (0.007) | FE187 | 1 |
| ACVR1 | 102576 | chr2_157799450_G/C | ENSG00000115170 | ENST00000440523 | missense_variant | 318 | 44 | 15 | A/G | gCt/gGt | rs13406336 | MODERATE | tolerated (0.18) | benign (0.007) | FE187 | 1 |
| ACVR1 | 102576 | chr2_157799450_G/C | ENSG00000115170 | ENST00000434821 | missense_variant | 761 | 44 | 15 | A/G | gCt/gGt | rs13406336 | MODERATE | deleterious_low_confidence (0.04) | benign (0.007) | FE187 | 1 |
| ACVR1 | 102576 | chr2_157799450_G/C | ENSG00000115170 | ENST00000672582 | missense_variant | 398 | 44 | 15 | A/G | gCt/gGt | rs13406336 | MODERATE | deleterious_low_confidence (0.04) | benign (0.007) | FE187 | 1 |
| ACVR1 | 102576 | chr2_157799450_G/C | ENSG00000115170 | ENST00000263640 | missense_variant | 474 | 44 | 15 | A/G | gCt/gGt | rs13406336 | MODERATE | deleterious_low_confidence (0.04) | benign (0.007) | FE187 | 1 |
| ACVR1 | 102576 | chr2_157799450_G/C | ENSG00000115170 | ENST00000673324 | missense_variant | 398 | 44 | 15 | A/G | gCt/gGt | rs13406336 | MODERATE | deleterious_low_confidence (0.04) | benign (0.007) | FE187 | 1 |
| ACVR1 | 102576 | chr2_157799450_G/C | ENSG00000115170 | ENST00000410057 | missense_variant | 383 | 44 | 15 | A/G | gCt/gGt | rs13406336 | MODERATE | deleterious_low_confidence (0.04) | benign (0.007) | FE187 | 1 |
| ACVR1 | 102576 | chr2_157799450_G/C | ENSG00000115170 | ENST00000412025 | missense_variant | 313 | 44 | 15 | A/G | gCt/gGt | rs13406336 | MODERATE | tolerated (0.18) | benign (0.007) | FE187 | 1 |
| ACVR1 | 102576 | chr2_157799450_G/C | ENSG00000115170 | ENST00000413751 | missense_variant | 515 | 44 | 15 | A/G | gCt/gGt | rs13406336 | MODERATE | - | benign (0.007) | FE187 | 1 |
| ACVR1 | 102576 | chr2_157799450_G/C | ENSG00000115170 | ENST00000409283 | missense_variant | 311 | 44 | 15 | A/G | gCt/gGt | rs13406336 | MODERATE | deleterious_low_confidence (0.04) | benign (0.007) | FE187 | 1 |
| ACVR1 | 102576 | chr2_157799450_G/C | ENSG00000115170 | ENST00000424669 | missense_variant | 380 | 44 | 15 | A/G | gCt/gGt | rs13406336 | MODERATE | deleterious_low_confidence (0.04) | benign (0.007) | FE187 | 1 |
| ADAM22 | 603709 | chr7_88075657_C/T | ENSG00000008277 | ENST00000265727 | missense_variant | 434 | 355 | 119 | H/Y | Cac/Tac | rs4728730 | MODERATE | tolerated (0.08) | benign (0.279) | FE193 | 1 |
| ADAMTS2 | 604539 | chr5_179114310_C/G | ENSG00000087116 | ENST00000251582 | missense_variant | 3326 | 3193 | 1065 | G/R | Ggc/Cgc | rs747348237 | MODERATE | deleterious (0.04) | probably_damaging (0.956) | FE193 | 1 |
| AFDN | 159559 | chr6_167896967_A/G | ENSG00000130396 | ENST00000392108 | missense_variant | 1454 | 1312 | 438 | I/V | Atc/Gtc | rs35140809 | MODERATE | deleterious (0.01) | possibly_damaging (0.685) | FE194 | 1 |
| Aug4 | 607356 | chr1_35850212_A/G | ENSG00000134698 | ENST00000373210 | missense_variant | 2632 | 2231 | 744 | H/R | cAt/cGt | rs1200837254 | MODERATE | deleterious (0) | probably_damaging (0.931) | FE187 | 1 |
| AHCY | 180960 | chr20_34295502_G/A | ENSG00000101444 | ENST00000217426 | missense_variant | 197 | 112 | 38 | R/W | Cgg/Tgg | rs13043752,CM096334 | MODERATE | deleterious_low_confidence (0) | possibly_damaging (0.673) | FE130 | 1 |
| ALAS1 | 125290 | chr3_52208160_G/A | ENSG00000023330 | ENST00000469224 | missense_variant | 1445 | 1243 | 415 | A/T | Gca/Aca | rs371320771 | MODERATE | deleterious (0) | probably_damaging (0.992) | FE193 | 1 |
| ALAS1 | 125290 | chr3_52208160_G/A | ENSG00000023330 | ENST00000484952 | missense_variant | 1548 | 1243 | 415 | A/T | Gca/Aca | rs371320771 | MODERATE | deleterious (0) | probably_damaging (0.992) | FE193 | 1 |
| ALAS1 | 125290 | chr3_52208160_G/A | ENSG00000023330 | ENST00000310271 | missense_variant | 1388 | 1243 | 415 | A/T | Gca/Aca | rs371320771 | MODERATE | deleterious (0) | probably_damaging (0.992) | FE193 | 1 |
| ALAS1 | 125290 | chr3_52208160_G/A | ENSG00000023330 | ENST00000394965 | missense_variant | 1603 | 1243 | 415 | A/T | Gca/Aca | rs371320771 | MODERATE | deleterious (0) | probably_damaging (0.992) | FE193 | 1 |
| ALDOA | 103850 | chr16_30070156_G/A | ENSG00000149925 | ENST00000412304 | missense_variant | 1348 | 1039 | 347 | G/S | Ggt/Agt | rs138824667,COSV57633016 | MODERATE | tolerated (0.32) | benign (0.013) | FE136 | 1 |
| ALDOA | 103850 | chr16_30070156_G/A | ENSG00000149925 | ENST00000395240 | missense_variant | 1223 | 1051 | 351 | G/S | Ggt/Agt | rs138824667,COSV57633016 | MODERATE | tolerated (0.33) | benign (0.047) | FE136 | 1 |
| ALDOA | 103850 | chr16_30070156_G/A | ENSG00000149925 | ENST00000642816 | missense_variant | 1382 | 1201 | 401 | G/S | Ggt/Agt | rs138824667,COSV57633016 | MODERATE | tolerated (0.26) | benign (0.058) | FE136 | 1 |
| ALDOA | 103850 | chr16_30070156_G/A | ENSG00000149925 | ENST00000563060 | missense_variant | 1292 | 1039 | 347 | G/S | Ggt/Agt | rs138824667,COSV57633016 | MODERATE | tolerated (0.32) | benign (0.013) | FE136 | 1 |
| ALDOA | 103850 | chr16_30070156_G/A | ENSG00000149925 | ENST00000565355 | missense_variant | 336 | 337 | 113 | G/S | Ggt/Agt | rs138824667,COSV57633016 | MODERATE | tolerated (0.28) | benign (0.028) | FE136 | 1 |
| ALDOA | 103850 | chr16_30070156_G/A | ENSG00000149925 | ENST00000569545 | missense_variant | 1258 | 1039 | 347 | G/S | Ggt/Agt | rs138824667,COSV57633016 | MODERATE | tolerated (0.32) | benign (0.013) | FE136 | 1 |
| ALDOA | 103850 | chr16_30070156_G/A | ENSG00000149925 | ENST00000643777 | missense_variant | 1228 | 1039 | 347 | G/S | Ggt/Agt | rs138824667,COSV57633016 | MODERATE | tolerated (0.32) | benign (0.013) | FE136 | 1 |
| ALG12 | 607144 | chr22_49910544_C/T | ENSG00000182858 | ENST00000330817 | missense_variant | 613 | 359 | 120 | R/Q | cGg/cAg | rs117687848 | MODERATE | - | benign (0.059) | FE130 | 1 |
| ALG12 | 607144 | chr22_49904344_G/A | ENSG00000182858 | ENST00000492791 | missense_variant,NMD_transcript_variant | 564 | 565 | 189 | P/S | Cca/Tca | rs113652023 | MODERATE | - | unknown (0) | FE154 | 1 |
| ALG9 | 606941 | chr11_111836229_G/A | ENSG00000086848 | ENST00000398006 | missense_variant | 1913 | 1004 | 335 | P/L | cCt/cTt | rs185149177 | MODERATE | tolerated (0.07) | benign (0.025) | FE194 | 1 |
| ALG9 | 606941 | chr11_111836229_G/A | ENSG00000086848 | ENST00000531154 | missense_variant | 1498 | 1025 | 342 | P/L | cCt/cTt | rs185149177 | MODERATE | tolerated (0.06) | benign (0.081) | FE194 | 1 |
| ALG9 | 606941 | chr11_111836229_G/A | ENSG00000086848 | ENST00000532425 | missense_variant | 271 | 272 | 91 | P/L | cCt/cTt | rs185149177 | MODERATE | deleterious (0.02) | benign (0.005) | FE194 | 1 |
| ALG9 | 606941 | chr11_111836229_G/A | ENSG00000086848 | ENST00000614444 | missense_variant | 1568 | 1517 | 506 | P/L | cCt/cTt | rs185149177 | MODERATE | tolerated (0.06) | benign (0.025) | FE194 | 1 |
| ALG9 | 606941 | chr11_111836229_G/A | ENSG00000086848 | ENST00000616540 | missense_variant | 1637 | 1538 | 513 | P/L | cCt/cTt | rs185149177 | MODERATE | tolerated (0.06) | benign (0.081) | FE194 | 1 |

| | | | | | | | | | | | | | | | | |
|---|---|---|---|---|---|---|---|---|---|---|---|---|---|---|---|---|
| ALX1 | 601527 | chr12_85280451_C/T | ENSG00000180318 | ENST00000316824 | missense_variant | 232 | 190 | 64 | R/C | Cgc/Tgc | rs145944049 | MODERATE | tolerated_low_confidence (0.06) | probably_damaging (0.973) | FE165 | 1 |
| AMBRA1 | 611359 | chr11_46408531_C/A | ENSG00000110497 | ENST00000314845 | missense_variant | 3475 | 3115 | 1039 | A/S | Gcc/Tcc | rs72910100 | MODERATE | tolerated_low_confidence (0.79) | benign (0.001) | FE190 | 1 |
| AMBRA1 | 611359 | chr11_46408531_C/A | ENSG00000110497 | ENST00000458649 | missense_variant | 3804 | 3385 | 1129 | A/S | Gcc/Tcc | rs72910100 | MODERATE | tolerated_low_confidence (0.82) | benign (0) | FE190 | 1 |
| AMBRA1 | 611359 | chr11_46408531_C/A | ENSG00000110497 | ENST00000526545 | missense_variant | 259 | 259 | 87 | A/S | Gcc/Tcc | rs72910100 | MODERATE | tolerated_low_confidence (0.79) | benign (0.034) | FE190 | 1 |
| AMBRA1 | 611359 | chr11_46408531_C/A | ENSG00000110497 | ENST00000528950 | missense_variant | 3590 | 3298 | 1100 | A/S | Gcc/Tcc | rs72910100 | MODERATE | tolerated_low_confidence (0.83) | benign (0.001) | FE190 | 1 |
| AMBRA1 | 611359 | chr11_46408531_C/A | ENSG00000110497 | ENST00000533727 | missense_variant | 3269 | 3028 | 1010 | A/S | Gcc/Tcc | rs72910100 | MODERATE | tolerated_low_confidence (0.88) | benign (0.001) | FE190 | 1 |
| AMBRA1 | 611359 | chr11_46408531_C/A | ENSG00000110497 | ENST00000534300 | missense_variant | 3519 | 3205 | 1069 | A/S | Gcc/Tcc | rs72910100 | MODERATE | tolerated_low_confidence (0.74) | benign (0.001) | FE190 | 1 |
| ANKHD1 | 610500 | chr5_140529328_G/A | ENSG00000131503 | ENST00000297183 | missense_variant | 6536 | 1531 | 511 | V/I | Gta/Ata | - | MODERATE | tolerated_low_confidence (0.13) | benign (0) | FE190 | 1 |
| APEX1 | 107748 | chr14_20456008_G/C | ENSG00000100823 | ENST00000398030 | missense_variant | 324 | 153 | 51 | Q/H | caG/caC | rs1048945,CM071559 | MODERATE | deleterious (0.03) | benign (0.013) | FE130 | 1 |
| APEX1 | 107748 | chr14_20456008_G/C | ENSG00000100823 | ENST00000553368 | missense_variant | 281 | 102 | 34 | Q/H | caG/caC | rs1048945,CM071559 | MODERATE | deleterious_low_confidence (0) | benign (0.237) | FE130 | 1 |
| APEX1 | 107748 | chr14_20456008_G/C | ENSG00000100823 | ENST00000216714 | missense_variant | 390 | 153 | 51 | Q/H | caG/caC | rs1048945,CM071559 | MODERATE | deleterious (0.03) | benign (0.013) | FE130 | 1 |
| APEX1 | 107748 | chr14_20456008_G/C | ENSG00000100823 | ENST00000555839 | missense_variant | 377 | 153 | 51 | Q/H | caG/caC | rs1048945,CM071559 | MODERATE | deleterious (0.05) | benign (0.168) | FE130 | 1 |
| APEX1 | 107748 | chr14_20456008_G/C | ENSG00000100823 | ENST00000556054 | missense_variant | 403 | 153 | 51 | Q/H | caG/caC | rs1048945,CM071559 | MODERATE | deleterious (0.02) | benign (0.013) | FE130 | 1 |
| APEX1 | 107748 | chr14_20456008_G/C | ENSG00000100823 | ENST00000553681 | missense_variant | 261 | 153 | 51 | Q/H | caG/caC | rs1048945,CM071559 | MODERATE | deleterious (0.04) | benign (0.013) | FE130 | 1 |
| APEX1 | 107748 | chr14_20456008_G/C | ENSG00000100823 | ENST00000555414 | missense_variant | 369 | 153 | 51 | Q/H | caG/caC | rs1048945,CM071559 | MODERATE | deleterious (0.03) | benign (0.013) | FE130 | 1 |
| APEX1 | 107748 | chr14_20456008_G/C | ENSG00000100823 | ENST00000557344 | missense_variant | 385 | 153 | 51 | Q/H | caG/caC | rs1048945,CM071559 | MODERATE | deleterious (0.01) | benign (0.013) | FE130 | 1 |
| APEX1 | 107748 | chr14_20456008_G/C | ENSG00000100823 | ENST00000557592 | missense_variant | 225 | 102 | 34 | Q/H | caG/caC | rs1048945,CM071559 | MODERATE | deleterious (0.03) | benign (0.237) | FE130 | 1 |
| APEX1 | 107748 | chr14_20456008_G/C | ENSG00000100823 | ENST00000557150 | missense_variant | 355 | 102 | 34 | Q/H | caG/caC | rs1048945,CM071559 | MODERATE | deleterious (0.02) | benign (0.237) | FE130 | 1 |
| APEX1 | 107748 | chr14_20456008_G/C | ENSG00000100823 | ENST00000557181 | missense_variant | 396 | 153 | 51 | Q/H | caG/caC | rs1048945,CM071559 | MODERATE | deleterious (0.02) | benign (0.013) | FE130 | 1 |
| ARHGAP1 | 602732 | chr11_46680520_G/A | ENSG00000175220 | ENST00000528837 | missense_variant | 647 | 649 | 217 | L/F | Ctc/Ttc | rs144801476 | MODERATE | tolerated (0.07) | possibly_damaging (0.517) | FE187 | 1 |
| ARHGAP1 | 602732 | chr11_46680520_G/A | ENSG00000175220 | ENST00000311956 | missense_variant | 905 | 787 | 263 | L/F | Ctc/Ttc | rs144801476 | MODERATE | tolerated (0.06) | possibly_damaging (0.649) | FE187 | 1 |
| ARHGAP21 | 609870 | chr10_24635037_G/A | ENSG00000107863 | ENST00000396432 | missense_variant | 1041 | 335 | 112 | P/L | cCt/cTt | COSV64541567 | MODERATE | deleterious (0) | probably_damaging (1) | FE165 | 1 |
| ARHGAP31 | 610911 | chr3_119402349_G/A | ENSG00000031081 | ENST00000264245 | missense_variant | 2119 | 1597 | 533 | G/R | Gga/Aga | rs75764457 | MODERATE | deleterious (0.01) | probably_damaging (0.95) | FE154 | 1 |
| ARHGEF2 | 607560 | chr1_155951529_G/A | ENSG00000116584 | ENST00000361247 | missense_variant | 2333 | 2213 | 738 | A/V | gCg/gTg | rs138090439 | MODERATE | tolerated (1) | benign (0) | FE154 | 1 |
| ASCL2 | 601886 | chr11_2269965_T/C | ENSG00000183734 | ENST00000331289 | missense_variant | 988 | 368 | 123 | Q/R | cAg/cGg | rs376558492 | MODERATE | tolerated (0.11) | benign (0) | FE165 | 1 |
| ASCL2 | 601886 | chr11_2269965_T/C | ENSG00000183734 | ENST00000331289 | missense_variant | 988 | 368 | 123 | Q/R | cAg/cGg | rs376558492 | MODERATE | tolerated (0.11) | benign (0) | FE164 | 1 |
| ASPM | 605481 | chr1_197104066_G/A | ENSG00000066279 | ENST00000367409 | missense_variant | 5417 | 5185 | 1729 | R/W | Cgg/Tgg | rs41299623 | MODERATE | deleterious (0) | probably_damaging (0.978) | FE130 | 1 |
| ASTN1 | 600904 | chr1_176864332_C/G | ENSG00000152092 | ENST00000361833 | missense_variant | 3873 | 3837 | 1279 | Q/H | caG/caC | - | MODERATE | deleterious_low_confidence (0) | probably_damaging (0.977) | FE190 | 1 |
| ATAD2B | 615347 | chr2_23757445_C/T | ENSG00000119778 | ENST00000238789 | missense_variant | 4404 | 4051 | 1351 | V/I | Gtt/Att | rs115642848 | MODERATE | tolerated (0.52) | benign (0.003) | FE193 | 1 |
| ATF7IP | 613644 | chr12_14424331_T/G | ENSG00000171681 | ENST00000261168 | missense_variant | 619 | 416 | 139 | L/R | cTg/cGg | rs200853057 | MODERATE | deleterious_low_confidence (0) | benign (0.08) | FE193 | 1 |
| ATG4B | 611338 | chr2_241653596_G/A | ENSG00000168397 | ENST00000404914 | missense_variant | 291 | 269 | 90 | R/Q | cGg/cAg | rs143448469 | MODERATE | tolerated (0.11) | benign (0.103) | FE136 | 1 |
| ATP13A3 | 610232 | chr3_194427130_C/T | ENSG00000133657 | ENST00000439040 | missense_variant | 3862 | 3070 | 1024 | G/S | Ggt/Agt | rs199767870 | MODERATE | deleterious (0.02) | benign (0.04) | FE190 | 1 |
| ATP6V1B2 | 606939 | chr8_20197417_G/A | ENSG00000147416 | ENST00000523478 | missense_variant,NMD_transcript_variant | 14 | 11 | 4 | R/Q | cGg/cAg | rs116941637 | MODERATE | deleterious_low_confidence (0.02) | probably_damaging (0.947) | FE164 | 1 |
| ATP6V1B2 | 606939 | chr8_20197417_G/A | ENSG00000147416 | ENST00000276390 | missense_variant | 37 | 11 | 4 | R/Q | cGg/cAg | rs116941637 | MODERATE | deleterious_low_confidence (0.02) | benign (0.007) | FE164 | 1 |
| ATP6V1F | 607160 | chr7_128865179_C/T | ENSG00000128524 | ENST00000492758 | missense_variant | 219 | 218 | 73 | P/L | cCg/cTg | rs117627878 | MODERATE | deleterious_low_confidence (0.01) | unknown (0) | FE136 | 1 |
| ATXN2L | 607931 | chr16_28829482_T/G | ENSG00000168488 | ENST00000325215 | missense_variant | 990 | 823 | 275 | S/A | Tct/Gct | - | MODERATE | tolerated (0.09) | benign (0.3) | FE130 | 1 |
| ATXN2L | 607931 | chr16_28829482_T/G | ENSG00000168488 | ENST00000336783 | missense_variant | 990 | 823 | 275 | S/A | Tct/Gct | - | MODERATE | tolerated (0.09) | benign (0.426) | FE130 | 1 |
| ATXN2L | 607931 | chr16_28829482_T/G | ENSG00000168488 | ENST00000340394 | missense_variant | 1035 | 823 | 275 | S/A | Tct/Gct | - | MODERATE | tolerated (0.09) | benign (0.3) | FE130 | 1 |
| ATXN2L | 607931 | chr16_28829482_T/G | ENSG00000168488 | ENST00000382686 | missense_variant | 990 | 823 | 275 | S/A | Tct/Gct | - | MODERATE | tolerated (0.09) | benign (0.426) | FE130 | 1 |
| ATXN2L | 607931 | chr16_28829482_T/G | ENSG00000168488 | ENST00000395547 | missense_variant | 990 | 823 | 275 | S/A | Tct/Gct | - | MODERATE | tolerated (0.08) | benign (0.3) | FE130 | 1 |
| ATXN2L | 607931 | chr16_28829482_T/G | ENSG00000168488 | ENST00000564304 | missense_variant | 988 | 823 | 275 | S/A | Tct/Gct | - | MODERATE | tolerated (0.08) | benign (0.075) | FE130 | 1 |
| ATXN2L | 607931 | chr16_28829482_T/G | ENSG00000168488 | ENST00000568266 | missense_variant | 929 | 643 | 215 | S/A | Tct/Gct | - | MODERATE | tolerated (0.1) | benign (0.001) | FE130 | 1 |

| | | | | | | | | | | | | | | | | |
|---|---|---|---|---|---|---|---|---|---|---|---|---|---|---|---|---|
| ATXN2L | 607931 | chr16_28829482_T/G | ENSG00000168488 | ENST00000570200 | missense_variant | 831 | 823 | 275 | S/A | Tct/Gct | - | MODERATE | tolerated (0.09) | benign (0.426) | FE130 | 1 |
| AURKB | 604970 | chr17_8207267_C/T | ENSG00000178999 | ENST00000577833 | missense_variant | 362 | 187 | 63 | V/M | Gtg/Atg | rs150216235 | MODERATE | deleterious (0.03) | probably_damaging (0.999) | FE193 | 1 |
| AURKB | 604970 | chr17_8207267_C/T | ENSG00000178999 | ENST00000581511 | missense_variant | 363 | 307 | 103 | V/M | Gtg/Atg | rs150216235 | MODERATE | deleterious (0.03) | possibly_damaging (0.759) | FE193 | 1 |
| AURKB | 604970 | chr17_8207267_C/T | ENSG00000178999 | ENST00000583915 | missense_variant | 372 | 184 | 62 | V/M | Gtg/Atg | rs150216235 | MODERATE | deleterious (0.02) | possibly_damaging (0.759) | FE193 | 1 |
| AURKB | 604970 | chr17_8207267_C/T | ENSG00000178999 | ENST00000584972 | missense_variant | 101 | 103 | 35 | V/M | Gtg/Atg | rs150216235 | MODERATE | deleterious (0.02) | probably_damaging (0.998) | FE193 | 1 |
| AURKB | 604970 | chr17_8207267_C/T | ENSG00000178999 | ENST00000585124 | missense_variant | 378 | 307 | 103 | V/M | Gtg/Atg | rs150216235 | MODERATE | deleterious (0.03) | possibly_damaging (0.759) | FE193 | 1 |
| AURKB | 604970 | chr17_8207267_C/T | ENSG00000178999 | ENST00000582368 | missense_variant | 439 | 184 | 62 | V/M | Gtg/Atg | rs150216235 | MODERATE | deleterious (0.02) | possibly_damaging (0.759) | FE193 | 1 |
| AURKB | 604970 | chr17_8207267_C/T | ENSG00000178999 | ENST00000316199 | missense_variant | 388 | 310 | 104 | V/M | Gtg/Atg | rs150216235 | MODERATE | deleterious (0.03) | probably_damaging (0.999) | FE193 | 1 |
| AURKB | 604970 | chr17_8207267_C/T | ENSG00000178999 | ENST00000534871 | missense_variant | 311 | 184 | 62 | V/M | Gtg/Atg | rs150216235 | MODERATE | deleterious (0.03) | possibly_damaging (0.759) | FE193 | 1 |
| BARD1 | 601593 | chr2_214770791_G/A | ENSG00000138376 | ENST00000650978 | missense_variant,NMD_transcript_variant | 1211 | 1213 | 405 | L/F | Ctc/Ttc | rs562168985 | MODERATE | deleterious_low_confidence (0.01) | benign (0.045) | FE154 | 1 |
| BBS4 | 600374 | chr15_72715321_A/G | ENSG00000140463 | ENST00000268057 | missense_variant | 272 | 251 | 84 | Q/R | cAa/cGa | - | MODERATE | tolerated (0.08) | benign (0.133) | FE164 | 1 |
| BBS4 | 600374 | chr15_72715321_A/G | ENSG00000140463 | ENST00000565160 | missense_variant,NMD_transcript_variant | 285 | 251 | 84 | Q/R | cAa/cGa | - | MODERATE | deleterious (0.02) | possibly_damaging (0.897) | FE164 | 1 |
| BBS4 | 600374 | chr15_72715321_A/G | ENSG00000140463 | ENST00000566829 | missense_variant | 396 | 269 | 90 | Q/R | cAa/cGa | - | MODERATE | deleterious (0.03) | possibly_damaging (0.876) | FE164 | 1 |
| BBS4 | 600374 | chr15_72715321_A/G | ENSG00000140463 | ENST00000569338 | missense_variant | 242 | 242 | 81 | Q/R | cAa/cGa | - | MODERATE | deleterious (0.05) | benign (0.093) | FE164 | 1 |
| BHLHE40 | 604256 | chr3_4982896_C/T | ENSG00000134107 | ENST00000256495 | missense_variant | 725 | 443 | 148 | T/I | aCa/aTa | rs148401238 | MODERATE | tolerated (0.45) | benign (0.062) | FE130 | 1 |
| BIN1 | 601248 | chr2_127050470_T/C | ENSG00000136717 | ENST00000393041 | missense_variant | 1616 | 1271 | 424 | K/R | aAg/aGg | rs138047593,CM1512481,COSV52116674 | MODERATE | deleterious (0.01) | benign (0.018) | FE136 | 1 |
| BIN1 | 601248 | chr2_127050470_T/C | ENSG00000136717 | ENST00000352848 | missense_variant | 1553 | 1208 | 403 | K/R | aAg/aGg | rs138047593,CM1512481,COSV52116674 | MODERATE | deleterious (0.04) | probably_damaging (0.997) | FE136 | 1 |
| BIN1 | 601248 | chr2_127050470_T/C | ENSG00000136717 | ENST00000409400 | missense_variant | 1498 | 1163 | 388 | K/R | aAg/aGg | rs138047593,CM1512481,COSV52116674 | MODERATE | deleterious (0) | probably_damaging (0.997) | FE136 | 1 |
| BIN1 | 601248 | chr2_127050470_T/C | ENSG00000136717 | ENST00000376113 | missense_variant | 1176 | 1118 | 373 | K/R | aAg/aGg | rs138047593,CM1512481,COSV52116674 | MODERATE | deleterious (0.05) | probably_damaging (0.993) | FE136 | 1 |
| BIN1 | 601248 | chr2_127050470_T/C | ENSG00000136717 | ENST00000348750 | missense_variant | 1418 | 1073 | 358 | K/R | aAg/aGg | rs138047593,CM1512481,COSV52116674 | MODERATE | tolerated (0.06) | probably_damaging (0.929) | FE136 | 1 |
| BIN1 | 601248 | chr2_127050470_T/C | ENSG00000136717 | ENST00000357970 | missense_variant | 1841 | 1496 | 499 | K/R | aAg/aGg | rs138047593,CM1512481,COSV52116674 | MODERATE | deleterious (0.01) | probably_damaging (0.92) | FE136 | 1 |
| BIN1 | 601248 | chr2_127050470_T/C | ENSG00000136717 | ENST00000316724 | missense_variant | 1836 | 1625 | 542 | K/R | aAg/aGg | rs138047593,CM1512481,COSV52116674 | MODERATE | deleterious (0.03) | possibly_damaging (0.491) | FE136 | 1 |
| BIN1 | 601248 | chr2_127050470_T/C | ENSG00000136717 | ENST00000393040 | missense_variant | 1637 | 1292 | 431 | K/R | aAg/aGg | rs138047593,CM1512481,COSV52116674 | MODERATE | tolerated (0.05) | benign (0.009) | FE136 | 1 |
| BIN1 | 601248 | chr2_127050470_T/C | ENSG00000136717 | ENST00000259238 | missense_variant | 1682 | 1337 | 446 | K/R | aAg/aGg | rs138047593,CM1512481,COSV52116674 | MODERATE | tolerated (0.05) | benign (0.009) | FE136 | 1 |
| BIN1 | 601248 | chr2_127050470_T/C | ENSG00000136717 | ENST00000351659 | missense_variant | 1709 | 1364 | 455 | K/R | aAg/aGg | rs138047593,CM1512481,COSV52116674 | MODERATE | deleterious (0.01) | benign (0.009) | FE136 | 1 |
| BIN1 | 601248 | chr2_127050470_T/C | ENSG00000136717 | ENST00000346226 | missense_variant | 1745 | 1400 | 467 | K/R | aAg/aGg | rs138047593,CM1512481,COSV52116674 | MODERATE | deleterious (0.01) | possibly_damaging (0.493) | FE136 | 1 |
| BNIP1 | 603291 | chr5_173159948_G/C | ENSG00000113734 | ENST00000231668 | missense_variant | 620 | 516 | 172 | E/D | gaG/gaC | - | MODERATE | tolerated (0.09) | benign (0.354) | FE164 | 1 |
| BNIP1 | 603291 | chr5_173159948_G/C | ENSG00000113734 | ENST00000351486 | missense_variant | 418 | 387 | 129 | E/D | gaG/gaC | - | MODERATE | tolerated (0.12) | benign (0.202) | FE164 | 1 |
| BNIP1 | 603291 | chr5_173159948_G/C | ENSG00000113734 | ENST00000352523 | missense_variant | 423 | 414 | 138 | E/D | gaG/gaC | - | MODERATE | tolerated (0.13) | benign (0.06) | FE164 | 1 |
| BNIP1 | 603291 | chr5_173159948_G/C | ENSG00000113734 | ENST00000393770 | missense_variant | 285 | 285 | 95 | E/D | gaG/gaC | - | MODERATE | tolerated (0.1) | possibly_damaging (0.446) | FE164 | 1 |
| BORA | 610510 | chr13_72745098_C/T | ENSG00000136122 | ENST00000377814 | missense_variant | 561 | 563 | 188 | S/L | tCg/tTg | rs9543107 | MODERATE | tolerated (0.13) | benign (0.007) | FE130 | 1 |
| BORA | 610510 | chr13_72745098_C/T | ENSG00000136122 | ENST00000390667 | missense_variant | 729 | 629 | 210 | S/L | tCg/tTg | rs9543107 | MODERATE | tolerated (0.15) | benign (0.003) | FE130 | 1 |
| BORA | 610510 | chr13_72745098_C/T | ENSG00000136122 | ENST00000613797 | missense_variant | 1001 | 854 | 285 | S/L | tCg/tTg | rs9543107 | MODERATE | tolerated (0.17) | benign (0.062) | FE130 | 1 |
| BORA | 610510 | chr13_72745098_C/T | ENSG00000136122 | ENST00000651477 | missense_variant | 1001 | 629 | 210 | S/L | tCg/tTg | rs9543107 | MODERATE | tolerated (0.15) | benign (0.003) | FE130 | 1 |
| BORA | 610510 | chr13_72745098_C/T | ENSG00000136122 | ENST00000652266 | missense_variant | 796 | 419 | 140 | S/L | tCg/tTg | rs9543107 | MODERATE | tolerated (0.15) | benign (0.003) | FE130 | 1 |
| BRINP1 | 602865 | chr9_119208791_C/T | ENSG00000078725 | ENST00000265922 | missense_variant | 1503 | 1073 | 358 | R/H | cGc/cAc | rs17476783,COSV56305264 | MODERATE | deleterious (0.02) | benign (0) | FE190 | 1 |
| BRIP1 | 605882 | chr17_61847211_G/A | ENSG00000136492 | ENST00000259008 | missense_variant | 792 | 517 | 173 | R/C | Cgt/Tgt | rs4988345,CM035889 | MODERATE | deleterious (0.01) | benign (0.069) | FE190 | 1 |
| BRIP1 | 605882 | chr17_61847211_G/A | ENSG00000136492 | ENST00000577598 | missense_variant | 517 | 517 | 173 | R/C | Cgt/Tgt | rs4988345,CM035889 | MODERATE | deleterious (0.01) | probably_damaging (0.973) | FE190 | 1 |
| BTF3 | 602542 | chr5_73498700_C/G | ENSG00000145741 | ENST00000380591 | missense_variant | 252 | 33 | 11 | D/E | gaC/gaG | rs375502008 | MODERATE | deleterious_low_confidence (0.02) | benign (0) | FE106 | 1 |
| BUB1 | 602452 | chr2_110657067_C/A | ENSG00000169679 | ENST00000671097 | missense_variant,NMD_transcript_variant | 127 | 128 | 43 | R/L | cGc/cTc | - | MODERATE | deleterious (0) | possibly_damaging (0.771) | FE130 | 1 |
| BUB1 | 602452 | chr2_110657067_C/A | ENSG00000169679 | ENST00000535254 | missense_variant | 1675 | 1607 | 536 | R/L | cGc/cTc | - | MODERATE | deleterious (0) | possibly_damaging (0.839) | FE130 | 1 |
| BUB1 | 602452 | chr2_110657067_C/A | ENSG00000169679 | ENST00000302759 | missense_variant | 1735 | 1667 | 556 | R/L | cGc/cTc | - | MODERATE | deleterious (0) | possibly_damaging (0.77) | FE130 | 1 |

| | | | | | | | | | | | | | | | | |
|---|---|---|---|---|---|---|---|---|---|---|---|---|---|---|---|---|
| BUB1 | 602452 | chr2_110657067_C/A | ENSG00000169679 | ENST00000409311 | missense_variant | 1735 | 1667 | 556 | R/L | cGc/cTc | - | MODERATE | deleterious (0) | possibly_damaging (0.76) | FE130 | 1 |
| CACNA1I | 608230 | chr22_39649648_C/T | ENSG00000100346 | ENST00000402142 | missense_variant | 1715 | 1715 | 572 | S/L | tCg/tTg | rs372904212 | MODERATE | tolerated (0.08) | probably_damaging (0.968) | FE194 | 1 |
| CASP9 | 602234 | chr1_15504674_C/T | ENSG00000132906 | ENST00000333868 | missense_variant | 820 | 805 | 269 | G/R | Ggg/Agg | rs745473642 | MODERATE | deleterious (0) | probably_damaging (0.967) | FE190 | 1 |
| CASP9 | 602234 | chr1_15504674_C/T | ENSG00000132906 | ENST00000375890 | missense_variant | 817 | 556 | 186 | G/R | Ggg/Agg | rs745473642 | MODERATE | deleterious (0) | probably_damaging (0.967) | FE190 | 1 |
| CASP9 | 602234 | chr1_15504674_C/T | ENSG00000132906 | ENST00000447522 | missense_variant | 711 | 556 | 186 | G/R | Ggg/Agg | rs745473642 | MODERATE | deleterious (0) | probably_damaging (0.967) | FE190 | 1 |
| CASP9 | 602234 | chr1_15504674_C/T | ENSG00000132906 | ENST00000546424 | missense_variant | 1050 | 805 | 269 | G/R | Ggg/Agg | rs745473642 | MODERATE | deleterious (0) | probably_damaging (0.999) | FE190 | 1 |
| CASP9 | 602234 | chr1_15504674_C/T | ENSG00000132906 | ENST00000424908 | missense_variant | 329 | 331 | 111 | G/R | Ggg/Agg | rs745473642 | MODERATE | deleterious (0) | probably_damaging (1) | FE190 | 1 |
| CASP9 | 602234 | chr1_15504674_C/T | ENSG00000132906 | ENST00000440484 | missense_variant | 732 | 715 | 239 | G/R | Ggg/Agg | rs745473642 | MODERATE | deleterious (0) | probably_damaging (0.997) | FE190 | 1 |
| CCDC22 | 300859 | chrX_49249509_G/A | ENSG00000101997 | ENST00000376227 | missense_variant,splice_region_variant | 1803 | 1636 | 546 | D/N | Gat/Aat | rs147222955 | MODERATE | deleterious (0.01) | probably_damaging (1) | FE190 | 2 |
| CCDC6 | 601985 | chr10_59793005_G/A | ENSG00000108091 | ENST00000263102 | missense_variant | 1469 | 1337 | 446 | P/L | cCg/cTg | rs61740504 | MODERATE | tolerated_low_confidence (0.05) | benign (0.003) | FE130 | 1 |
| CDC42BPA | 603412 | chr1_227028712_C/A | ENSG00000143776 | ENST00000366769 | missense_variant | 5564 | 4272 | 1424 | Q/H | caG/caT | rs996572366 | MODERATE | tolerated (0.07) | benign (0.085) | FE187 | 1 |
| CDC6 | 602627 | chr17_40293996_G/A | ENSG00000094804 | ENST00000649662 | missense_variant | 913 | 883 | 295 | D/N | Gat/Aat | rs4135012 | MODERATE | deleterious (0) | possibly_damaging (0.831) | FE187 | 1 |
| CDC6 | 602627 | chr17_40293996_G/A | ENSG00000094804 | ENST00000209728 | missense_variant | 1108 | 883 | 295 | D/N | Gat/Aat | rs4135012 | MODERATE | deleterious (0) | possibly_damaging (0.831) | FE187 | 1 |
| CEBPZ | 612828 | chr2_37201897_T/C | ENSG00000115816 | ENST00000234170 | missense_variant | 3061 | 3032 | 1011 | K/R | aAa/aGa | rs1028215374 | MODERATE | deleterious (0.04) | probably_damaging (0.998) | FE165 | 1 |
| CELSR1 | 604523 | chr22_46397724_T/C | ENSG00000075275 | ENST00000674312 | missense_variant | 211 | 212 | 71 | N/S | aAt/aGt | rs144122307 | MODERATE | tolerated (0.16) | benign (0.061) | FE164 | 1 |
| CELSR1 | 604523 | chr22_46397724_T/C | ENSG00000075275 | ENST00000674359 | missense_variant | 664 | 665 | 222 | N/S | aAt/aGt | rs144122307 | MODERATE | tolerated (0.12) | benign (0.068) | FE164 | 1 |
| CELSR1 | 604523 | chr22_46397724_T/C | ENSG00000075275 | ENST00000674500 | missense_variant | 1597 | 1598 | 533 | N/S | aAt/aGt | rs144122307 | MODERATE | tolerated (0.06) | benign (0.019) | FE164 | 1 |
| CELSR1 | 604523 | chr22_46397724_T/C | ENSG00000075275 | ENST00000262738 | missense_variant | 6081 | 5651 | 1884 | N/S | aAt/aGt | rs144122307 | MODERATE | tolerated (0.05) | benign (0.024) | FE164 | 1 |
| CENPH | 605607 | chr5_69191816_A/T | ENSG00000153044 | ENST00000283006 | missense_variant | 208 | 156 | 52 | Q/H | caA/caT | rs149837292 | MODERATE | deleterious (0) | probably_damaging (0.997) | FE194 | 1 |
| CENPH | 605607 | chr5_69191816_A/T | ENSG00000153044 | ENST00000502689 | missense_variant | 49 | 51 | 17 | Q/H | caA/caT | rs149837292 | MODERATE | deleterious (0) | probably_damaging (0.979) | FE194 | 1 |
| CENPH | 605607 | chr5_69191816_A/T | ENSG00000153044 | ENST00000513575 | missense_variant,NMD_transcript_variant | 199 | 156 | 52 | Q/H | caA/caT | rs149837292 | MODERATE | deleterious (0) | probably_damaging (0.997) | FE194 | 1 |
| CENPH | 605607 | chr5_69191816_A/T | ENSG00000153044 | ENST00000515001 | missense_variant | 217 | 156 | 52 | Q/H | caA/caT | rs149837292 | MODERATE | deleterious (0) | probably_damaging (0.995) | FE194 | 1 |
| CFAP298 | 615494 | chr21_32604187_A/G | ENSG00000159079 | ENST00000300260 | missense_variant,NMD_transcript_variant | 764 | 404 | 135 | V/A | gTt/gCt | - | MODERATE | tolerated_low_confidence (0.58) | benign (0) | FE194 | 1 |
| CFAP47 | NA | chrX_35967712_C/T | ENSG00000165164 | ENST00000297866 | missense_variant | 1760 | 1694 | 565 | S/L | tCa/tTa | rs139917087,COSV52888497 | MODERATE | deleterious (0) | probably_damaging (0.964) | FE193 | 1 |
| CHTOP | 614206 | chr1_153643344_G/A | ENSG00000160679 | ENST00000368694 | missense_variant | 816 | 521 | 174 | R/H | cGt/cAt | rs74844193 | MODERATE | tolerated (0.15) | probably_damaging (0.954) | FE187 | 1 |
| CHTOP | 614206 | chr1_153643344_G/A | ENSG00000160679 | ENST00000368690 | missense_variant | 903 | 524 | 175 | R/H | cGt/cAt | rs74844193 | MODERATE | tolerated (0.14) | probably_damaging (0.923) | FE187 | 1 |
| CHTOP | 614206 | chr1_153643344_G/A | ENSG00000160679 | ENST00000368687 | missense_variant | 667 | 446 | 149 | R/H | cGt/cAt | rs74844193 | MODERATE | tolerated (0.14) | probably_damaging (0.954) | FE187 | 1 |
| CINP | 613362 | chr14_102359436_G/C | ENSG00000100865 | ENST00000541568 | missense_variant | 176 | 159 | 53 | N/K | aaC/aaG | rs11544426 | MODERATE | deleterious (0) | probably_damaging (0.998) | FE130 | 1 |
| CINP | 613362 | chr14_102359436_G/C | ENSG00000100865 | ENST00000558764 | missense_variant,NMD_transcript_variant | 222 | 159 | 53 | N/K | aaC/aaG | rs11544426 | MODERATE | deleterious (0) | probably_damaging (0.999) | FE130 | 1 |
| CINP | 613362 | chr14_102359436_G/C | ENSG00000100865 | ENST00000216756 | missense_variant | 198 | 159 | 53 | N/K | aaC/aaG | rs11544426 | MODERATE | deleterious (0) | probably_damaging (0.995) | FE130 | 1 |
| CINP | 613362 | chr14_102359436_G/C | ENSG00000100865 | ENST00000536961 | missense_variant | 293 | 204 | 68 | N/K | aaC/aaG | rs11544426 | MODERATE | deleterious (0) | probably_damaging (0.998) | FE130 | 1 |
| CINP | 613362 | chr14_102359436_G/C | ENSG00000100865 | ENST00000559504 | missense_variant,NMD_transcript_variant | 160 | 162 | 54 | N/K | aaC/aaG | rs11544426 | MODERATE | deleterious (0) | probably_damaging (0.994) | FE130 | 1 |
| CINP | 613362 | chr14_102359436_G/C | ENSG00000100865 | ENST00000559514 | missense_variant,NMD_transcript_variant | 224 | 159 | 53 | N/K | aaC/aaG | rs11544426 | MODERATE | deleterious (0) | probably_damaging (0.995) | FE130 | 1 |
| CLASP1 | 605852 | chr2_121503187_T/C | ENSG00000074054 | ENST00000263710 | missense_variant | 1082 | 692 | 231 | N/S | aAc/aGc | rs770261375 | MODERATE | deleterious (0.04) | benign (0.031) | FE106 | 1 |
| CLK3 | 602990 | chr15_74620124_C/T | ENSG00000179335 | ENST00000395066 | missense_variant | 1173 | 712 | 238 | R/C | Cgt/Tgt | rs767369515 | MODERATE | deleterious (0) | benign (0.328) | FE193 | 1 |
| CLSTN3 | 611324 | chr12_7130691_G/T | ENSG00000139182 | ENST00000266546 | missense_variant | 321 | 43 | 15 | A/S | Gcg/Tcg | rs145190321,COSV56940497 | MODERATE | tolerated (0.61) | benign (0.001) | FE130 | 1 |
| CMPK1 | 191710 | chr1_47375214_A/G | ENSG00000162368 | ENST00000371873 | missense_variant | 715 | 566 | 189 | Q/R | cAg/cGg | - | MODERATE | tolerated (0.06) | benign (0.059) | FE193 | 1 |
| CMPK1 | 191710 | chr1_47375214_A/G | ENSG00000162368 | ENST00000450808 | missense_variant | 451 | 419 | 140 | Q/R | cAg/cGg | - | MODERATE | tolerated (0.06) | benign (0.038) | FE193 | 1 |
| COL4A1 | 120130 | chr13_110178066_G/C | ENSG00000187498 | ENST00000375820 | missense_variant,splice_region_variant | 2754 | 2624 | 875 | P/R | cCa/cGa | rs201964644 | MODERATE | tolerated (0.23) | possibly_damaging (0.89) | FE136 | 1 |
| COL4A5 | 303630 | chrX_108601436_G/T | ENSG00000188153 | ENST00000328300 | missense_variant | 2280 | 1992 | 664 | K/N | aaG/aaT | CD129404,COSV60375567 | MODERATE | tolerated (0.27) | benign (0) | FE165 | 2 |
| COL5A1 | 120215 | chr9_134750808_G/A | ENSG00000130635 | ENST00000618395 | missense_variant | 1971 | 1588 | 530 | G/S | Ggc/Agc | rs61735045,CM020927 | MODERATE | tolerated (0.15) | probably_damaging (1) | FE193 | 1 |
| COL5A1 | 120215 | chr9_134700009_G/T | ENSG00000130635 | ENST00000371817 | missense_variant | 763 | 378 | 126 | Q/H | caG/caT | rs145178917,COSV65675551 | MODERATE | deleterious (0) | probably_damaging (0.911) | FE194 | 1 |

| | | | | | | | | | | | | | | | | |
|---|---|---|---|---|---|---|---|---|---|---|---|---|---|---|---|---|
| COL5A1 | 120215 | chr9_134750808_G/A | ENSG00000130635 | ENST00000371817 | missense_variant | 1973 | 1588 | 530 | G/S | Ggc/Agc | rs61735045,CM020927 | MODERATE | tolerated (0.15) | probably_damaging (0.998) | FE193 | 1 |
| COL5A1 | 120215 | chr9_134700009_G/T | ENSG00000130635 | ENST00000618395 | missense_variant | 761 | 378 | 126 | Q/H | caG/caT | rs145178917,COSV65675551 | MODERATE | deleterious (0) | probably_damaging (0.998) | FE194 | 1 |
| COP1 | 608067 | chr1_176206953_G/C | ENSG00000143207 | ENST00000367669 | missense_variant | 334 | 26 | 9 | S/W | tCg/tGg | rs868382148 | MODERATE | deleterious_low_confidence (0) | benign (0.214) | FE164 | 1 |
| COP1 | 608067 | chr1_176206953_G/C | ENSG00000143207 | ENST00000474194 | missense_variant,NMD_transcript_variant | 131 | 26 | 9 | S/W | tCg/tGg | rs868382148 | MODERATE | deleterious_low_confidence (0) | benign (0.214) | FE164 | 1 |
| COP1 | 608067 | chr1_176206953_G/C | ENSG00000143207 | ENST00000308769 | missense_variant | 26 | 26 | 9 | S/W | tCg/tGg | rs868382148 | MODERATE | deleterious_low_confidence (0) | benign (0.217) | FE164 | 1 |
| COPG1 | 615525 | chr3_129250754_C/G | ENSG00000181789 | ENST00000504350 | missense_variant,NMD_transcript_variant | 167 | 110 | 37 | P/R | cCt/cGt | rs368427024 | MODERATE | - | unknown (0) | FE187 | 1 |
| COPS4 | 616008 | chr4_83073303_G/C | ENSG00000138663 | ENST00000511653 | missense_variant | 1185 | 1185 | 395 | L/F | ttG/ttC | rs774050810 | MODERATE | tolerated_low_confidence (0.18) | possibly_damaging (0.587) | FE154 | 1 |
| COQ2 | 609825 | chr4_83284851_T/A | ENSG00000173085 | ENST00000311469 | stop_gained | 64 | 64 | 22 | R/* | Aga/Tga | rs112033303,CM135690 | HIGH | - | - | FE164 | 1 |
| COQ5 | 616359 | chr12_120503842_A/G | ENSG00000110871 | ENST00000288532 | missense_variant | 943 | 926 | 309 | V/A | gTg/gCg | rs141304059 | MODERATE | deleterious (0.02) | possibly_damaging (0.874) | FE190 | 1 |
| COQ5 | 616359 | chr12_120503842_A/G | ENSG00000110871 | ENST00000445328 | missense_variant | 724 | 704 | 235 | V/A | gTg/gCg | rs141304059 | MODERATE | tolerated (0.09) | benign (0.241) | FE190 | 1 |
| COX19 | 610429 | chr7_935399_G/A | ENSG00000240230 | ENST00000457254 | missense_variant,NMD_transcript_variant | 341 | 301 | 101 | R/C | Cgc/Tgc | rs370559460 | MODERATE | deleterious_low_confidence (0.01) | unknown (0) | FE193 | 1 |
| CPT1A | 600528 | chr11_68807618_G/A | ENSG00000110090 | ENST00000265641 | missense_variant | 457 | 302 | 101 | T/M | aCg/aTg | rs61731903 | MODERATE | tolerated (0.23) | benign (0.028) | FE106 | 1 |
| CRNKL1 | 610952 | chr20_20052554_C/T | ENSG00000101343 | ENST00000377327 | missense_variant | 268 | 236 | 79 | C/Y | tGc/tAc | rs145079188 | MODERATE | deleterious_low_confidence (0.01) | benign (0.025) | FE136 | 1 |
| CRNKL1 | 610952 | chr20_20052554_C/T | ENSG00000101343 | ENST00000377340 | missense_variant | 304 | 272 | 91 | C/Y | tGc/tAc | rs145079188 | MODERATE | deleterious_low_confidence (0.01) | benign (0) | FE136 | 1 |
| CRNKL1 | 610952 | chr20_20052554_C/T | ENSG00000101343 | ENST00000490910 | missense_variant,NMD_transcript_variant | 268 | 236 | 79 | C/Y | tGc/tAc | rs145079188 | MODERATE | tolerated_low_confidence (0.09) | benign (0.025) | FE136 | 1 |
| CRNKL1 | 610952 | chr20_20052554_C/T | ENSG00000101343 | ENST00000496549 | missense_variant,NMD_transcript_variant | 174 | 142 | 48 | A/T | Gcc/Acc | rs145079188 | MODERATE | - | unknown (0) | FE136 | 1 |
| CRY2 | 603732 | chr11_45870365_G/A | ENSG00000121671 | ENST00000443527 | missense_variant | 1467 | 1445 | 482 | R/Q | cGa/cAa | rs139385343 | MODERATE | tolerated (0.14) | benign (0.019) | FE154 | 1 |
| CSNK1G1 | 606274 | chr15_64195080_C/T | ENSG00000169118 | ENST00000635414 | splice_acceptor_variant | - | - | - | - | - | - | HIGH | - | - | FE106 | 1 |
| CSTB | 601145 | chr21_43774605_G/A | ENSG00000160213 | ENST00000640406 | missense_variant | 270 | 221 | 74 | P/L | cCg/cTg | rs180832281 | MODERATE | tolerated_low_confidence (0.19) | probably_damaging (0.988) | FE154 | 1 |
| CTNNA2 | 114025 | chr2_80393245_T/C | ENSG00000066032 | ENST00000409550 | missense_variant | 110 | 86 | 29 | I/T | aTt/aCt | rs61754542 | MODERATE | tolerated (0.63) | benign (0) | FE154 | 1 |
| CTNNA2 | 114025 | chr2_80393245_T/C | ENSG00000066032 | ENST00000629316 | missense_variant | 1169 | 1091 | 364 | I/T | aTt/aCt | rs61754542 | MODERATE | tolerated (0.62) | benign (0) | FE154 | 1 |
| CTNNA2 | 114025 | chr2_80393245_T/C | ENSG00000066032 | ENST00000466387 | missense_variant | 1815 | 1091 | 364 | I/T | aTt/aCt | rs61754542 | MODERATE | tolerated (0.61) | benign (0) | FE154 | 1 |
| CTNNA2 | 114025 | chr2_80393245_T/C | ENSG00000066032 | ENST00000496558 | missense_variant | 1304 | 1091 | 364 | I/T | aTt/aCt | rs61754542 | MODERATE | tolerated (0.61) | benign (0) | FE154 | 1 |
| CTNNA2 | 114025 | chr2_80393245_T/C | ENSG00000066032 | ENST00000402739 | missense_variant | 1096 | 1091 | 364 | I/T | aTt/aCt | rs61754542 | MODERATE | tolerated (0.61) | benign (0.001) | FE154 | 1 |
| CTNNA2 | 114025 | chr2_80393245_T/C | ENSG00000066032 | ENST00000343114 | missense_variant | 311 | 128 | 43 | I/T | aTt/aCt | rs61754542 | MODERATE | tolerated (0.61) | benign (0.001) | FE154 | 1 |
| CUL2 | 603135 | chr10_35033271_C/T | ENSG00000108094 | ENST00000374749 | missense_variant,splice_region_variant | 1215 | 1005 | 335 | M/I | atG/atA | rs61749171 | MODERATE | tolerated (1) | benign (0) | FE187 | 1 |
| CUL2 | 603135 | chr10_35033271_C/T | ENSG00000108094 | ENST00000374751 | missense_variant,splice_region_variant | 1240 | 1005 | 335 | M/I | atG/atA | rs61749171 | MODERATE | tolerated (1) | benign (0) | FE187 | 1 |
| CUL2 | 603135 | chr10_35033271_C/T | ENSG00000108094 | ENST00000374746 | missense_variant,splice_region_variant | 1027 | 1005 | 335 | M/I | atG/atA | rs61749171 | MODERATE | tolerated (1) | benign (0) | FE187 | 1 |
| CUL2 | 603135 | chr10_35033271_C/T | ENSG00000108094 | ENST00000537177 | missense_variant,splice_region_variant | 1279 | 1044 | 348 | M/I | atG/atA | rs61749171 | MODERATE | tolerated (1) | probably_damaging (0.986) | FE187 | 1 |
| CUL2 | 603135 | chr10_35033271_C/T | ENSG00000108094 | ENST00000626172 | missense_variant,splice_region_variant | 1240 | 1005 | 335 | M/I | atG/atA | rs61749171 | MODERATE | tolerated (1) | benign (0) | FE187 | 1 |
| CUL2 | 603135 | chr10_35033271_C/T | ENSG00000108094 | ENST00000673636 | missense_variant,splice_region_variant | 1314 | 1005 | 335 | M/I | atG/atA | rs61749171 | MODERATE | tolerated (1) | benign (0) | FE187 | 1 |
| CUL2 | 603135 | chr10_35033271_C/T | ENSG00000108094 | ENST00000421317 | missense_variant,splice_region_variant | 1124 | 1062 | 354 | M/I | atG/atA | rs61749171 | MODERATE | tolerated (0.99) | benign (0) | FE187 | 1 |
| CUL2 | 603135 | chr10_35033271_C/T | ENSG00000108094 | ENST00000374748 | missense_variant,splice_region_variant | 1319 | 1005 | 335 | M/I | atG/atA | rs61749171 | MODERATE | tolerated (1) | benign (0) | FE187 | 1 |
| CWC22 | 615186 | chr2_179950685_G/A | ENSG00000163510 | ENST00000404136 | missense_variant | 2124 | 1967 | 656 | A/V | gCg/gTg | rs17778270 | MODERATE | tolerated (0.15) | possibly_damaging (0.453) | FE190 | 1 |
| CWC22 | 615186 | chr2_179950685_G/A | ENSG00000163510 | ENST00000410053 | missense_variant | 2511 | 1967 | 656 | A/V | gCg/gTg | rs17778270 | MODERATE | tolerated (0.17) | possibly_damaging (0.453) | FE190 | 1 |
| DBN1 | 126660 | chr5_177457924_T/A | ENSG00000113758 | ENST00000512501 | missense_variant | 1321 | 1106 | 369 | H/L | cAc/cTc | rs188055232 | MODERATE | tolerated_low_confidence (0.06) | unknown (0) | FE164 | 1 |
| DBR1 | 607024 | chr3_138162242_C/G | ENSG00000138231 | ENST00000260803 | missense_variant | 1408 | 1282 | 428 | E/Q | Gaa/Caa | rs36061810 | MODERATE | tolerated (0.09) | probably_damaging (0.996) | FE190 | 1 |
| DCLK1 | 604742 | chr13_35822787_A/T | ENSG00000133083 | ENST00000255448 | missense_variant | 1708 | 1496 | 499 | I/N | aTc/aAc | - | MODERATE | deleterious (0) | possibly_damaging (0.8) | FE164 | 1 |
| DCP2 | 609844 | chr5_112976805_C/A | ENSG00000172795 | ENST00000513585 | missense_variant | 96 | 98 | 33 | S/Y | tCc/tAc | rs61746772 | MODERATE | deleterious_low_confidence (0) | unknown (0) | FE164 | 2 |
| DCTN5 | 612962 | chr16_23657502_G/A | ENSG00000166847 | ENST00000563188 | missense_variant,NMD_transcript_variant | 164 | 134 | 45 | R/H | cGc/cAc | rs143714353 | MODERATE | tolerated (1) | benign (0.035) | FE154 | 1 |
| DDHD1 | 614603 | chr14_53096146_G/A | ENSG00000100523 | ENST00000323669 | missense_variant | 392 | 392 | 131 | A/V | gCg/gTg | rs144016382 | MODERATE | tolerated (0.13) | benign (0.013) | FE187 | 1 |
| DDN | 610588 | chr12_48998401_G/C | ENSG00000181418 | ENST00000421952 | missense_variant | 563 | 475 | 159 | L/V | Ctc/Gtc | rs202154840 | MODERATE | tolerated_low_confidence (0.05) | benign (0.081) | FE154 | 1 |

| | | | | | | | | | | | | | | | | |
|---|---|---|---|---|---|---|---|---|---|---|---|---|---|---|---|---|
| DDX19A | NA | chr16_70361466_C/T | ENSG00000168872 | ENST00000569319 | missense_variant,NMD_transcript_variant | 293 | 206 | 69 | P/L | cCt/cTt | rs1055783 | MODERATE | - | unknown (0) | FE190 | 1 |
| DDX20 | 606168 | chr1_111766100_A/G | ENSG00000064703 | ENST00000369702 | missense_variant | 1700 | 1676 | 559 | N/S | aAc/aGc | rs41310098 | MODERATE | tolerated (0.35) | benign (0.001) | FE136 | 1 |
| DDX20 | 606168 | chr1_111766100_A/G | ENSG00000064703 | ENST00000475700 | missense_variant | 2858 | 500 | 167 | N/S | aAc/aGc | rs41310098 | MODERATE | tolerated (0.29) | benign (0.005) | FE136 | 1 |
| DEPDC5 | 614191 | chr22_31822741_C/A | ENSG00000100150 | ENST00000382112 | missense_variant | 2141 | 2055 | 685 | F/L | ttC/ttA | rs61731667 | MODERATE | tolerated (0.69) | benign (0.026) | FE130 | 1 |
| DEPDC5 | 614191 | chr22_31822741_C/A | ENSG00000100150 | ENST00000382112 | missense_variant | 2141 | 2055 | 685 | F/L | ttC/ttA | rs61731667 | MODERATE | tolerated (0.69) | benign (0.026) | FE136 | 1 |
| DGKA | 125855 | chr12_55938925_A/G | ENSG00000065357 | ENST00000331886 | missense_variant | 674 | 410 | 137 | K/R | aAa/aGa | rs149792147 | MODERATE | tolerated (1) | benign (0) | FE106 | 1 |
| DIP2B | 611379 | chr12_50714504_T/A | ENSG00000066084 | ENST00000301180 | missense_variant | 2915 | 2759 | 920 | L/H | cTc/cAc | - | MODERATE | tolerated (0.18) | benign (0.001) | FE190 | 1 |
| DLG1 | 601014 | chr3_197194489_T/C | ENSG00000075711 | ENST00000357674 | missense_variant | 639 | 419 | 140 | K/R | aAg/aGg | rs1802668 | MODERATE | tolerated (0.4) | benign (0.013) | FE136 | 1 |
| DLG1 | 601014 | chr3_197194489_T/C | ENSG00000075711 | ENST00000419227 | missense_variant,NMD_transcript_variant | 609 | 419 | 140 | K/R | aAg/aGg | rs1802668 | MODERATE | tolerated (0.26) | benign (0.05) | FE136 | 1 |
| DLG1 | 601014 | chr3_197194489_T/C | ENSG00000075711 | ENST00000436682 | missense_variant | 732 | 419 | 140 | K/R | aAg/aGg | rs1802668 | MODERATE | tolerated_low_confidence (0.87) | probably_damaging (0.994) | FE136 | 1 |
| DLG1 | 601014 | chr3_197194489_T/C | ENSG00000075711 | ENST00000419354 | missense_variant | 706 | 419 | 140 | K/R | aAg/aGg | rs1802668 | MODERATE | tolerated_low_confidence (0.25) | benign (0.043) | FE136 | 1 |
| DLG1 | 601014 | chr3_197194489_T/C | ENSG00000075711 | ENST00000392382 | missense_variant | 493 | 419 | 140 | K/R | aAg/aGg | rs1802668 | MODERATE | tolerated (0.4) | benign (0.027) | FE136 | 1 |
| DLG1 | 601014 | chr3_197194489_T/C | ENSG00000075711 | ENST00000346964 | missense_variant | 609 | 419 | 140 | K/R | aAg/aGg | rs1802668 | MODERATE | tolerated_low_confidence (0.26) | benign (0.027) | FE136 | 1 |
| DLG1 | 601014 | chr3_197194489_T/C | ENSG00000075711 | ENST00000469073 | missense_variant,NMD_transcript_variant | 604 | 293 | 98 | K/R | aAg/aGg | rs1802668 | MODERATE | tolerated_low_confidence (0.2) | possibly_damaging (0.818) | FE136 | 1 |
| DLG1 | 601014 | chr3_197194489_T/C | ENSG00000075711 | ENST00000419553 | missense_variant,NMD_transcript_variant | 323 | 323 | 108 | K/R | aAg/aGg | rs1802668 | MODERATE | tolerated (0.26) | benign (0.01) | FE136 | 1 |
| DLG1 | 601014 | chr3_197194489_T/C | ENSG00000075711 | ENST00000422288 | missense_variant | 477 | 419 | 140 | K/R | aAg/aGg | rs1802668 | MODERATE | tolerated (0.21) | benign (0.006) | FE136 | 1 |
| DLG1 | 601014 | chr3_197194489_T/C | ENSG00000075711 | ENST00000655488 | missense_variant | 620 | 419 | 140 | K/R | aAg/aGg | rs1802668 | MODERATE | tolerated (0.4) | benign (0.027) | FE136 | 1 |
| DLG1 | 601014 | chr3_197194489_T/C | ENSG00000075711 | ENST00000661808 | missense_variant | 569 | 131 | 44 | K/R | aAg/aGg | rs1802668 | MODERATE | tolerated_low_confidence (0.16) | probably_damaging (0.927) | FE136 | 1 |
| DLG1 | 601014 | chr3_197194489_T/C | ENSG00000075711 | ENST00000662727 | missense_variant | 732 | 419 | 140 | K/R | aAg/aGg | rs1802668 | MODERATE | tolerated (0.3) | benign (0.073) | FE136 | 1 |
| DLG1 | 601014 | chr3_197194489_T/C | ENSG00000075711 | ENST00000663148 | missense_variant | 473 | 419 | 140 | K/R | aAg/aGg | rs1802668 | MODERATE | tolerated (0.4) | benign (0.027) | FE136 | 1 |
| DLG1 | 601014 | chr3_197194489_T/C | ENSG00000075711 | ENST00000664564 | missense_variant,NMD_transcript_variant | 612 | 419 | 140 | K/R | aAg/aGg | rs1802668 | MODERATE | tolerated (0.34) | benign (0.156) | FE136 | 1 |
| DLG1 | 601014 | chr3_197194489_T/C | ENSG00000075711 | ENST00000664991 | missense_variant | 573 | 419 | 140 | K/R | aAg/aGg | rs1802668 | MODERATE | tolerated (0.29) | benign (0.191) | FE136 | 1 |
| DLG1 | 601014 | chr3_197194489_T/C | ENSG00000075711 | ENST00000665728 | missense_variant,NMD_transcript_variant | 576 | 131 | 44 | K/R | aAg/aGg | rs1802668 | MODERATE | tolerated_low_confidence (0.06) | possibly_damaging (0.776) | FE136 | 1 |
| DLG1 | 601014 | chr3_197194489_T/C | ENSG00000075711 | ENST00000666007 | missense_variant | 606 | 419 | 140 | K/R | aAg/aGg | rs1802668 | MODERATE | tolerated (0.21) | benign (0.006) | FE136 | 1 |
| DLG1 | 601014 | chr3_197194489_T/C | ENSG00000075711 | ENST00000667104 | missense_variant | 730 | 419 | 140 | K/R | aAg/aGg | rs1802668 | MODERATE | tolerated (0.21) | benign (0.007) | FE136 | 1 |
| DLG1 | 601014 | chr3_197194489_T/C | ENSG00000075711 | ENST00000667157 | missense_variant | 527 | 419 | 140 | K/R | aAg/aGg | rs1802668 | MODERATE | tolerated (0.4) | benign (0.013) | FE136 | 1 |
| DLG1 | 601014 | chr3_197194489_T/C | ENSG00000075711 | ENST00000667971 | missense_variant | 631 | 419 | 140 | K/R | aAg/aGg | rs1802668 | MODERATE | tolerated (0.3) | benign (0.03) | FE136 | 1 |
| DLG1 | 601014 | chr3_197194489_T/C | ENSG00000075711 | ENST00000669332 | missense_variant,NMD_transcript_variant | 436 | 131 | 44 | K/R | aAg/aGg | rs1802668 | MODERATE | tolerated_low_confidence (0.31) | possibly_damaging (0.492) | FE136 | 1 |
| DLG1 | 601014 | chr3_197194489_T/C | ENSG00000075711 | ENST00000392380 | missense_variant | 511 | 419 | 140 | K/R | aAg/aGg | rs1802668 | MODERATE | tolerated (0.14) | benign (0.043) | FE136 | 1 |
| DLG1 | 601014 | chr3_197194489_T/C | ENSG00000075711 | ENST00000392381 | missense_variant,NMD_transcript_variant | 893 | 419 | 140 | K/R | aAg/aGg | rs1802668 | MODERATE | tolerated (0.27) | benign (0.082) | FE136 | 1 |
| DLG1 | 601014 | chr3_197194489_T/C | ENSG00000075711 | ENST00000670366 | missense_variant,NMD_transcript_variant | 516 | 131 | 44 | K/R | aAg/aGg | rs1802668 | MODERATE | tolerated_low_confidence (0.27) | benign (0.006) | FE136 | 1 |
| DLG1 | 601014 | chr3_197194489_T/C | ENSG00000075711 | ENST00000670455 | missense_variant | 750 | 419 | 140 | K/R | aAg/aGg | rs1802668 | MODERATE | tolerated (0.4) | benign (0.013) | FE136 | 1 |
| DLG1 | 601014 | chr3_197194489_T/C | ENSG00000075711 | ENST00000670935 | missense_variant | 708 | 419 | 140 | K/R | aAg/aGg | rs1802668 | MODERATE | tolerated_low_confidence (0.25) | benign (0.043) | FE136 | 1 |
| DLG1 | 601014 | chr3_197194489_T/C | ENSG00000075711 | ENST00000671185 | missense_variant | 631 | 419 | 140 | K/R | aAg/aGg | rs1802668 | MODERATE | tolerated (0.4) | benign (0.027) | FE136 | 1 |
| DLG1 | 601014 | chr3_197194489_T/C | ENSG00000075711 | ENST00000657381 | missense_variant | 538 | 419 | 140 | K/R | aAg/aGg | rs1802668 | MODERATE | tolerated (0.4) | benign (0.024) | FE136 | 1 |
| DLG1 | 601014 | chr3_197194489_T/C | ENSG00000075711 | ENST00000658155 | missense_variant | 212 | 131 | 44 | K/R | aAg/aGg | rs1802668 | MODERATE | tolerated_low_confidence (0.27) | benign (0.012) | FE136 | 1 |
| DLG1 | 601014 | chr3_197194489_T/C | ENSG00000075711 | ENST00000448528 | missense_variant | 609 | 419 | 140 | K/R | aAg/aGg | rs1802668 | MODERATE | tolerated_low_confidence (0.25) | benign (0.043) | FE136 | 1 |
| DLG1 | 601014 | chr3_197194489_T/C | ENSG00000075711 | ENST00000450955 | missense_variant | 419 | 419 | 140 | K/R | aAg/aGg | rs1802668 | MODERATE | tolerated (0.4) | benign (0.013) | FE136 | 1 |
| DLG1 | 601014 | chr3_197194489_T/C | ENSG00000075711 | ENST00000453607 | missense_variant | 304 | 131 | 44 | K/R | aAg/aGg | rs1802668 | MODERATE | tolerated (0.26) | benign (0.074) | FE136 | 1 |
| DLG1 | 601014 | chr3_197194489_T/C | ENSG00000075711 | ENST00000456699 | missense_variant | 511 | 419 | 140 | K/R | aAg/aGg | rs1802668 | MODERATE | tolerated (0.18) | possibly_damaging (0.717) | FE136 | 1 |
| DLG1 | 601014 | chr3_197194489_T/C | ENSG00000075711 | ENST00000661013 | missense_variant | 472 | 419 | 140 | K/R | aAg/aGg | rs1802668 | MODERATE | tolerated (0.4) | benign (0.024) | FE136 | 1 |
| DLG1 | 601014 | chr3_197194489_T/C | ENSG00000075711 | ENST00000654733 | missense_variant | 565 | 131 | 44 | K/R | aAg/aGg | rs1802668 | MODERATE | tolerated_low_confidence (0.25) | benign (0.381) | FE136 | 1 |

| | | | | | | | | | | | | | | | | |
|---|---|---|---|---|---|---|---|---|---|---|---|---|---|---|---|---|
| DLG1 | 601014 | chr3_197194489_T/C | ENSG00000075711 | ENST00000654737 | missense_variant | 601 | 419 | 140 | K/R | aAg/aGg | rs1802668 | MODERATE | tolerated (0.21) | benign (0.029) | FE136 | 1 |
| DLG1 | 601014 | chr3_197194489_T/C | ENSG00000075711 | ENST00000661453 | missense_variant | 778 | 419 | 140 | K/R | aAg/aGg | rs1802668 | MODERATE | tolerated (0.4) | benign (0.027) | FE136 | 1 |
| DLG1 | 601014 | chr3_197194489_T/C | ENSG00000075711 | ENST00000656087 | missense_variant | 587 | 419 | 140 | K/R | aAg/aGg | rs1802668 | MODERATE | tolerated (0.3) | benign (0.073) | FE136 | 1 |
| DLG1 | 601014 | chr3_197194489_T/C | ENSG00000075711 | ENST00000656428 | missense_variant | 597 | 293 | 98 | K/R | aAg/aGg | rs1802668 | MODERATE | tolerated_low_confidence (0.16) | possibly_damaging (0.818) | FE136 | 1 |
| DLG1 | 601014 | chr3_197194489_T/C | ENSG00000075711 | ENST00000656944 | missense_variant | 591 | 419 | 140 | K/R | aAg/aGg | rs1802668 | MODERATE | tolerated (0.4) | benign (0.024) | FE136 | 1 |
| DLG1 | 601014 | chr3_197194489_T/C | ENSG00000075711 | ENST00000659716 | missense_variant | 527 | 419 | 140 | K/R | aAg/aGg | rs1802668 | MODERATE | tolerated (0.4) | benign (0.027) | FE136 | 1 |
| DLG1 | 601014 | chr3_197194489_T/C | ENSG00000075711 | ENST00000660898 | missense_variant | 631 | 419 | 140 | K/R | aAg/aGg | rs1802668 | MODERATE | tolerated_low_confidence (0.29) | benign (0.191) | FE136 | 1 |
| DLG1 | 601014 | chr3_197194489_T/C | ENSG00000075711 | ENST00000658701 | missense_variant | 736 | 419 | 140 | K/R | aAg/aGg | rs1802668 | MODERATE | tolerated (0.39) | benign (0.073) | FE136 | 1 |
| DLG1 | 601014 | chr3_197194489_T/C | ENSG00000075711 | ENST00000659221 | missense_variant | 626 | 419 | 140 | K/R | aAg/aGg | rs1802668 | MODERATE | tolerated (0.4) | benign (0.024) | FE136 | 1 |
| DLG1 | 601014 | chr3_197194489_T/C | ENSG00000075711 | ENST00000661336 | missense_variant | 645 | 419 | 140 | K/R | aAg/aGg | rs1802668 | MODERATE | tolerated (0.24) | benign (0.02) | FE136 | 1 |
| DLG1 | 601014 | chr3_197194489_T/C | ENSG00000075711 | ENST00000669714 | missense_variant,NMD_transcript_variant | 582 | 131 | 44 | K/R | aAg/aGg | rs1802668 | MODERATE | tolerated_low_confidence (0.23) | benign (0.082) | FE136 | 1 |
| DLG1 | 601014 | chr3_197194489_T/C | ENSG00000075711 | ENST00000661229 | missense_variant,NMD_transcript_variant | 393 | 395 | 132 | K/R | aAg/aGg | rs1802668 | MODERATE | tolerated (0.2) | benign (0.024) | FE136 | 1 |
| DLG1 | 601014 | chr3_197194489_T/C | ENSG00000075711 | ENST00000669565 | missense_variant | 631 | 419 | 140 | K/R | aAg/aGg | rs1802668 | MODERATE | tolerated (0.4) | benign (0.013) | FE136 | 1 |
| DLST | 126063 | chr14_74893363_C/T | ENSG00000119689 | ENST00000334220 | missense_variant | 649 | 611 | 204 | P/L | cCc/cTc | rs142872233 | MODERATE | tolerated (0.06) | possibly_damaging (0.454) | FE194 | 1 |
| DLST | 126063 | chr14_74893363_C/T | ENSG00000119689 | ENST00000554806 | missense_variant | 560 | 560 | 187 | P/L | cCc/cTc | rs142872233 | MODERATE | tolerated (0.12) | possibly_damaging (0.454) | FE194 | 1 |
| DNLZ | NA | chr9_136363600_G/A | ENSG00000213221 | ENST00000371738 | missense_variant | 145 | 115 | 39 | R/W | Cgg/Tgg | rs759742424 | MODERATE | deleterious (0) | benign (0.001) | FE194 | 1 |
| DNLZ | NA | chr9_136363600_G/A | ENSG00000213221 | ENST00000371739 | missense_variant | 120 | 115 | 39 | R/W | Cgg/Tgg | rs759742424 | MODERATE | deleterious_low_confidence (0) | benign (0.005) | FE194 | 1 |
| DOLK | 610746 | chr9_128946342_T/C | ENSG00000175283 | ENST00000372586 | missense_variant | 1262 | 962 | 321 | K/R | aAg/aGg | - | MODERATE | tolerated (0.23) | benign (0.033) | FE154 | 1 |
| DOLK | 610746 | chr9_128946225_T/C | ENSG00000175283 | ENST00000372586 | missense_variant | 1379 | 1079 | 360 | Y/C | tAt/tGt | rs138453255 | MODERATE | tolerated (0.14) | possibly_damaging (0.784) | FE164 | 1 |
| DYNC1LI1 | 615890 | chr3_32545029_C/T | ENSG00000144635 | ENST00000424991 | missense_variant | 626 | 538 | 180 | V/I | Gta/Ata | rs138677120 | MODERATE | tolerated (0.13) | benign (0.006) | FE165 | 1 |
| DYNC1LI1 | 615890 | chr3_32545029_C/T | ENSG00000144635 | ENST00000413350 | missense_variant | 308 | 262 | 88 | V/I | Gta/Ata | rs138677120 | MODERATE | tolerated (0.09) | benign (0) | FE165 | 1 |
| DYNC1LI1 | 615890 | chr3_32545029_C/T | ENSG00000144635 | ENST00000273130 | missense_variant | 503 | 415 | 139 | V/I | Gta/Ata | rs138677120 | MODERATE | tolerated (0.19) | benign (0) | FE165 | 1 |
| DYRK2 | 603496 | chr12_67649940_C/T | ENSG00000127334 | ENST00000344096 | missense_variant | 582 | 193 | 65 | H/Y | Cac/Tac | rs557502236 | MODERATE | tolerated_low_confidence (0.1) | benign (0.001) | FE165 | 1 |
| E4F1 | 603022 | chr16_2233107_C/G | ENSG00000167967 | ENST00000301727 | missense_variant | 1003 | 980 | 327 | A/G | gCc/gGc | - | MODERATE | tolerated (0.2) | benign (0.358) | FE106 | 1 |
| E4F1 | 603022 | chr16_2233107_C/G | ENSG00000167967 | ENST00000564139 | missense_variant | 1000 | 980 | 327 | A/G | gCc/gGc | - | MODERATE | tolerated (0.14) | probably_damaging (0.999) | FE106 | 1 |
| E4F1 | 603022 | chr16_2233107_C/G | ENSG00000167967 | ENST00000565090 | missense_variant | 1014 | 980 | 327 | A/G | gCc/gGc | - | MODERATE | tolerated (0.14) | possibly_damaging (0.839) | FE106 | 1 |
| EAF1 | 608315 | chr3_15427876_A/G | ENSG00000144597 | ENST00000396842 | missense_variant | 279 | 97 | 33 | I/V | Att/Gtt | rs41284029 | MODERATE | tolerated (0.9) | benign (0.009) | FE190 | 1 |
| EAF1 | 608315 | chr3_15427876_A/G | ENSG00000144597 | ENST00000396842 | missense_variant | 279 | 97 | 33 | I/V | Att/Gtt | rs41284029 | MODERATE | tolerated (0.9) | benign (0.009) | FE193 | 1 |
| EAF1 | 608315 | chr3_15427876_A/G | ENSG00000144597 | ENST00000449565 | missense_variant,NMD_transcript_variant | 279 | 97 | 33 | I/V | Att/Gtt | rs41284029 | MODERATE | tolerated (0.67) | benign (0.001) | FE190 | 1 |
| EAF1 | 608315 | chr3_15427876_A/G | ENSG00000144597 | ENST00000449565 | missense_variant,NMD_transcript_variant | 279 | 97 | 33 | I/V | Att/Gtt | rs41284029 | MODERATE | tolerated (0.67) | benign (0.001) | FE193 | 1 |
| ECE1 | 600423 | chr1_21217754_T/A | ENSG00000117298 | ENST00000649812 | missense_variant | 2680 | 2486 | 829 | E/V | gAa/gTa | rs74343247 | MODERATE | - | unknown (0) | FE164 | 1 |
| ECE1 | 600423 | chr1_21217842_G/A | ENSG00000117298 | ENST00000649812 | stop_gained | 2592 | 2398 | 800 | Q/* | Cag/Tag | rs1008867347 | HIGH | - | - | FE187 | 1 |
| ECEL1 | 605896 | chr2_232484141_C/G | ENSG00000171551 | ENST00000304546 | missense_variant | 1484 | 1267 | 423 | E/Q | Gag/Cag | rs41265123,COSV58812016 | MODERATE | tolerated (0.22) | possibly_damaging (0.516) | FE130 | 1 |
| ECEL1 | 605896 | chr2_232484141_C/G | ENSG00000171551 | ENST00000409941 | missense_variant | 1267 | 1267 | 423 | E/Q | Gag/Cag | rs41265123,COSV58812016 | MODERATE | tolerated (0.22) | benign (0.185) | FE130 | 1 |
| EGLN1 | 606425 | chr1_231421578_G/A | ENSG00000135766 | ENST00000366641 | missense_variant | 710 | 311 | 104 | S/F | tCc/tTc | rs551207815 | MODERATE | tolerated (0.36) | benign (0.02) | FE154 | 1 |
| EIF3I | 603911 | chr1_32228574_A/G | ENSG00000084623 | ENST00000373586 | missense_variant | 676 | 604 | 202 | M/V | Atg/Gtg | rs202239183 | MODERATE | deleterious (0.04) | possibly_damaging (0.542) | FE106 | 1 |
| EIF5B | 606086 | chr2_99361611_A/T | ENSG00000158417 | ENST00000617677 | missense_variant | 894 | 710 | 237 | E/V | gAg/gTg | rs201583340 | MODERATE | deleterious_low_confidence (0) | probably_damaging (0.996) | FE136 | 1 |
| EIF5B | 606086 | chr2_99361611_A/T | ENSG00000158417 | ENST00000289371 | missense_variant | 876 | 710 | 237 | E/V | gAg/gTg | rs201583340 | MODERATE | deleterious_low_confidence (0) | probably_damaging (0.991) | FE136 | 1 |
| ELMO1 | 606420 | chr7_37315930_C/T | ENSG00000155849 | ENST00000310758 | missense_variant | 757 | 109 | 37 | V/I | Gtc/Atc | rs148906394 | MODERATE | tolerated (0.32) | benign (0.074) | FE130 | 1 |
| ELP5 | 615019 | chr17_7252983_C/G | ENSG00000170291 | ENST00000396627 | missense_variant | 404 | 221 | 74 | S/C | tCt/tGt | - | MODERATE | deleterious (0.02) | possibly_damaging (0.759) | FE164 | 1 |
| ELP5 | 615019 | chr17_7252983_C/G | ENSG00000170291 | ENST00000396628 | missense_variant | 438 | 221 | 74 | S/C | tCt/tGt | - | MODERATE | deleterious (0.02) | possibly_damaging (0.759) | FE164 | 1 |
| ELP5 | 615019 | chr17_7252983_C/G | ENSG00000170291 | ENST00000354429 | missense_variant | 328 | 221 | 74 | S/C | tCt/tGt | - | MODERATE | deleterious (0.02) | possibly_damaging (0.759) | FE164 | 1 |

| | | | | | | | | | | | | | | | | |
|---|---|---|---|---|---|---|---|---|---|---|---|---|---|---|---|---|
| ELP5 | 615019 | chr17_7252983_C/G | ENSG00000170291 | ENST00000356683 | missense_variant | 435 | 221 | 74 | S/C | tCt/tGt | - | MODERATE | deleterious (0.02) | possibly_damaging (0.823) | FE164 | 1 |
| ELP5 | 615019 | chr17_7252983_C/G | ENSG00000170291 | ENST00000571146 | missense_variant,NMD_transcript_variant | 127 | 128 | 43 | S/C | tCt/tGt | - | MODERATE | deleterious (0.02) | probably_damaging (0.917) | FE164 | 1 |
| ELP5 | 615019 | chr17_7252983_C/G | ENSG00000170291 | ENST00000574993 | missense_variant | 438 | 221 | 74 | S/C | tCt/tGt | - | MODERATE | deleterious (0.02) | possibly_damaging (0.823) | FE164 | 1 |
| ELP5 | 615019 | chr17_7252983_C/G | ENSG00000170291 | ENST00000570322 | missense_variant | 199 | 200 | 67 | S/C | tCt/tGt | - | MODERATE | deleterious (0.04) | probably_damaging (0.959) | FE164 | 1 |
| ELP5 | 615019 | chr17_7252983_C/G | ENSG00000170291 | ENST00000570500 | missense_variant | 407 | 221 | 74 | S/C | tCt/tGt | - | MODERATE | deleterious (0.03) | possibly_damaging (0.759) | FE164 | 1 |
| ELP5 | 615019 | chr17_7252983_C/G | ENSG00000170291 | ENST00000573657 | missense_variant | 348 | 221 | 74 | S/C | tCt/tGt | - | MODERATE | deleterious (0.01) | possibly_damaging (0.847) | FE164 | 1 |
| ELP5 | 615019 | chr17_7252983_C/G | ENSG00000170291 | ENST00000573699 | missense_variant | 456 | 221 | 74 | S/C | tCt/tGt | - | MODERATE | deleterious (0.04) | possibly_damaging (0.759) | FE164 | 1 |
| ELP5 | 615019 | chr17_7252983_C/G | ENSG00000170291 | ENST00000576496 | missense_variant | 169 | 170 | 57 | S/C | tCt/tGt | - | MODERATE | tolerated (0.05) | probably_damaging (0.94) | FE164 | 1 |
| ELP5 | 615019 | chr17_7252983_C/G | ENSG00000170291 | ENST00000573513 | missense_variant | 357 | 221 | 74 | S/C | tCt/tGt | - | MODERATE | deleterious (0.05) | possibly_damaging (0.759) | FE164 | 1 |
| ELP5 | 615019 | chr17_7252983_C/G | ENSG00000170291 | ENST00000574255 | missense_variant | 325 | 221 | 74 | S/C | tCt/tGt | - | MODERATE | deleterious (0.01) | possibly_damaging (0.847) | FE164 | 1 |
| EML1 | 602033 | chr14_99865619_A/G | ENSG00000066629 | ENST00000334192 | missense_variant | 490 | 356 | 119 | K/R | aAa/aGa | rs199650308 | MODERATE | tolerated (0.25) | benign (0.015) | FE194 | 1 |
| ENDOG | 600440 | chr9_128818847_C/T | ENSG00000167136 | ENST00000372642 | missense_variant | 348 | 163 | 55 | P/S | Ccc/Tcc | rs938890042 | MODERATE | tolerated (0.4) | benign (0.015) | FE106 | 1 |
| ENDOG | 600440 | chr9_128818882_G/C | ENSG00000167136 | ENST00000372642 | missense_variant | 383 | 198 | 66 | K/N | aaG/aaC | rs200885264 | MODERATE | deleterious (0) | probably_damaging (0.961) | FE130 | 1 |
| EOMES | 604615 | chr3_27721936_G/GCGGCGC/C | ENSG00000163508 | ENST00000295743 | inframe_insertion | 563 | 359 | 120 | A/GAA | gGCGCCGCc | - | MODERATE | - | - | FE106 | 1 |
| EOMES | 604615 | chr3_27721936_G/GCGGCGC/C | ENSG00000163508 | ENST00000449599 | inframe_insertion | 388 | 359 | 120 | A/GAA | gGCGCCGCc | - | MODERATE | - | - | FE106 | 1 |
| EOMES | 604615 | chr3_27721936_G/GCGGCGC/C | ENSG00000163508 | ENST00000449599 | inframe_insertion | 388 | 359 | 120 | A/GAA | gGCGCCGCc | - | MODERATE | - | - | FE130 | 1 |
| EOMES | 604615 | chr3_27721936_G/GCGGCGC/C | ENSG00000163508 | ENST00000295743 | inframe_insertion | 563 | 359 | 120 | A/GAA | gGCGCCGCc | - | MODERATE | - | - | FE164 | 1 |
| EOMES | 604615 | chr3_27721936_G/GCGGCGC/C | ENSG00000163508 | ENST00000295743 | inframe_insertion | 563 | 359 | 120 | A/GAA | gGCGCCGCc | - | MODERATE | - | - | FE136 | 1 |
| EOMES | 604615 | chr3_27721936_G/GCGGCGC/C | ENSG00000163508 | ENST00000295743 | inframe_insertion | 563 | 359 | 120 | A/GAA | gGCGCCGCc | - | MODERATE | - | - | FE130 | 1 |
| EOMES | 604615 | chr3_27721936_G/GCGGCGC/C | ENSG00000163508 | ENST00000449599 | inframe_insertion | 388 | 359 | 120 | A/GAA | gGCGCCGCc | - | MODERATE | - | - | FE165 | 1 |
| EOMES | 604615 | chr3_27721936_G/GCGGCGC/C | ENSG00000163508 | ENST00000295743 | inframe_insertion | 563 | 359 | 120 | A/GAA | gGCGCCGCc | - | MODERATE | - | - | FE187 | 1 |
| EOMES | 604615 | chr3_27721936_G/GCGGCGC/C | ENSG00000163508 | ENST00000295743 | inframe_insertion | 563 | 359 | 120 | A/GAA | gGCGCCGCc | - | MODERATE | - | - | FE193 | 1 |
| EOMES | 604615 | chr3_27721936_G/GCGGCGC/C | ENSG00000163508 | ENST00000449599 | inframe_insertion | 388 | 359 | 120 | A/GAA | gGCGCCGCc | - | MODERATE | - | - | FE136 | 1 |
| EOMES | 604615 | chr3_27721936_G/GCGGCGC/C | ENSG00000163508 | ENST00000449599 | inframe_insertion | 388 | 359 | 120 | A/GAA | gGCGCCGCc | - | MODERATE | - | - | FE187 | 1 |
| EOMES | 604615 | chr3_27721936_G/GCGGCGC/C | ENSG00000163508 | ENST00000449599 | inframe_insertion | 388 | 359 | 120 | A/GAA | gGCGCCGCc | - | MODERATE | - | - | FE193 | 1 |
| EOMES | 604615 | chr3_27721936_G/GCGGCGC/C | ENSG00000163508 | ENST00000295743 | inframe_insertion | 563 | 359 | 120 | A/GAA | gGCGCCGCc | - | MODERATE | - | - | FE165 | 1 |
| EOMES | 604615 | chr3_27721936_G/GCGGCGC/C | ENSG00000163508 | ENST00000449599 | inframe_insertion | 388 | 359 | 120 | A/GAA | gGCGCCGCc | - | MODERATE | - | - | FE164 | 1 |
| EOMES | 604615 | chr3_27721936_G/GCGGCGC/C | ENSG00000163508 | ENST00000449599 | inframe_insertion | 388 | 359 | 120 | A/GAA | gGCGCCGCc | - | MODERATE | - | - | FE194 | 1 |
| EOMES | 604615 | chr3_27721936_G/GCGGCGC/C | ENSG00000163508 | ENST00000295743 | inframe_insertion | 563 | 359 | 120 | A/GAA | gGCGCCGCc | - | MODERATE | - | - | FE194 | 1 |
| EP300 | 602700 | chr22_41178379_A/C | ENSG00000100393 | ENST00000263253 | missense_variant | 7081 | 6668 | 2223 | Q/P | cAg/cCg | rs1046088,COSV54331579 | MODERATE | tolerated_low_confidence (0.17) | benign (0) | FE164 | 1 |
| EP300 | 602700 | chr22_41178379_A/C | ENSG00000100393 | ENST00000263253 | missense_variant | 7081 | 6668 | 2223 | Q/P | cAg/cCg | rs1046088,COSV54331579 | MODERATE | tolerated_low_confidence (0.17) | benign (0) | FE190 | 1 |
| EP300 | 602700 | chr22_41178379_A/C | ENSG00000100393 | ENST00000674155 | missense_variant | 6590 | 6590 | 2197 | Q/P | cAg/cCg | rs1046088,COSV54331579 | MODERATE | tolerated_low_confidence (0.21) | probably_damaging (0.915) | FE164 | 1 |
| EP300 | 602700 | chr22_41178379_A/C | ENSG00000100393 | ENST00000674155 | missense_variant | 6590 | 6590 | 2197 | Q/P | cAg/cCg | rs1046088,COSV54331579 | MODERATE | tolerated_low_confidence (0.21) | probably_damaging (0.915) | FE190 | 1 |
| EPC2 | 611000 | chr2_148771256_C/T | ENSG00000135999 | ENST00000258484 | missense_variant | 1856 | 1589 | 530 | P/L | cCa/cTa | - | MODERATE | deleterious (0) | probably_damaging (0.994) | FE136 | 1 |
| EPHB1 | 600600 | chr3_135132960_C/T | ENSG00000154928 | ENST00000398015 | missense_variant | 1580 | 1208 | 403 | T/I | aCc/aTc | rs200876961 | MODERATE | tolerated (0.08) | possibly_damaging (0.8) | FE106 | 1 |
| ESCO1 | 609674 | chr18_21574272_T/C | ENSG00000141446 | ENST00000269214 | missense_variant | 1478 | 572 | 191 | N/S | aAt/aGt | rs35087820,CM0911485 | MODERATE | tolerated (0.18) | benign (0) | FE136 | 1 |
| EXOC7 | 608163 | chr17_76101230_G/C | ENSG00000182473 | ENST00000406660 | stop_gained | 500 | 458 | 153 | S/* | tCa/tGa | rs113648734 | HIGH | - | - | FE106 | 1 |
| FAM193B | 615813 | chr5_177536398_G/C | ENSG00000146067 | ENST00000514747 | missense_variant | 1141 | 1036 | 346 | L/V | Ctc/Gtc | - | MODERATE | tolerated (0.21) | benign (0.011) | FE106 | 1 |
| FAM98B | 616142 | chr15_38484608_T/A | ENSG00000171262 | ENST00000559431 | missense_variant | 275 | 275 | 92 | V/E | gTg/gAg | rs374461368 | MODERATE | deleterious_low_confidence (0) | unknown (0) | FE154 | 1 |
| FANCE | 613976 | chr6_35460568_C/T | ENSG00000112039 | ENST00000229769 | missense_variant | 1541 | 1333 | 445 | P/S | Ccc/Tcc | rs141551053 | MODERATE | tolerated (0.12) | benign (0.033) | FE165 | 1 |
| FANCE | 613976 | chr6_35460568_C/T | ENSG00000112039 | ENST00000648059 | missense_variant,NMD_transcript_variant | 1523 | 1333 | 445 | P/S | Ccc/Tcc | rs141551053 | MODERATE | tolerated (0.11) | benign (0.033) | FE165 | 1 |
| FBLN1 | 135820 | chr22_45523158_G/A | ENSG00000077942 | ENST00000402984 | missense_variant | 356 | 253 | 85 | A/T | Gca/Aca | rs150868662 | MODERATE | tolerated_low_confidence (0.17) | benign (0) | FE106 | 1 |

| | | | | | | | | | | | | | | | | |
|---|---|---|---|---|---|---|---|---|---|---|---|---|---|---|---|---|
| FBXO30 | 609101 | chr6_145805444_G/A | ENSG00000118496 | ENST00000237281 | missense_variant | 1171 | 962 | 321 | S/L | tCa/tTa | rs148771547 | MODERATE | tolerated (0.31) | benign (0.006) | FE136 | 1 |
| FBXO30 | 609101 | chr6_145805444_G/A | ENSG00000118496 | ENST00000237281 | missense_variant | 1171 | 962 | 321 | S/L | tCa/tTa | rs148771547 | MODERATE | tolerated (0.31) | benign (0.006) | FE194 | 1 |
| FITM2 | 612029 | chr20_44311053_C/T | ENSG00000197296 | ENST00000396825 | missense_variant | 150 | 96 | 32 | M/I | atG/atA | rs148377517,COSV67679219 | MODERATE | tolerated (0.46) | benign (0.015) | FE136 | 1 |
| FITM2 | 612029 | chr20_44306641_T/C | ENSG00000197296 | ENST00000396825 | missense_variant | 827 | 773 | 258 | D/G | gAt/gGt | rs142318812 | MODERATE | tolerated (0.31) | benign (0.003) | FE154 | 1 |
| FLAD1 | 610595 | chr1_154992654_C/T | ENSG00000160688 | ENST00000295530 | stop_gained | 982 | 505 | 169 | Q/* | Cag/Tag | - | HIGH | - | - | FE130 | 1 |
| FLVCR2 | 610865 | chr14_75624635_C/T | ENSG00000119686 | ENST00000539311 | missense_variant | 329 | 220 | 74 | P/S | Ccc/Tcc | rs45479302 | MODERATE | deleterious (0.02) | probably_damaging (0.973) | FE194 | 1 |
| FLVCR2 | 610865 | chr14_75624635_C/T | ENSG00000119686 | ENST00000553587 | missense_variant | 185 | 79 | 27 | P/S | Ccc/Tcc | rs45479302 | MODERATE | deleterious (0.01) | possibly_damaging (0.813) | FE194 | 1 |
| FLVCR2 | 610865 | chr14_75624635_C/T | ENSG00000119686 | ENST00000555058 | missense_variant | 346 | 79 | 27 | P/S | Ccc/Tcc | rs45479302 | MODERATE | deleterious (0.01) | probably_damaging (0.956) | FE194 | 1 |
| FLVCR2 | 610865 | chr14_75624635_C/T | ENSG00000119686 | ENST00000556856 | missense_variant | 153 | 79 | 27 | P/S | Ccc/Tcc | rs45479302 | MODERATE | deleterious (0.02) | probably_damaging (0.999) | FE194 | 1 |
| FLVCR2 | 610865 | chr14_75624635_C/T | ENSG00000119686 | ENST00000238667 | missense_variant | 1188 | 835 | 279 | P/S | Ccc/Tcc | rs45479302 | MODERATE | deleterious (0.03) | probably_damaging (0.956) | FE194 | 1 |
| FMNL2 | 616285 | chr2_152549066_C/G | ENSG00000157827 | ENST00000288670 | missense_variant | 758 | 328 | 110 | L/V | Ctg/Gtg | rs189416564 | MODERATE | deleterious (0) | probably_damaging (0.969) | FE165 | 1 |
| FMNL2 | 616285 | chr2_152560918_G/T | ENSG00000157827 | ENST00000288670 | missense_variant | 909 | 479 | 160 | S/I | aGc/aTc | rs866373641,COSV56495414,COSV56491380 | MODERATE | tolerated (0.14) | benign (0.02) | FE136 | 1 |
| FMNL2 | 616285 | chr2_152560914_G/T | ENSG00000157827 | ENST00000288670 | stop_gained | 905 | 475 | 159 | E/* | Gag/Tag | rs750755379,COSV56492433 | HIGH | - | - | FE187 | 1 |
| FMNL2 | 616285 | chr2_152560918_G/T | ENSG00000157827 | ENST00000288670 | missense_variant | 909 | 479 | 160 | S/I | aGc/aTc | rs866373641,COSV56495414,COSV56491380 | MODERATE | tolerated (0.14) | benign (0.02) | FE165 | 1 |
| FMNL2 | 616285 | chr2_152560918_G/T | ENSG00000157827 | ENST00000288670 | missense_variant | 909 | 479 | 160 | S/I | aGc/aTc | rs866373641,COSV56495414,COSV56491380 | MODERATE | tolerated (0.14) | benign (0.02) | FE190 | 1 |
| FMNL2 | 616285 | chr2_152560914_G/T | ENSG00000157827 | ENST00000288670 | stop_gained | 905 | 475 | 159 | E/* | Gag/Tag | rs750755379,COSV56492433 | HIGH | - | - | FE165 | 1 |
| FMNL2 | 616285 | chr2_152560914_G/T | ENSG00000157827 | ENST00000288670 | stop_gained | 905 | 475 | 159 | E/* | Gag/Tag | rs750755379,COSV56492433 | HIGH | - | - | FE193 | 1 |
| FMNL2 | 616285 | chr2_152560914_G/T | ENSG00000157827 | ENST00000288670 | stop_gained | 905 | 475 | 159 | E/* | Gag/Tag | rs750755379,COSV56492433 | HIGH | - | - | FE190 | 1 |
| FMNL2 | 616285 | chr2_152560918_G/T | ENSG00000157827 | ENST00000288670 | missense_variant | 909 | 479 | 160 | S/I | aGc/aTc | rs866373641,COSV56495414,COSV56491380 | MODERATE | tolerated (0.14) | benign (0.02) | FE193 | 1 |
| FMNL2 | 616285 | chr2_152560918_G/T | ENSG00000157827 | ENST00000288670 | missense_variant | 909 | 479 | 160 | S/I | aGc/aTc | rs866373641,COSV56495414,COSV56491380 | MODERATE | tolerated (0.14) | benign (0.02) | FE187 | 1 |
| FMNL2 | 616285 | chr2_152560914_G/T | ENSG00000157827 | ENST00000288670 | stop_gained | 905 | 475 | 159 | E/* | Gag/Tag | rs750755379,COSV56492433 | HIGH | - | - | FE136 | 1 |
| FOXA1 | 602294 | chr14_37592342_G/C | ENSG00000129514 | ENST00000250448 | missense_variant | 719 | 442 | 148 | L/V | Ctg/Gtg | rs112819884 | MODERATE | deleterious (0.01) | benign (0.273) | FE130 | 1 |
| FOXE3 | 601094 | chr1_47416331_G/A | ENSG00000186790 | ENST00000335071 | missense_variant | 47 | 16 | 6 | D/N | Gac/Aac | rs765169217 | MODERATE | tolerated_low_confidence (0.11) | benign (0.068) | FE154 | 1 |
| FOXE3 | 601094 | chr1_47417213_A/G | ENSG00000186790 | ENST00000335071 | missense_variant | 929 | 898 | 300 | S/G | Agc/Ggc | rs552420470,COSV58633000 | MODERATE | tolerated (0.46) | benign (0) | FE190 | 1 |
| FRMPD3 | 301005 | chrX_107603212_C/T | ENSG00000147234 | ENST00000276185 | stop_gained | 5272 | 5272 | 1758 | Q/* | Cag/Tag | rs1235926839,COSV52187413 | HIGH | - | - | FE106 | 1 |
| FRMPD3 | 301005 | chrX_107603212_C/T | ENSG00000147234 | ENST00000276185 | stop_gained | 5272 | 5272 | 1758 | Q/* | Cag/Tag | rs1235926839,COSV52187413 | HIGH | - | - | FE165 | 1 |
| FTH1 | 134770 | chr11_61965469_T/C | ENSG00000167996 | ENST00000273550 | missense_variant | 370 | 161 | 54 | K/R | aAa/aGa | rs186448909,CM034816 | MODERATE | tolerated (0.37) | benign (0.005) | FE164 | 1 |
| FTH1 | 134770 | chr11_61967377_A/C | ENSG00000167996 | ENST00000273550 | missense_variant | 258 | 49 | 17 | S/A | Tca/Gca | - | MODERATE | tolerated (0.08) | benign (0.003) | FE136 | 1 |
| FUT7 | 602030 | chr9_137031531_G/A | ENSG00000180549 | ENST00000314412 | missense_variant | 305 | 208 | 70 | R/C | Cgc/Tgc | rs117125309 | MODERATE | deleterious (0.02) | possibly_damaging (0.849) | FE130 | 1 |
| GBF1 | 603698 | chr10_102382332_A/G | ENSG00000107862 | ENST00000369983 | missense_variant | 5836 | 5576 | 1859 | N/S | aAc/aGc | rs769397523 | MODERATE | deleterious_low_confidence (0) | benign (0.001) | FE193 | 1 |
| GBF1 | 603698 | chr10_102382332_A/G | ENSG00000107862 | ENST00000673650 | missense_variant | 5935 | 5675 | 1892 | N/S | aAc/aGc | rs769397523 | MODERATE | deleterious_low_confidence (0) | probably_damaging (0.998) | FE193 | 1 |
| GBF1 | 603698 | chr10_102382332_A/G | ENSG00000107862 | ENST00000674034 | missense_variant | 5911 | 5651 | 1884 | N/S | aAc/aGc | rs769397523 | MODERATE | deleterious_low_confidence (0) | probably_damaging (0.998) | FE193 | 1 |
| GDA | 139260 | chr9_72225752_G/A | ENSG00000119125 | ENST00000238018 | missense_variant | 999 | 790 | 264 | V/M | Gtg/Atg | rs61752956 | MODERATE | deleterious (0) | probably_damaging (0.975) | FE165 | 1 |
| GDF3 | 606522 | chr12_7690338_G/A | ENSG00000184344 | ENST00000329913 | missense_variant | 682 | 635 | 212 | S/L | tCa/tTa | rs372790667,CM1412384 | MODERATE | tolerated (0.79) | benign (0) | FE193 | 1 |
| GEMIN5 | 607005 | chr5_154917072_G/C | ENSG00000082516 | ENST00000285873 | missense_variant | 1859 | 1781 | 594 | P/R | cCa/cGa | rs142530738 | MODERATE | deleterious (0.04) | probably_damaging (0.999) | FE106 | 1 |
| GEMIN5 | 607005 | chr5_154917072_G/C | ENSG00000082516 | ENST00000285873 | missense_variant | 1859 | 1781 | 594 | P/R | cCa/cGa | rs142530738 | MODERATE | deleterious (0.04) | probably_damaging (0.999) | FE130 | 1 |
| GEMIN5 | 607005 | chr5_154938122_C/A | ENSG00000082516 | ENST00000285873 | missense_variant | 90 | 12 | 4 | E/D | gaG/gaT | rs535567224 | MODERATE | tolerated_low_confidence (0.07) | benign (0.3) | FE193 | 1 |
| GGA1 | 606004 | chr22_37623556_T/C | ENSG00000100083 | ENST00000343632 | missense_variant | 782 | 755 | 252 | L/P | cTg/cCg | - | MODERATE | deleterious (0) | probably_damaging (0.979) | FE190 | 1 |
| GID4 | 617699 | chr17_18039601_A/C | ENSG00000141034 | ENST00000268719 | missense_variant | 194 | 137 | 46 | H/P | cAc/cCc | rs1049247858 | MODERATE | tolerated_low_confidence (0.27) | benign (0) | FE187 | 1 |
| GJB5 | 604493 | chr1_34757613_G/A | ENSG00000189280 | ENST00000338513 | missense_variant | 456 | 283 | 95 | V/M | Gtg/Atg | rs137982501 | MODERATE | deleterious (0) | probably_damaging (0.999) | FE193 | 1 |
| GNB1L | 610778 | chr22_19788842_G/A | ENSG00000185838 | ENST00000403325 | missense_variant | 1272 | 851 | 284 | T/M | aCg/aTg | rs73148914 | MODERATE | deleterious (0.01) | benign (0.326) | FE190 | 1 |
| GNB1L | 610778 | chr22_19788842_G/A | ENSG00000185838 | ENST00000329517 | missense_variant | 1023 | 851 | 284 | T/M | aCg/aTg | rs73148914 | MODERATE | deleterious (0.01) | benign (0.326) | FE190 | 1 |

| | | | | | | | | | | | | | | | | |
|---|---|---|---|---|---|---|---|---|---|---|---|---|---|---|---|---|
| GNL2 | 609365 | chr1_37581532_G/A | ENSG00000134697 | ENST00000538069 | missense_variant | 274 | 275 | 92 | T/M | aCg/aTg | rs555289288 | MODERATE | deleterious (0.03) | unknown (0) | FE165 | 1 |
| GPX4 | 138322 | chr19_1106733_G/A | ENSG00000167468 | ENST00000585480 | missense_variant | 455 | 455 | 152 | R/Q | cGg/cAg | rs34865773 | MODERATE | deleterious_low_confidence (0) | unknown (0) | FE106 | 1 |
| GRM3 | 601115 | chr7_86838938_G/A | ENSG00000198822 | ENST00000361669 | missense_variant | 2523 | 1424 | 475 | G/D | gGt/gAt | rs17161026 | MODERATE | tolerated (0.41) | benign (0) | FE164 | 1 |
| GTPBP4 | NA | chr10_1015825_C/G | ENSG00000107937 | ENST00000360803 | missense_variant | 1727 | 1681 | 561 | P/A | Ccg/Gcg | rs41294962 | MODERATE | tolerated (0.4) | benign (0.005) | FE165 | 1 |
| HAUS3 | 613430 | chr4_2174697_A/G | ENSG00000214367 | ENST00000672725 | missense_variant,NMD_transcript_variant | 3415 | 2666 | 889 | F/S | tTt/tCt | rs138077251 | MODERATE | - | benign (0.163) | FE165 | 1 |
| HCN1 | 602780 | chr5_45695954_C/A | ENSG00000164588 | ENST00000303230 | missense_variant | 427 | 140 | 47 | G/V | gGg/gTg | rs544994462,CM145435,COSV57523921 | MODERATE | tolerated_low_confidence (0.14) | benign (0.084) | FE187 | 1 |
| HEATR3 | 614951 | chr16_50094737_A/G | ENSG00000155393 | ENST00000299192 | missense_variant | 1705 | 1543 | 515 | I/V | Ata/Gta | rs73580039 | MODERATE | tolerated (0.94) | benign (0) | FE154 | 1 |
| HECW2 | 617245 | chr2_196343657_G/A | ENSG00000138411 | ENST00000260983 | missense_variant,splice_region_variant | 656 | 400 | 134 | P/S | Ccg/Tcg | rs61752169 | MODERATE | tolerated (0.28) | benign (0.005) | FE154 | 1 |
| HERC4 | 609248 | chr10_67932719_C/T | ENSG00000148634 | ENST00000395198 | missense_variant | 2988 | 2740 | 914 | D/N | Gat/Aat | rs143083420 | MODERATE | tolerated (0.36) | benign (0.007) | FE165 | 1 |
| HERC4 | 609248 | chr10_67932719_C/T | ENSG00000148634 | ENST00000395198 | missense_variant | 2988 | 2740 | 914 | D/N | Gat/Aat | rs143083420 | MODERATE | tolerated (0.36) | benign (0.007) | FE190 | 1 |
| HERPUD2 | NA | chr7_35670253_T/C | ENSG00000122557 | ENST00000396081 | missense_variant | 1106 | 301 | 101 | R/G | Aga/Gga | rs140296640 | MODERATE | tolerated (0.31) | benign (0) | FE136 | 1 |
| HES3 | 609971 | chr1_6244394_A/G | ENSG00000173673 | ENST00000377898 | missense_variant | 107 | 29 | 10 | N/S | aAt/aGt | rs114198341 | MODERATE | deleterious (0) | probably_damaging (0.997) | FE136 | 1 |
| HHIP | 606178 | chr4_144707143_A/G | ENSG00000164161 | ENST00000296575 | missense_variant | 1560 | 1040 | 347 | E/G | gAa/gGa | - | MODERATE | deleterious (0) | possibly_damaging (0.856) | FE165 | 1 |
| HK2 | 601125 | chr2_74867692_G/T | ENSG00000159399 | ENST00000290573 | missense_variant | 737 | 283 | 95 | V/L | Gtg/Ttg | - | MODERATE | deleterious (0) | probably_damaging (0.993) | FE193 | 1 |
| HK2 | 601125 | chr2_74867692_G/T | ENSG00000159399 | ENST00000409174 | missense_variant | 389 | 199 | 67 | V/L | Gtg/Ttg | - | MODERATE | deleterious (0.01) | probably_damaging (0.993) | FE193 | 1 |
| HOXD9 | 142982 | chr2_176123331_C/A | ENSG00000128709 | ENST00000249499 | missense_variant | 613 | 563 | 188 | S/Y | tCc/tAc | rs34862292 | MODERATE | tolerated (0.24) | benign (0) | FE190 | 1 |
| HSF1 | 140580 | chr8_144312189_T/C | ENSG00000185122 | ENST00000400780 | missense_variant | 1254 | 1087 | 363 | S/P | Tcc/Ccc | rs1299053211,COSV60047043 | MODERATE | deleterious (0.03) | benign (0.398) | FE130 | 1 |
| HSF1 | 140580 | chr8_144312189_T/C | ENSG00000185122 | ENST00000528838 | missense_variant | 1241 | 1087 | 363 | S/P | Tcc/Ccc | rs1299053211,COSV60047043 | MODERATE | tolerated (0.08) | benign (0.003) | FE130 | 1 |
| HSF1 | 140580 | chr8_144312189_T/C | ENSG00000185122 | ENST00000530661 | missense_variant,NMD_transcript_variant | 119 | 119 | 40 | L/P | cTc/cCc | rs1299053211,COSV60047043 | MODERATE | - | unknown (0) | FE130 | 1 |
| HSF1 | 140580 | chr8_144312189_T/C | ENSG00000185122 | ENST00000400780 | missense_variant | 1254 | 1087 | 363 | S/P | Tcc/Ccc | rs1299053211,COSV60047043 | MODERATE | deleterious (0.03) | benign (0.398) | FE136 | 1 |
| HSF1 | 140580 | chr8_144312189_T/C | ENSG00000185122 | ENST00000528838 | missense_variant | 1241 | 1087 | 363 | S/P | Tcc/Ccc | rs1299053211,COSV60047043 | MODERATE | tolerated (0.08) | benign (0.003) | FE136 | 1 |
| HSF1 | 140580 | chr8_144312189_T/C | ENSG00000185122 | ENST00000530661 | missense_variant,NMD_transcript_variant | 119 | 119 | 40 | L/P | cTc/cCc | rs1299053211,COSV60047043 | MODERATE | - | unknown (0) | FE136 | 1 |
| HSF1 | 140580 | chr8_144312189_T/C | ENSG00000185122 | ENST00000530661 | missense_variant,NMD_transcript_variant | 119 | 119 | 40 | L/P | cTc/cCc | rs1299053211,COSV60047043 | MODERATE | - | unknown (0) | FE165 | 1 |
| HSF1 | 140580 | chr8_144312189_T/C | ENSG00000185122 | ENST00000400780 | missense_variant | 1254 | 1087 | 363 | S/P | Tcc/Ccc | rs1299053211,COSV60047043 | MODERATE | deleterious (0.03) | benign (0.398) | FE193 | 1 |
| HSF1 | 140580 | chr8_144312189_T/C | ENSG00000185122 | ENST00000400780 | missense_variant | 1254 | 1087 | 363 | S/P | Tcc/Ccc | rs1299053211,COSV60047043 | MODERATE | deleterious (0.03) | benign (0.398) | FE165 | 1 |
| HSF1 | 140580 | chr8_144312189_T/C | ENSG00000185122 | ENST00000528838 | missense_variant | 1241 | 1087 | 363 | S/P | Tcc/Ccc | rs1299053211,COSV60047043 | MODERATE | tolerated (0.08) | benign (0.003) | FE165 | 1 |
| HSF1 | 140580 | chr8_144312189_T/C | ENSG00000185122 | ENST00000528838 | missense_variant | 1241 | 1087 | 363 | S/P | Tcc/Ccc | rs1299053211,COSV60047043 | MODERATE | tolerated (0.08) | benign (0.003) | FE193 | 1 |
| HSF1 | 140580 | chr8_144312189_T/C | ENSG00000185122 | ENST00000530661 | missense_variant,NMD_transcript_variant | 119 | 119 | 40 | L/P | cTc/cCc | rs1299053211,COSV60047043 | MODERATE | - | unknown (0) | FE193 | 1 |
| HSPA14 | 610369 | chr10_14867875_A/C | ENSG00000187522 | ENST00000378372 | missense_variant | 1443 | 1346 | 449 | K/T | aAg/aCg | - | MODERATE | tolerated (0.3) | benign (0.174) | FE106 | 1 |
| HSPD1 | 118190 | chr2_197487080_C/G | ENSG00000144381 | ENST00000388968 | missense_variant | 1749 | 1688 | 563 | G/A | gGa/gCa | rs41265953,CM088535 | MODERATE | deleterious (0.04) | probably_damaging (0.981) | FE187 | 1 |
| HSPD1 | 118190 | chr2_197487080_C/G | ENSG00000144381 | ENST00000388968 | missense_variant | 1749 | 1688 | 563 | G/A | gGa/gCa | rs41265953,CM088535 | MODERATE | deleterious (0.04) | probably_damaging (0.981) | FE164 | 1 |
| HSPH1 | 610703 | chr13_31138435_C/T | ENSG00000120694 | ENST00000320027 | missense_variant | 2664 | 2342 | 781 | R/H | cGt/cAt | rs73171026 | MODERATE | tolerated (0.1) | benign (0.302) | FE106 | 1 |
| HTRA2 | 606441 | chr2_74532698_G/A | ENSG00000115317 | ENST00000258080 | missense_variant | 1825 | 1195 | 399 | G/S | Ggc/Agc | rs72470545,CM052355 | MODERATE | deleterious (0.02) | probably_damaging (0.936) | FE190 | 1 |
| HTRA2 | 606441 | chr2_74530221_T/C | ENSG00000115317 | ENST00000258080 | missense_variant | 845 | 215 | 72 | L/P | cTg/cCg | rs150047108,COSV51206225 | MODERATE | tolerated_low_confidence (0.06) | benign (0) | FE194 | 1 |
| HTRA2 | 606441 | chr2_74530221_T/C | ENSG00000115317 | ENST00000437202 | missense_variant | 176 | 176 | 59 | L/P | cTg/cCg | rs150047108,COSV51206225 | MODERATE | tolerated_low_confidence (0.06) | benign (0.003) | FE194 | 1 |
| HTRA2 | 606441 | chr2_74530221_T/C | ENSG00000115317 | ENST00000352222 | missense_variant | 232 | 215 | 72 | L/P | cTg/cCg | rs150047108,COSV51206225 | MODERATE | tolerated_low_confidence (0.07) | benign (0.003) | FE194 | 1 |
| IDH3A | 601149 | chr15_78168982_C/T | ENSG00000166411 | ENST00000299518 | missense_variant | 1120 | 1078 | 360 | R/C | Cgc/Tgc | rs116374996,COSV51908489 | MODERATE | deleterious (0.01) | benign (0.205) | FE190 | 1 |
| IDH3A | 601149 | chr15_78168982_C/T | ENSG00000166411 | ENST00000557960 | missense_variant,NMD_transcript_variant | 123 | 124 | 42 | R/C | Cgc/Tgc | rs116374996,COSV51908489 | MODERATE | deleterious (0.02) | possibly_damaging (0.54) | FE190 | 1 |
| IDH3A | 601149 | chr15_78168982_C/T | ENSG00000166411 | ENST00000558554 | missense_variant | 1017 | 973 | 325 | R/C | Cgc/Tgc | rs116374996,COSV51908489 | MODERATE | deleterious (0.01) | possibly_damaging (0.66) | FE190 | 1 |
| IDH3A | 601149 | chr15_78168982_C/T | ENSG00000166411 | ENST00000559205 | missense_variant | 265 | 241 | 81 | R/C | Cgc/Tgc | rs116374996,COSV51908489 | MODERATE | deleterious (0.01) | possibly_damaging (0.594) | FE190 | 1 |
| IGF2R | 147280 | chr6_160050497_A/G | ENSG00000197081 | ENST00000356956 | missense_variant | 2687 | 2539 | 847 | I/V | Atc/Gtc | rs939293106 | MODERATE | tolerated (0.46) | benign (0.145) | FE165 | 1 |
| IKZF2 | 606234 | chr2_213056961_T/C | ENSG00000030419 | ENST00000374319 | missense_variant | 588 | 278 | 93 | N/S | aAc/aGc | rs16849611 | MODERATE | tolerated (0.49) | benign (0.006) | FE106 | 1 |

| | | | | | | | | | | | | | | | | |
|---|---|---|---|---|---|---|---|---|---|---|---|---|---|---|---|---|
| IKZF2 | 606234 | chr2_213056961_T/C | ENSG00000030419 | ENST00000431520 | missense_variant,NMD_transcript_variant | 278 | 278 | 93 | N/S | aAc/aGc | rs16849611 | MODERATE | tolerated (0.61) | benign (0.003) | FE106 | 1 |
| IKZF2 | 606234 | chr2_213056961_T/C | ENSG00000030419 | ENST00000433134 | missense_variant | 521 | 296 | 99 | N/S | aAc/aGc | rs16849611 | MODERATE | tolerated (0.36) | benign (0.351) | FE106 | 1 |
| IKZF2 | 606234 | chr2_213056961_T/C | ENSG00000030419 | ENST00000342002 | missense_variant | 447 | 296 | 99 | N/S | aAc/aGc | rs16849611 | MODERATE | tolerated (0.69) | benign (0.001) | FE106 | 1 |
| IKZF2 | 606234 | chr2_213056961_T/C | ENSG00000030419 | ENST00000439848 | missense_variant,NMD_transcript_variant | 278 | 278 | 93 | N/S | aAc/aGc | rs16849611 | MODERATE | tolerated (0.38) | probably_damaging (0.979) | FE106 | 1 |
| IKZF2 | 606234 | chr2_213056961_T/C | ENSG00000030419 | ENST00000451136 | missense_variant | 594 | 149 | 50 | N/S | aAc/aGc | rs16849611 | MODERATE | tolerated (0.47) | benign (0.005) | FE106 | 1 |
| IKZF2 | 606234 | chr2_213056961_T/C | ENSG00000030419 | ENST00000457361 | missense_variant | 594 | 149 | 50 | N/S | aAc/aGc | rs16849611 | MODERATE | tolerated (0.36) | benign (0.011) | FE106 | 1 |
| IKZF2 | 606234 | chr2_213056961_T/C | ENSG00000030419 | ENST00000431520 | missense_variant,NMD_transcript_variant | 278 | 278 | 93 | N/S | aAc/aGc | rs16849611 | MODERATE | tolerated (0.61) | benign (0.003) | FE194 | 1 |
| IKZF2 | 606234 | chr2_213056961_T/C | ENSG00000030419 | ENST00000433134 | missense_variant | 521 | 296 | 99 | N/S | aAc/aGc | rs16849611 | MODERATE | tolerated (0.36) | benign (0.351) | FE194 | 1 |
| IKZF2 | 606234 | chr2_213056961_T/C | ENSG00000030419 | ENST00000342002 | missense_variant | 447 | 296 | 99 | N/S | aAc/aGc | rs16849611 | MODERATE | tolerated (0.69) | benign (0.001) | FE194 | 1 |
| IKZF2 | 606234 | chr2_213056961_T/C | ENSG00000030419 | ENST00000374319 | missense_variant | 588 | 278 | 93 | N/S | aAc/aGc | rs16849611 | MODERATE | tolerated (0.49) | benign (0.006) | FE194 | 1 |
| IKZF2 | 606234 | chr2_213056961_T/C | ENSG00000030419 | ENST00000451136 | missense_variant | 594 | 149 | 50 | N/S | aAc/aGc | rs16849611 | MODERATE | tolerated (0.47) | benign (0.005) | FE194 | 1 |
| IKZF2 | 606234 | chr2_213056961_T/C | ENSG00000030419 | ENST00000457361 | missense_variant | 594 | 149 | 50 | N/S | aAc/aGc | rs16849611 | MODERATE | tolerated (0.36) | benign (0.011) | FE194 | 1 |
| IKZF2 | 606234 | chr2_213056961_T/C | ENSG00000030419 | ENST00000434687 | missense_variant | 588 | 278 | 93 | N/S | aAc/aGc | rs16849611 | MODERATE | tolerated (0.61) | benign (0.001) | FE194 | 1 |
| IKZF2 | 606234 | chr2_213056961_T/C | ENSG00000030419 | ENST00000439848 | missense_variant,NMD_transcript_variant | 278 | 278 | 93 | N/S | aAc/aGc | rs16849611 | MODERATE | tolerated (0.38) | probably_damaging (0.979) | FE194 | 1 |
| IKZF2 | 606234 | chr2_213056961_T/C | ENSG00000030419 | ENST00000434687 | missense_variant | 588 | 278 | 93 | N/S | aAc/aGc | rs16849611 | MODERATE | tolerated (0.61) | benign (0.001) | FE106 | 1 |
| IKZF3 | 606221 | chr17_39788266_C/G | ENSG00000161405 | ENST00000346872 | missense_variant | 887 | 701 | 234 | G/A | gGg/gCg | rs112301322,COSV53099829 | MODERATE | tolerated (0.55) | benign (0.007) | FE136 | 1 |
| IL25 | 605658 | chr14_23375631_C/G | ENSG00000166090 | ENST00000329715 | missense_variant | 543 | 285 | 95 | D/E | gaC/gaG | - | MODERATE | deleterious (0) | possibly_damaging (0.906) | FE193 | 1 |
| IL25 | 605658 | chr14_23375631_C/G | ENSG00000166090 | ENST00000397242 | missense_variant | 395 | 237 | 79 | D/E | gaC/gaG | - | MODERATE | deleterious (0) | probably_damaging (0.994) | FE193 | 1 |
| IL3 | 147740 | chr5_132060983_A/G | ENSG00000164399 | ENST00000296870 | missense_variant | 231 | 179 | 60 | N/S | aAt/aGt | rs35482671 | MODERATE | tolerated (0.05) | benign (0.031) | FE165 | 1 |
| INPP5A | 600106 | chr10_132781888_C/T | ENSG00000068383 | ENST00000368594 | missense_variant | 1496 | 1186 | 396 | P/S | Ccc/Tcc | rs145146330 | MODERATE | tolerated (0.47) | benign (0.015) | FE154 | 1 |
| INPP5A | 600106 | chr10_132781888_C/T | ENSG00000068383 | ENST00000445580 | missense_variant | 232 | 232 | 78 | P/S | Ccc/Tcc | rs145146330 | MODERATE | tolerated (0.47) | benign (0.121) | FE154 | 1 |
| INTS13 | 615079 | chr12_26928865_C/T | ENSG00000064102 | ENST00000261191 | missense_variant | 538 | 341 | 114 | R/Q | cGg/cAg | rs771730672 | MODERATE | tolerated (0.48) | benign (0.003) | FE193 | 1 |
| INTS13 | 615079 | chr12_26928865_C/T | ENSG00000064102 | ENST00000537336 | missense_variant | 411 | 341 | 114 | R/Q | cGg/cAg | rs771730672 | MODERATE | tolerated (0.46) | benign (0.003) | FE193 | 1 |
| INTS13 | 615079 | chr12_26928865_C/T | ENSG00000064102 | ENST00000538727 | missense_variant | 218 | 38 | 13 | R/Q | cGg/cAg | rs771730672 | MODERATE | tolerated (0.39) | benign (0.003) | FE193 | 1 |
| INTS13 | 615079 | chr12_26928865_C/T | ENSG00000064102 | ENST00000544548 | missense_variant | 541 | 341 | 114 | R/Q | cGg/cAg | rs771730672 | MODERATE | tolerated (0.38) | benign (0.003) | FE193 | 1 |
| INTS4 | 611348 | chr11_77883877_G/A | ENSG00000149262 | ENST00000534064 | missense_variant | 2693 | 2668 | 890 | H/Y | Cac/Tac | rs749672057 | MODERATE | tolerated (0.4) | benign (0.201) | FE190 | 1 |
| INTS4 | 611348 | chr11_77883877_G/A | ENSG00000149262 | ENST00000535943 | missense_variant | 931 | 793 | 265 | H/Y | Cac/Tac | rs749672057 | MODERATE | tolerated (0.38) | benign (0.366) | FE190 | 1 |
| INTS5 | 611349 | chr11_62649230_C/T | ENSG00000185085 | ENST00000330574 | missense_variant | 903 | 850 | 284 | A/T | Gcg/Acg | - | MODERATE | tolerated (0.54) | benign (0.003) | FE193 | 1 |
| IP6K2 | 606992 | chr3_48688737_T/G | ENSG00000068745 | ENST00000328631 | missense_variant | 1012 | 817 | 273 | N/H | Aac/Cac | rs150229636 | MODERATE | deleterious (0) | probably_damaging (0.983) | FE106 | 1 |
| IPO11 | 610889 | chr5_62627199_A/G | ENSG00000086200 | ENST00000409296 | missense_variant | 3059 | 2929 | 977 | I/V | Atc/Gtc | rs11544795 | MODERATE | tolerated (1) | benign (0) | FE154 | 1 |
| IPO11 | 610889 | chr5_62627199_A/G | ENSG00000086200 | ENST00000409534 | missense_variant | 342 | 166 | 56 | I/V | Atc/Gtc | rs11544795 | MODERATE | tolerated (1) | benign (0) | FE154 | 1 |
| IPO11 | 610889 | chr5_62627199_A/G | ENSG00000086200 | ENST00000325324 | missense_variant | 2978 | 2809 | 937 | I/V | Atc/Gtc | rs11544795 | MODERATE | tolerated (1) | benign (0) | FE154 | 1 |
| ITGB8 | 604160 | chr7_20363679_G/T | ENSG00000105855 | ENST00000222573 | missense_variant | 1077 | 170 | 57 | R/M | aGg/aTg | rs376774573,COSV56006322 | MODERATE | tolerated (0.07) | possibly_damaging (0.446) | FE187 | 1 |
| ITGB8 | 604160 | chr7_20363679_G/T | ENSG00000105855 | ENST00000222573 | missense_variant | 1077 | 170 | 57 | R/M | aGg/aTg | rs376774573,COSV56006322 | MODERATE | tolerated (0.07) | possibly_damaging (0.446) | FE193 | 1 |
| JADE2 | 610515 | chr5_134566249_G/T | ENSG00000043143 | ENST00000395003 | missense_variant | 1282 | 1103 | 368 | G/V | gGc/gTc | rs115315395 | MODERATE | tolerated (0.06) | benign (0.065) | FE106 | 1 |
| KANSL3 | 617742 | chr2_96604271_A/C | ENSG00000114982 | ENST00000431828 | missense_variant | 2208 | 2128 | 710 | S/A | Tct/Gct | rs116798525 | MODERATE | tolerated_low_confidence (0.17) | benign (0.038) | FE164 | 1 |
| KANSL3 | 617742 | chr2_96604271_A/C | ENSG00000114982 | ENST00000420155 | missense_variant,NMD_transcript_variant | 2305 | 2206 | 736 | S/A | Tct/Gct | rs116798525 | MODERATE | tolerated_low_confidence (0.19) | benign (0) | FE164 | 1 |
| KANSL3 | 617742 | chr2_96604271_A/C | ENSG00000114982 | ENST00000670907 | missense_variant,NMD_transcript_variant | 2286 | 2206 | 736 | S/A | Tct/Gct | rs116798525 | MODERATE | tolerated_low_confidence (0.18) | benign (0.012) | FE164 | 1 |
| KANSL3 | 617742 | chr2_96604271_A/C | ENSG00000114982 | ENST00000666923 | missense_variant | 2290 | 2206 | 736 | S/A | Tct/Gct | rs116798525 | MODERATE | tolerated_low_confidence (0.19) | benign (0) | FE164 | 1 |
| KDM7A | NA | chr7_140091845_G/A | ENSG00000006459 | ENST00000397560 | missense_variant | 2736 | 2690 | 897 | P/L | cCg/cTg | rs192601953 | MODERATE | tolerated (0.07) | benign (0.018) | FE106 | 1 |
| KDM7A | NA | chr7_140091845_G/A | ENSG00000006459 | ENST00000397560 | missense_variant | 2736 | 2690 | 897 | P/L | cCg/cTg | rs192601953 | MODERATE | tolerated (0.07) | benign (0.018) | FE187 | 1 |
| KIF22 | 603213 | chr16_29804916_G/A | ENSG00000079616 | ENST00000561482 | missense_variant | 2213 | 1576 | 526 | D/N | Gat/Aat | rs146561986 | MODERATE | tolerated (0.25) | benign (0.021) | FE190 | 1 |

| | | | | | | | | | | | | | | | | |
|---|---|---|---|---|---|---|---|---|---|---|---|---|---|---|---|---|
| KIF22 | 603213 | chr16_29804916_G/A | ENSG00000079616 | ENST00000160827 | missense_variant | 1789 | 1780 | 594 | D/N | Gat/Aat | rs146561986 | MODERATE | tolerated (0.31) | benign (0.021) | FE190 | 1 |
| KIF22 | 603213 | chr16_29804916_G/A | ENSG00000079616 | ENST00000400751 | missense_variant | 2201 | 1576 | 526 | D/N | Gat/Aat | rs146561986 | MODERATE | tolerated (0.25) | benign (0.021) | FE190 | 1 |
| KIF22 | 603213 | chr16_29804916_G/A | ENSG00000079616 | ENST00000569382 | missense_variant | 1759 | 1618 | 540 | D/N | Gat/Aat | rs146561986 | MODERATE | tolerated (0.21) | benign (0.005) | FE190 | 1 |
| L1CAM | 308840 | chrX_153866778_C/T | ENSG00000198910 | ENST00000361699 | missense_variant | 2313 | 2302 | 768 | V/I | Gtc/Atc | rs36021462,CM950738, CM120261 | MODERATE | tolerated (0.21) | benign (0.227) | FE187 | 1 |
| L1CAM | 308840 | chrX_153866778_C/T | ENSG00000198910 | ENST00000361981 | missense_variant | 2395 | 2287 | 763 | V/I | Gtc/Atc | rs36021462,CM950738, CM120261 | MODERATE | tolerated (0.19) | benign (0.403) | FE187 | 1 |
| L1CAM | 308840 | chrX_153866778_C/T | ENSG00000198910 | ENST00000370055 | missense_variant | 2485 | 2287 | 763 | V/I | Gtc/Atc | rs36021462,CM950738, CM120261 | MODERATE | tolerated (0.19) | benign (0.403) | FE187 | 1 |
| L1CAM | 308840 | chrX_153866778_C/T | ENSG00000198910 | ENST00000370060 | missense_variant | 2519 | 2302 | 768 | V/I | Gtc/Atc | rs36021462,CM950738, CM120261 | MODERATE | tolerated (0.21) | possibly_damaging (0.539) | FE187 | 1 |
| L1CAM | 308840 | chrX_153866778_C/T | ENSG00000198910 | ENST00000455590 | missense_variant | 564 | 565 | 189 | V/I | Gtc/Atc | rs36021462,CM950738, CM120261 | MODERATE | deleterious (0.02) | benign (0.355) | FE187 | 1 |
| LCP2 | 601603 | chr5_170253135_G/C | ENSG00000043462 | ENST00000046794 | missense_variant | 1395 | 1229 | 410 | S/C | tCc/tGc | rs34192428 | MODERATE | tolerated (0.07) | possibly_damaging (0.784) | FE165 | 1 |
| LIAS | 607031 | chr4_39467554_T/A | ENSG00000121897 | ENST00000261434 | missense_variant | 367 | 336 | 112 | D/E | gaT/gaA | rs869312808,CM141141 | MODERATE | deleterious (0) | possibly_damaging (0.789) | FE165 | 1 |
| LIAS | 607031 | chr4_39467554_T/A | ENSG00000121897 | ENST00000340169 | missense_variant | 654 | 645 | 215 | D/E | gaT/gaA | rs869312808,CM141141 | MODERATE | deleterious (0) | probably_damaging (1) | FE165 | 1 |
| LIAS | 607031 | chr4_39467554_T/A | ENSG00000121897 | ENST00000513731 | missense_variant | 307 | 255 | 85 | D/E | gaT/gaA | rs869312808,CM141141 | MODERATE | deleterious (0) | probably_damaging (1) | FE165 | 1 |
| LIAS | 607031 | chr4_39467554_T/A | ENSG00000121897 | ENST00000638422 | missense_variant,NMD_transcript_variant | 718 | 645 | 215 | D/E | gaT/gaA | rs869312808,CM141141 | MODERATE | deleterious (0) | probably_damaging (0.999) | FE165 | 1 |
| LIAS | 607031 | chr4_39467554_T/A | ENSG00000121897 | ENST00000640349 | missense_variant | 535 | 531 | 177 | D/E | gaT/gaA | rs869312808,CM141141 | MODERATE | deleterious (0) | probably_damaging (1) | FE165 | 1 |
| LIAS | 607031 | chr4_39467554_T/A | ENSG00000121897 | ENST00000640888 | missense_variant | 707 | 645 | 215 | D/E | gaT/gaA | rs869312808,CM141141 | MODERATE | deleterious (0) | probably_damaging (1) | FE165 | 1 |
| LNPEP | 151300 | chr5_96979616_C/G | ENSG00000113441 | ENST00000231368 | missense_variant | 574 | 498 | 166 | I/M | atC/atG | rs61752351 | MODERATE | tolerated (0.69) | benign (0.017) | FE165 | 1 |
| LRP12 | 618299 | chr8_104490924_T/G | ENSG00000147650 | ENST00000276654 | missense_variant | 2438 | 2329 | 777 | I/L | Att/Ctt | rs115192762 | MODERATE | tolerated_low_confidence (0.18) | possibly_damaging (0.847) | FE154 | 1 |
| LRP1B | 608766 | chr2_141015881_T/C | ENSG00000168702 | ENST00000389484 | missense_variant | 2292 | 2005 | 669 | I/V | Ata/Gta | rs75995642,COSV67288977 | MODERATE | tolerated (0.53) | benign (0.001) | FE190 | 1 |
| LRP2 | 600073 | chr2_169185871_C/T | ENSG00000081479 | ENST00000649046 | missense_variant | 9612 | 9477 | 3159 | M/I | atG/atA | rs144322413 | MODERATE | deleterious (0.03) | benign (0.005) | FE106 | 1 |
| LRP2 | 600073 | chr2_169185871_C/T | ENSG00000081479 | ENST00000649153 | missense_variant,NMD_transcript_variant | 377 | 378 | 126 | M/I | atG/atA | rs144322413 | MODERATE | deleterious (0.03) | benign (0.062) | FE106 | 1 |
| LRP2 | 600073 | chr2_169226426_T/C | ENSG00000081479 | ENST00000649046 | missense_variant | 5525 | 5390 | 1797 | N/S | aAt/aGt | rs138070797 | MODERATE | tolerated (0.38) | benign (0.416) | FE164 | 1 |
| LRR1 | 609193 | chr14_49607523_A/G | ENSG00000165501 | ENST00000298288 | missense_variant | 487 | 406 | 136 | T/A | Act/Gct | rs34947556 | MODERATE | deleterious (0.04) | benign (0.012) | FE106 | 1 |
| LRR1 | 609193 | chr14_49607523_A/G | ENSG00000165501 | ENST00000298288 | missense_variant | 487 | 406 | 136 | T/A | Act/Gct | rs34947556 | MODERATE | deleterious (0.04) | benign (0.012) | FE154 | 1 |
| LRR1 | 609193 | chr14_49607784_A/G | ENSG00000165501 | ENST00000298288 | missense_variant | 748 | 667 | 223 | T/A | Aca/Gca | rs61754305 | MODERATE | tolerated (0.18) | benign (0.005) | FE164 | 1 |
| LRRTM1 | 610867 | chr2_80302937_T/C | ENSG00000162951 | ENST00000295057 | missense_variant | 1543 | 883 | 295 | I/V | Atc/Gtc | rs76300062 | MODERATE | tolerated (1) | benign (0.005) | FE164 | 1 |
| LRRTM1 | 610867 | chr2_80302937_T/C | ENSG00000162951 | ENST00000295057 | missense_variant | 1543 | 883 | 295 | I/V | Atc/Gtc | rs76300062 | MODERATE | tolerated (1) | benign (0.005) | FE130 | 1 |
| LSG1 | 610780 | chr3_194653007_T/C | ENSG00000041802 | ENST00000265245 | missense_variant | 924 | 895 | 299 | T/A | Aca/Gca | rs151257941 | MODERATE | tolerated (0.75) | benign (0.001) | FE193 | 1 |
| LSG1 | 610780 | chr3_194653007_T/C | ENSG00000041802 | ENST00000437613 | missense_variant | 96 | 97 | 33 | T/A | Aca/Gca | rs151257941 | MODERATE | tolerated_low_confidence (0.82) | benign (0.001) | FE193 | 1 |
| LYL1 | 151440 | chr19_13100877_T/C | ENSG00000104903 | ENST00000264824 | missense_variant | 647 | 295 | 99 | T/A | Act/Gct | - | MODERATE | tolerated (0.75) | benign (0) | FE154 | 1 |
| LYL1 | 151440 | chr19_13100877_T/C | ENSG00000104903 | ENST00000590974 | missense_variant | 803 | 73 | 25 | T/A | Act/Gct | - | MODERATE | tolerated_low_confidence (1) | benign (0) | FE154 | 1 |
| MAN1A2 | 604345 | chr1_117522919_G/A | ENSG00000198162 | ENST00000356554 | missense_variant | 2623 | 1888 | 630 | A/T | Gcc/Acc | rs41296190 | MODERATE | tolerated (0.56) | benign (0.005) | FE130 | 1 |
| MAP2 | 157130 | chr2_209695141_G/A | ENSG00000078018 | ENST00000360351 | missense_variant | 3477 | 2971 | 991 | G/R | Gga/Aga | rs35927101 | MODERATE | deleterious_low_confidence (0) | possibly_damaging (0.685) | FE130 | 1 |
| MAP2K4 | 601335 | chr17_12020983_G/A | ENSG00000065559 | ENST00000353533 | missense_variant | 107 | 97 | 33 | A/T | Gcc/Acc | rs1404280869 | MODERATE | tolerated (0.6) | benign (0) | FE136 | 1 |
| MAP3K12 | 600447 | chr12_53493697_A/T | ENSG00000139625 | ENST00000547035 | splice_donor_variant | - | - | - | - | - | rs117247527 | HIGH | - | - | FE164 | 1 |
| MARCHF6 | 613297 | chr5_10407181_C/T | ENSG00000145495 | ENST00000274140 | missense_variant | 1736 | 1532 | 511 | P/L | cCa/cTa | - | MODERATE | deleterious (0) | probably_damaging (0.999) | FE154 | 1 |
| MARK4 | 606495 | chr19_45294407_C/T | ENSG00000007047 | ENST00000262891 | missense_variant | 1871 | 1553 | 518 | P/L | cCg/cTg | rs144086640 | MODERATE | tolerated (0.07) | benign (0.028) | FE190 | 1 |
| MBD2 | 603547 | chr18_54224418_C/G | ENSG00000134046 | ENST00000256429 | missense_variant | 252 | 142 | 48 | G/R | Ggc/Cgc | rs534204537 | MODERATE | deleterious_low_confidence (0) | benign (0) | FE193 | 1 |
| MBD2 | 603547 | chr18_54224418_C/G | ENSG00000134046 | ENST00000398398 | missense_variant | 230 | 142 | 48 | G/R | Ggc/Cgc | rs534204537 | MODERATE | deleterious_low_confidence (0) | benign (0.003) | FE193 | 1 |
| MBD2 | 603547 | chr18_54224418_C/G | ENSG00000134046 | ENST00000583046 | missense_variant | 200 | 142 | 48 | G/R | Ggc/Cgc | rs534204537 | MODERATE | deleterious_low_confidence (0) | benign (0.045) | FE193 | 1 |
| MBD3 | 603573 | chr19_1578384_C/G | ENSG00000071655 | ENST00000156825 | missense_variant | 906 | 736 | 246 | E/Q | Gag/Cag | - | MODERATE | deleterious_low_confidence (0.02) | benign (0) | FE130 | 1 |
| MBD3 | 603573 | chr19_1578384_C/G | ENSG00000071655 | ENST00000434436 | missense_variant | 1066 | 832 | 278 | E/Q | Gag/Cag | - | MODERATE | deleterious_low_confidence (0.02) | benign (0) | FE130 | 1 |
| MBD3 | 603573 | chr19_1578384_C/G | ENSG00000071655 | ENST00000590550 | missense_variant | 1038 | 664 | 222 | E/Q | Gag/Cag | - | MODERATE | deleterious_low_confidence (0.02) | benign (0) | FE130 | 1 |

| | | | | | | | | | | | | | | | | |
|---|---|---|---|---|---|---|---|---|---|---|---|---|---|---|---|---|
| MED17 | 603810 | chr11_93812147_A/T | ENSG00000042429 | ENST00000639724 | missense_variant | 2086 | 1879 | 627 | I/L | Ata/Tta | rs116795572 | MODERATE | deleterious_low_confidence (0) | unknown (0) | FE154 | 1 |
| MED19 | 612385 | chr11_57712035_C/A | ENSG00000156603 | ENST00000337672 | missense_variant | 181 | 145 | 49 | A/S | Gcc/Tcc | rs199862897 | MODERATE | tolerated_low_confidence (0.73) | benign (0.015) | FE194 | 1 |
| MED19 | 612385 | chr11_57712035_C/A | ENSG00000156603 | ENST00000431606 | missense_variant | 289 | 145 | 49 | A/S | Gcc/Tcc | rs199862897 | MODERATE | tolerated (0.69) | benign (0.015) | FE194 | 1 |
| MED19 | 612385 | chr11_57712035_C/A | ENSG00000156603 | ENST00000645681 | missense_variant,NMD_transcript_variant | 180 | 145 | 49 | A/S | Gcc/Tcc | rs199862897 | MODERATE | tolerated_low_confidence (0.73) | benign (0.015) | FE194 | 1 |
| MED24 | 607000 | chr17_40019205_T/CACACAT/C | ENSG00000008838 | ENST00000614384 | inframe_insertion | 378 | 361 | 121 | M/MCV | ATGTGTGtg | - | MODERATE | - | - | FE165 | 1 |
| MED24 | 607000 | chr17_40019175_A/AACACACAC/C | ENSG00000008838 | ENST00000614384 | frameshift_variant | 408 | 391 | 131 | L/VCVX | GTGTGTGTTg | COSV56641415 | HIGH | - | - | FE193 | 1 |
| MED30 | 610237 | chr8_117521023_G/A | ENSG00000164758 | ENST00000522839 | missense_variant | 191 | 147 | 49 | M/I | atG/atA | - | MODERATE | tolerated (0.28) | benign (0.003) | FE190 | 1 |
| MED30 | 610237 | chr8_117521023_G/A | ENSG00000164758 | ENST00000297347 | missense_variant | 311 | 147 | 49 | M/I | atG/atA | - | MODERATE | deleterious (0.04) | benign (0.023) | FE190 | 1 |
| MEGF10 | 612453 | chr5_127339133_G/A | ENSG00000145794 | ENST00000274473 | missense_variant | 397 | 130 | 44 | V/M | Gtg/Atg | rs780558660 | MODERATE | deleterious (0.01) | probably_damaging (0.994) | FE187 | 1 |
| MEPCE | 611478 | chr7_100430034_G/A | ENSG00000146834 | ENST00000310512 | missense_variant | 204 | 16 | 6 | A/T | Gcg/Acg | - | MODERATE | deleterious_low_confidence (0.02) | benign (0.055) | FE106 | 1 |
| METTL17 | 616091 | chr14_20996699_G/A | ENSG00000165792 | ENST00000339374 | missense_variant | 1486 | 1253 | 418 | R/H | cGc/cAc | rs139452603 | MODERATE | tolerated (0.11) | benign (0.111) | FE165 | 1 |
| METTL17 | 616091 | chr14_20996699_G/A | ENSG00000165792 | ENST00000382985 | missense_variant | 1276 | 1253 | 418 | R/H | cGc/cAc | rs139452603 | MODERATE | tolerated (0.25) | benign (0.097) | FE165 | 1 |
| METTL17 | 616091 | chr14_20996699_G/A | ENSG00000165792 | ENST00000556670 | missense_variant | 1269 | 1253 | 418 | R/H | cGc/cAc | rs139452603 | MODERATE | tolerated (0.12) | benign (0.067) | FE165 | 1 |
| METTL17 | 616091 | chr14_20996699_G/A | ENSG00000165792 | ENST00000556733 | missense_variant | 279 | 281 | 94 | R/H | cGc/cAc | rs139452603 | MODERATE | tolerated (0.14) | benign (0.02) | FE165 | 1 |
| MGAT2 | 602616 | chr14_49621814_G/T | ENSG00000168282 | ENST00000305386 | missense_variant | 1016 | 546 | 182 | Q/H | caG/caT | rs780633686 | MODERATE | deleterious (0) | probably_damaging (0.999) | FE164 | 1 |
| MLLT3 | 159558 | chr9_20414275_T/C | ENSG00000171843 | ENST00000380338 | missense_variant | 814 | 571 | 191 | T/A | Acc/Gcc | rs1224384763 | MODERATE | tolerated (0.24) | benign (0) | FE190 | 1 |
| MMP14 | 600754 | chr14_22836827_G/A | ENSG00000157227 | ENST00000311852 | missense_variant | 243 | 10 | 4 | A/T | Gcc/Acc | rs17882219 | MODERATE | tolerated_low_confidence (0.21) | benign (0.012) | FE154 | 1 |
| MMP14 | 600754 | chr14_22836827_G/A | ENSG00000157227 | ENST00000547279 | missense_variant | 208 | 10 | 4 | A/T | Gcc/Acc | rs17882219 | MODERATE | - | benign (0.026) | FE154 | 1 |
| MOGS | 601336 | chr2_74461727_C/T | ENSG00000115275 | ENST00000452063 | missense_variant | 1885 | 1744 | 582 | A/T | Gct/Act | rs186098891 | MODERATE | tolerated (0.56) | possibly_damaging (0.75) | FE190 | 1 |
| MOGS | 601336 | chr2_74461727_C/T | ENSG00000115275 | ENST00000648810 | missense_variant | 1519 | 1237 | 413 | A/T | Gct/Act | rs186098891 | MODERATE | tolerated (1) | possibly_damaging (0.75) | FE190 | 1 |
| MOGS | 601336 | chr2_74461727_C/T | ENSG00000115275 | ENST00000448666 | missense_variant | 2197 | 2062 | 688 | A/T | Gct/Act | rs186098891 | MODERATE | tolerated (0.27) | possibly_damaging (0.75) | FE190 | 1 |
| MOGS | 601336 | chr2_74461727_C/T | ENSG00000115275 | ENST00000462443 | missense_variant | 1580 | 1237 | 413 | A/T | Gct/Act | rs186098891 | MODERATE | tolerated (1) | possibly_damaging (0.75) | FE190 | 1 |
| MOGS | 601336 | chr2_74461727_C/T | ENSG00000115275 | ENST00000649854 | missense_variant | 1695 | 1696 | 566 | A/T | Gct/Act | rs186098891 | MODERATE | tolerated (0.85) | possibly_damaging (0.841) | FE190 | 1 |
| MRPL18 | 611831 | chr6_159797489_C/T | ENSG00000112110 | ENST00000367034 | missense_variant | 563 | 442 | 148 | P/S | Cca/Tca | rs142227430 | MODERATE | tolerated (0.68) | benign (0.012) | FE154 | 1 |
| MRPL21 | 611834 | chr11_68892929_T/C | ENSG00000197345 | ENST00000362034 | missense_variant | 536 | 514 | 172 | M/V | Atg/Gtg | rs865881697 | MODERATE | deleterious (0.05) | benign (0.038) | FE106 | 1 |
| MRPL21 | 611834 | chr11_68892929_T/C | ENSG00000197345 | ENST00000450904 | missense_variant | 503 | 259 | 87 | M/V | Atg/Gtg | rs865881697 | MODERATE | deleterious (0.04) | benign (0.038) | FE106 | 1 |
| MRPL21 | 611834 | chr11_68892929_T/C | ENSG00000197345 | ENST00000567045 | missense_variant | 495 | 259 | 87 | M/V | Atg/Gtg | rs865881697 | MODERATE | tolerated (0.05) | probably_damaging (0.968) | FE106 | 1 |
| MRPL21 | 611834 | chr11_68898012_T/C | ENSG00000197345 | ENST00000544567 | n_variant,NMD_transcript_variant | 93 | 89 | 30 | D/G | gAt/gGt | rs117105758 | MODERATE | - | unknown (0) | FE165 | 1 |
| MRPL53 | 611857 | chr2_74472588_T/C | ENSG00000204822 | ENST00000409710 | missense_variant | 100 | 73 | 25 | K/E | Aaa/Gaa | rs141704877 | MODERATE | tolerated_low_confidence (0.07) | benign (0.034) | FE165 | 1 |
| MRPL53 | 611857 | chr2_74472588_T/C | ENSG00000204822 | ENST00000258105 | missense_variant | 93 | 73 | 25 | K/E | Aaa/Gaa | rs141704877 | MODERATE | tolerated (0.73) | benign (0.006) | FE165 | 1 |
| MRPS10 | 611976 | chr6_42214185_T/A | ENSG00000048544 | ENST00000053468 | missense_variant | 133 | 121 | 41 | N/Y | Aat/Tat | - | MODERATE | deleterious (0.01) | benign (0.054) | FE130 | 1 |
| MRPS28 | 611990 | chr8_80030106_C/G | ENSG00000147586 | ENST00000276585 | missense_variant | 154 | 143 | 48 | R/P | cGc/cCc | rs78308259 | MODERATE | tolerated (0.06) | benign (0.005) | FE106 | 1 |
| MRPS28 | 611990 | chr8_80030106_C/G | ENSG00000147586 | ENST00000518271 | missense_variant | 127 | 128 | 43 | R/P | cGc/cCc | rs78308259 | MODERATE | tolerated (0.06) | possibly_damaging (0.793) | FE106 | 1 |
| MRPS28 | 611990 | chr8_80030106_C/G | ENSG00000147586 | ENST00000521605 | missense_variant | 184 | 143 | 48 | R/P | cGc/cCc | rs78308259 | MODERATE | deleterious (0) | benign (0.012) | FE106 | 1 |
| MRPS34 | 611994 | chr16_1772442_C/G | ENSG00000074071 | ENST00000397375 | missense_variant | 451 | 436 | 146 | E/Q | Gag/Cag | rs139953295 | MODERATE | tolerated (0.07) | probably_damaging (0.948) | FE165 | 1 |
| MRPS34 | 611994 | chr16_1772442_C/G | ENSG00000074071 | ENST00000177742 | missense_variant | 488 | 457 | 153 | E/Q | Gag/Cag | rs139953295 | MODERATE | tolerated (0.07) | probably_damaging (0.948) | FE165 | 1 |
| MRPS5 | 611972 | chr2_95100877_A/G | ENSG00000144029 | ENST00000345084 | missense_variant,NMD_transcript_variant | 745 | 737 | 246 | F/S | tTc/tCc | rs113198616 | MODERATE | tolerated_low_confidence (0.63) | benign (0.006) | FE165 | 1 |
| MRPS5 | 611972 | chr2_95100854_C/T | ENSG00000144029 | ENST00000272418 | missense_variant | 868 | 851 | 284 | R/Q | cGa/cAa | rs115258802 | MODERATE | deleterious (0.03) | probably_damaging (0.939) | FE190 | 1 |
| MRTFB | 609463 | chr16_14240269_C/A | ENSG00000186260 | ENST00000318282 | missense_variant | 994 | 864 | 288 | H/Q | caC/caA | rs75963814 | MODERATE | tolerated (0.96) | probably_damaging (0.996) | FE136 | 1 |
| MRTFB | 609463 | chr16_14240269_C/A | ENSG00000186260 | ENST00000571589 | missense_variant | 1039 | 864 | 288 | H/Q | caC/caA | rs75963814 | MODERATE | tolerated (0.67) | possibly_damaging (0.469) | FE136 | 1 |
| MRTFB | 609463 | chr16_14240269_C/A | ENSG00000186260 | ENST00000572567 | missense_variant | 1029 | 831 | 277 | H/Q | caC/caA | rs75963814 | MODERATE | tolerated (0.86) | benign (0.059) | FE136 | 1 |
| MRTFB | 609463 | chr16_14240269_C/A | ENSG00000186260 | ENST00000573051 | missense_variant | 869 | 711 | 237 | H/Q | caC/caA | rs75963814 | MODERATE | tolerated (0.7) | possibly_damaging (0.578) | FE136 | 1 |

| | | | | | | | | | | | | | | | | |
|---|---|---|---|---|---|---|---|---|---|---|---|---|---|---|---|---|
| MRTFB | 609463 | chr16_14240269_C/A | ENSG00000186260 | ENST00000574045 | missense_variant | 1019 | 864 | 288 | H/Q | caC/caA | rs75963814 | MODERATE | tolerated (0.96) | probably_damaging (0.996) | FE136 | 1 |
| MRTFB | 609463 | chr16_14247091_C/A | ENSG00000186260 | ENST00000574045 | missense_variant | 1986 | 1831 | 611 | Q/K | Cag/Aag | rs1442945069 | MODERATE | tolerated (0.49) | benign (0.031) | FE194 | 1 |
| MRTFB | 609463 | chr16_14247091_C/A | ENSG00000186260 | ENST00000318282 | missense_variant | 1961 | 1831 | 611 | Q/K | Cag/Aag | rs1442945069 | MODERATE | tolerated (0.49) | benign (0.031) | FE194 | 1 |
| MRTFB | 609463 | chr16_14247091_C/A | ENSG00000186260 | ENST00000571589 | missense_variant | 2006 | 1831 | 611 | Q/K | Cag/Aag | rs1442945069 | MODERATE | tolerated (0.55) | benign (0.054) | FE194 | 1 |
| MTERF3 | 616930 | chr8_96257096_G/T | ENSG00000156469 | ENST00000287025 | missense_variant | 473 | 353 | 118 | P/Q | cCa/cAa | rs75477527 | MODERATE | tolerated (0.06) | possibly_damaging (0.867) | FE190 | 1 |
| MTERF3 | 616930 | chr8_96257096_G/T | ENSG00000156469 | ENST00000523821 | missense_variant | 473 | 353 | 118 | P/Q | cCa/cAa | rs75477527 | MODERATE | deleterious (0.05) | possibly_damaging (0.867) | FE190 | 1 |
| MTMR3 | 603558 | chr22_30019994_C/T | ENSG00000100330 | ENST00000401950 | missense_variant | 2658 | 2335 | 779 | L/F | Ctc/Ttc | rs61737780 | MODERATE | tolerated_low_confidence (0.15) | benign (0.109) | FE165 | 1 |
| MUS81 | 606591 | chr11_65863890_C/T | ENSG00000172732 | ENST00000308110 | missense_variant | 1369 | 1048 | 350 | R/W | Cgg/Tgg | rs34891773 | MODERATE | deleterious (0) | probably_damaging (0.93) | FE106 | 1 |
| MUS81 | 606591 | chr11_65863890_C/T | ENSG00000172732 | ENST00000524647 | missense_variant,NMD_transcript_variant | 942 | 943 | 315 | R/W | Cgg/Tgg | rs34891773 | MODERATE | deleterious (0) | probably_damaging (0.928) | FE106 | 1 |
| MUS81 | 606591 | chr11_65863890_C/T | ENSG00000172732 | ENST00000529374 | missense_variant | 824 | 826 | 276 | R/W | Cgg/Tgg | rs34891773 | MODERATE | deleterious (0) | probably_damaging (0.987) | FE106 | 1 |
| MUS81 | 606591 | chr11_65863890_C/T | ENSG00000172732 | ENST00000533035 | missense_variant | 1184 | 823 | 275 | R/W | Cgg/Tgg | rs34891773 | MODERATE | deleterious (0) | probably_damaging (0.93) | FE106 | 1 |
| MUS81 | 606591 | chr11_65863890_C/T | ENSG00000172732 | ENST00000529374 | missense_variant | 824 | 826 | 276 | R/W | Cgg/Tgg | rs34891773 | MODERATE | deleterious (0) | probably_damaging (0.987) | FE130 | 1 |
| MUS81 | 606591 | chr11_65863890_C/T | ENSG00000172732 | ENST00000533035 | missense_variant | 1184 | 823 | 275 | R/W | Cgg/Tgg | rs34891773 | MODERATE | deleterious (0) | probably_damaging (0.93) | FE130 | 1 |
| MUS81 | 606591 | chr11_65863890_C/T | ENSG00000172732 | ENST00000308110 | missense_variant | 1369 | 1048 | 350 | R/W | Cgg/Tgg | rs34891773 | MODERATE | deleterious (0) | probably_damaging (0.93) | FE130 | 1 |
| MUS81 | 606591 | chr11_65863890_C/T | ENSG00000172732 | ENST00000524647 | missense_variant,NMD_transcript_variant | 942 | 943 | 315 | R/W | Cgg/Tgg | rs34891773 | MODERATE | deleterious (0) | probably_damaging (0.928) | FE130 | 1 |
| MYBL1 | 159405 | chr8_66576116_C/T | ENSG00000185697 | ENST00000522677 | missense_variant | 1772 | 1361 | 454 | R/Q | cGa/cAa | rs760647023 | MODERATE | tolerated (0.15) | benign (0.226) | FE193 | 1 |
| NADK2 | 615787 | chr5_36219608_C/T | ENSG00000152620 | ENST00000282512 | missense_variant | 379 | 143 | 48 | R/H | cGt/cAt | rs138373837 | MODERATE | tolerated (0.1) | benign (0.013) | FE164 | 1 |
| NADK2 | 615787 | chr5_36219608_C/T | ENSG00000152620 | ENST00000381937 | missense_variant | 759 | 632 | 211 | R/H | cGt/cAt | rs138373837 | MODERATE | tolerated (0.06) | benign (0.013) | FE164 | 1 |
| NADK2 | 615787 | chr5_36219608_C/T | ENSG00000152620 | ENST00000397338 | missense_variant | 365 | 143 | 48 | R/H | cGt/cAt | rs138373837 | MODERATE | tolerated (0.1) | benign (0.013) | FE164 | 1 |
| NADK2 | 615787 | chr5_36219608_C/T | ENSG00000152620 | ENST00000511088 | missense_variant | 476 | 143 | 48 | R/H | cGt/cAt | rs138373837 | MODERATE | tolerated (0.12) | benign (0.013) | FE164 | 1 |
| NADK2 | 615787 | chr5_36219608_C/T | ENSG00000152620 | ENST00000514504 | missense_variant | 632 | 632 | 211 | R/H | cGt/cAt | rs138373837 | MODERATE | tolerated (0.05) | benign (0.006) | FE164 | 1 |
| NADK2 | 615787 | chr5_36219608_C/T | ENSG00000152620 | ENST00000617628 | missense_variant | 143 | 143 | 48 | R/H | cGt/cAt | rs138373837 | MODERATE | tolerated (0.09) | benign (0.006) | FE164 | 1 |
| NADK2 | 615787 | chr5_36219608_C/T | ENSG00000152620 | ENST00000506945 | missense_variant | 516 | 143 | 48 | R/H | cGt/cAt | rs138373837 | MODERATE | tolerated (0.07) | benign (0.014) | FE164 | 1 |
| NCAPD3 | 609276 | chr11_134152972_C/T | ENSG00000151503 | ENST00000534548 | missense_variant | 4510 | 4469 | 1490 | R/Q | cGa/cAa | rs147905924 | MODERATE | tolerated (0.05) | possibly_damaging (0.575) | FE154 | 1 |
| NCAPH2 | 611230 | chr22_50519304_C/T | ENSG00000025770 | ENST00000299821 | missense_variant | 923 | 845 | 282 | S/F | tCc/tTc | rs17850507 | MODERATE | deleterious (0.03) | possibly_damaging (0.542) | FE194 | 1 |
| NCAPH2 | 611230 | chr22_50519304_C/T | ENSG00000025770 | ENST00000395701 | missense_variant | 939 | 845 | 282 | S/F | tCc/tTc | rs17850507 | MODERATE | deleterious (0.03) | benign (0.123) | FE194 | 1 |
| NCAPH2 | 611230 | chr22_50519304_C/T | ENSG00000025770 | ENST00000523045 | missense_variant | 835 | 743 | 248 | S/F | tCc/tTc | rs17850507 | MODERATE | deleterious (0.03) | possibly_damaging (0.549) | FE194 | 1 |
| NCAPH2 | 611230 | chr22_50519304_C/T | ENSG00000025770 | ENST00000395698 | missense_variant | 940 | 845 | 282 | S/F | tCc/tTc | rs17850507 | MODERATE | deleterious (0) | benign (0.122) | FE194 | 1 |
| NCAPH2 | 611230 | chr22_50519304_C/T | ENSG00000025770 | ENST00000420993 | missense_variant | 959 | 845 | 282 | S/F | tCc/tTc | rs17850507 | MODERATE | deleterious (0.03) | possibly_damaging (0.671) | FE194 | 1 |
| NDUFB7 | 603842 | chr19_14566920_T/C | ENSG00000099795 | ENST00000593353 | missense_variant,NMD_transcript_variant | 186 | 124 | 42 | T/A | Aca/Gca | rs2228463 | MODERATE | deleterious_low_confidence (0.02) | benign (0) | FE136 | 1 |
| NELFA | 606026 | chr4_1986122_T/C | ENSG00000185049 | ENST00000382882 | missense_variant | 842 | 827 | 276 | K/R | aAg/aGg | rs150291014 | MODERATE | tolerated (0.36) | benign (0.015) | FE136 | 1 |
| NELFA | 606026 | chr4_1986122_T/C | ENSG00000185049 | ENST00000416258 | missense_variant | 839 | 839 | 280 | K/R | aAg/aGg | rs150291014 | MODERATE | tolerated (0.24) | benign (0.162) | FE136 | 1 |
| NELFA | 606026 | chr4_1986122_T/C | ENSG00000185049 | ENST00000431323 | missense_variant | 880 | 842 | 281 | K/R | aAg/aGg | rs150291014 | MODERATE | tolerated (0.15) | benign (0.274) | FE136 | 1 |
| NELFA | 606026 | chr4_1986122_T/C | ENSG00000185049 | ENST00000453740 | missense_variant | 529 | 530 | 177 | K/R | aAg/aGg | rs150291014 | MODERATE | tolerated (0.29) | benign (0.328) | FE136 | 1 |
| NELFA | 606026 | chr4_1986122_T/C | ENSG00000185049 | ENST00000455762 | missense_variant | 1015 | 617 | 206 | K/R | aAg/aGg | rs150291014 | MODERATE | tolerated (0.15) | benign (0.015) | FE136 | 1 |
| NELFA | 606026 | chr4_1986122_T/C | ENSG00000185049 | ENST00000542778 | missense_variant | 1103 | 860 | 287 | K/R | aAg/aGg | rs150291014 | MODERATE | tolerated (0.33) | benign (0.025) | FE136 | 1 |
| NELL2 | 602320 | chr12_44776143_C/T | ENSG00000184613 | ENST00000437801 | missense_variant | 1292 | 920 | 307 | R/Q | cGa/cAa | rs111769386 | MODERATE | tolerated (0.29) | benign (0.003) | FE130 | 1 |
| NFAT5 | 604708 | chr16_69693607_G/A | ENSG00000102908 | ENST00000354436 | missense_variant | 4046 | 3728 | 1243 | S/N | aGt/aAt | rs893101984 | MODERATE | tolerated_low_confidence (0.43) | benign (0) | FE154 | 1 |
| NFAT5 | 604708 | chr16_69693607_G/A | ENSG00000102908 | ENST00000349945 | missense_variant | 4118 | 3782 | 1261 | S/N | aGt/aAt | rs893101984 | MODERATE | tolerated_low_confidence (0.37) | benign (0) | FE154 | 1 |
| NFAT5 | 604708 | chr16_69693607_G/A | ENSG00000102908 | ENST00000567239 | missense_variant | 4019 | 3779 | 1260 | S/N | aGt/aAt | rs893101984 | MODERATE | tolerated_low_confidence (0.39) | benign (0) | FE154 | 1 |
| NFIB | 600728 | chr9_14307355_G/A | ENSG00000147862 | ENST00000380934 | missense_variant | 626 | 274 | 92 | P/S | Cct/Tct | rs140030018 | MODERATE | deleterious (0.02) | probably_damaging (0.994) | FE187 | 1 |
| NFIB | 600728 | chr9_14307355_G/A | ENSG00000147862 | ENST00000380953 | missense_variant | 826 | 196 | 66 | P/S | Cct/Tct | rs140030018 | MODERATE | deleterious (0.02) | possibly_damaging (0.605) | FE187 | 1 |

| | | | | | | | | | | | | | | | | |
|---|---|---|---|---|---|---|---|---|---|---|---|---|---|---|---|---|
| NFIB | 600728 | chr9_14307355_G/A | ENSG00000147862 | ENST00000380959 | missense_variant | 670 | 196 | 66 | P/S | Cct/Tct | rs140030018 | MODERATE | deleterious (0.02) | benign (0.054) | FE187 | 1 |
| NFIB | 600728 | chr9_14307355_G/A | ENSG00000147862 | ENST00000397579 | missense_variant | 1267 | 196 | 66 | P/S | Cct/Tct | rs140030018 | MODERATE | deleterious (0.02) | benign (0.054) | FE187 | 1 |
| NFIB | 600728 | chr9_14307355_G/A | ENSG00000147862 | ENST00000397581 | missense_variant | 1204 | 196 | 66 | P/S | Cct/Tct | rs140030018 | MODERATE | deleterious (0.01) | benign (0.054) | FE187 | 1 |
| NFIB | 600728 | chr9_14307355_G/A | ENSG00000147862 | ENST00000493697 | missense_variant | 272 | 181 | 61 | P/S | Cct/Tct | rs140030018 | MODERATE | tolerated (0.07) | benign (0.115) | FE187 | 1 |
| NFIB | 600728 | chr9_14307355_G/A | ENSG00000147862 | ENST00000397575 | missense_variant | 754 | 196 | 66 | P/S | Cct/Tct | rs140030018 | MODERATE | deleterious (0.01) | benign (0.054) | FE187 | 1 |
| NFIB | 600728 | chr9_14307355_G/A | ENSG00000147862 | ENST00000622520 | missense_variant | 648 | 196 | 66 | P/S | Cct/Tct | rs140030018 | MODERATE | tolerated (0.09) | benign (0.023) | FE187 | 1 |
| NFIB | 600728 | chr9_14307355_G/A | ENSG00000147862 | ENST00000635877 | missense_variant | 174 | 175 | 59 | P/S | Cct/Tct | rs140030018 | MODERATE | deleterious (0.02) | probably_damaging (0.986) | FE187 | 1 |
| NFIB | 600728 | chr9_14307355_G/A | ENSG00000147862 | ENST00000636057 | missense_variant | 320 | 169 | 57 | P/S | Cct/Tct | rs140030018 | MODERATE | deleterious (0.02) | possibly_damaging (0.615) | FE187 | 1 |
| NFIB | 600728 | chr9_14307355_G/A | ENSG00000147862 | ENST00000606230 | missense_variant | 564 | 184 | 62 | P/S | Cct/Tct | rs140030018 | MODERATE | deleterious (0.02) | probably_damaging (0.991) | FE187 | 1 |
| NFIB | 600728 | chr9_14307355_G/A | ENSG00000147862 | ENST00000380921 | missense_variant | 642 | 196 | 66 | P/S | Cct/Tct | rs140030018 | MODERATE | tolerated (0.07) | benign (0.103) | FE187 | 1 |
| NFIB | 600728 | chr9_14307355_G/A | ENSG00000147862 | ENST00000637640 | missense_variant | 175 | 169 | 57 | P/S | Cct/Tct | rs140030018 | MODERATE | deleterious (0.02) | probably_damaging (0.986) | FE187 | 1 |
| NFIB | 600728 | chr9_14307355_G/A | ENSG00000147862 | ENST00000637742 | missense_variant | 333 | 196 | 66 | P/S | Cct/Tct | rs140030018 | MODERATE | deleterious (0.02) | benign (0.054) | FE187 | 1 |
| NFIB | 600728 | chr9_14307355_G/A | ENSG00000147862 | ENST00000646622 | missense_variant | 286 | 184 | 62 | P/S | Cct/Tct | rs140030018 | MODERATE | deleterious (0.01) | probably_damaging (0.994) | FE187 | 1 |
| NFIB | 600728 | chr9_14307355_G/A | ENSG00000147862 | ENST00000636063 | missense_variant | 528 | 196 | 66 | P/S | Cct/Tct | rs140030018 | MODERATE | deleterious_low_confidence (0.02) | possibly_damaging (0.837) | FE187 | 1 |
| NFIB | 600728 | chr9_14307355_G/A | ENSG00000147862 | ENST00000636432 | missense_variant | 227 | 184 | 62 | P/S | Cct/Tct | rs140030018 | MODERATE | deleterious (0) | probably_damaging (0.913) | FE187 | 1 |
| NFYC | 605344 | chr1_40763472_G/T | ENSG00000066136 | ENST00000372669 | missense_variant | 896 | 830 | 277 | R/L | cGa/cTa | rs145040282 | MODERATE | tolerated_low_confidence (0.79) | unknown (0) | FE154 | 1 |
| NIFK | 611970 | chr2_121735687_C/T | ENSG00000155438 | ENST00000285814 | missense_variant | 194 | 169 | 57 | D/N | Gac/Aac | rs75350722 | MODERATE | tolerated (0.32) | benign (0) | FE106 | 1 |
| NOL4 | 603577 | chr18_33883412_G/A | ENSG00000101746 | ENST00000261592 | missense_variant | 1853 | 1555 | 519 | P/S | Cca/Tca | - | MODERATE | tolerated (0.49) | benign (0) | FE154 | 1 |
| NOM1 | 611269 | chr7_156949921_C/T | ENSG00000146909 | ENST00000275820 | missense_variant | 210 | 184 | 62 | P/S | Ccc/Tcc | rs144364133 | MODERATE | tolerated_low_confidence (0.08) | benign (0.001) | FE187 | 1 |
| NR5A2 | 604453 | chr1_200057561_C/A | ENSG00000116833 | ENST00000367357 | missense_variant | 930 | 931 | 311 | L/I | Ctc/Atc | rs71633889 | MODERATE | tolerated_low_confidence (0.16) | benign (0) | FE164 | 1 |
| NTRK3 | 191316 | chr15_87885716_T/A | ENSG00000140538 | ENST00000360948 | missense_variant | 2459 | 2153 | 718 | N/I | aAt/aTt | rs1460807501 | MODERATE | tolerated_low_confidence (0.18) | possibly_damaging (0.904) | FE193 | 1 |
| NTRK3 | 191316 | chr15_87885702_A/T | ENSG00000140538 | ENST00000360948 | missense_variant | 2473 | 2167 | 723 | W/R | Tgg/Agg | - | MODERATE | tolerated_low_confidence (0.33) | possibly_damaging (0.86) | FE193 | 1 |
| NVL | 602426 | chr1_224236546_A/T | ENSG00000143748 | ENST00000340871 | missense_variant | 2014 | 1759 | 587 | L/M | Ttg/Atg | COSV55874335 | MODERATE | deleterious (0.03) | possibly_damaging (0.828) | FE164 | 1 |
| NVL | 602426 | chr1_224236546_A/T | ENSG00000143748 | ENST00000391875 | missense_variant | 2295 | 2008 | 670 | L/M | Ttg/Atg | COSV55874335 | MODERATE | deleterious (0.02) | probably_damaging (0.989) | FE164 | 1 |
| NVL | 602426 | chr1_224236546_A/T | ENSG00000143748 | ENST00000281701 | missense_variant | 2371 | 2326 | 776 | L/M | Ttg/Atg | COSV55874335 | MODERATE | deleterious (0) | probably_damaging (0.989) | FE164 | 1 |
| NVL | 602426 | chr1_224236546_A/T | ENSG00000143748 | ENST00000482491 | missense_variant | 2469 | 1759 | 587 | L/M | Ttg/Atg | COSV55874335 | MODERATE | deleterious (0.03) | possibly_damaging (0.828) | FE164 | 1 |
| NVL | 602426 | chr1_224236546_A/T | ENSG00000143748 | ENST00000469075 | missense_variant | 2064 | 2053 | 685 | L/M | Ttg/Atg | COSV55874335 | MODERATE | deleterious (0) | possibly_damaging (0.828) | FE164 | 1 |
| NVL | 602426 | chr1_224236546_A/T | ENSG00000143748 | ENST00000469968 | missense_variant | 1973 | 1735 | 579 | L/M | Ttg/Atg | COSV55874335 | MODERATE | deleterious (0.03) | possibly_damaging (0.828) | FE164 | 1 |
| NYAP2 | 615478 | chr2_225582441_G/C | ENSG00000144460 | ENST00000272907 | missense_variant | 1199 | 1024 | 342 | A/P | Gcc/Ccc | rs761168029,COSV56001990 | MODERATE | deleterious (0.01) | probably_damaging (0.999) | FE193 | 1 |
| NYAP2 | 615478 | chr2_225582441_G/C | ENSG00000144460 | ENST00000272907 | missense_variant | 1199 | 1024 | 342 | A/P | Gcc/Ccc | rs761168029,COSV56001990 | MODERATE | deleterious (0.01) | probably_damaging (0.999) | FE136 | 1 |
| OBSL1 | 610991 | chr2_219552642_G/T | ENSG00000124006 | ENST00000373876 | missense_variant | 4935 | 4926 | 1642 | D/E | gaC/gaA | rs181520135 | MODERATE | tolerated (0.89) | benign (0.131) | FE190 | 1 |
| OBSL1 | 610991 | chr2_219552642_G/T | ENSG00000124006 | ENST00000404537 | missense_variant | 5509 | 5202 | 1734 | D/E | gaC/gaA | rs181520135 | MODERATE | tolerated (1) | benign (0.156) | FE190 | 1 |
| OPA1 | 605290 | chr3_193644073_C/A | ENSG00000198836 | ENST00000361908 | missense_variant | 1714 | 1522 | 508 | L/M | Ctg/Atg | COSV62480739 | MODERATE | tolerated (0.12) | benign (0.42) | FE190 | 1 |
| OPN1LW | 300822 | chrX_154154684_T/C | ENSG00000102076 | ENST00000369951 | missense_variant | 730 | 689 | 230 | I/T | aTc/aCc | rs148583295,CM045801,COSV64049257 | MODERATE | tolerated (0.15) | benign (0.005) | FE165 | 1 |
| ORC4 | 603056 | chr2_147958332_A/G | ENSG00000115947 | ENST00000264169 | missense_variant | 527 | 353 | 118 | L/S | tTa/tCa | rs61750441 | MODERATE | deleterious (0) | probably_damaging (1) | FE130 | 1 |
| ORC4 | 603056 | chr2_147958332_A/G | ENSG00000115947 | ENST00000457954 | missense_variant | 616 | 353 | 118 | L/S | tTa/tCa | rs61750441 | MODERATE | deleterious (0) | probably_damaging (1) | FE130 | 1 |
| ORC4 | 603056 | chr2_147958332_A/G | ENSG00000115947 | ENST00000392857 | missense_variant | 475 | 353 | 118 | L/S | tTa/tCa | rs61750441 | MODERATE | deleterious (0) | probably_damaging (1) | FE130 | 1 |
| ORC4 | 603056 | chr2_147958332_A/G | ENSG00000115947 | ENST00000440042 | missense_variant | 523 | 353 | 118 | L/S | tTa/tCa | rs61750441 | MODERATE | deleterious (0) | probably_damaging (1) | FE130 | 1 |
| ORC4 | 603056 | chr2_147958332_A/G | ENSG00000115947 | ENST00000535373 | missense_variant | 639 | 353 | 118 | L/S | tTa/tCa | rs61750441 | MODERATE | deleterious (0) | probably_damaging (1) | FE130 | 1 |
| ORC4 | 603056 | chr2_147958332_A/G | ENSG00000115947 | ENST00000536575 | missense_variant | 257 | 101 | 34 | L/S | tTa/tCa | rs61750441 | MODERATE | deleterious (0) | probably_damaging (1) | FE130 | 1 |
| ORC4 | 603056 | chr2_147958332_A/G | ENSG00000115947 | ENST00000540442 | missense_variant | 401 | 131 | 44 | L/S | tTa/tCa | rs61750441 | MODERATE | deleterious (0) | probably_damaging (1) | FE130 | 1 |
| ORC4 | 603056 | chr2_147958332_A/G | ENSG00000115947 | ENST00000416719 | missense_variant | 488 | 353 | 118 | L/S | tTa/tCa | rs61750441 | MODERATE | deleterious (0) | probably_damaging (1) | FE130 | 1 |

| | | | | | | | | | | | | | | | | |
|---|---|---|---|---|---|---|---|---|---|---|---|---|---|---|---|---|
| OXA1L | 601066 | chr14_22766613_G/C | ENSG00000155463 | ENST00000285848 | missense_variant | 92 | 92 | 31 | W/S | tGg/tCg | rs143108324 | MODERATE | tolerated_low_confidence (0.2) | possibly_damaging (0.494) | FE106 | 1 |
| PAFAH1B1 | 601545 | chr17_2674283_T/A | ENSG00000007168 | ENST00000574468 | missense_variant | 391 | 391 | 131 | S/T | Tct/Act | - | MODERATE | tolerated (0.15) | benign (0.003) | FE130 | 1 |
| PAFAH1B1 | 601545 | chr17_2674283_T/A | ENSG00000007168 | ENST00000397195 | missense_variant | 1438 | 895 | 299 | S/T | Tct/Act | - | MODERATE | tolerated (0.59) | benign (0) | FE130 | 1 |
| PAK3 | 300142 | chrX_111123170_C/T | ENSG00000077264 | ENST00000360648 | missense_variant | 94 | 67 | 23 | R/W | Cgg/Tgg | rs978487268,COSV53276222,COSV53272582 | MODERATE | deleterious_low_confidence (0) | possibly_damaging (0.761) | FE187 | 2 |
| PAX8 | 167415 | chr2_113227201_G/A | ENSG00000125618 | ENST00000263335 | missense_variant | 989 | 833 | 278 | A/V | gCg/gTg | rs145036350 | MODERATE | tolerated_low_confidence (0.38) | benign (0) | FE154 | 1 |
| PAX8 | 167415 | chr2_113227201_G/A | ENSG00000125618 | ENST00000348715 | missense_variant | 1220 | 1064 | 355 | A/V | gCg/gTg | rs145036350 | MODERATE | tolerated_low_confidence (0.91) | benign (0) | FE154 | 1 |
| PCCB | 232050 | chr3_136298060_G/A | ENSG00000114054 | ENST00000462637 | missense_variant | 824 | 803 | 268 | C/Y | tGc/tAc | rs77820367,CM1110010 | MODERATE | tolerated_low_confidence (0.11) | possibly_damaging (0.722) | FE165 | 1 |
| PCCB | 232050 | chr3_136298060_G/A | ENSG00000114054 | ENST00000466072 | missense_variant | 893 | 872 | 291 | C/Y | tGc/tAc | rs77820367,CM1110010 | MODERATE | tolerated_low_confidence (0.1) | possibly_damaging (0.863) | FE165 | 1 |
| PCCB | 232050 | chr3_136298060_G/A | ENSG00000114054 | ENST00000484181 | missense_variant,NMD_transcript_variant | 893 | 872 | 291 | C/Y | tGc/tAc | rs77820367,CM1110010 | MODERATE | tolerated_low_confidence (0.09) | possibly_damaging (0.694) | FE165 | 1 |
| PCCB | 232050 | chr3_136298060_G/A | ENSG00000114054 | ENST00000471595 | missense_variant | 890 | 872 | 291 | C/Y | tGc/tAc | rs77820367,CM1110010 | MODERATE | tolerated_low_confidence (0.14) | possibly_damaging (0.722) | FE165 | 1 |
| PCCB | 232050 | chr3_136298060_G/A | ENSG00000114054 | ENST00000251654 | missense_variant | 908 | 872 | 291 | C/Y | tGc/tAc | rs77820367,CM1110010 | MODERATE | tolerated_low_confidence (0.13) | possibly_damaging (0.722) | FE165 | 1 |
| PCCB | 232050 | chr3_136259204_C/G | ENSG00000114054 | ENST00000469217 | missense_variant | 418 | 401 | 134 | P/R | cCg/cGg | rs756629698 | MODERATE | deleterious_low_confidence (0.02) | benign (0) | FE106 | 1 |
| PCCB | 232050 | chr3_136298060_G/A | ENSG00000114054 | ENST00000469217 | missense_variant | 949 | 932 | 311 | C/Y | tGc/tAc | rs77820367,CM1110010 | MODERATE | tolerated_low_confidence (0.11) | possibly_damaging (0.722) | FE165 | 1 |
| PCCB | 232050 | chr3_136298060_G/A | ENSG00000114054 | ENST00000468777 | missense_variant | 986 | 965 | 322 | C/Y | tGc/tAc | rs77820367,CM1110010 | MODERATE | tolerated_low_confidence (0.11) | possibly_damaging (0.722) | FE165 | 1 |
| PCCB | 232050 | chr3_136298060_G/A | ENSG00000114054 | ENST00000483687 | missense_variant | 836 | 815 | 272 | C/Y | tGc/tAc | rs77820367,CM1110010 | MODERATE | tolerated_low_confidence (0.11) | possibly_damaging (0.722) | FE165 | 1 |
| PCCB | 232050 | chr3_136298060_G/A | ENSG00000114054 | ENST00000478469 | missense_variant | 873 | 872 | 291 | C/Y | tGc/tAc | rs77820367,CM1110010 | MODERATE | deleterious_low_confidence (0.05) | possibly_damaging (0.793) | FE165 | 1 |
| PCCB | 232050 | chr3_136298060_G/A | ENSG00000114054 | ENST00000482086 | missense_variant | 545 | 524 | 175 | C/Y | tGc/tAc | rs77820367,CM1110010 | MODERATE | tolerated_low_confidence (0.14) | possibly_damaging (0.722) | FE165 | 1 |
| PCCB | 232050 | chr3_136298060_G/A | ENSG00000114054 | ENST00000490504 | missense_variant | 722 | 701 | 234 | C/Y | tGc/tAc | rs77820367,CM1110010 | MODERATE | tolerated (0.16) | benign (0.024) | FE165 | 1 |
| PCDH11X | 300246 | chrX_92263139_G/A | ENSG00000102290 | ENST00000373094 | missense_variant | 3985 | 3140 | 1047 | G/E | gGa/gAa | rs147493192,COSV99036178 | MODERATE | tolerated (0.1) | benign (0.009) | FE136 | 1 |
| PCID2 | 613713 | chr13_113180195_G/T | ENSG00000126226 | ENST00000246505 | missense_variant | 1020 | 985 | 329 | H/N | Cac/Aac | rs546330602 | MODERATE | tolerated (0.8) | benign (0.001) | FE193 | 1 |
| PCID2 | 613713 | chr13_113180195_G/T | ENSG00000126226 | ENST00000337344 | missense_variant | 858 | 823 | 275 | H/N | Cac/Aac | rs546330602 | MODERATE | tolerated (0.74) | benign (0) | FE193 | 1 |
| PCID2 | 613713 | chr13_113180195_G/T | ENSG00000126226 | ENST00000375457 | missense_variant | 1414 | 817 | 273 | H/N | Cac/Aac | rs546330602 | MODERATE | tolerated (0.73) | benign (0.031) | FE193 | 1 |
| PCID2 | 613713 | chr13_113180195_G/T | ENSG00000126226 | ENST00000375459 | missense_variant | 870 | 817 | 273 | H/N | Cac/Aac | rs546330602 | MODERATE | tolerated (0.73) | benign (0.031) | FE193 | 1 |
| PCID2 | 613713 | chr13_113180195_G/T | ENSG00000126226 | ENST00000375477 | missense_variant | 855 | 823 | 275 | H/N | Cac/Aac | rs546330602 | MODERATE | tolerated (0.74) | benign (0) | FE193 | 1 |
| PCID2 | 613713 | chr13_113180195_G/T | ENSG00000126226 | ENST00000375479 | missense_variant | 904 | 823 | 275 | H/N | Cac/Aac | rs546330602 | MODERATE | tolerated (0.74) | benign (0) | FE193 | 1 |
| PCID2 | 613713 | chr13_113180195_G/T | ENSG00000126226 | ENST00000622406 | missense_variant | 1066 | 985 | 329 | H/N | Cac/Aac | rs546330602 | MODERATE | tolerated (0.8) | benign (0.001) | FE193 | 1 |
| PCYT2 | 602679 | chr17_81909549_A/G | ENSG00000185813 | ENST00000570391 | missense_variant | 421 | 47 | 16 | M/T | aTg/aCg | rs150714189 | MODERATE | deleterious (0) | benign (0.034) | FE165 | 1 |
| PCYT2 | 602679 | chr17_81909549_A/G | ENSG00000185813 | ENST00000576343 | missense_variant | 206 | 143 | 48 | M/T | aTg/aCg | rs150714189 | MODERATE | deleterious (0) | probably_damaging (0.936) | FE165 | 1 |
| PCYT2 | 602679 | chr17_81909549_A/G | ENSG00000185813 | ENST00000538721 | missense_variant | 194 | 143 | 48 | M/T | aTg/aCg | rs150714189 | MODERATE | deleterious (0) | benign (0.033) | FE194 | 1 |
| PCYT2 | 602679 | chr17_81909549_A/G | ENSG00000185813 | ENST00000572157 | missense_variant | 329 | 47 | 16 | M/T | aTg/aCg | rs150714189 | MODERATE | deleterious_low_confidence (0) | benign (0.007) | FE165 | 1 |
| PCYT2 | 602679 | chr17_81909549_A/G | ENSG00000185813 | ENST00000573636 | missense_variant | 198 | 143 | 48 | M/T | aTg/aCg | rs150714189 | MODERATE | deleterious (0) | possibly_damaging (0.6) | FE165 | 1 |
| PCYT2 | 602679 | chr17_81909549_A/G | ENSG00000185813 | ENST00000538936 | missense_variant | 187 | 143 | 48 | M/T | aTg/aCg | rs150714189 | MODERATE | deleterious (0) | benign (0.034) | FE165 | 1 |
| PCYT2 | 602679 | chr17_81909549_A/G | ENSG00000185813 | ENST00000571105 | missense_variant | 201 | 143 | 48 | M/T | aTg/aCg | rs150714189 | MODERATE | deleterious (0) | possibly_damaging (0.624) | FE165 | 1 |
| PCYT2 | 602679 | chr17_81909549_A/G | ENSG00000185813 | ENST00000576343 | missense_variant | 206 | 143 | 48 | M/T | aTg/aCg | rs150714189 | MODERATE | deleterious (0) | probably_damaging (0.936) | FE194 | 1 |
| PCYT2 | 602679 | chr17_81909549_A/G | ENSG00000185813 | ENST00000538721 | missense_variant | 194 | 143 | 48 | M/T | aTg/aCg | rs150714189 | MODERATE | deleterious (0) | benign (0.033) | FE165 | 1 |
| PCYT2 | 602679 | chr17_81909549_A/G | ENSG00000185813 | ENST00000571105 | missense_variant | 201 | 143 | 48 | M/T | aTg/aCg | rs150714189 | MODERATE | deleterious (0) | possibly_damaging (0.624) | FE194 | 1 |
| PCYT2 | 602679 | chr17_81909549_A/G | ENSG00000185813 | ENST00000573636 | missense_variant | 198 | 143 | 48 | M/T | aTg/aCg | rs150714189 | MODERATE | deleterious (0) | possibly_damaging (0.6) | FE194 | 1 |
| PCYT2 | 602679 | chr17_81909549_A/G | ENSG00000185813 | ENST00000538936 | missense_variant | 187 | 143 | 48 | M/T | aTg/aCg | rs150714189 | MODERATE | deleterious (0) | benign (0.034) | FE194 | 1 |
| PCYT2 | 602679 | chr17_81909549_A/G | ENSG00000185813 | ENST00000570391 | missense_variant | 421 | 47 | 16 | M/T | aTg/aCg | rs150714189 | MODERATE | deleterious (0) | benign (0.034) | FE194 | 1 |
| PCYT2 | 602679 | chr17_81909549_A/G | ENSG00000185813 | ENST00000572157 | missense_variant | 329 | 47 | 16 | M/T | aTg/aCg | rs150714189 | MODERATE | deleterious_low_confidence (0) | benign (0.007) | FE194 | 1 |
| PEX12 | 601758 | chr17_35577920_T/A | ENSG00000108733 | ENST00000586663 | missense_variant | 633 | 102 | 34 | R/S | agA/agT | rs147530802,CM073260 | MODERATE | deleterious (0.01) | possibly_damaging (0.597) | FE130 | 1 |
| PEX12 | 601758 | chr17_35577920_T/A | ENSG00000108733 | ENST00000613219 | missense_variant | 718 | 102 | 34 | R/S | agA/agT | rs147530802,CM073260 | MODERATE | deleterious (0.01) | possibly_damaging (0.597) | FE130 | 1 |

| | | | | | | | | | | | | | | | |
|---|---|---|---|---|---|---|---|---|---|---|---|---|---|---|---|
| PEX12 | 601758 | chr17_35577920_T/A | ENSG00000108733 | ENST00000225873 | missense_variant | 652 | 102 | 34 | R/S | agA/agT | rs147530802,CM073260 | MODERATE | deleterious (0.01) | possibly_damaging (0.597) | FE130 | 1 |
| PEX12 | 601758 | chr17_35577920_T/A | ENSG00000108733 | ENST00000585380 | missense_variant | 336 | 102 | 34 | R/S | agA/agT | rs147530802,CM073260 | MODERATE | deleterious (0.01) | possibly_damaging (0.597) | FE130 | 1 |
| PEX26 | 608666 | chr22_18083474_G/C | ENSG00000215193 | ENST00000329627 | missense_variant | 615 | 409 | 137 | V/L | Gtg/Ctg | rs142648687 | MODERATE | tolerated (0.06) | benign (0.031) | FE164 | 1 |
| PEX26 | 608666 | chr22_18083474_G/C | ENSG00000215193 | ENST00000399744 | missense_variant | 796 | 409 | 137 | V/L | Gtg/Ctg | rs142648687 | MODERATE | tolerated (0.06) | benign (0.031) | FE164 | 1 |
| PEX26 | 608666 | chr22_18083474_G/C | ENSG00000215193 | ENST00000428061 | missense_variant | 409 | 409 | 137 | V/L | Gtg/Ctg | rs142648687 | MODERATE | tolerated (0.06) | benign (0.018) | FE164 | 1 |
| PEX26 | 608666 | chr22_18083474_G/C | ENSG00000215193 | ENST00000610387 | missense_variant | 618 | 409 | 137 | V/L | Gtg/Ctg | rs142648687 | MODERATE | tolerated (0.06) | benign (0.018) | FE164 | 1 |
| PEX26 | 608666 | chr22_18079968_T/C | ENSG00000215193 | ENST00000610387 | missense_variant | 534 | 325 | 109 | Y/H | Tat/Cat | rs45567240 | MODERATE | tolerated (0.12) | probably_damaging (0.991) | FE194 | 1 |
| PEX26 | 608666 | chr22_18079968_T/C | ENSG00000215193 | ENST00000329627 | missense_variant | 531 | 325 | 109 | Y/H | Tat/Cat | rs45567240 | MODERATE | tolerated (0.07) | probably_damaging (1) | FE194 | 1 |
| PEX26 | 608666 | chr22_18079968_T/C | ENSG00000215193 | ENST00000399744 | missense_variant | 712 | 325 | 109 | Y/H | Tat/Cat | rs45567240 | MODERATE | tolerated (0.07) | probably_damaging (1) | FE194 | 1 |
| PEX26 | 608666 | chr22_18079968_T/C | ENSG00000215193 | ENST00000428061 | missense_variant | 325 | 325 | 109 | Y/H | Tat/Cat | rs45567240 | MODERATE | tolerated (0.12) | probably_damaging (0.991) | FE194 | 1 |
| PEX26 | 608666 | chr22_18079968_T/C | ENSG00000215193 | ENST00000474897 | missense_variant,NMD_transcript_variant | 687 | 325 | 109 | Y/H | Tat/Cat | rs45567240 | MODERATE | tolerated_low_confidence (0.09) | benign (0.104) | FE194 | 1 |
| PEX3 | 603164 | chr6_143471401_G/T | ENSG00000034693 | ENST00000367592 | missense_variant | 579 | 343 | 115 | D/Y | Gat/Tat | - | MODERATE | deleterious (0.01) | benign (0.021) | FE190 | 1 |
| PEX3 | 603164 | chr6_143471401_G/T | ENSG00000034693 | ENST00000367591 | missense_variant | 713 | 475 | 159 | D/Y | Gat/Tat | - | MODERATE | deleterious (0.01) | benign (0.031) | FE190 | 1 |
| PEX5 | 600414 | chr12_7189938_T/C | ENSG00000139197 | ENST00000412720 | missense_variant | 94 | 5 | 2 | L/P | cTc/cCc | rs186539500 | MODERATE | - | benign (0) | FE154 | 1 |
| PGM3 | 172100 | chr6_83166409_G/T | ENSG00000013375 | ENST00000512866 | missense_variant | 1734 | 1679 | 560 | A/D | gCt/gAt | - | MODERATE | - | unknown (0) | FE154 | 1 |
| PGM3 | 172100 | chr6_83191240_A/G | ENSG00000013375 | ENST00000508748 | missense_variant | 199 | 29 | 10 | I/T | aTc/aCc | rs373825865 | MODERATE | deleterious_low_confidence (0) | benign (0.003) | FE154 | 1 |
| PGM3 | 172100 | chr6_83191240_A/G | ENSG00000013375 | ENST00000506587 | missense_variant | 176 | 29 | 10 | I/T | aTc/aCc | rs373825865 | MODERATE | deleterious_low_confidence (0.01) | benign (0.003) | FE154 | 1 |
| PGM3 | 172100 | chr6_83191240_A/G | ENSG00000013375 | ENST00000503094 | missense_variant | 245 | 29 | 10 | I/T | aTc/aCc | rs373825865 | MODERATE | deleterious_low_confidence (0) | benign (0.003) | FE154 | 1 |
| PHC1 | 602978 | chr12_8933192_C/G | ENSG00000111752 | ENST00000433083 | missense_variant | 1745 | 1600 | 534 | P/A | Cct/Gct | rs201210657 | MODERATE | tolerated (0.36) | benign (0.01) | FE164 | 1 |
| PHC1 | 602978 | chr12_8933192_C/G | ENSG00000111752 | ENST00000543824 | missense_variant | 2067 | 1735 | 579 | P/A | Cct/Gct | rs201210657 | MODERATE | tolerated (0.31) | benign (0.003) | FE164 | 1 |
| PHC1 | 602978 | chr12_8933192_C/G | ENSG00000111752 | ENST00000544916 | missense_variant | 2102 | 1735 | 579 | P/A | Cct/Gct | rs201210657 | MODERATE | tolerated (0.31) | benign (0.003) | FE164 | 1 |
| PHF12 | 618645 | chr17_28911117_T/C | ENSG00000109118 | ENST00000332830 | missense_variant | 2768 | 2210 | 737 | N/S | aAt/aGt | rs148347485 | MODERATE | tolerated (0.51) | benign (0.001) | FE130 | 1 |
| PHF21A | 608325 | chr11_45935703_C/T | ENSG00000135365 | ENST00000323180 | missense_variant | 1948 | 1580 | 527 | S/N | aGt/aAt | rs751045065 | MODERATE | tolerated (0.15) | probably_damaging (0.995) | FE193 | 1 |
| PHF21A | 608325 | chr11_45935703_C/T | ENSG00000135365 | ENST00000418153 | missense_variant | 1918 | 1718 | 573 | S/N | aGt/aAt | rs751045065 | MODERATE | tolerated (0.16) | possibly_damaging (0.554) | FE193 | 1 |
| PHF21A | 608325 | chr11_45935703_C/T | ENSG00000135365 | ENST00000525676 | missense_variant | 51 | 53 | 18 | S/N | aGt/aAt | rs751045065 | MODERATE | deleterious_low_confidence (0) | probably_damaging (0.969) | FE193 | 1 |
| PHF21A | 608325 | chr11_45935703_C/T | ENSG00000135365 | ENST00000530587 | missense_variant,NMD_transcript_variant | 221 | 221 | 74 | S/N | aGt/aAt | rs751045065 | MODERATE | tolerated (0.26) | probably_damaging (0.954) | FE193 | 1 |
| PHF21A | 608325 | chr11_45935703_C/T | ENSG00000135365 | ENST00000532028 | missense_variant | 143 | 143 | 48 | S/N | aGt/aAt | rs751045065 | MODERATE | tolerated (0.18) | probably_damaging (0.953) | FE193 | 1 |
| PHF21B | 616727 | chr22_44883123_G/A | ENSG00000056487 | ENST00000313237 | missense_variant | 2015 | 1559 | 520 | T/I | aCa/aTa | rs78473374 | MODERATE | deleterious_low_confidence (0.04) | benign (0.001) | FE194 | 1 |
| PI4K2A | 609763 | chr10_97656887_G/A | ENSG00000155252 | ENST00000370631 | missense_variant | 892 | 835 | 279 | A/T | Gca/Aca | rs146387356 | MODERATE | tolerated (0.26) | benign (0.065) | FE164 | 1 |
| PI4K2A | 609763 | chr10_97662967_G/A | ENSG00000155252 | ENST00000370631 | missense_variant,splice_region_variant | 1040 | 983 | 328 | R/Q | cGg/cAg | rs61760983 | MODERATE | tolerated (0.25) | benign (0.009) | FE187 | 1 |
| PI4KB | 602758 | chr1_151293457_G/A | ENSG00000143393 | ENST00000455060 | missense_variant | 445 | 445 | 149 | R/C | Cgc/Tgc | rs148244140 | MODERATE | tolerated (0.11) | unknown (0) | FE106 | 1 |
| PI4KB | 602758 | chr1_151293457_G/A | ENSG00000143393 | ENST00000455060 | missense_variant | 445 | 445 | 149 | R/C | Cgc/Tgc | rs148244140 | MODERATE | tolerated (0.11) | unknown (0) | FE154 | 1 |
| PIK3CD | 602839 | chr1_9717541_C/G | ENSG00000171608 | ENST00000377346 | missense_variant | 1144 | 935 | 312 | S/C | tCc/tGc | rs61755420 | MODERATE | deleterious (0.05) | benign (0.353) | FE194 | 1 |
| PIM3 | 610580 | chr22_49961135_C/G | ENSG00000198355 | ENST00000360612 | missense_variant | 272 | 96 | 32 | D/E | gaC/gaG | rs201604409 | MODERATE | tolerated (1) | benign (0.003) | FE154 | 1 |
| PKN2 | 602549 | chr1_88805742_G/A | ENSG00000065243 | ENST00000316005 | missense_variant | 2106 | 1747 | 583 | G/S | Ggt/Agt | - | MODERATE | tolerated_low_confidence (0.37) | benign (0) | FE194 | 1 |
| PLAA | 603873 | chr9_26917138_G/A | ENSG00000137055 | ENST00000517642 | missense_variant | 462 | 464 | 155 | S/L | tCg/tTg | rs140970730 | MODERATE | tolerated (0.06) | probably_damaging (0.996) | FE194 | 1 |
| PLAA | 603873 | chr9_26917138_G/A | ENSG00000137055 | ENST00000520884 | missense_variant | 1633 | 1445 | 482 | S/L | tCg/tTg | rs140970730 | MODERATE | tolerated (0.08) | benign (0.041) | FE194 | 1 |
| PLAA | 603873 | chr9_26917138_G/A | ENSG00000137055 | ENST00000397292 | missense_variant | 1642 | 1445 | 482 | S/L | tCg/tTg | rs140970730 | MODERATE | tolerated (0.05) | benign (0.035) | FE194 | 1 |
| PLEC | 601282 | chr8_143916616_G/A | ENSG00000178209 | ENST00000398774 | missense_variant | 13183 | 13109 | 4370 | T/M | aCg/aTg | rs113513807 | MODERATE | tolerated (0.24) | benign (0.003) | FE164 | 1 |
| PLEC | 601282 | chr8_143916616_G/A | ENSG00000178209 | ENST00000436759 | missense_variant | 13324 | 13286 | 4429 | T/M | aCg/aTg | rs113513807 | MODERATE | tolerated (0.26) | benign (0.003) | FE164 | 1 |
| PLEC | 601282 | chr8_143916616_G/A | ENSG00000178209 | ENST00000527096 | missense_variant | 13274 | 13274 | 4425 | T/M | aCg/aTg | rs113513807 | MODERATE | tolerated (0.26) | benign (0.003) | FE164 | 1 |
| PLEC | 601282 | chr8_143916616_G/A | ENSG00000178209 | ENST00000527303 | missense_variant | 9943 | 9905 | 3302 | T/M | aCg/aTg | rs113513807 | MODERATE | tolerated (0.27) | benign (0.01) | FE164 | 1 |

| | | | | | | | | | | | | | | | | |
|---|---|---|---|---|---|---|---|---|---|---|---|---|---|---|---|---|
| PLEC | 601282 | chr8_143925331_C/T | ENSG00000178209 | ENST00000322810 | missense_variant | 5179 | 5009 | 1670 | R/Q | cGg/cAg | rs201430180 | MODERATE | tolerated_low_confidence (0.09) | benign (0.003) | FE164 | 1 |
| PLEC | 601282 | chr8_143925331_C/T | ENSG00000178209 | ENST00000345136 | missense_variant | 4727 | 4598 | 1533 | R/Q | cGg/cAg | rs201430180 | MODERATE | tolerated (0.09) | benign (0.009) | FE164 | 1 |
| PLEC | 601282 | chr8_143925331_C/T | ENSG00000178209 | ENST00000354589 | missense_variant | 4666 | 4598 | 1533 | R/Q | cGg/cAg | rs201430180 | MODERATE | tolerated (0.1) | benign (0.009) | FE164 | 1 |
| PLEC | 601282 | chr8_143916616_G/A | ENSG00000178209 | ENST00000322810 | missense_variant | 13786 | 13616 | 4539 | T/M | aCg/aTg | rs113513807 | MODERATE | tolerated (0.12) | benign (0.005) | FE164 | 1 |
| PLEC | 601282 | chr8_143916616_G/A | ENSG00000178209 | ENST00000345136 | missense_variant | 13334 | 13205 | 4402 | T/M | aCg/aTg | rs113513807 | MODERATE | tolerated (0.09) | benign (0.003) | FE164 | 1 |
| PLEC | 601282 | chr8_143916616_G/A | ENSG00000178209 | ENST00000354589 | missense_variant | 13273 | 13205 | 4402 | T/M | aCg/aTg | rs113513807 | MODERATE | tolerated (0.15) | benign (0.003) | FE164 | 1 |
| PLEC | 601282 | chr8_143925331_C/T | ENSG00000178209 | ENST00000398774 | missense_variant | 4576 | 4502 | 1501 | R/Q | cGg/cAg | rs201430180 | MODERATE | tolerated (0.09) | benign (0.009) | FE164 | 1 |
| PLEC | 601282 | chr8_143925331_C/T | ENSG00000178209 | ENST00000436759 | missense_variant | 4717 | 4679 | 1560 | R/Q | cGg/cAg | rs201430180 | MODERATE | tolerated (0.09) | benign (0.009) | FE164 | 1 |
| PLEC | 601282 | chr8_143925331_C/T | ENSG00000178209 | ENST00000527096 | missense_variant | 4667 | 4667 | 1556 | R/Q | cGg/cAg | rs201430180 | MODERATE | tolerated (0.09) | benign (0.009) | FE164 | 1 |
| PLEC | 601282 | chr8_143934868_C/T | ENSG00000178209 | ENST00000527303 | missense_variant | 1006 | 968 | 323 | R/Q | cGa/cAa | rs138924815,CM101531 | MODERATE | deleterious (0) | probably_damaging (0.994) | FE194 | 1 |
| PLEC | 601282 | chr8_143934868_C/T | ENSG00000178209 | ENST00000528025 | missense_variant | 1025 | 1019 | 340 | R/Q | cGa/cAa | rs138924815,CM101531 | MODERATE | deleterious (0.01) | benign (0.04) | FE194 | 1 |
| PLEC | 601282 | chr8_143922157_G/C | ENSG00000178209 | ENST00000357649 | missense_variant | 7685 | 7676 | 2559 | A/G | gCg/gGg | - | MODERATE | deleterious_low_confidence (0) | probably_damaging (0.999) | FE165 | 1 |
| PLEC | 601282 | chr8_143922157_G/C | ENSG00000178209 | ENST00000398774 | missense_variant | 7642 | 7568 | 2523 | A/G | gCg/gGg | - | MODERATE | deleterious_low_confidence (0) | probably_damaging (0.999) | FE165 | 1 |
| PLEC | 601282 | chr8_143922157_G/C | ENSG00000178209 | ENST00000436759 | missense_variant | 7783 | 7745 | 2582 | A/G | gCg/gGg | - | MODERATE | deleterious_low_confidence (0) | probably_damaging (0.999) | FE165 | 1 |
| PLEC | 601282 | chr8_143922157_G/C | ENSG00000178209 | ENST00000527096 | missense_variant | 7733 | 7733 | 2578 | A/G | gCg/gGg | - | MODERATE | deleterious_low_confidence (0) | probably_damaging (0.999) | FE165 | 1 |
| PLEC | 601282 | chr8_143922157_G/C | ENSG00000178209 | ENST00000527303 | missense_variant | 4402 | 4364 | 1455 | A/G | gCg/gGg | - | MODERATE | deleterious_low_confidence (0.01) | probably_damaging (0.999) | FE165 | 1 |
| PLEC | 601282 | chr8_143925331_C/T | ENSG00000178209 | ENST00000354958 | missense_variant | 4681 | 4532 | 1511 | R/Q | cGg/cAg | rs201430180 | MODERATE | tolerated (0.09) | benign (0.009) | FE164 | 1 |
| PLEC | 601282 | chr8_143925331_C/T | ENSG00000178209 | ENST00000356346 | missense_variant | 4613 | 4556 | 1519 | R/Q | cGg/cAg | rs201430180 | MODERATE | tolerated (0.09) | benign (0.009) | FE164 | 1 |
| PLEC | 601282 | chr8_143925331_C/T | ENSG00000178209 | ENST00000357649 | missense_variant | 4619 | 4610 | 1537 | R/Q | cGg/cAg | rs201430180 | MODERATE | tolerated (0.09) | benign (0.009) | FE164 | 1 |
| PLEC | 601282 | chr8_143922157_G/C | ENSG00000178209 | ENST00000322810 | missense_variant | 8245 | 8075 | 2692 | A/G | gCg/gGg | - | MODERATE | deleterious_low_confidence (0) | probably_damaging (0.997) | FE165 | 1 |
| PLEC | 601282 | chr8_143922157_G/C | ENSG00000178209 | ENST00000345136 | missense_variant | 7793 | 7664 | 2555 | A/G | gCg/gGg | - | MODERATE | deleterious_low_confidence (0) | probably_damaging (0.999) | FE165 | 1 |
| PLEC | 601282 | chr8_143922157_G/C | ENSG00000178209 | ENST00000354589 | missense_variant | 7732 | 7664 | 2555 | A/G | gCg/gGg | - | MODERATE | deleterious_low_confidence (0) | probably_damaging (0.999) | FE165 | 1 |
| PLEC | 601282 | chr8_143922157_G/C | ENSG00000178209 | ENST00000354958 | missense_variant | 7747 | 7598 | 2533 | A/G | gCg/gGg | - | MODERATE | deleterious_low_confidence (0) | probably_damaging (0.999) | FE165 | 1 |
| PLEC | 601282 | chr8_143922157_G/C | ENSG00000178209 | ENST00000356346 | missense_variant | 7679 | 7622 | 2541 | A/G | gCg/gGg | - | MODERATE | deleterious_low_confidence (0) | probably_damaging (0.999) | FE165 | 1 |
| PLEC | 601282 | chr8_143934868_C/T | ENSG00000178209 | ENST00000345136 | missense_variant | 1016 | 887 | 296 | R/Q | cGa/cAa | rs138924815,CM101531 | MODERATE | deleterious (0.02) | probably_damaging (0.994) | FE194 | 1 |
| PLEC | 601282 | chr8_143934868_C/T | ENSG00000178209 | ENST00000354589 | missense_variant | 955 | 887 | 296 | R/Q | cGa/cAa | rs138924815,CM101531 | MODERATE | deleterious (0.02) | probably_damaging (0.994) | FE194 | 1 |
| PLEC | 601282 | chr8_143934868_C/T | ENSG00000178209 | ENST00000354958 | missense_variant | 970 | 821 | 274 | R/Q | cGa/cAa | rs138924815,CM101531 | MODERATE | deleterious (0.02) | probably_damaging (0.994) | FE194 | 1 |
| PLEC | 601282 | chr8_143934868_C/T | ENSG00000178209 | ENST00000322810 | missense_variant | 1468 | 1298 | 433 | R/Q | cGa/cAa | rs138924815,CM101531 | MODERATE | deleterious (0.02) | probably_damaging (0.986) | FE194 | 1 |
| PLEC | 601282 | chr8_143934868_C/T | ENSG00000178209 | ENST00000357649 | missense_variant | 908 | 899 | 300 | R/Q | cGa/cAa | rs138924815,CM101531 | MODERATE | deleterious (0.02) | probably_damaging (0.994) | FE194 | 1 |
| PLEC | 601282 | chr8_143916616_G/A | ENSG00000178209 | ENST00000354958 | missense_variant | 13288 | 13139 | 4380 | T/M | aCg/aTg | rs113513807 | MODERATE | tolerated (0.15) | benign (0.003) | FE164 | 1 |
| PLEC | 601282 | chr8_143916616_G/A | ENSG00000178209 | ENST00000356346 | missense_variant | 13220 | 13163 | 4388 | T/M | aCg/aTg | rs113513807 | MODERATE | tolerated (0.16) | benign (0.003) | FE164 | 1 |
| PLEC | 601282 | chr8_143934868_C/T | ENSG00000178209 | ENST00000356346 | missense_variant | 902 | 845 | 282 | R/Q | cGa/cAa | rs138924815,CM101531 | MODERATE | deleterious (0.02) | probably_damaging (0.994) | FE194 | 1 |
| PLEC | 601282 | chr8_143934868_C/T | ENSG00000178209 | ENST00000527096 | missense_variant | 956 | 956 | 319 | R/Q | cGa/cAa | rs138924815,CM101531 | MODERATE | deleterious (0.02) | probably_damaging (0.994) | FE194 | 1 |
| PLEC | 601282 | chr8_143934868_C/T | ENSG00000178209 | ENST00000398774 | missense_variant | 865 | 791 | 264 | R/Q | cGa/cAa | rs138924815,CM101531 | MODERATE | deleterious (0.02) | probably_damaging (0.994) | FE194 | 1 |
| PLEC | 601282 | chr8_143916616_G/A | ENSG00000178209 | ENST00000357649 | missense_variant | 13226 | 13217 | 4406 | T/M | aCg/aTg | rs113513807 | MODERATE | tolerated (0.14) | benign (0.003) | FE164 | 1 |
| PLEC | 601282 | chr8_143934868_C/T | ENSG00000178209 | ENST00000436759 | missense_variant | 1006 | 968 | 323 | R/Q | cGa/cAa | rs138924815,CM101531 | MODERATE | deleterious (0.02) | probably_damaging (0.994) | FE194 | 1 |
| PLEKHA6 | 607771 | chr1_204248971_C/G | ENSG00000143850 | ENST00000272203 | splice_acceptor_variant | - | - | - | - | - | - | HIGH | - | - | FE165 | 1 |
| PLOD1 | 153454 | chr1_11965543_C/T | ENSG00000083444 | ENST00000196061 | missense_variant | 1597 | 1534 | 512 | R/C | Cgc/Tgc | rs138490756 | MODERATE | deleterious (0) | benign (0.2) | FE164 | 1 |
| PLXNA3 | 300022 | chrX_154464783_C/T | ENSG00000130827 | ENST00000369682 | missense_variant | 2133 | 1958 | 653 | P/L | cCc/cTc | rs139336954 | MODERATE | deleterious (0.02) | benign (0.407) | FE190 | 2 |
| PNISR | 616653 | chr6_99401630_T/C | ENSG00000132424 | ENST00000369239 | missense_variant,splice_region_variant | 1533 | 1328 | 443 | E/G | gAa/gGa | - | MODERATE | deleterious (0.01) | possibly_damaging (0.897) | FE194 | 1 |
| PNISR | 616653 | chr6_99401630_T/C | ENSG00000132424 | ENST00000438806 | missense_variant,splice_region_variant | 1476 | 1328 | 443 | E/G | gAa/gGa | - | MODERATE | deleterious (0.01) | possibly_damaging (0.897) | FE194 | 1 |
| PNISR | 616653 | chr6_99401630_T/C | ENSG00000132424 | ENST00000647811 | missense_variant,splice_region_variant | 1610 | 1328 | 443 | E/G | gAa/gGa | - | MODERATE | deleterious (0.01) | possibly_damaging (0.897) | FE194 | 1 |

| | | | | | | | | | | | | | | | | |
|---|---|---|---|---|---|---|---|---|---|---|---|---|---|---|---|---|
| POGK | NA | chr1_166841027_G/A | ENSG00000143157 | ENST00000367875 | missense_variant | 431 | 71 | 24 | R/Q | cGg/cAg | rs141874882 | MODERATE | tolerated_low_confidence (0.35) | benign (0) | FE130 | 1 |
| POLR1E | NA | chr9_37495920_A/G | ENSG00000137054 | ENST00000377792 | missense_variant | 1160 | 872 | 291 | Q/R | cAg/cGg | - | MODERATE | tolerated (0.57) | benign (0.003) | FE193 | 1 |
| POLR1E | NA | chr9_37495920_A/G | ENSG00000137054 | ENST00000377798 | missense_variant | 786 | 686 | 229 | Q/R | cAg/cGg | - | MODERATE | tolerated (0.87) | benign (0.001) | FE193 | 1 |
| POLR3H | NA | chr22_41543765_G/A | ENSG00000100413 | ENST00000432789 | missense_variant,NMD_transcript_variant | 663 | 337 | 113 | L/F | Ctt/Ttt | rs563673428 | MODERATE | - | unknown (0) | FE154 | 1 |
| PPARGC1B | 608886 | chr5_149832735_C/A | ENSG00000155846 | ENST00000309241 | missense_variant | 695 | 662 | 221 | T/N | aCc/aAc | rs139476696 | MODERATE | deleterious (0) | probably_damaging (0.997) | FE165 | 1 |
| PPAT | 172450 | chr4_56404115_A/C | ENSG00000128059 | ENST00000510643 | missense_variant,NMD_transcript_variant | 437 | 406 | 136 | S/A | Tcc/Gcc | rs558435178 | MODERATE | tolerated (0.42) | benign (0.001) | FE190 | 1 |
| PPCS | 609853 | chr1_42456878_C/G | ENSG00000127125 | ENST00000372561 | missense_variant | 336 | 313 | 105 | L/V | Ctg/Gtg | - | MODERATE | deleterious (0.01) | benign (0.219) | FE165 | 1 |
| PPCS | 609853 | chr1_42456878_C/G | ENSG00000127125 | ENST00000372560 | missense_variant | 340 | 313 | 105 | L/V | Ctg/Gtg | - | MODERATE | tolerated (0.06) | benign (0.185) | FE165 | 1 |
| PPP1CA | 176875 | chr11_67421025_G/C | ENSG00000172531 | ENST00000542876 | missense_variant | 159 | 110 | 37 | P/R | cCg/cGg | rs545975688 | MODERATE | deleterious_low_confidence (0) | benign (0.415) | FE194 | 1 |
| PPP1CA | 176875 | chr11_67421025_G/C | ENSG00000172531 | ENST00000546202 | missense_variant | 157 | 110 | 37 | P/R | cCg/cGg | rs545975688 | MODERATE | - | possibly_damaging (0.619) | FE194 | 1 |
| PPP1R12A | 602021 | chr12_79806178_C/G | ENSG00000058272 | ENST00000261207 | missense_variant | 1934 | 1811 | 604 | G/A | gGc/gCc | rs61756418 | MODERATE | tolerated (0.64) | possibly_damaging (0.572) | FE130 | 1 |
| PPP1R12A | 602021 | chr12_79806178_C/G | ENSG00000058272 | ENST00000437004 | missense_variant | 1939 | 1811 | 604 | G/A | gGc/gCc | rs61756418 | MODERATE | tolerated (0.43) | possibly_damaging (0.754) | FE130 | 1 |
| PPP1R12A | 602021 | chr12_79806178_C/G | ENSG00000058272 | ENST00000450142 | missense_variant | 1977 | 1811 | 604 | G/A | gGc/gCc | rs61756418 | MODERATE | tolerated (0.64) | possibly_damaging (0.572) | FE130 | 1 |
| PPP1R12A | 602021 | chr12_79806178_C/G | ENSG00000058272 | ENST00000546369 | missense_variant | 1689 | 1550 | 517 | G/A | gGc/gCc | rs61756418 | MODERATE | tolerated (0.35) | possibly_damaging (0.572) | FE130 | 1 |
| PPP1R12A | 602021 | chr12_79806178_C/G | ENSG00000058272 | ENST00000547330 | missense_variant | 2010 | 1811 | 604 | G/A | gGc/gCc | rs61756418 | MODERATE | tolerated (0.74) | possibly_damaging (0.459) | FE130 | 1 |
| PPP1R12A | 602021 | chr12_79806178_C/G | ENSG00000058272 | ENST00000553081 | missense_variant | 586 | 587 | 196 | G/A | gGc/gCc | rs61756418 | MODERATE | tolerated (0.42) | possibly_damaging (0.754) | FE130 | 1 |
| PPP1R12A | 602021 | chr12_79806178_C/G | ENSG00000058272 | ENST00000650220 | missense_variant | 814 | 815 | 272 | G/A | gGc/gCc | rs61756418 | MODERATE | tolerated (0.4) | benign (0.23) | FE130 | 1 |
| PPP1R7 | 602877 | chr2_241158503_A/G | ENSG00000115685 | ENST00000423280 | missense_variant | 172 | 80 | 27 | Y/C | tAt/tGt | rs116718216 | MODERATE | tolerated (0.71) | benign (0) | FE136 | 1 |
| PPP1R7 | 602877 | chr2_241158503_A/G | ENSG00000115685 | ENST00000450367 | missense_variant | 199 | 200 | 67 | Y/C | tAt/tGt | rs116718216 | MODERATE | tolerated (0.25) | benign (0) | FE136 | 1 |
| PPP1R7 | 602877 | chr2_241158503_A/G | ENSG00000115685 | ENST00000438799 | missense_variant | 338 | 209 | 70 | Y/C | tAt/tGt | rs116718216 | MODERATE | tolerated (0.29) | benign (0) | FE136 | 1 |
| PPP1R7 | 602877 | chr2_241158503_A/G | ENSG00000115685 | ENST00000401987 | missense_variant | 128 | 128 | 43 | Y/C | tAt/tGt | rs116718216 | MODERATE | tolerated (0.23) | benign (0) | FE136 | 1 |
| PPP1R7 | 602877 | chr2_241158503_A/G | ENSG00000115685 | ENST00000427172 | missense_variant | 284 | 284 | 95 | Y/C | tAt/tGt | rs116718216 | MODERATE | tolerated (0.19) | benign (0.062) | FE136 | 1 |
| PPP1R7 | 602877 | chr2_241158503_A/G | ENSG00000115685 | ENST00000404405 | missense_variant | 335 | 257 | 86 | Y/C | tAt/tGt | rs116718216 | MODERATE | tolerated (0.25) | benign (0) | FE136 | 1 |
| PPP1R7 | 602877 | chr2_241158503_A/G | ENSG00000115685 | ENST00000407025 | missense_variant | 565 | 257 | 86 | Y/C | tAt/tGt | rs116718216 | MODERATE | tolerated (0.31) | benign (0) | FE136 | 1 |
| PPP1R7 | 602877 | chr2_241158503_A/G | ENSG00000115685 | ENST00000406106 | missense_variant | 273 | 257 | 86 | Y/C | tAt/tGt | rs116718216 | MODERATE | tolerated (0.26) | benign (0.003) | FE136 | 1 |
| PPP1R7 | 602877 | chr2_241158503_A/G | ENSG00000115685 | ENST00000234038 | missense_variant | 286 | 257 | 86 | Y/C | tAt/tGt | rs116718216 | MODERATE | tolerated (0.31) | benign (0) | FE136 | 1 |
| PPP1R7 | 602877 | chr2_241158503_A/G | ENSG00000115685 | ENST00000272983 | missense_variant | 966 | 128 | 43 | Y/C | tAt/tGt | rs116718216 | MODERATE | tolerated (0.27) | benign (0) | FE136 | 1 |
| PPP1R7 | 602877 | chr2_241158503_A/G | ENSG00000115685 | ENST00000439916 | missense_variant | 304 | 275 | 92 | Y/C | tAt/tGt | rs116718216 | MODERATE | tolerated (0.26) | benign (0) | FE136 | 1 |
| PPP1R7 | 602877 | chr2_241158503_A/G | ENSG00000115685 | ENST00000402734 | missense_variant | 193 | 80 | 27 | Y/C | tAt/tGt | rs116718216 | MODERATE | tolerated (0.48) | benign (0) | FE136 | 1 |
| PPP6R1 | 610875 | chr19_55231829_G/C | ENSG00000105063 | ENST00000412770 | missense_variant | 2868 | 2279 | 760 | P/R | cCt/cGt | rs781518117 | MODERATE | deleterious (0.02) | benign (0.159) | FE136 | 1 |
| PRDM10 | 618319 | chr11_129925052_A/T | ENSG00000170325 | ENST00000358825 | missense_variant | 1952 | 1720 | 574 | S/T | Tcc/Acc | - | MODERATE | deleterious (0.02) | benign (0.297) | FE164 | 1 |
| PRDM10 | 618319 | chr11_129925052_A/T | ENSG00000170325 | ENST00000360871 | missense_variant | 1940 | 1708 | 570 | S/T | Tcc/Acc | - | MODERATE | deleterious (0.02) | benign (0.164) | FE164 | 1 |
| PRDM10 | 618319 | chr11_129925052_A/T | ENSG00000170325 | ENST00000528746 | missense_variant | 1862 | 1630 | 544 | S/T | Tcc/Acc | - | MODERATE | deleterious (0.01) | benign (0.253) | FE164 | 1 |
| PRDM10 | 618319 | chr11_129925052_A/T | ENSG00000170325 | ENST00000533431 | missense_variant | 907 | 859 | 287 | S/T | Tcc/Acc | - | MODERATE | tolerated (0.06) | benign (0.253) | FE164 | 1 |
| PRDM10 | 618319 | chr11_129925052_A/T | ENSG00000170325 | ENST00000423662 | missense_variant | 1678 | 1462 | 488 | S/T | Tcc/Acc | - | MODERATE | deleterious (0.01) | benign (0.164) | FE164 | 1 |
| PRDM10 | 618319 | chr11_129925052_A/T | ENSG00000170325 | ENST00000526082 | missense_variant | 1517 | 1462 | 488 | S/T | Tcc/Acc | - | MODERATE | deleterious (0.02) | benign (0.164) | FE164 | 1 |
| PRDM10 | 618319 | chr11_129925052_A/T | ENSG00000170325 | ENST00000304538 | missense_variant | 1597 | 1450 | 484 | S/T | Tcc/Acc | - | MODERATE | deleterious (0.01) | benign (0.213) | FE164 | 1 |
| PRDM10 | 618319 | chr11_129918644_G/C | ENSG00000170325 | ENST00000304538 | missense_variant | 1998 | 1851 | 617 | I/M | atC/atG | rs545610604 | MODERATE | deleterious (0.02) | possibly_damaging (0.66) | FE187 | 1 |
| PRDM10 | 618319 | chr11_129918644_G/C | ENSG00000170325 | ENST00000358825 | missense_variant | 2353 | 2121 | 707 | I/M | atC/atG | rs545610604 | MODERATE | deleterious (0.04) | possibly_damaging (0.531) | FE187 | 1 |
| PRDM10 | 618319 | chr11_129918644_G/C | ENSG00000170325 | ENST00000360871 | missense_variant | 2341 | 2109 | 703 | I/M | atC/atG | rs545610604 | MODERATE | deleterious (0.03) | benign (0.255) | FE187 | 1 |
| PRDM10 | 618319 | chr11_129918644_G/C | ENSG00000170325 | ENST00000423662 | missense_variant | 2079 | 1863 | 621 | I/M | atC/atG | rs545610604 | MODERATE | deleterious (0.03) | benign (0.255) | FE187 | 1 |
| PRDM10 | 618319 | chr11_129918644_G/C | ENSG00000170325 | ENST00000526082 | missense_variant | 1918 | 1863 | 621 | I/M | atC/atG | rs545610604 | MODERATE | deleterious (0.03) | benign (0.255) | FE187 | 1 |

| | | | | | | | | | | | | | | | | |
|---|---|---|---|---|---|---|---|---|---|---|---|---|---|---|---|---|
| PRDM10 | 618319 | chr11_129918644_G/C | ENSG00000170325 | ENST00000528746 | missense_variant | 2263 | 2031 | 677 | I/M | atC/atG | rs545610604 | MODERATE | deleterious (0.02) | benign (0.131) | FE187 | 1 |
| PRDM10 | 618319 | chr11_129918644_G/C | ENSG00000170325 | ENST00000533431 | missense_variant | 1308 | 1260 | 420 | I/M | atC/atG | rs545610604 | MODERATE | tolerated (0.08) | benign (0.131) | FE187 | 1 |
| PREP | 600400 | chr6_105368935_G/T | ENSG00000085377 | ENST00000369110 | missense_variant | 1450 | 487 | 163 | P/T | Cct/Act | rs202011848 | MODERATE | deleterious (0.02) | possibly_damaging (0.898) | FE136 | 1 |
| PRPF18 | 604993 | chr10_13600257_A/G | ENSG00000165630 | ENST00000320054 | missense_variant | 398 | 113 | 38 | E/G | gAg/gGg | rs142307573 | MODERATE | tolerated (0.33) | benign (0.116) | FE164 | 1 |
| PRPF18 | 604993 | chr10_13600257_A/G | ENSG00000165630 | ENST00000378572 | missense_variant | 280 | 158 | 53 | E/G | gAg/gGg | rs142307573 | MODERATE | tolerated (0.26) | benign (0) | FE164 | 1 |
| PRPF18 | 604993 | chr10_13600257_A/G | ENSG00000165630 | ENST00000417658 | missense_variant | 454 | 140 | 47 | E/G | gAg/gGg | rs142307573 | MODERATE | tolerated (0.12) | benign (0.033) | FE164 | 1 |
| PRR14L | NA | chr22_31714903_G/A | ENSG00000183530 | ENST00000327423 | missense_variant | 3127 | 2936 | 979 | P/L | cCa/cTa | rs774171969 | MODERATE | tolerated (0.13) | benign (0.039) | FE187 | 1 |
| PSAP | 176801 | chr10_71820257_C/T | ENSG00000197746 | ENST00000394934 | missense_variant | 1101 | 997 | 333 | D/N | Gac/Aac | rs774816225 | MODERATE | tolerated (0.14) | benign (0.012) | FE193 | 1 |
| PSAP | 176801 | chr10_71820257_C/T | ENSG00000197746 | ENST00000394936 | missense_variant | 1018 | 988 | 330 | D/N | Gac/Aac | rs774816225 | MODERATE | tolerated (0.14) | possibly_damaging (0.449) | FE193 | 1 |
| PSAP | 176801 | chr10_71820257_C/T | ENSG00000197746 | ENST00000633965 | missense_variant | 398 | 400 | 134 | D/N | Gac/Aac | rs774816225 | MODERATE | tolerated (0.22) | benign (0.042) | FE193 | 1 |
| PSME3IP1 | 617766 | chr16_57167154_T/A | ENSG00000172775 | ENST00000309137 | missense_variant | 655 | 421 | 141 | I/L | Ata/Tta | rs143746498,COSV58429330 | MODERATE | tolerated (0.38) | benign (0) | FE164 | 1 |
| PSMG3 | 617528 | chr7_1569176_C/G | ENSG00000157778 | ENST00000252329 | missense_variant | 717 | 164 | 55 | S/T | aGt/aCt | rs34407549 | MODERATE | tolerated (0.36) | benign (0) | FE130 | 1 |
| PSMG3 | 617528 | chr7_1569176_C/G | ENSG00000157778 | ENST00000404674 | missense_variant | 222 | 164 | 55 | S/T | aGt/aCt | rs34407549 | MODERATE | tolerated (0.36) | benign (0) | FE130 | 1 |
| PSMG3 | 617528 | chr7_1569176_C/G | ENSG00000157778 | ENST00000288607 | missense_variant | 857 | 164 | 55 | S/T | aGt/aCt | rs34407549 | MODERATE | tolerated (0.36) | benign (0) | FE164 | 1 |
| PSMG3 | 617528 | chr7_1569176_C/G | ENSG00000157778 | ENST00000404674 | missense_variant | 222 | 164 | 55 | S/T | aGt/aCt | rs34407549 | MODERATE | tolerated (0.36) | benign (0) | FE164 | 1 |
| PSMG3 | 617528 | chr7_1569176_C/G | ENSG00000157778 | ENST00000252329 | missense_variant | 717 | 164 | 55 | S/T | aGt/aCt | rs34407549 | MODERATE | tolerated (0.36) | benign (0) | FE164 | 1 |
| PSMG3 | 617528 | chr7_1569176_C/G | ENSG00000157778 | ENST00000288607 | missense_variant | 857 | 164 | 55 | S/T | aGt/aCt | rs34407549 | MODERATE | tolerated (0.36) | benign (0) | FE130 | 1 |
| PTPMT1 | 609538 | chr11_47571628_G/C | ENSG00000110536 | ENST00000326656 | stop_lost | 437 | 413 | 138 | */S | tGa/tCa | rs190036505 | HIGH | - | - | FE187 | 1 |
| PTPMT1 | 609538 | chr11_47571628_G/C | ENSG00000110536 | ENST00000326674 | stop_lost | 629 | 605 | 202 | */S | tGa/tCa | rs190036505 | HIGH | - | - | FE187 | 1 |
| RAB11FIP2 | 608599 | chr10_118015066_G/A | ENSG00000107560 | ENST00000355624 | missense_variant,splice_region_variant | 2088 | 1310 | 437 | P/L | cCg/cTg | rs141500760 | MODERATE | tolerated (0.36) | benign (0) | FE187 | 1 |
| RAB21 | 612398 | chr12_71785666_C/T | ENSG00000080371 | ENST00000261263 | missense_variant | 938 | 671 | 224 | S/F | tCt/tTt | rs61754230 | MODERATE | deleterious_low_confidence (0) | probably_damaging (0.974) | FE165 | 1 |
| RACGAP1 | 604980 | chr12_49976972_G/A | ENSG00000161800 | ENST00000548961 | missense_variant | 338 | 332 | 111 | S/F | tCt/tTt | rs148060886 | MODERATE | deleterious_low_confidence (0.01) | benign (0.003) | FE190 | 1 |
| RAD1 | 603153 | chr5_34911809_G/C | ENSG00000113456 | ENST00000341754 | missense_variant | 1210 | 311 | 104 | T/S | aCt/aGt | rs1805328 | MODERATE | tolerated (1) | benign (0) | FE165 | 2 |
| RAD1 | 603153 | chr5_34911809_G/C | ENSG00000113456 | ENST00000382038 | missense_variant | 469 | 311 | 104 | T/S | aCt/aGt | rs1805328 | MODERATE | tolerated (1) | benign (0) | FE165 | 2 |
| RAD1 | 603153 | chr5_34911809_G/C | ENSG00000113456 | ENST00000325577 | missense_variant,NMD_transcript_variant | 404 | 202 | 68 | L/V | Ctt/Gtt | rs1805328 | MODERATE | deleterious_low_confidence (0) | possibly_damaging (0.578) | FE165 | 2 |
| RAN | 601179 | chr12_130872100_G/A | ENSG00000132341 | ENST00000448750 | missense_variant | 64 | 64 | 22 | G/R | Gga/Aga | rs11546491 | MODERATE | deleterious_low_confidence (0.01) | probably_damaging (0.969) | FE130 | 1 |
| RAN | 601179 | chr12_130872100_G/A | ENSG00000132341 | ENST00000536606 | missense_variant,NMD_transcript_variant | 54 | 55 | 19 | G/R | Gga/Aga | rs11546491 | MODERATE | deleterious (0.01) | benign (0.324) | FE130 | 1 |
| RANBP2 | 601181 | chr2_108748943_C/T | ENSG00000153201 | ENST00000283195 | missense_variant | 1212 | 1087 | 363 | R/C | Cgt/Tgt | rs149207118,COSV51704283 | MODERATE | tolerated (0.13) | benign (0.001) | FE154 | 1 |
| RANBP2 | 601181 | chr2_108765299_G/T | ENSG00000153201 | ENST00000283195 | missense_variant | 4885 | 4760 | 1587 | G/V | gGt/gTt | rs148677577 | MODERATE | deleterious (0) | possibly_damaging (0.651) | FE194 | 1 |
| RAPGEF6 | 610499 | chr5_131431342_G/C | ENSG00000158987 | ENST00000296859 | missense_variant | 4087 | 4006 | 1336 | L/V | Ctc/Gtc | rs34896346 | MODERATE | tolerated_low_confidence (0.58) | benign (0) | FE165 | 1 |
| RASA1 | 139150 | chr5_87349234_C/T | ENSG00000145715 | ENST00000274376 | missense_variant | 1692 | 1123 | 375 | L/F | Ctt/Ttt | COSV57199839 | MODERATE | deleterious (0) | probably_damaging (0.998) | FE190 | 1 |
| RBBP8 | 604124 | chr18_23022190_G/A | ENSG00000101773 | ENST00000327155 | missense_variant | 2851 | 2516 | 839 | R/Q | cGa/cAa | rs140196819 | MODERATE | deleterious (0) | probably_damaging (0.998) | FE130 | 1 |
| RBBP8 | 604124 | chr18_23022190_G/A | ENSG00000101773 | ENST00000360790 | missense_variant | 2654 | 2531 | 844 | R/Q | cGa/cAa | rs140196819 | MODERATE | deleterious (0) | probably_damaging (0.999) | FE130 | 1 |
| RBBP8 | 604124 | chr18_23022190_G/A | ENSG00000101773 | ENST00000399722 | missense_variant | 2867 | 2516 | 839 | R/Q | cGa/cAa | rs140196819 | MODERATE | deleterious (0) | probably_damaging (0.998) | FE130 | 1 |
| RBBP8 | 604124 | chr18_23022190_G/A | ENSG00000101773 | ENST00000399725 | missense_variant | 2836 | 2419 | 807 | D/N | Gat/Aat | rs140196819 | MODERATE | deleterious_low_confidence (0.03) | possibly_damaging (0.632) | FE130 | 1 |
| RBBP8 | 604124 | chr18_23022190_G/A | ENSG00000101773 | ENST00000581687 | missense_variant | 191 | 50 | 17 | R/Q | cGa/cAa | rs140196819 | MODERATE | deleterious (0) | probably_damaging (0.998) | FE130 | 1 |
| RBBP8 | 604124 | chr18_23022190_G/A | ENSG00000101773 | ENST00000583057 | missense_variant | 790 | 790 | 264 | D/N | Gat/Aat | rs140196819 | MODERATE | deleterious_low_confidence (0.04) | probably_damaging (0.99) | FE130 | 1 |
| RBM19 | 616444 | chr12_113945854_C/T | ENSG00000122965 | ENST00000261741 | missense_variant | 1698 | 1600 | 534 | A/T | Gcc/Acc | rs62621126 | MODERATE | tolerated (1) | benign (0.003) | FE165 | 1 |
| RBM19 | 616444 | chr12_113945854_C/T | ENSG00000122965 | ENST00000392561 | missense_variant | 1683 | 1600 | 534 | A/T | Gcc/Acc | rs62621126 | MODERATE | tolerated (1) | benign (0.003) | FE165 | 1 |
| RBM19 | 616444 | chr12_113945854_C/T | ENSG00000122965 | ENST00000545145 | missense_variant | 1679 | 1600 | 534 | A/T | Gcc/Acc | rs62621126 | MODERATE | tolerated (1) | benign (0.003) | FE165 | 1 |
| RBM25 | 612427 | chr14_73096937_C/T | ENSG00000119707 | ENST00000261973 | missense_variant | 753 | 566 | 189 | T/I | aCt/aTt | - | MODERATE | tolerated (0.19) | benign (0.074) | FE106 | 1 |
| RBM25 | 612427 | chr14_73096937_C/T | ENSG00000119707 | ENST00000525321 | missense_variant | 740 | 566 | 189 | T/I | aCt/aTt | - | MODERATE | tolerated (0.19) | benign (0.003) | FE106 | 1 |

| | | | | | | | | | | | | | | | | |
|---|---|---|---|---|---|---|---|---|---|---|---|---|---|---|---|---|
| RBM25 | 612427 | chr14_73096937_C/T | ENSG00000119707 | ENST00000526754 | missense_variant | 738 | 566 | 189 | T/I | aCt/aTt | - | MODERATE | tolerated (0.19) | benign (0.04) | FE106 | 1 |
| RBM25 | 612427 | chr14_73096937_C/T | ENSG00000119707 | ENST00000527432 | missense_variant | 823 | 566 | 189 | T/I | aCt/aTt | - | MODERATE | tolerated (0.19) | benign (0.074) | FE106 | 1 |
| RBM25 | 612427 | chr14_73096937_C/T | ENSG00000119707 | ENST00000531500 | missense_variant | 876 | 566 | 189 | T/I | aCt/aTt | - | MODERATE | tolerated (0.19) | benign (0.074) | FE106 | 1 |
| RCC1L | NA | chr7_75058755_T/C | ENSG00000274523 | ENST00000614461 | missense_variant | 946 | 802 | 268 | N/D | Aat/Gat | rs868974204 | MODERATE | tolerated (0.35) | benign (0.001) | FE154 | 1 |
| RCC1L | NA | chr7_75058755_T/C | ENSG00000274523 | ENST00000618035 | missense_variant | 946 | 802 | 268 | N/D | Aat/Gat | rs868974204 | MODERATE | tolerated (0.27) | benign (0.001) | FE154 | 1 |
| RCC1L | NA | chr7_75058755_T/C | ENSG00000274523 | ENST00000610322 | missense_variant | 867 | 802 | 268 | N/D | Aat/Gat | rs868974204 | MODERATE | tolerated (0.35) | benign (0.005) | FE154 | 1 |
| RCE1 | 605385 | chr11_66844366_T/G | ENSG00000173653 | ENST00000524506 | splice_donor_variant | - | - | - | - | - | - | HIGH | - | - | FE106 | 1 |
| RCE1 | 605385 | chr11_66844366_T/G | ENSG00000173653 | ENST00000524849 | splice_donor_variant,NMD_transcript_variant | - | - | - | - | - | - | HIGH | - | - | FE106 | 1 |
| RCE1 | 605385 | chr11_66844366_T/G | ENSG00000173653 | ENST00000525356 | splice_donor_variant | - | - | - | - | - | - | HIGH | - | - | FE106 | 1 |
| RCE1 | 605385 | chr11_66844366_T/G | ENSG00000173653 | ENST00000532775 | splice_donor_variant,non_coding_transcript_variant | - | - | - | - | - | - | HIGH | - | - | FE106 | 1 |
| RCE1 | 605385 | chr11_66844366_T/G | ENSG00000173653 | ENST00000533277 | splice_donor_variant,non_coding_transcript_variant | - | - | - | - | - | - | HIGH | - | - | FE106 | 1 |
| RCE1 | 605385 | chr11_66844366_T/G | ENSG00000173653 | ENST00000534645 | splice_donor_variant,non_coding_transcript_variant | - | - | - | - | - | - | HIGH | - | - | FE106 | 1 |
| RCE1 | 605385 | chr11_66844366_T/G | ENSG00000173653 | ENST00000309657 | splice_donor_variant | - | - | - | - | - | - | HIGH | - | - | FE106 | 1 |
| RELA | 164014 | chr11_65658293_C/T | ENSG00000173039 | ENST00000308639 | missense_variant | 1120 | 862 | 288 | D/N | Gat/Aat | rs61759893 | MODERATE | deleterious (0.02) | possibly_damaging (0.732) | FE136 | 1 |
| RELA | 164014 | chr11_65658293_C/T | ENSG00000173039 | ENST00000406246 | missense_variant | 955 | 871 | 291 | D/N | Gat/Aat | rs61759893 | MODERATE | deleterious (0.01) | possibly_damaging (0.543) | FE136 | 1 |
| RELA | 164014 | chr11_65658293_C/T | ENSG00000173039 | ENST00000532999 | missense_variant | 990 | 904 | 302 | D/N | Gat/Aat | rs61759893 | MODERATE | deleterious (0.04) | possibly_damaging (0.481) | FE136 | 1 |
| RELA | 164014 | chr11_65658293_C/T | ENSG00000173039 | ENST00000612991 | missense_variant | 1011 | 871 | 291 | D/N | Gat/Aat | rs61759893 | MODERATE | deleterious (0.05) | benign (0.378) | FE136 | 1 |
| RELA | 164014 | chr11_65658293_C/T | ENSG00000173039 | ENST00000525693 | missense_variant | 934 | 871 | 291 | D/N | Gat/Aat | rs61759893 | MODERATE | tolerated (0.28) | benign (0.028) | FE136 | 1 |
| RELA | 164014 | chr11_65658293_C/T | ENSG00000173039 | ENST00000526283 | missense_variant,NMD_transcript_variant | 752 | 689 | 230 | R/K | aGa/aAa | rs61759893 | MODERATE | tolerated_low_confidence (0.74) | benign (0.069) | FE136 | 1 |
| RFC2 | 600404 | chr7_74238987_G/A | ENSG00000049541 | ENST00000352131 | missense_variant,splice_region_variant | 599 | 593 | 198 | A/V | gCg/gTg | rs3135684 | MODERATE | deleterious (0.04) | benign (0.334) | FE187 | 1 |
| RFC2 | 600404 | chr7_74238987_G/A | ENSG00000049541 | ENST00000485545 | missense_variant,splice_region_variant | 72 | 74 | 25 | A/V | gCg/gTg | rs3135684 | MODERATE | tolerated (0.22) | probably_damaging (0.923) | FE187 | 1 |
| RFC2 | 600404 | chr7_74238987_G/A | ENSG00000049541 | ENST00000497430 | missense_variant,splice_region_variant | 261 | 263 | 88 | A/V | gCg/gTg | rs3135684 | MODERATE | deleterious (0.02) | benign (0.058) | FE187 | 1 |
| RFC2 | 600404 | chr7_74238987_G/A | ENSG00000049541 | ENST00000055077 | missense_variant,splice_region_variant | 711 | 695 | 232 | A/V | gCg/gTg | rs3135684 | MODERATE | deleterious (0.04) | benign (0.125) | FE187 | 1 |
| RFC2 | 600404 | chr7_74238987_G/A | ENSG00000049541 | ENST00000621097 | missense_variant,splice_region_variant | 663 | 386 | 129 | A/V | gCg/gTg | rs3135684 | MODERATE | deleterious (0.03) | benign (0.058) | FE187 | 1 |
| RFWD3 | 614151 | chr16_74649159_G/C | ENSG00000168411 | ENST00000361070 | missense_variant | 859 | 765 | 255 | I/M | atC/atG | rs762552515,COSV63099165 | MODERATE | tolerated (0.2) | benign (0.338) | FE193 | 1 |
| RFWD3 | 614151 | chr16_74649159_G/C | ENSG00000168411 | ENST00000571750 | missense_variant | 974 | 765 | 255 | I/M | atC/atG | rs762552515,COSV63099165 | MODERATE | tolerated (0.2) | benign (0.338) | FE193 | 1 |
| RHOT1 | 613888 | chr17_32209372_C/T | ENSG00000126858 | ENST00000358365 | missense_variant | 1989 | 1762 | 588 | L/F | Ctc/Ttc | rs144578639 | MODERATE | tolerated (0.68) | benign (0.029) | FE193 | 1 |
| RNF4 | 602850 | chr4_2468928_G/C | ENSG00000063978 | ENST00000504224 | splice_donor_variant | - | - | - | - | - | rs17132599 | HIGH | - | - | FE154 | 1 |
| RNMT | 603514 | chr18_13731575_G/C | ENSG00000101654 | ENST00000262173 | missense_variant | 102 | 58 | 20 | A/P | Gcg/Ccg | rs61730997 | MODERATE | tolerated_low_confidence (0.06) | benign (0.003) | FE194 | 1 |
| RNMT | 603514 | chr18_13731575_G/C | ENSG00000101654 | ENST00000383314 | missense_variant | 298 | 58 | 20 | A/P | Gcg/Ccg | rs61730997 | MODERATE | tolerated_low_confidence (0.06) | benign (0.003) | FE194 | 1 |
| RNMT | 603514 | chr18_13731575_G/C | ENSG00000101654 | ENST00000543302 | missense_variant | 170 | 58 | 20 | A/P | Gcg/Ccg | rs61730997 | MODERATE | tolerated_low_confidence (0.06) | benign (0.003) | FE194 | 1 |
| RNMT | 603514 | chr18_13731575_G/C | ENSG00000101654 | ENST00000589866 | missense_variant | 418 | 58 | 20 | A/P | Gcg/Ccg | rs61730997 | MODERATE | tolerated_low_confidence (0.06) | benign (0.003) | FE194 | 1 |
| RNMT | 603514 | chr18_13731575_G/C | ENSG00000101654 | ENST00000591746 | missense_variant | 326 | 58 | 20 | A/P | Gcg/Ccg | rs61730997 | MODERATE | tolerated (0.05) | benign (0.003) | FE194 | 1 |
| RNMT | 603514 | chr18_13731575_G/C | ENSG00000101654 | ENST00000592764 | missense_variant | 255 | 58 | 20 | A/P | Gcg/Ccg | rs61730997 | MODERATE | deleterious_low_confidence (0.05) | benign (0.011) | FE194 | 1 |
| ROR2 | 602337 | chr9_91724099_G/A | ENSG00000169071 | ENST00000375708 | missense_variant | 2660 | 2395 | 799 | P/S | Ccc/Tcc | rs141235720 | MODERATE | tolerated_low_confidence (0.06) | probably_damaging (0.996) | FE136 | 1 |
| RORB | 601972 | chr9_74642608_G/A | ENSG00000198963 | ENST00000376896 | missense_variant | 1072 | 430 | 144 | G/R | Ggg/Agg | rs571973540 | MODERATE | deleterious (0.02) | possibly_damaging (0.588) | FE193 | 1 |
| RORB | 601972 | chr9_74662477_A/G | ENSG00000198963 | ENST00000376896 | missense_variant | 1405 | 763 | 255 | R/G | Agg/Ggg | - | MODERATE | tolerated (0.38) | possibly_damaging (0.49) | FE154 | 1 |
| RPL18 | 604179 | chr19_48617819_T/C | ENSG00000063177 | ENST00000084795 | missense_variant | 66 | 68 | 23 | Q/R | cAg/cGg | - | MODERATE | deleterious (0.05) | possibly_damaging (0.782) | FE190 | 1 |
| RPL18 | 604179 | chr19_48617819_T/C | ENSG00000063177 | ENST00000547897 | missense_variant | 87 | 62 | 21 | Q/R | cAg/cGg | - | MODERATE | tolerated (0.06) | possibly_damaging (0.795) | FE190 | 1 |
| RPL18 | 604179 | chr19_48617819_T/C | ENSG00000063177 | ENST00000549273 | missense_variant | 96 | 62 | 21 | Q/R | cAg/cGg | - | MODERATE | tolerated (0.06) | benign (0.088) | FE190 | 1 |
| RPL18 | 604179 | chr19_48617819_T/C | ENSG00000063177 | ENST00000549370 | missense_variant,NMD_transcript_variant | 95 | 62 | 21 | Q/R | cAg/cGg | - | MODERATE | deleterious (0) | possibly_damaging (0.888) | FE190 | 1 |
| RPL18 | 604179 | chr19_48617819_T/C | ENSG00000063177 | ENST00000549920 | missense_variant | 97 | 62 | 21 | Q/R | cAg/cGg | - | MODERATE | deleterious (0.05) | benign (0.088) | FE190 | 1 |

| | | | | | | | | | | | | | | | | |
|---|---|---|---|---|---|---|---|---|---|---|---|---|---|---|---|---|
| RPL18 | 604179 | chr19_48617819_T/C | ENSG00000063177 | ENST00000550645 | missense_variant | 82 | 62 | 21 | Q/R | cAg/cGg | - | MODERATE | tolerated (0.08) | benign (0.234) | FE190 | 1 |
| RRS1 | 618311 | chr8_66430017_G/A | ENSG00000179041 | ENST00000320270 | missense_variant | 1004 | 886 | 296 | D/N | Gac/Aac | rs115935831 | MODERATE | tolerated (0.12) | benign (0.021) | FE154 | 1 |
| RSPRY1 | 616585 | chr16_57204737_A/G | ENSG00000159579 | ENST00000537866 | missense_variant | 952 | 79 | 27 | I/V | Ata/Gta | rs61746362 | MODERATE | tolerated_low_confidence (0.88) | benign (0) | FE193 | 1 |
| RTL9 | 300965 | chrX_110454215_T/C | ENSG00000243978 | ENST00000465301 | missense_variant | 3844 | 3598 | 1200 | W/R | Tgg/Cgg | rs41306249,COSV99081697 | MODERATE | deleterious_low_confidence (0) | probably_damaging (0.971) | FE193 | 1 |
| RTN4RL2 | 610462 | chr11_57476823_C/T | ENSG00000186907 | ENST00000335099 | missense_variant | 1513 | 1175 | 392 | A/V | gCg/gTg | rs141304388 | MODERATE | tolerated (0.38) | benign (0.001) | FE136 | 1 |
| RXYLT1 | 605862 | chr12_63808890_A/G | ENSG00000118600 | ENST00000261234 | missense_variant | 1182 | 1130 | 377 | K/R | aAg/aGg | rs763122658 | MODERATE | deleterious (0) | probably_damaging (0.997) | FE194 | 1 |
| RXYLT1 | 605862 | chr12_63808890_A/G | ENSG00000118600 | ENST00000537373 | missense_variant | 1298 | 350 | 117 | K/R | aAg/aGg | rs763122658 | MODERATE | deleterious (0) | probably_damaging (0.997) | FE194 | 1 |
| RYR2 | 180902 | chr1_237591776_A/G | ENSG00000198626 | ENST00000366574 | missense_variant | 4536 | 4198 | 1400 | S/G | Agc/Ggc | rs56229512 | MODERATE | tolerated (0.16) | benign (0.006) | FE193 | 1 |
| SACM1L | 606569 | chr3_45705193_A/C | ENSG00000211456 | ENST00000438671 | missense_variant | 338 | 134 | 45 | Q/P | cAa/cCa | rs143985480 | MODERATE | deleterious_low_confidence (0) | benign (0.001) | FE154 | 1 |
| SAMHD1 | 606754 | chr20_36911251_T/C | ENSG00000101347 | ENST00000262878 | missense_variant | 1437 | 1237 | 413 | I/V | Att/Gtt | - | MODERATE | tolerated (0.19) | possibly_damaging (0.66) | FE106 | 1 |
| SAMHD1 | 606754 | chr20_36911251_T/C | ENSG00000101347 | ENST00000643918 | missense_variant | 1275 | 1237 | 413 | I/V | Att/Gtt | - | MODERATE | tolerated (0.17) | possibly_damaging (0.773) | FE106 | 1 |
| SAMHD1 | 606754 | chr20_36911251_T/C | ENSG00000101347 | ENST00000644114 | missense_variant,NMD_transcript_variant | 1163 | 1165 | 389 | I/V | Att/Gtt | - | MODERATE | tolerated (0.25) | probably_damaging (0.996) | FE106 | 1 |
| SAMHD1 | 606754 | chr20_36911251_T/C | ENSG00000101347 | ENST00000646066 | missense_variant | 1027 | 1027 | 343 | I/V | Att/Gtt | - | MODERATE | tolerated (0.19) | possibly_damaging (0.829) | FE106 | 1 |
| SAMHD1 | 606754 | chr20_36911251_T/C | ENSG00000101347 | ENST00000646673 | missense_variant | 1302 | 1237 | 413 | I/V | Att/Gtt | - | MODERATE | tolerated (0.15) | benign (0.01) | FE106 | 1 |
| SAMHD1 | 606754 | chr20_36911251_T/C | ENSG00000101347 | ENST00000646869 | missense_variant,NMD_transcript_variant | 1308 | 1237 | 413 | I/V | Att/Gtt | - | MODERATE | tolerated (0.15) | benign (0.01) | FE106 | 1 |
| SCFD1 | 618207 | chr14_30694827_A/G | ENSG00000092108 | ENST00000396629 | missense_variant | 1207 | 1021 | 341 | T/A | Act/Gct | rs61754285 | MODERATE | tolerated (0.38) | benign (0.015) | FE164 | 1 |
| SCFD1 | 618207 | chr14_30694827_A/G | ENSG00000092108 | ENST00000458591 | missense_variant | 1317 | 1297 | 433 | T/A | Act/Gct | rs61754285 | MODERATE | tolerated (0.49) | benign (0.015) | FE164 | 1 |
| SCFD1 | 618207 | chr14_30694827_A/G | ENSG00000092108 | ENST00000544052 | missense_variant | 1246 | 1096 | 366 | T/A | Act/Gct | rs61754285 | MODERATE | tolerated (0.45) | benign (0.015) | FE164 | 1 |
| SCFD1 | 618207 | chr14_30630553_T/C | ENSG00000092108 | ENST00000557713 | missense_variant,NMD_transcript_variant | 291 | 134 | 45 | I/T | aTc/aCc | rs61754480 | MODERATE | deleterious (0) | possibly_damaging (0.743) | FE194 | 1 |
| SCFD1 | 618207 | chr14_30630553_T/C | ENSG00000092108 | ENST00000311943 | missense_variant,NMD_transcript_variant | 221 | 209 | 70 | I/T | aTc/aCc | rs61754480 | MODERATE | deleterious (0) | possibly_damaging (0.863) | FE194 | 1 |
| SCFD1 | 618207 | chr14_30630553_T/C | ENSG00000092108 | ENST00000458591 | missense_variant | 229 | 209 | 70 | I/T | aTc/aCc | rs61754480 | MODERATE | deleterious (0) | possibly_damaging (0.768) | FE194 | 1 |
| SCFD1 | 618207 | chr14_30630553_T/C | ENSG00000092108 | ENST00000484733 | missense_variant,NMD_transcript_variant | 158 | 8 | 3 | I/T | aTc/aCc | rs61754480 | MODERATE | deleterious (0) | benign (0.127) | FE194 | 1 |
| SCFD1 | 618207 | chr14_30630553_T/C | ENSG00000092108 | ENST00000544052 | missense_variant | 158 | 8 | 3 | I/T | aTc/aCc | rs61754480 | MODERATE | deleterious (0) | possibly_damaging (0.768) | FE194 | 1 |
| SCFD1 | 618207 | chr14_30630553_T/C | ENSG00000092108 | ENST00000554776 | missense_variant,NMD_transcript_variant | 148 | 149 | 50 | I/T | aTc/aCc | rs61754480 | MODERATE | deleterious (0) | possibly_damaging (0.716) | FE194 | 1 |
| SCFD1 | 618207 | chr14_30630553_T/C | ENSG00000092108 | ENST00000555259 | missense_variant,NMD_transcript_variant | 237 | 209 | 70 | I/T | aTc/aCc | rs61754480 | MODERATE | deleterious (0) | possibly_damaging (0.74) | FE194 | 1 |
| SCFD1 | 618207 | chr14_30630553_T/C | ENSG00000092108 | ENST00000556768 | missense_variant,NMD_transcript_variant | 229 | 209 | 70 | I/T | aTc/aCc | rs61754480 | MODERATE | deleterious (0) | possibly_damaging (0.642) | FE194 | 1 |
| SCFD1 | 618207 | chr14_30630553_T/C | ENSG00000092108 | ENST00000557076 | missense_variant | 283 | 134 | 45 | I/T | aTc/aCc | rs61754480 | MODERATE | deleterious (0) | possibly_damaging (0.768) | FE194 | 1 |
| SCN1A | 182389 | chr2_166015636_G/C | ENSG00000144285 | ENST00000303395 | missense_variant | 3939 | 3521 | 1174 | T/S | aCt/aGt | rs121918799,CM076496 | MODERATE | tolerated (1) | benign (0) | FE136 | 1 |
| SCUBE1 | 611746 | chr22_43339179_T/C | ENSG00000159307 | ENST00000360835 | missense_variant | 256 | 145 | 49 | I/V | Atc/Gtc | rs41307223 | MODERATE | deleterious (0.05) | possibly_damaging (0.491) | FE164 | 1 |
| SDF2L1 | 607551 | chr22_21643991_G/A | ENSG00000128228 | ENST00000248958 | missense_variant | 517 | 482 | 161 | R/H | cGc/cAc | rs73166641 | MODERATE | deleterious (0) | probably_damaging (0.998) | FE130 | 1 |
| SDHB | 185470 | chr1_17027802_A/G | ENSG00000117118 | ENST00000375499 | missense_variant | 500 | 487 | 163 | S/P | Tct/Cct | rs33927012,CM056695 | MODERATE | tolerated (0.96) | benign (0) | FE194 | 1 |
| SDHB | 185470 | chr1_17027802_A/G | ENSG00000117118 | ENST00000463045 | missense_variant | 649 | 316 | 106 | S/P | Tct/Cct | rs33927012,CM056695 | MODERATE | tolerated_low_confidence (1) | benign (0) | FE194 | 1 |
| SDHB | 185470 | chr1_17027802_A/G | ENSG00000117118 | ENST00000491274 | missense_variant | 494 | 445 | 149 | S/P | Tct/Cct | rs33927012,CM056695 | MODERATE | tolerated_low_confidence (1) | possibly_damaging (0.812) | FE194 | 1 |
| SEC16A | 612854 | chr9_136475364_G/C | ENSG00000148396 | ENST00000290037 | missense_variant | 2571 | 2252 | 751 | A/G | gCa/gGa | rs200394508 | MODERATE | tolerated (0.15) | benign (0) | FE106 | 1 |
| SEC16A | 612854 | chr9_136475364_G/C | ENSG00000148396 | ENST00000313050 | missense_variant | 2326 | 2252 | 751 | A/G | gCa/gGa | rs200394508 | MODERATE | tolerated (0.14) | benign (0.003) | FE106 | 1 |
| SEC16A | 612854 | chr9_136475364_G/C | ENSG00000148396 | ENST00000371706 | missense_variant | 1752 | 1718 | 573 | A/G | gCa/gGa | rs200394508 | MODERATE | tolerated (0.15) | benign (0.003) | FE106 | 1 |
| SEC16A | 612854 | chr9_136475364_G/C | ENSG00000148396 | ENST00000431893 | missense_variant | 1718 | 1718 | 573 | A/G | gCa/gGa | rs200394508 | MODERATE | tolerated (0.15) | benign (0.001) | FE106 | 1 |
| SEL1L | 602329 | chr14_81477058_G/T | ENSG00000071537 | ENST00000336735 | missense_variant | 2408 | 2299 | 767 | Q/K | Caa/Aaa | - | MODERATE | tolerated (0.07) | possibly_damaging (0.581) | FE164 | 1 |
| SELENOI | 607915 | chr2_26373468_G/A | ENSG00000138018 | ENST00000260585 | missense_variant | 502 | 412 | 138 | V/M | Gtg/Atg | COSV53146718 | MODERATE | deleterious (0.02) | benign (0.107) | FE193 | 1 |
| SELENOI | 607915 | chr2_26373468_G/A | ENSG00000138018 | ENST00000442141 | missense_variant | 466 | 316 | 106 | V/M | Gtg/Atg | COSV53146718 | MODERATE | tolerated (0.09) | benign (0.107) | FE193 | 1 |
| SELENOI | 607915 | chr2_26373468_G/A | ENSG00000138018 | ENST00000613142 | missense_variant | 559 | 412 | 138 | V/M | Gtg/Atg | COSV53146718 | MODERATE | deleterious (0.03) | benign (0.281) | FE193 | 1 |
| SEMA4C | 604462 | chr2_96865914_G/A | ENSG00000168758 | ENST00000449330 | missense_variant | 694 | 274 | 92 | P/S | Ccc/Tcc | rs141610691 | MODERATE | tolerated (0.81) | benign (0.047) | FE136 | 1 |

| | | | | | | | | | | | | | | | | |
|---|---|---|---|---|---|---|---|---|---|---|---|---|---|---|---|---|
| SEMA4C | 604462 | chr2_96865914_G/A | ENSG00000168758 | ENST00000305476 | missense_variant | 516 | 274 | 92 | P/S | Ccc/Tcc | rs141610691 | MODERATE | tolerated (0.64) | benign (0.047) | FE136 | 1 |
| SEMA4C | 604462 | chr2_96865914_G/A | ENSG00000168758 | ENST00000442264 | missense_variant | 487 | 274 | 92 | P/S | Ccc/Tcc | rs141610691 | MODERATE | tolerated (1) | benign (0.047) | FE136 | 1 |
| SENP2 | 608261 | chr3_185582696_G/A | ENSG00000163904 | ENST00000430355 | missense_variant | 201 | 142 | 48 | V/M | Gtg/Atg | - | MODERATE | deleterious_low_confidence (0.04) | unknown (0) | FE165 | 1 |
| SENP6 | 605003 | chr6_75663290_A/G | ENSG00000112701 | ENST00000424947 | missense_variant | 436 | 436 | 146 | I/V | Att/Gtt | rs145031402 | MODERATE | tolerated (0.12) | benign (0.374) | FE164 | 1 |
| SENP6 | 605003 | chr6_75663290_A/G | ENSG00000112701 | ENST00000447266 | missense_variant | 1411 | 766 | 256 | I/V | Att/Gtt | rs145031402 | MODERATE | deleterious (0.03) | benign (0.374) | FE164 | 1 |
| SENP6 | 605003 | chr6_75663290_A/G | ENSG00000112701 | ENST00000370010 | missense_variant | 1761 | 745 | 249 | I/V | Att/Gtt | rs145031402 | MODERATE | deleterious (0.03) | possibly_damaging (0.578) | FE164 | 1 |
| SENP6 | 605003 | chr6_75663290_A/G | ENSG00000112701 | ENST00000327284 | missense_variant | 1343 | 745 | 249 | I/V | Att/Gtt | rs145031402 | MODERATE | tolerated (0.06) | possibly_damaging (0.578) | FE164 | 1 |
| SENP6 | 605003 | chr6_75663290_A/G | ENSG00000112701 | ENST00000483859 | missense_variant | 789 | 436 | 146 | I/V | Att/Gtt | rs145031402 | MODERATE | tolerated (0.07) | benign (0.164) | FE164 | 1 |
| SGK1 | 602958 | chr6_134262124_T/C | ENSG00000118515 | ENST00000367858 | missense_variant | 692 | 94 | 32 | M/V | Atg/Gtg | rs147657480,CM152696,COSV63116122 | MODERATE | tolerated_low_confidence (0.05) | benign (0) | FE136 | 1 |
| SGK1 | 602958 | chr6_134262124_T/C | ENSG00000118515 | ENST00000367858 | missense_variant | 692 | 94 | 32 | M/V | Atg/Gtg | rs147657480,CM152696,COSV63116122 | MODERATE | tolerated_low_confidence (0.05) | benign (0) | FE187 | 1 |
| SIK2 | 608973 | chr11_111721842_C/G | ENSG00000170145 | ENST00000304987 | missense_variant | 2072 | 1957 | 653 | Q/E | Cag/Gag | rs571853253 | MODERATE | tolerated (0.09) | benign (0.015) | FE193 | 1 |
| SIPA1L2 | 611609 | chr1_232404167_C/G | ENSG00000116991 | ENST00000366630 | missense_variant | 5133 | 4774 | 1592 | D/H | Gat/Cat | rs923838728 | MODERATE | tolerated (0.06) | possibly_damaging (0.765) | FE130 | 1 |
| SIPA1L2 | 611609 | chr1_232514233_C/T | ENSG00000116991 | ENST00000366630 | missense_variant | 1466 | 1107 | 369 | M/I | atG/atA | rs113255944 | MODERATE | tolerated (0.35) | benign (0) | FE154 | 1 |
| SLAIN1 | 610491 | chr13_77746840_A/C | ENSG00000139737 | ENST00000488699 | missense_variant | 794 | 751 | 251 | N/H | Aat/Cat | - | MODERATE | tolerated (0.34) | possibly_damaging (0.773) | FE130 | 1 |
| SLC13A4 | 604309 | chr7_135691293_G/A | ENSG00000164707 | ENST00000354042 | missense_variant | 2041 | 1351 | 451 | P/S | Ccc/Tcc | rs36004833 | MODERATE | tolerated (0.09) | benign (0.095) | FE164 | 1 |
| SLC25A19 | 606521 | chr17_75273251_C/T | ENSG00000125454 | ENST00000582822 | missense_variant | 189 | 191 | 64 | R/H | cGt/cAt | rs62622012 | MODERATE | deleterious (0.02) | possibly_damaging (0.651) | FE106 | 1 |
| SLC33A1 | 603690 | chr3_155829719_T/G | ENSG00000169359 | ENST00000475842 | missense_variant | 609 | 611 | 204 | N/T | aAc/aCc | rs144015992 | MODERATE | tolerated (1) | benign (0.003) | FE190 | 1 |
| SLC33A1 | 603690 | chr3_155829719_T/G | ENSG00000169359 | ENST00000359479 | missense_variant | 1883 | 1451 | 484 | N/T | aAc/aCc | rs144015992 | MODERATE | tolerated (0.75) | benign (0.012) | FE190 | 1 |
| SLC33A1 | 603690 | chr3_155829719_T/G | ENSG00000169359 | ENST00000496772 | missense_variant | 359 | 359 | 120 | N/T | aAc/aCc | rs144015992 | MODERATE | tolerated (0.74) | benign (0.01) | FE190 | 1 |
| SLC33A1 | 603690 | chr3_155829719_T/G | ENSG00000169359 | ENST00000646424 | missense_variant | 1594 | 1145 | 382 | N/T | aAc/aCc | rs144015992 | MODERATE | tolerated (0.77) | benign (0.003) | FE190 | 1 |
| SLC33A1 | 603690 | chr3_155829719_T/G | ENSG00000169359 | ENST00000643144 | missense_variant | 1881 | 1451 | 484 | N/T | aAc/aCc | rs144015992 | MODERATE | tolerated (0.75) | benign (0.012) | FE190 | 1 |
| SLC35A2 | 314375 | chrX_48911594_G/T | ENSG00000102100 | ENST00000247138 | missense_variant | 53 | 43 | 15 | P/T | Cca/Aca | rs55719932,COSV55946578 | MODERATE | tolerated_low_confidence (0.38) | benign (0.014) | FE194 | 1 |
| SLC35A2 | 314375 | chrX_48911594_G/T | ENSG00000102100 | ENST00000376512 | missense_variant | 84 | 43 | 15 | P/T | Cca/Aca | rs55719932,COSV55946578 | MODERATE | tolerated_low_confidence (0.43) | benign (0.014) | FE194 | 1 |
| SLC35A2 | 314375 | chrX_48911594_G/T | ENSG00000102100 | ENST00000376521 | missense_variant | 365 | 43 | 15 | P/T | Cca/Aca | rs55719932,COSV55946578 | MODERATE | tolerated_low_confidence (0.44) | benign (0.026) | FE194 | 1 |
| SLC35A2 | 314375 | chrX_48911594_G/T | ENSG00000102100 | ENST00000376529 | missense_variant | 53 | 43 | 15 | P/T | Cca/Aca | rs55719932,COSV55946578 | MODERATE | tolerated_low_confidence (0.13) | benign (0.014) | FE194 | 1 |
| SLC35A2 | 314375 | chrX_48911594_G/T | ENSG00000102100 | ENST00000445167 | missense_variant | 76 | 43 | 15 | P/T | Cca/Aca | rs55719932,COSV55946578 | MODERATE | deleterious_low_confidence (0.05) | benign (0.055) | FE194 | 1 |
| SLC35A2 | 314375 | chrX_48911594_G/T | ENSG00000102100 | ENST00000452555 | missense_variant | 53 | 43 | 15 | P/T | Cca/Aca | rs55719932,COSV55946578 | MODERATE | tolerated_low_confidence (0.12) | benign (0.007) | FE194 | 1 |
| SLC35A2 | 314375 | chrX_48911594_G/T | ENSG00000102100 | ENST00000616181 | missense_variant | 53 | 43 | 15 | P/T | Cca/Aca | rs55719932,COSV55946578 | MODERATE | tolerated_low_confidence (0.23) | benign (0) | FE194 | 1 |
| SLC35A2 | 314375 | chrX_48911594_G/T | ENSG00000102100 | ENST00000634461 | missense_variant | 76 | 43 | 15 | P/T | Cca/Aca | rs55719932,COSV55946578 | MODERATE | tolerated_low_confidence (0.05) | unknown (0) | FE194 | 1 |
| SLC35A2 | 314375 | chrX_48911594_G/T | ENSG00000102100 | ENST00000634665 | missense_variant | 53 | 43 | 15 | P/T | Cca/Aca | rs55719932,COSV55946578 | MODERATE | tolerated_low_confidence (0.45) | benign (0.031) | FE194 | 1 |
| SLC35A2 | 314375 | chrX_48911594_G/T | ENSG00000102100 | ENST00000635015 | missense_variant | 53 | 43 | 15 | P/T | Cca/Aca | rs55719932,COSV55946578 | MODERATE | tolerated_low_confidence (0.05) | benign (0.025) | FE194 | 1 |
| SLC35A2 | 314375 | chrX_48911594_G/T | ENSG00000102100 | ENST00000635238 | missense_variant | 53 | 43 | 15 | P/T | Cca/Aca | rs55719932,COSV55946578 | MODERATE | tolerated (0.21) | possibly_damaging (0.448) | FE194 | 1 |
| SLC35A2 | 314375 | chrX_48911594_G/T | ENSG00000102100 | ENST00000635285 | missense_variant,NMD_transcript_variant | 58 | 43 | 15 | P/T | Cca/Aca | rs55719932,COSV55946578 | MODERATE | tolerated_low_confidence (0.44) | benign (0.026) | FE194 | 1 |
| SLC35A2 | 314375 | chrX_48911594_G/T | ENSG00000102100 | ENST00000635460 | missense_variant | 41 | 43 | 15 | P/T | Cca/Aca | rs55719932,COSV55946578 | MODERATE | tolerated (0.16) | benign (0.031) | FE194 | 1 |
| SLC35A2 | 314375 | chrX_48911594_G/T | ENSG00000102100 | ENST00000635589 | missense_variant | 53 | 43 | 15 | P/T | Cca/Aca | rs55719932,COSV55946578 | MODERATE | tolerated_low_confidence (0.13) | benign (0.011) | FE194 | 1 |
| SLC35A2 | 314375 | chrX_48911594_G/T | ENSG00000102100 | ENST00000635628 | missense_variant,NMD_transcript_variant | 53 | 43 | 15 | P/T | Cca/Aca | rs55719932,COSV55946578 | MODERATE | deleterious_low_confidence (0) | benign (0.031) | FE194 | 1 |
| SLC35B1 | 610790 | chr17_49707012_G/C | ENSG00000121073 | ENST00000502268 | missense_variant,NMD_transcript_variant | 56 | 56 | 19 | T/S | aCt/aGt | rs142518431 | MODERATE | tolerated_low_confidence (0.37) | benign (0.007) | FE165 | 1 |
| SLC35B1 | 610790 | chr17_49707012_G/C | ENSG00000121073 | ENST00000508520 | missense_variant | 253 | 170 | 57 | T/S | aCt/aGt | rs142518431 | MODERATE | tolerated (0.58) | benign (0.01) | FE165 | 1 |
| SLC35B1 | 610790 | chr17_49707012_G/C | ENSG00000121073 | ENST00000240333 | missense_variant | 292 | 161 | 54 | T/S | aCt/aGt | rs142518431 | MODERATE | tolerated (0.62) | benign (0.003) | FE165 | 1 |
| SLC35B1 | 610790 | chr17_49707012_G/C | ENSG00000121073 | ENST00000508520 | missense_variant | 253 | 170 | 57 | T/S | aCt/aGt | rs142518431 | MODERATE | tolerated (0.58) | benign (0.01) | FE190 | 1 |
| SLC35B1 | 610790 | chr17_49707012_G/C | ENSG00000121073 | ENST00000515850 | missense_variant | 379 | 263 | 88 | T/S | aCt/aGt | rs142518431 | MODERATE | tolerated (0.61) | benign (0.012) | FE190 | 1 |
| SLC35B1 | 610790 | chr17_49707012_G/C | ENSG00000121073 | ENST00000649906 | missense_variant | 279 | 272 | 91 | T/S | aCt/aGt | rs142518431 | MODERATE | tolerated (0.56) | benign (0.006) | FE190 | 1 |

| | | | | | | | | | | | | | | | | |
|---|---|---|---|---|---|---|---|---|---|---|---|---|---|---|---|---|
| SLC35B1 | 610790 | chr17_49707012_G/C | ENSG00000121073 | ENST00000514907 | missense_variant | 139 | 68 | 23 | T/S | aCt/aGt | rs142518431 | MODERATE | tolerated (0.57) | benign (0.006) | FE165 | 1 |
| SLC35B1 | 610790 | chr17_49707012_G/C | ENSG00000121073 | ENST00000514907 | missense_variant | 139 | 68 | 23 | T/S | aCt/aGt | rs142518431 | MODERATE | tolerated (0.57) | benign (0.006) | FE190 | 1 |
| SLC35B1 | 610790 | chr17_49707012_G/C | ENSG00000121073 | ENST00000507773 | missense_variant,NMD_transcript_variant | 418 | 161 | 54 | T/S | aCt/aGt | rs142518431 | MODERATE | tolerated (0.71) | benign (0.033) | FE165 | 1 |
| SLC35B1 | 610790 | chr17_49707012_G/C | ENSG00000121073 | ENST00000502268 | missense_variant,NMD_transcript_variant | 56 | 56 | 19 | T/S | aCt/aGt | rs142518431 | MODERATE | tolerated_low_confidence (0.37) | benign (0.007) | FE190 | 1 |
| SLC35B1 | 610790 | chr17_49707012_G/C | ENSG00000121073 | ENST00000507773 | missense_variant,NMD_transcript_variant | 418 | 161 | 54 | T/S | aCt/aGt | rs142518431 | MODERATE | tolerated (0.71) | benign (0.033) | FE190 | 1 |
| SLC35B1 | 610790 | chr17_49707012_G/C | ENSG00000121073 | ENST00000649906 | missense_variant | 279 | 272 | 91 | T/S | aCt/aGt | rs142518431 | MODERATE | tolerated (0.56) | benign (0.006) | FE165 | 1 |
| SLC35B1 | 610790 | chr17_49707012_G/C | ENSG00000121073 | ENST00000240333 | missense_variant | 292 | 161 | 54 | T/S | aCt/aGt | rs142518431 | MODERATE | tolerated (0.62) | benign (0.003) | FE190 | 1 |
| SLC35B1 | 610790 | chr17_49707012_G/C | ENSG00000121073 | ENST00000515850 | missense_variant | 379 | 263 | 88 | T/S | aCt/aGt | rs142518431 | MODERATE | tolerated (0.61) | benign (0.012) | FE165 | 1 |
| SLC35C1 | 605881 | chr11_45811294_C/T | ENSG00000181830 | ENST00000314134 | missense_variant | 1707 | 1054 | 352 | P/S | Ccc/Tcc | rs145613857 | MODERATE | tolerated (0.45) | benign (0.007) | FE136 | 1 |
| SLC35C1 | 605881 | chr11_45811294_C/T | ENSG00000181830 | ENST00000442528 | missense_variant | 1492 | 1015 | 339 | P/S | Ccc/Tcc | rs145613857 | MODERATE | tolerated (0.49) | benign (0.007) | FE136 | 1 |
| SLC4A4 | 603345 | chr4_71451250_G/A | ENSG00000080493 | ENST00000425175 | missense_variant | 1388 | 1271 | 424 | G/D | gGt/gAt | rs148635969 | MODERATE | tolerated (0.41) | benign (0.356) | FE136 | 1 |
| SLIT2 | 603746 | chr4_20510514_A/G | ENSG00000145147 | ENST00000504154 | missense_variant | 2845 | 934 | 312 | I/V | Atc/Gtc | rs139850475 | MODERATE | deleterious (0.01) | probably_damaging (0.996) | FE136 | 1 |
| SMURF2 | 605532 | chr17_64593460_T/C | ENSG00000108854 | ENST00000262435 | missense_variant | 741 | 314 | 105 | N/S | aAc/aGc | rs80215473 | MODERATE | tolerated (0.83) | benign (0) | FE130 | 1 |
| SMURF2 | 605532 | chr17_64593460_T/C | ENSG00000108854 | ENST00000585301 | missense_variant | 307 | 275 | 92 | N/S | aAc/aGc | rs80215473 | MODERATE | tolerated (0.61) | benign (0) | FE130 | 1 |
| SMURF2 | 605532 | chr17_64661852_C/T | ENSG00000108854 | ENST00000578386 | missense_variant,NMD_transcript_variant | 77 | 29 | 10 | G/E | gGg/gAg | rs866321574 | MODERATE | tolerated (0.11) | benign (0.012) | FE193 | 1 |
| SMURF2 | 605532 | chr17_64661852_C/T | ENSG00000108854 | ENST00000582081 | missense_variant,NMD_transcript_variant | 46 | 29 | 10 | G/E | gGg/gAg | rs866321574 | MODERATE | deleterious_low_confidence (0) | benign (0.048) | FE193 | 1 |
| SMURF2 | 605532 | chr17_64593460_T/C | ENSG00000108854 | ENST00000262435 | missense_variant | 741 | 314 | 105 | N/S | aAc/aGc | rs80215473 | MODERATE | tolerated (0.83) | benign (0) | FE190 | 1 |
| SMURF2 | 605532 | chr17_64593460_T/C | ENSG00000108854 | ENST00000585301 | missense_variant | 307 | 275 | 92 | N/S | aAc/aGc | rs80215473 | MODERATE | tolerated (0.61) | benign (0) | FE190 | 1 |
| SMURF2 | 605532 | chr17_64661852_C/T | ENSG00000108854 | ENST00000262435 | missense_variant | 456 | 29 | 10 | G/E | gGg/gAg | rs866321574 | MODERATE | tolerated (0.05) | benign (0.036) | FE193 | 1 |
| SMURF2 | 605532 | chr17_64661852_C/T | ENSG00000108854 | ENST00000585301 | missense_variant | 61 | 29 | 10 | G/E | gGg/gAg | rs866321574 | MODERATE | deleterious (0) | benign (0.206) | FE193 | 1 |
| SNRPA1 | 603521 | chr15_101293080_C/T | ENSG00000131876 | ENST00000254193 | missense_variant | 245 | 175 | 59 | G/S | Ggt/Agt | rs748156963 | MODERATE | deleterious (0.01) | probably_damaging (0.984) | FE164 | 1 |
| SNRPC | 603522 | chr6_34757881_G/ATCT | ENSG00000124562 | ENST00000374017 | protein_altering_variant | 328 | 41 | 14 | R/HL | cGt/cATCTt | rs67671088,COSV55074695 | MODERATE | - | - | FE106 | 1 |
| SNRPC | 603522 | chr6_34757881_G/ATCT | ENSG00000124562 | ENST00000374017 | protein_altering_variant | 328 | 41 | 14 | R/HL | cGt/cATCTt | rs67671088,COSV55074695 | MODERATE | - | - | FE130 | 1 |
| SNRPC | 603522 | chr6_34757881_G/ATCT | ENSG00000124562 | ENST00000374017 | protein_altering_variant | 328 | 41 | 14 | R/HL | cGt/cATCTt | rs67671088,COSV55074695 | MODERATE | - | - | FE164 | 1 |
| SNRPC | 603522 | chr6_34757881_G/ATCT | ENSG00000124562 | ENST00000374017 | protein_altering_variant | 328 | 41 | 14 | R/HL | cGt/cATCTt | rs67671088,COSV55074695 | MODERATE | - | - | FE190 | 1 |
| SNRPC | 603522 | chr6_34757881_G/ATCT | ENSG00000124562 | ENST00000374017 | protein_altering_variant | 328 | 41 | 14 | R/HL | cGt/cATCTt | rs67671088,COSV55074695 | MODERATE | - | - | FE136 | 2 |
| SNRPC | 603522 | chr6_34757881_G/ATCT | ENSG00000124562 | ENST00000374017 | protein_altering_variant | 328 | 41 | 14 | R/HL | cGt/cATCTt | rs67671088,COSV55074695 | MODERATE | - | - | FE193 | 1 |
| SNRPC | 603522 | chr6_34757881_G/ATCT | ENSG00000124562 | ENST00000374017 | protein_altering_variant | 328 | 41 | 14 | R/HL | cGt/cATCTt | rs67671088,COSV55074695 | MODERATE | - | - | FE154 | 1 |
| SNX13 | 606589 | chr7_17796898_G/A | ENSG00000071189 | ENST00000428135 | missense_variant | 2754 | 2555 | 852 | P/L | cCa/cTa | rs757491303 | MODERATE | tolerated (0.09) | benign (0.015) | FE187 | 1 |
| SORT1 | 602458 | chr1_109323008_C/T | ENSG00000134243 | ENST00000256637 | missense_variant | 1974 | 1948 | 650 | V/M | Gtg/Atg | rs72646577 | MODERATE | deleterious (0.02) | possibly_damaging (0.808) | FE154 | 1 |
| SORT1 | 602458 | chr1_109367478_T/C | ENSG00000134243 | ENST00000256637 | missense_variant | 396 | 370 | 124 | I/V | Att/Gtt | rs61797119,CM145240 | MODERATE | tolerated (0.36) | benign (0.056) | FE190 | 1 |
| SORT1 | 602458 | chr1_109323008_C/T | ENSG00000134243 | ENST00000538502 | missense_variant | 1970 | 1537 | 513 | V/M | Gtg/Atg | rs72646577 | MODERATE | deleterious (0.02) | possibly_damaging (0.519) | FE154 | 1 |
| SOS1 | 182530 | chr2_39013966_G/A | ENSG00000115904 | ENST00000395038 | missense_variant | 1992 | 1964 | 655 | P/L | cCa/cTa | rs56219475,CM070277 | MODERATE | tolerated (0.27) | benign (0.003) | FE194 | 1 |
| SOS1 | 182530 | chr2_39013966_G/A | ENSG00000115904 | ENST00000402219 | missense_variant | 2005 | 1964 | 655 | P/L | cCa/cTa | rs56219475,CM070277 | MODERATE | tolerated (0.3) | benign (0.099) | FE194 | 1 |
| SOS2 | 601247 | chr14_50188589_C/T | ENSG00000100485 | ENST00000216373 | missense_variant | 917 | 622 | 208 | A/T | Gca/Aca | rs61755579 | MODERATE | tolerated (0.45) | benign (0.201) | FE165 | 1 |
| SOX13 | 604748 | chr1_204125859_C/T | ENSG00000143842 | ENST00000367204 | missense_variant,splice_region_variant | 2192 | 1594 | 532 | P/S | Ccg/Tcg | rs34758764 | MODERATE | tolerated (0.25) | benign (0.001) | FE190 | 1 |
| SP1 | 189906 | chr12_53409514_G/A | ENSG00000185591 | ENST00000327443 | missense_variant | 2113 | 1997 | 666 | R/H | cGc/cAc | - | MODERATE | deleterious (0) | probably_damaging (0.991) | FE164 | 1 |
| SP1 | 189906 | chr12_53409514_G/A | ENSG00000185591 | ENST00000426431 | missense_variant | 2036 | 1976 | 659 | R/H | cGc/cAc | - | MODERATE | deleterious (0) | probably_damaging (0.991) | FE164 | 1 |
| SPEG | 615950 | chr2_219444966_G/A | ENSG00000072195 | ENST00000312358 | missense_variant | 755 | 620 | 207 | R/H | cGc/cAc | rs202020343,COSV56663654 | MODERATE | tolerated (0.54) | benign (0) | FE187 | 1 |
| SPI1 | 165170 | chr11_47358804_G/A | ENSG00000066336 | ENST00000533968 | missense_variant | 667 | 533 | 178 | P/L | cCg/cTg | rs146054989 | MODERATE | tolerated_low_confidence (0.12) | benign (0) | FE194 | 1 |
| SRF | 600589 | chr6_43172050_A/T | ENSG00000112658 | ENST00000265354 | missense_variant | 782 | 394 | 132 | S/C | Agc/Tgc | - | MODERATE | deleterious (0.04) | possibly_damaging (0.719) | FE130 | 1 |
| SRGAP1 | 606523 | chr12_64043534_G/A | ENSG00000196935 | ENST00000355086 | missense_variant | 877 | 760 | 254 | V/I | Gtt/Att | - | MODERATE | tolerated (0.29) | benign (0.007) | FE164 | 1 |

| | | | | | | | | | | | | | | | | |
|---|---|---|---|---|---|---|---|---|---|---|---|---|---|---|---|---|
| SRRM1 | 605975 | chr1_24651436_A/T | ENSG00000133226 | ENST00000323848 | missense_variant | 574 | 549 | 183 | R/S | agA/agT | - | MODERATE | deleterious_low_confidence (0.04) | probably_damaging (0.95) | FE190 | 1 |
| SRRM1 | 605975 | chr1_24670297_G/A | ENSG00000133226 | ENST00000447431 | missense_variant | 2382 | 2377 | 793 | E/K | Gag/Aag | rs373063309 | MODERATE | - | unknown (0) | FE164 | 1 |
| SRRM1 | 605975 | chr1_24651436_A/T | ENSG00000133226 | ENST00000447431 | missense_variant | 554 | 549 | 183 | R/S | agA/agT | - | MODERATE | deleterious_low_confidence (0.01) | probably_damaging (0.978) | FE190 | 1 |
| SRRM1 | 605975 | chr1_24651436_A/T | ENSG00000133226 | ENST00000596378 | missense_variant | 702 | 432 | 144 | R/S | agA/agT | - | MODERATE | deleterious_low_confidence (0) | probably_damaging (0.978) | FE190 | 1 |
| SRRM1 | 605975 | chr1_24651436_A/T | ENSG00000133226 | ENST00000374389 | missense_variant | 577 | 549 | 183 | R/S | agA/agT | - | MODERATE | deleterious_low_confidence (0.01) | probably_damaging (0.95) | FE190 | 1 |
| ST7 | 600833 | chr7_117222922_G/T | ENSG00000004866 | ENST00000265437 | missense_variant | 1914 | 1700 | 567 | R/L | cGt/cTt | rs35196356 | MODERATE | deleterious_low_confidence (0.02) | benign (0) | FE130 | 1 |
| ST7 | 600833 | chr7_117222922_G/T | ENSG00000004866 | ENST00000265437 | missense_variant | 1914 | 1700 | 567 | R/L | cGt/cTt | rs35196356 | MODERATE | deleterious_low_confidence (0.02) | benign (0) | FE164 | 1 |
| STAG2 | 300826 | chrX_123961437_T/G | ENSG00000101972 | ENST00000371160 | splice_donor_variant | - | - | - | - | - | rs913664484 | HIGH | - | - | FE130 | 2 |
| STAG2 | 300826 | chrX_123961437_T/G | ENSG00000101972 | ENST00000435103 | splice_donor_variant | - | - | - | - | - | rs913664484 | HIGH | - | - | FE130 | 2 |
| STRA6 | 610745 | chr15_74180851_C/T | ENSG00000137868 | ENST00000323940 | missense_variant | 2017 | 1771 | 591 | A/T | Gcg/Acg | rs115331762 | MODERATE | tolerated (0.65) | benign (0.003) | FE154 | 1 |
| STRA6 | 610745 | chr15_74180851_C/T | ENSG00000137868 | ENST00000395105 | missense_variant | 1861 | 1771 | 591 | A/T | Gcg/Acg | rs115331762 | MODERATE | tolerated (0.65) | benign (0.003) | FE154 | 1 |
| STRA6 | 610745 | chr15_74180851_C/T | ENSG00000137868 | ENST00000416286 | missense_variant | 1903 | 1747 | 583 | A/T | Gcg/Acg | rs115331762 | MODERATE | tolerated (0.65) | benign (0.001) | FE154 | 1 |
| STRA6 | 610745 | chr15_74180851_C/T | ENSG00000137868 | ENST00000423167 | missense_variant | 1929 | 1744 | 582 | A/T | Gcg/Acg | rs115331762 | MODERATE | tolerated (0.58) | benign (0.003) | FE154 | 1 |
| STRA6 | 610745 | chr15_74180851_C/T | ENSG00000137868 | ENST00000449139 | missense_variant | 1885 | 1771 | 591 | A/T | Gcg/Acg | rs115331762 | MODERATE | tolerated (0.65) | benign (0.003) | FE154 | 1 |
| STRA6 | 610745 | chr15_74180851_C/T | ENSG00000137868 | ENST00000535552 | missense_variant | 2133 | 1882 | 628 | A/T | Gcg/Acg | rs115331762 | MODERATE | tolerated (0.51) | benign (0.007) | FE154 | 1 |
| STRA6 | 610745 | chr15_74180851_C/T | ENSG00000137868 | ENST00000563965 | missense_variant | 2227 | 1888 | 630 | A/T | Gcg/Acg | rs115331762 | MODERATE | tolerated (0.48) | benign (0.01) | FE154 | 1 |
| STRA6 | 610745 | chr15_74180851_C/T | ENSG00000137868 | ENST00000572785 | missense_variant | 626 | 628 | 210 | A/T | Gcg/Acg | rs115331762 | MODERATE | tolerated (0.46) | benign (0.001) | FE154 | 1 |
| STRA6 | 610745 | chr15_74180851_C/T | ENSG00000137868 | ENST00000574278 | missense_variant | 2144 | 1816 | 606 | A/T | Gcg/Acg | rs115331762 | MODERATE | tolerated (0.56) | benign (0.003) | FE154 | 1 |
| STRA6 | 610745 | chr15_74180851_C/T | ENSG00000137868 | ENST00000616000 | missense_variant | 1999 | 1771 | 591 | A/T | Gcg/Acg | rs115331762 | MODERATE | tolerated (0.65) | benign (0.003) | FE154 | 1 |
| SULF2 | 610013 | chr20_47676579_T/C | ENSG00000196562 | ENST00000359930 | missense_variant | 2147 | 1295 | 432 | E/G | gAg/gGg | - | MODERATE | deleterious (0.02) | probably_damaging (0.999) | FE136 | 1 |
| SULF2 | 610013 | chr20_47676579_T/C | ENSG00000196562 | ENST00000467815 | missense_variant | 1902 | 1295 | 432 | E/G | gAg/gGg | - | MODERATE | deleterious (0.02) | probably_damaging (0.969) | FE136 | 1 |
| SULF2 | 610013 | chr20_47676579_T/C | ENSG00000196562 | ENST00000484875 | missense_variant | 1964 | 1295 | 432 | E/G | gAg/gGg | - | MODERATE | deleterious (0.02) | probably_damaging (0.999) | FE136 | 1 |
| SURF6 | 185642 | chr9_133332023_G/A | ENSG00000148296 | ENST00000372022 | missense_variant | 988 | 932 | 311 | T/M | aCg/aTg | rs1800867 | MODERATE | deleterious (0) | probably_damaging (0.938) | FE193 | 1 |
| SUZ12 | 606245 | chr17_31983078_A/T | ENSG00000178691 | ENST00000322652 | missense_variant | 1237 | 997 | 333 | T/S | Aca/Tca | - | MODERATE | tolerated (0.41) | benign (0.007) | FE164 | 1 |
| SUZ12 | 606245 | chr17_31983078_A/T | ENSG00000178691 | ENST00000580398 | missense_variant | 1108 | 928 | 310 | T/S | Aca/Tca | - | MODERATE | tolerated (0.48) | benign (0.163) | FE164 | 1 |
| SYDE1 | 617377 | chr19_15113953_T/C | ENSG00000105137 | ENST00000342784 | missense_variant | 2231 | 2198 | 733 | V/A | gTg/gCg | - | MODERATE | deleterious (0) | possibly_damaging (0.872) | FE190 | 1 |
| TAF6L | 602946 | chr11_62775854_C/T | ENSG00000162227 | ENST00000525405 | missense_variant,NMD_transcript_variant | 208 | 71 | 24 | T/M | aCg/aTg | rs76769410 | MODERATE | tolerated_low_confidence (0.1) | benign (0.025) | FE164 | 1 |
| TAF6L | 602946 | chr11_62775854_C/T | ENSG00000162227 | ENST00000526261 | missense_variant | 339 | 71 | 24 | T/M | aCg/aTg | rs76769410 | MODERATE | tolerated_low_confidence (0.28) | benign (0.017) | FE164 | 1 |
| TAF6L | 602946 | chr11_62775854_C/T | ENSG00000162227 | ENST00000294168 | missense_variant | 179 | 71 | 24 | T/M | aCg/aTg | rs76769410 | MODERATE | tolerated (0.16) | benign (0.017) | FE164 | 1 |
| TAF6L | 602946 | chr11_62775926_C/T | ENSG00000162227 | ENST00000294168 | missense_variant | 251 | 143 | 48 | T/M | aCg/aTg | rs144825291 | MODERATE | tolerated (0.08) | benign (0.336) | FE165 | 1 |
| TAF6L | 602946 | chr11_62775854_C/T | ENSG00000162227 | ENST00000529509 | missense_variant | 163 | 71 | 24 | T/M | aCg/aTg | rs76769410 | MODERATE | tolerated (0.15) | benign (0.017) | FE164 | 1 |
| TAF6L | 602946 | chr11_62775926_C/T | ENSG00000162227 | ENST00000529509 | missense_variant | 235 | 143 | 48 | T/M | aCg/aTg | rs144825291 | MODERATE | tolerated (0.1) | benign (0.336) | FE165 | 1 |
| TAF6L | 602946 | chr11_62775926_C/T | ENSG00000162227 | ENST00000525405 | missense_variant,NMD_transcript_variant | 280 | 143 | 48 | T/M | aCg/aTg | rs144825291 | MODERATE | tolerated_low_confidence (0.06) | probably_damaging (0.982) | FE165 | 1 |
| TAF6L | 602946 | chr11_62775926_C/T | ENSG00000162227 | ENST00000526261 | missense_variant | 411 | 143 | 48 | T/M | aCg/aTg | rs144825291 | MODERATE | tolerated_low_confidence (0.1) | benign (0.336) | FE165 | 1 |
| TBC1D22A | 616879 | chr22_46793586_A/C | ENSG00000054611 | ENST00000337137 | missense_variant | 342 | 205 | 69 | S/R | Agc/Cgc | - | MODERATE | deleterious (0.01) | probably_damaging (0.957) | FE187 | 1 |
| TBX2 | 600747 | chr17_61408397_C/T | ENSG00000121068 | ENST00000240328 | missense_variant | 2364 | 2030 | 677 | P/L | cCc/cTc | rs61751978,COSV53599330 | MODERATE | tolerated (0.06) | possibly_damaging (0.879) | FE106 | 1 |
| TFPI | 152310 | chr2_187466977_C/T | ENSG00000003436 | ENST00000233156 | missense_variant | 1112 | 874 | 292 | V/M | Gtg/Atg | rs5940,CM991170,COSV51903633 | MODERATE | deleterious (0.03) | benign (0.142) | FE165 | 1 |
| TFPI | 152310 | chr2_187466977_C/T | ENSG00000003436 | ENST00000392365 | missense_variant | 902 | 874 | 292 | V/M | Gtg/Atg | rs5940,CM991170,COSV51903633 | MODERATE | deleterious (0.03) | benign (0.142) | FE165 | 1 |
| TGFB2 | 190220 | chr1_218346973_G/A | ENSG00000092969 | ENST00000366929 | missense_variant | 739 | 272 | 91 | R/H | cGc/cAc | rs10482721,CM157997 | MODERATE | deleterious (0.02) | possibly_damaging (0.855) | FE136 | 1 |
| TGFB2 | 190220 | chr1_218346973_G/A | ENSG00000092969 | ENST00000366930 | missense_variant | 1638 | 272 | 91 | R/H | cGc/cAc | rs10482721,CM157997 | MODERATE | deleterious (0.02) | possibly_damaging (0.767) | FE136 | 1 |
| THOC5 | 612733 | chr22_29543425_C/T | ENSG00000100296 | ENST00000358079 | missense_variant,NMD_transcript_variant | 368 | 358 | 120 | V/I | Gta/Ata | rs79004872 | MODERATE | tolerated_low_confidence (0.58) | unknown (0) | FE194 | 1 |
| THOC5 | 612733 | chr22_29544567_G/C | ENSG00000100296 | ENST00000455450 | missense_variant | 121 | 70 | 24 | L/V | Ctg/Gtg | rs61740613 | MODERATE | tolerated (0.28) | benign (0.029) | FE194 | 1 |

| | | | | | | | | | | | | | | | | |
|---|---|---|---|---|---|---|---|---|---|---|---|---|---|---|---|---|
| THOC5 | 612733 | chr22_29544567_G/C | ENSG00000100296 | ENST00000490103 | missense_variant | 256 | 133 | 45 | L/V | Ctg/Gtg | rs61740613 | MODERATE | tolerated (0.28) | benign (0) | FE194 | 1 |
| THOC5 | 612733 | chr22_29544567_G/C | ENSG00000100296 | ENST00000428374 | missense_variant | 377 | 133 | 45 | L/V | Ctg/Gtg | rs61740613 | MODERATE | tolerated (0.28) | benign (0) | FE194 | 1 |
| THOC5 | 612733 | chr22_29544567_G/C | ENSG00000100296 | ENST00000442555 | missense_variant,NMD_transcript_variant | 844 | 133 | 45 | L/V | Ctg/Gtg | rs61740613 | MODERATE | tolerated (0.28) | benign (0.006) | FE194 | 1 |
| THOC5 | 612733 | chr22_29544567_G/C | ENSG00000100296 | ENST00000397873 | missense_variant | 356 | 133 | 45 | L/V | Ctg/Gtg | rs61740613 | MODERATE | tolerated (0.28) | benign (0) | FE194 | 1 |
| THOC5 | 612733 | chr22_29544567_G/C | ENSG00000100296 | ENST00000418021 | missense_variant | 591 | 133 | 45 | L/V | Ctg/Gtg | rs61740613 | MODERATE | tolerated (0.52) | benign (0) | FE194 | 1 |
| THOC5 | 612733 | chr22_29544567_G/C | ENSG00000100296 | ENST00000358079 | missense_variant,NMD_transcript_variant | 143 | 133 | 45 | L/V | Ctg/Gtg | rs61740613 | MODERATE | tolerated (0.28) | benign (0.007) | FE194 | 1 |
| THOC5 | 612733 | chr22_29544567_G/C | ENSG00000100296 | ENST00000440771 | missense_variant | 468 | 133 | 45 | L/V | Ctg/Gtg | rs61740613 | MODERATE | tolerated (0.28) | benign (0) | FE194 | 1 |
| THOC5 | 612733 | chr22_29544567_G/C | ENSG00000100296 | ENST00000397872 | missense_variant | 468 | 133 | 45 | L/V | Ctg/Gtg | rs61740613 | MODERATE | tolerated (0.28) | benign (0) | FE194 | 1 |
| THOC5 | 612733 | chr22_29544567_G/C | ENSG00000100296 | ENST00000397871 | missense_variant | 352 | 133 | 45 | L/V | Ctg/Gtg | rs61740613 | MODERATE | tolerated (0.28) | benign (0) | FE194 | 1 |
| TLE4 | 605132 | chr9_79652696_G/C | ENSG00000106829 | ENST00000376552 | missense_variant | 1512 | 494 | 165 | S/T | aGc/aCc | rs41307447 | MODERATE | tolerated (0.18) | benign (0.003) | FE136 | 1 |
| TMEM131L | 616243 | chr4_153550099_A/G | ENSG00000121210 | ENST00000409959 | missense_variant | 304 | 266 | 89 | K/R | aAa/aGa | rs1303654700 | MODERATE | tolerated (0.6) | benign (0.006) | FE154 | 1 |
| TMEM183A | NA | chr1_203007494_G/T | ENSG00000163444 | ENST00000367242 | missense_variant | 121 | 29 | 10 | R/M | aGg/aTg | rs11558253 | MODERATE | tolerated_low_confidence (0.08) | benign (0.173) | FE130 | 1 |
| TMEM183A | NA | chr1_203007494_G/T | ENSG00000163444 | ENST00000367242 | missense_variant | 121 | 29 | 10 | R/M | aGg/aTg | rs11558253 | MODERATE | tolerated_low_confidence (0.08) | benign (0.173) | FE136 | 1 |
| TNS1 | 600076 | chr2_217848746_G/A | ENSG00000079308 | ENST00000171887 | missense_variant | 1849 | 1396 | 466 | R/C | Cgc/Tgc | rs3815849 | MODERATE | tolerated (0.19) | benign (0.326) | FE106 | 1 |
| TNS1 | 600076 | chr2_217848746_G/A | ENSG00000079308 | ENST00000171887 | missense_variant | 1849 | 1396 | 466 | R/C | Cgc/Tgc | rs3815849 | MODERATE | tolerated (0.19) | benign (0.326) | FE190 | 1 |
| TNS1 | 600076 | chr2_217847726_T/G | ENSG00000079308 | ENST00000171887 | missense_variant | 2869 | 2416 | 806 | N/H | Aac/Cac | rs141901890 | MODERATE | tolerated (0.13) | benign (0.121) | FE190 | 1 |
| TOMM40 | 608061 | chr19_44891562_T/G | ENSG00000130204 | ENST00000252487 | missense_variant | 248 | 147 | 49 | S/R | agT/agG | rs11556510 | MODERATE | tolerated_low_confidence (0.49) | benign (0) | FE165 | 1 |
| TOMM40 | 608061 | chr19_44891562_T/G | ENSG00000130204 | ENST00000405636 | missense_variant | 251 | 147 | 49 | S/R | agT/agG | rs11556510 | MODERATE | tolerated_low_confidence (0.49) | benign (0) | FE165 | 1 |
| TOMM40 | 608061 | chr19_44891562_T/G | ENSG00000130204 | ENST00000426677 | missense_variant | 309 | 147 | 49 | S/R | agT/agG | rs11556510 | MODERATE | tolerated_low_confidence (0.49) | benign (0) | FE165 | 1 |
| TOMM40 | 608061 | chr19_44891562_T/G | ENSG00000130204 | ENST00000589649 | missense_variant | 234 | 147 | 49 | S/R | agT/agG | rs11556510 | MODERATE | tolerated (0.53) | benign (0) | FE165 | 1 |
| TOMM40 | 608061 | chr19_44891562_T/G | ENSG00000130204 | ENST00000592434 | missense_variant | 240 | 147 | 49 | S/R | agT/agG | rs11556510 | MODERATE | tolerated_low_confidence (0.44) | benign (0) | FE165 | 1 |
| TRAPPC11 | 614138 | chr4_183664012_G/C | ENSG00000168538 | ENST00000334690 | missense_variant | 321 | 145 | 49 | V/L | Gta/Cta | rs141909783 | MODERATE | tolerated (0.1) | benign (0.056) | FE136 | 1 |
| TRAPPC11 | 614138 | chr4_183664012_G/C | ENSG00000168538 | ENST00000357207 | missense_variant | 347 | 145 | 49 | V/L | Gta/Cta | rs141909783 | MODERATE | tolerated (0.11) | benign (0.12) | FE136 | 1 |
| TRAPPC11 | 614138 | chr4_183664012_G/C | ENSG00000168538 | ENST00000505676 | missense_variant,NMD_transcript_variant | 238 | 145 | 49 | V/L | Gta/Cta | rs141909783 | MODERATE | tolerated_low_confidence (0.46) | probably_damaging (0.98) | FE136 | 1 |
| TRAPPC11 | 614138 | chr4_183679401_C/T | ENSG00000168538 | ENST00000334690 | stop_gained | 1056 | 880 | 294 | Q/* | Cag/Tag | - | HIGH | - | - | FE154 | 1 |
| TRAPPC11 | 614138 | chr4_183679401_C/T | ENSG00000168538 | ENST00000357207 | stop_gained | 1082 | 880 | 294 | Q/* | Cag/Tag | - | HIGH | - | - | FE154 | 1 |
| TRIM33 | 605769 | chr1_114510883_C/G | ENSG00000197323 | ENST00000358465 | missense_variant | 321 | 194 | 65 | G/A | gGg/gCg | rs956535181 | MODERATE | tolerated_low_confidence (0.07) | benign (0) | FE190 | 1 |
| TRIM46 | 600986 | chr1_155176044_G/A | ENSG00000163462 | ENST00000334634 | missense_variant | 600 | 482 | 161 | R/H | cGc/cAc | rs80254867 | MODERATE | deleterious (0) | probably_damaging (0.99) | FE165 | 1 |
| TSG101 | 601387 | chr11_18506904_C/T | ENSG00000074319 | ENST00000536719 | missense_variant | 636 | 501 | 167 | M/I | atG/atA | rs34385327 | MODERATE | tolerated (0.43) | benign (0) | FE154 | 1 |
| TSG101 | 601387 | chr11_18506904_C/T | ENSG00000074319 | ENST00000251968 | missense_variant | 627 | 501 | 167 | M/I | atG/atA | rs34385327 | MODERATE | tolerated (0.43) | benign (0.01) | FE154 | 1 |
| TTF1 | 600777 | chr9_132402137_G/A | ENSG00000125482 | ENST00000334270 | missense_variant | 743 | 685 | 229 | R/W | Cgg/Tgg | rs139662903 | MODERATE | deleterious_low_confidence (0.01) | benign (0.001) | FE193 | 1 |
| TTI2 | 614426 | chr8_33512496_G/A | ENSG00000129696 | ENST00000431156 | missense_variant | 271 | 118 | 40 | P/S | Ccg/Tcg | rs78781527 | MODERATE | tolerated (0.26) | benign (0.018) | FE165 | 1 |
| TTI2 | 614426 | chr8_33512496_G/A | ENSG00000129696 | ENST00000360742 | missense_variant | 614 | 118 | 40 | P/S | Ccg/Tcg | rs78781527 | MODERATE | tolerated (0.26) | benign (0.018) | FE165 | 1 |
| TTI2 | 614426 | chr8_33512496_G/A | ENSG00000129696 | ENST00000520636 | missense_variant | 217 | 118 | 40 | P/S | Ccg/Tcg | rs78781527 | MODERATE | tolerated (0.26) | benign (0.018) | FE165 | 1 |
| TTI2 | 614426 | chr8_33512496_G/A | ENSG00000129696 | ENST00000613904 | missense_variant | 443 | 118 | 40 | P/S | Ccg/Tcg | rs78781527 | MODERATE | tolerated (0.26) | benign (0.018) | FE165 | 1 |
| TTI2 | 614426 | chr8_33512496_G/A | ENSG00000129696 | ENST00000523305 | missense_variant | 300 | 118 | 40 | P/S | Ccg/Tcg | rs78781527 | MODERATE | tolerated (0.19) | benign (0.018) | FE165 | 1 |
| TUBGCP4 | 609610 | chr15_43398133_G/A | ENSG00000137822 | ENST00000260383 | missense_variant | 1629 | 1375 | 459 | V/I | Gta/Ata | - | MODERATE | deleterious (0.03) | possibly_damaging (0.531) | FE165 | 1 |
| TUBGCP4 | 609610 | chr15_43398133_G/A | ENSG00000137822 | ENST00000561691 | missense_variant,NMD_transcript_variant | 1131 | 1132 | 378 | V/I | Gta/Ata | - | MODERATE | deleterious (0.03) | probably_damaging (0.996) | FE165 | 1 |
| TUBGCP4 | 609610 | chr15_43398133_G/A | ENSG00000137822 | ENST00000563147 | missense_variant | 4 | 4 | 2 | V/I | Gta/Ata | - | MODERATE | deleterious (0.01) | probably_damaging (0.932) | FE165 | 1 |
| TUBGCP4 | 609610 | chr15_43398133_G/A | ENSG00000137822 | ENST00000564079 | missense_variant | 1626 | 1372 | 458 | V/I | Gta/Ata | - | MODERATE | deleterious (0.03) | possibly_damaging (0.525) | FE165 | 1 |
| TULP4 | NA | chr6_158489711_G/A | ENSG00000130338 | ENST00000367097 | missense_variant | 3162 | 1610 | 537 | S/N | aGc/aAc | rs61742077 | MODERATE | tolerated (0.28) | benign (0.01) | FE193 | 1 |
| TUT7 | 613467 | chr9_86301455_G/A | ENSG00000083223 | ENST00000375963 | missense_variant | 4412 | 4241 | 1414 | T/I | aCa/aTa | rs564674138 | MODERATE | deleterious_low_confidence (0.04) | benign (0.035) | FE187 | 1 |

| | | | | | | | | | | | | | | | | |
|---|---|---|---|---|---|---|---|---|---|---|---|---|---|---|---|---|
| UBE3C | 614454 | chr7_157267726_G/C | ENSG00000009335 | ENST00000348165 | missense_variant | 3570 | 3223 | 1075 | E/Q | Gaa/Caa | rs1330234039,COSV61953054,COSV61952883 | MODERATE | tolerated (0.49) | benign (0.401) | FE165 | 1 |
| UBR3 | 613831 | chr2_170073459_G/A | ENSG00000144357 | ENST00000392632 | missense_variant | 2236 | 2237 | 746 | C/Y | tGc/tAc | - | MODERATE | tolerated (0.91) | probably_damaging (0.996) | FE130 | 1 |
| UBR3 | 613831 | chr2_170073459_G/A | ENSG00000144357 | ENST00000418381 | missense_variant | 5051 | 5051 | 1684 | C/Y | tGc/tAc | - | MODERATE | tolerated (1) | probably_damaging (0.986) | FE130 | 1 |
| UBR3 | 613831 | chr2_170073459_G/A | ENSG00000144357 | ENST00000439681 | missense_variant | 1151 | 1151 | 384 | C/Y | tGc/tAc | - | MODERATE | tolerated (0.95) | probably_damaging (0.986) | FE130 | 1 |
| UBR3 | 613831 | chr2_170073459_G/A | ENSG00000144357 | ENST00000272793 | missense_variant | 5105 | 5051 | 1684 | C/Y | tGc/tAc | - | MODERATE | tolerated (1) | probably_damaging (0.986) | FE130 | 1 |
| UBR5 | 608413 | chr8_102311743_C/A | ENSG00000104517 | ENST00000220959 | missense_variant | 2865 | 2410 | 804 | A/S | Gct/Tct | - | MODERATE | tolerated (0.43) | possibly_damaging (0.835) | FE106 | 1 |
| UBR5 | 608413 | chr8_102311743_C/A | ENSG00000104517 | ENST00000520539 | missense_variant | 2876 | 2410 | 804 | A/S | Gct/Tct | - | MODERATE | tolerated (0.43) | probably_damaging (0.935) | FE106 | 1 |
| UBR5 | 608413 | chr8_102311743_C/A | ENSG00000104517 | ENST00000521922 | missense_variant | 2917 | 2392 | 798 | A/S | Gct/Tct | - | MODERATE | tolerated (0.41) | probably_damaging (0.935) | FE106 | 1 |
| UFC1 | 610554 | chr1_161152992_G/C | ENSG00000143222 | ENST00000467540 | splice_donor_variant,non_coding_transcript_variant | - | - | - | - | - | rs181482336 | HIGH | - | - | FE190 | 1 |
| UMPS | 613891 | chr3_124731522_G/A | ENSG00000114491 | ENST00000460034 | missense_variant,NMD_transcript_variant | 193 | 173 | 58 | R/H | cGt/cAt | rs17843787 | MODERATE | tolerated (0.37) | benign (0.006) | FE193 | 1 |
| UMPS | 613891 | chr3_124731522_G/A | ENSG00000114491 | ENST00000467167 | missense_variant,NMD_transcript_variant | 200 | 173 | 58 | R/H | cGt/cAt | rs17843787 | MODERATE | tolerated (0.37) | benign (0.006) | FE193 | 1 |
| UMPS | 613891 | chr3_124731522_G/A | ENSG00000114491 | ENST00000628619 | missense_variant | 200 | 173 | 58 | R/H | cGt/cAt | rs17843787 | MODERATE | tolerated (0.37) | benign (0.006) | FE193 | 1 |
| URB2 | NA | chr1_229637152_A/C | ENSG00000135763 | ENST00000258243 | missense_variant | 2662 | 2539 | 847 | M/L | Atg/Ctg | rs116500601 | MODERATE | tolerated (0.43) | benign (0) | FE106 | 1 |
| USF2 | 600390 | chr19_35270591_G/A | ENSG00000105698 | ENST00000222305 | missense_variant | 714 | 574 | 192 | A/T | Gct/Act | rs764602073 | MODERATE | tolerated (0.19) | benign (0.074) | FE187 | 1 |
| USF2 | 600390 | chr19_35270591_G/A | ENSG00000105698 | ENST00000343550 | missense_variant | 497 | 373 | 125 | A/T | Gct/Act | rs764602073 | MODERATE | tolerated (0.26) | benign (0.155) | FE187 | 1 |
| USF2 | 600390 | chr19_35270591_G/A | ENSG00000105698 | ENST00000594064 | missense_variant | 568 | 568 | 190 | A/T | Gct/Act | rs764602073 | MODERATE | tolerated (0.15) | benign (0.074) | FE187 | 1 |
| USF2 | 600390 | chr19_35270591_G/A | ENSG00000105698 | ENST00000595068 | missense_variant | 611 | 574 | 192 | A/T | Gct/Act | rs764602073 | MODERATE | tolerated (0.18) | benign (0.155) | FE187 | 1 |
| USF2 | 600390 | chr19_35270591_G/A | ENSG00000105698 | ENST00000596380 | missense_variant | 329 | 208 | 70 | A/T | Gct/Act | rs764602073 | MODERATE | tolerated (0.4) | possibly_damaging (0.898) | FE187 | 1 |
| USF2 | 600390 | chr19_35270591_G/A | ENSG00000105698 | ENST00000597671 | missense_variant,NMD_transcript_variant | 98 | 100 | 34 | A/T | Gct/Act | rs764602073 | MODERATE | tolerated (0.31) | probably_damaging (0.99) | FE187 | 1 |
| USF2 | 600390 | chr19_35270591_G/A | ENSG00000105698 | ENST00000599471 | missense_variant | 142 | 142 | 48 | A/T | Gct/Act | rs764602073 | MODERATE | tolerated (0.23) | benign (0.086) | FE187 | 1 |
| USP25 | 604736 | chr21_15878373_C/G | ENSG00000155313 | ENST00000285679 | missense_variant | 3435 | 3066 | 1022 | F/L | ttC/ttG | rs545115233 | MODERATE | tolerated (1) | benign (0.003) | FE193 | 1 |
| USP37 | NA | chr2_218454943_C/T | ENSG00000135913 | ENST00000418019 | missense_variant | 3225 | 2927 | 976 | R/H | cGt/cAt | rs61752208 | MODERATE | tolerated (0.12) | benign (0.277) | FE136 | 1 |
| USP37 | NA | chr2_218454943_C/T | ENSG00000135913 | ENST00000415516 | missense_variant | 3074 | 2645 | 882 | R/H | cGt/cAt | rs61752208 | MODERATE | tolerated (0.12) | possibly_damaging (0.591) | FE136 | 1 |
| USP37 | NA | chr2_218549837_C/G | ENSG00000135913 | ENST00000418019 | missense_variant | 699 | 401 | 134 | S/T | aGc/aCc | rs150600881 | MODERATE | deleterious (0.05) | benign (0.001) | FE165 | 1 |
| USP37 | NA | chr2_218454943_C/T | ENSG00000135913 | ENST00000454775 | missense_variant | 3344 | 2927 | 976 | R/H | cGt/cAt | rs61752208 | MODERATE | tolerated (0.12) | benign (0.277) | FE136 | 1 |
| USP37 | NA | chr2_218454943_C/T | ENSG00000135913 | ENST00000258399 | missense_variant | 3330 | 2927 | 976 | R/H | cGt/cAt | rs61752208 | MODERATE | tolerated (0.12) | benign (0.277) | FE136 | 1 |
| USP37 | NA | chr2_218549837_C/G | ENSG00000135913 | ENST00000415516 | missense_variant | 614 | 185 | 62 | S/T | aGc/aCc | rs150600881 | MODERATE | deleterious (0.04) | benign (0.003) | FE165 | 1 |
| USP37 | NA | chr2_218549837_C/G | ENSG00000135913 | ENST00000454775 | missense_variant | 818 | 401 | 134 | S/T | aGc/aCc | rs150600881 | MODERATE | deleterious (0.05) | benign (0.001) | FE165 | 1 |
| USP37 | NA | chr2_218549837_C/G | ENSG00000135913 | ENST00000258399 | missense_variant | 804 | 401 | 134 | S/T | aGc/aCc | rs150600881 | MODERATE | deleterious (0.05) | benign (0.001) | FE165 | 1 |
| USP37 | NA | chr2_218549837_C/G | ENSG00000135913 | ENST00000338465 | missense_variant | 779 | 401 | 134 | S/T | aGc/aCc | rs150600881 | MODERATE | tolerated (0.06) | benign (0.023) | FE165 | 1 |
| UTP25 | NA | chr1_209837179_G/A | ENSG00000117597 | ENST00000491415 | missense_variant | 1122 | 1030 | 344 | D/N | Gac/Aac | rs41274840 | MODERATE | deleterious (0.01) | possibly_damaging (0.688) | FE194 | 1 |
| VIRMA | 616447 | chr8_94519082_G/A | ENSG00000164944 | ENST00000421249 | missense_variant | 2498 | 2416 | 806 | P/S | Cct/Tct | rs139266529 | MODERATE | tolerated (0.47) | benign (0.042) | FE193 | 1 |
| VIRMA | 616447 | chr8_94519082_G/A | ENSG00000164944 | ENST00000522263 | missense_variant,NMD_transcript_variant | 475 | 475 | 159 | P/S | Cct/Tct | rs139266529 | MODERATE | tolerated (0.33) | benign (0.042) | FE193 | 1 |
| VIRMA | 616447 | chr8_94519082_G/A | ENSG00000164944 | ENST00000297591 | missense_variant | 2438 | 2416 | 806 | P/S | Cct/Tct | rs139266529 | MODERATE | tolerated (0.21) | benign (0.042) | FE193 | 1 |
| VPS16 | 608550 | chr20_2860549_C/T | ENSG00000215305 | ENST00000380445 | missense_variant | 500 | 470 | 157 | A/V | gCc/gTc | rs142580838 | MODERATE | deleterious (0.04) | benign (0.136) | FE154 | 1 |
| VPS16 | 608550 | chr20_2860549_C/T | ENSG00000215305 | ENST00000380469 | missense_variant | 542 | 470 | 157 | A/V | gCc/gTc | rs142580838 | MODERATE | deleterious (0.02) | benign (0.142) | FE154 | 1 |
| VPS16 | 608550 | chr20_2860549_C/T | ENSG00000215305 | ENST00000417508 | missense_variant | 255 | 116 | 39 | A/V | gCc/gTc | rs142580838 | MODERATE | deleterious (0.01) | benign (0.048) | FE154 | 1 |
| VPS16 | 608550 | chr20_2860549_C/T | ENSG00000215305 | ENST00000453689 | missense_variant | 258 | 116 | 39 | A/V | gCc/gTc | rs142580838 | MODERATE | deleterious (0.01) | possibly_damaging (0.531) | FE154 | 1 |
| WBP11 | 618083 | chr12_14790617_G/A | ENSG00000084463 | ENST00000261167 | missense_variant | 1320 | 1148 | 383 | T/I | aCa/aTa | rs148994715 | MODERATE | tolerated_low_confidence (0.2) | benign (0.003) | FE136 | 1 |
| WDR24 | NA | chr16_686126_C/T | ENSG00000127580 | ENST00000248142 | missense_variant | 1783 | 1783 | 595 | A/T | Gca/Aca | - | MODERATE | tolerated (0.63) | benign (0) | FE136 | 1 |
| WDR24 | NA | chr16_686126_C/T | ENSG00000127580 | ENST00000293883 | missense_variant | 2151 | 1393 | 465 | A/T | Gca/Aca | - | MODERATE | tolerated (0.59) | benign (0) | FE136 | 1 |
| WDR24 | NA | chr16_686126_C/T | ENSG00000127580 | ENST00000647644 | missense_variant | 2222 | 1615 | 539 | A/T | Gca/Aca | - | MODERATE | tolerated (1) | benign (0) | FE136 | 1 |

| | | | | | | | | | | | | | | | | |
|---|---|---|---|---|---|---|---|---|---|---|---|---|---|---|---|---|
| WDR75 | NA | chr2_189448811_A/G | ENSG00000115368 | ENST00000436347 | missense_variant,NMD_transcript_variant | 274 | 227 | 76 | H/R | cAt/cGt | rs190390933 | MODERATE | tolerated_low_confidence (0.06) | unknown (0) | FE165 | 1 |
| WDR75 | NA | chr2_189448811_A/G | ENSG00000115368 | ENST00000631047 | missense_variant | 227 | 227 | 76 | H/R | cAt/cGt | rs190390933 | MODERATE | tolerated_low_confidence (0.06) | unknown (0) | FE165 | 1 |
| XPNPEP3 | 613553 | chr22_40907611_A/G | ENSG00000196236 | ENST00000357137 | missense_variant | 851 | 817 | 273 | S/G | Agt/Ggt | rs138501598 | MODERATE | deleterious (0) | possibly_damaging (0.622) | FE165 | 1 |
| XPOT | 603180 | chr12_64418048_A/T | ENSG00000184575 | ENST00000332707 | missense_variant,splice_region_variant | 690 | 203 | 68 | Y/F | tAc/tTc | - | MODERATE | deleterious (0.01) | possibly_damaging (0.625) | FE165 | 1 |
| XPOT | 603180 | chr12_64418048_A/T | ENSG00000184575 | ENST00000400935 | missense_variant,splice_region_variant | 582 | 203 | 68 | Y/F | tAc/tTc | - | MODERATE | deleterious (0.01) | possibly_damaging (0.625) | FE165 | 1 |
| XRN2 | 608851 | chr20_21346525_G/C | ENSG00000088930 | ENST00000377191 | missense_variant | 1708 | 1640 | 547 | C/S | tGc/tCc | rs150230004 | MODERATE | deleterious (0.04) | probably_damaging (0.999) | FE193 | 1 |
| YIPF5 | 611483 | chr5_144165488_G/A | ENSG00000145817 | ENST00000274496 | missense_variant | 362 | 227 | 76 | A/V | gCt/gTt | rs35429531 | MODERATE | tolerated (0.56) | benign (0.001) | FE130 | 1 |
| YY1 | 600013 | chr14_100239816_G/C | ENSG00000100811 | ENST00000262238 | missense_variant | 673 | 572 | 191 | G/A | gGc/gCc | rs772586727 | MODERATE | tolerated (0.85) | benign (0.005) | FE130 | 1 |
| YY1 | 600013 | chr14_100239816_G/C | ENSG00000100811 | ENST00000553625 | missense_variant | 63 | 65 | 22 | G/A | gGc/gCc | rs772586727 | MODERATE | tolerated (0.77) | benign (0.05) | FE130 | 1 |
| YY1 | 600013 | chr14_100239816_G/C | ENSG00000100811 | ENST00000554804 | missense_variant | 58 | 59 | 20 | G/A | gGc/gCc | rs772586727 | MODERATE | tolerated (0.82) | benign (0.05) | FE130 | 1 |
| ZC3H12A | 610562 | chr1_37475595_G/T | ENSG00000163874 | ENST00000373087 | missense_variant | 187 | 99 | 33 | R/S | agG/agT | rs116208741 | MODERATE | tolerated_low_confidence (0.77) | benign (0.001) | FE165 | 1 |
| ZC3H12A | 610562 | chr1_37475591_C/T | ENSG00000163874 | ENST00000373087 | missense_variant | 183 | 95 | 32 | P/L | cCa/cTa | rs115805535 | MODERATE | deleterious_low_confidence (0.01) | benign (0.001) | FE165 | 1 |
| ZEB1 | 189909 | chr10_31521751_A/G | ENSG00000148516 | ENST00000320985 | missense_variant | 2526 | 2416 | 806 | T/A | Aca/Gca | rs141194628 | MODERATE | tolerated (0.3) | probably_damaging (0.97) | FE190 | 1 |
| ZEB1 | 189909 | chr10_31521751_A/G | ENSG00000148516 | ENST00000446923 | missense_variant | 2759 | 2368 | 790 | T/A | Aca/Gca | rs141194628 | MODERATE | tolerated (0.48) | probably_damaging (0.97) | FE190 | 1 |
| ZEB1 | 189909 | chr10_31521751_A/G | ENSG00000148516 | ENST00000542815 | missense_variant | 2228 | 2215 | 739 | T/A | Aca/Gca | rs141194628 | MODERATE | tolerated (0.48) | benign (0.051) | FE190 | 1 |
| ZEB1 | 189909 | chr10_31521751_A/G | ENSG00000148516 | ENST00000560721 | missense_variant | 2379 | 2356 | 786 | T/A | Aca/Gca | rs141194628 | MODERATE | tolerated (0.28) | benign (0.001) | FE190 | 1 |
| ZEB1 | 189909 | chr10_31521751_A/G | ENSG00000148516 | ENST00000361642 | missense_variant | 2482 | 2419 | 807 | T/A | Aca/Gca | rs141194628 | MODERATE | tolerated (0.3) | probably_damaging (0.97) | FE190 | 1 |
| ZFYVE9 | 603755 | chr1_52238036_A/G | ENSG00000157077 | ENST00000287727 | missense_variant | 1071 | 619 | 207 | M/V | Atg/Gtg | rs138765001,COSV55098873 | MODERATE | tolerated_low_confidence (0.48) | benign (0) | FE193 | 1 |
| ZFYVE9 | 603755 | chr1_52237616_C/T | ENSG00000157077 | ENST00000287727 | missense_variant | 651 | 199 | 67 | P/S | Cca/Tca | rs41313248 | MODERATE | tolerated_low_confidence (0.09) | benign (0.057) | FE165 | 1 |
| ZNF335 | 610827 | chr20_45959287_A/C | ENSG00000198026 | ENST00000322927 | missense_variant | 2299 | 2167 | 723 | F/V | Ttc/Gtc | rs41305805 | MODERATE | deleterious (0.05) | benign (0.158) | FE106 | 1 |
| ZNF638 | 614349 | chr2_71349728_A/G | ENSG00000075292 | ENST00000409544 | missense_variant | 1404 | 774 | 258 | I/M | atA/atG | - | MODERATE | tolerated (0.35) | benign (0.02) | FE165 | 1 |
| ZNF750 | 610226 | chr17_82832031_C/T | ENSG00000141579 | ENST00000269394 | missense_variant | 702 | 424 | 142 | A/T | Gct/Act | rs146491968,CM1110630 | MODERATE | tolerated (0.3) | benign (0.033) | FE136 | 1 |
| ZNHIT6 | NA | chr1_85707948_C/G | ENSG00000117174 | ENST00000370574 | missense_variant | 486 | 337 | 113 | G/R | Ggc/Cgc | - | MODERATE | deleterious_low_confidence (0.01) | possibly_damaging (0.694) | FE190 | 1 |

| Supplementary Table 3 – Overrepresentation tests – Reactome - Pathways | | | | | | |
|---|---|---|---|---|---|---|
| **Reactome_term** | **pathway_id** | **Total genes in the pathway** | **Genes prioritized in embryos** | **Fold Enrichment** | **Raw pvalue** | **FDR** |
| Mitochondrial translation elongation | (R-HSA-5389840) | 88 | 9 | 5.19 | 0.11 | 10.29 |
| Mitochondrial translation termination | (R-HSA-5419276) | 88 | 9 | 5.19 | 0.11 | 8.23 |
| Mitochondrial translation initiation | (R-HSA-5368286) | 88 | 9 | 5.19 | 0.11 | 6.59 |
| Mitochondrial translation | (R-HSA-5368287) | 94 | 9 | 5.26 | 2.55 | 6.39 |
| PPARA activates gene expression | (R-HSA-1989781) | 114 | 10 | 4.45 | 0.15 | 8.11 |
| Regulation of lipid metabolism by PPARalpha | (R-HSA-400206) | 115 | 10 | 4.41 | 2.41 | 6.48 |
| Cell Cycle Checkpoints | (R-HSA-69620) | 270 | 16 | 3.01 | 2.34 | 0.44 |
| Signaling by Rho GTPases | (R-HSA-194315) | 417 | 23 | 2.8 | 1.76E-05 | 3.21 |

**Supplementary Table 4 – Panther enrichment analy**

| Cellular component | Gene ontology | Total gene in the pathway | Genes prioritized in embryos | Fold Enrichment | Raw pvalue | FDR |
|---|---|---|---|---|---|---|
| mitotic spindle pole | (GO:0097431) | 33 | 5 | 8.09 | 13.02 | 7.04 |
| organellar ribosome | (GO:0000313) | 89 | 8 | 4.56 | 9.48 | 6.08 |
| mitochondrial ribosome | (GO:0005761) | 89 | 8 | 4.56 | 9.48 | 5.57 |
| mitotic spindle | (GO:0072686) | 137 | 10 | 3.7 | 9.52 | 5.49 |
| kinetochore | (GO:0000776) | 137 | 10 | 3.7 | 9.52 | 5.39 |
| nuclear speck | (GO:0016607) | 403 | 24 | 3.02 | 3.42E-06 | 5.12 |
| condensed chromosome | (GO:0000793) | 222 | 13 | 3.37 | 11.36 | 6.28 |
| transcription regulator complex | (GO:0005667) | 427 | 25 | 3.37 | 2.85E-06 | 4.32 |
| nuclear body | (GO:0016604) | 796 | 42 | 3.08 | 1.86E-08 | 2.07E-06 |
| mitochondrial membrane | (GO:0031966) | 739 | 31 | 2.13 | 2.16 | 16.15 |
| nucleolus | (GO:0005730) | 945 | 39 | 2.09 | 2.35E-05 | 3.25 |
| mitochondrial envelope | (GO:0005740) | 787 | 32 | 2.06 | 3.38 | 2.31 |
| nucleoplasm | (GO:0005654) | 3991 | 156 | 2.38 | 2.13E-18 | 1.07E-15 |
| envelope | (GO:0031975) | 1238 | 47 | 2.33 | 2.41E-05 | 3.21 |
| organelle envelope | (GO:0031967) | 1238 | 47 | 2.33 | 2.41E-05 | 3.13 |
| nuclear lumen | (GO:0031981) | 4992 | 172 | 2.15 | 2.72E-15 | 6.82E-13 |
| catalytic complex | (GO:1902494) | 1411 | 48 | 2.13 | 0.34 | 0.22 |
| organelle lumen | (GO:0043233) | 6118 | 208 | 2.12 | 4.41E-19 | 8.84E-16 |
| intracellular organelle lumen | (GO:0070013) | 6118 | 208 | 2.12 | 4.41E-19 | 4.42E-16 |
| membrane-enclosed lumen | (GO:0031974) | 6118 | 208 | 2.12 | 4.41E-19 | 2.95E-16 |
| chromosome | (GO:0005694) | 2034 | 67 | 2.07 | 3.85E-05 | 4.57 |
| mitochondrion | (GO:0005739) | 1656 | 54 | 2.05 | 5.27 | 3.39 |
| protein-containing complex | (GO:0032991) | 5563 | 180 | 2.04 | 1.57E-13 | 2.86E-11 |
| cytosol | (GO:0005829) | 5305 | 166 | 1.59 | 5.51E-11 | 8.5E-09 |
| nucleus | (GO:0005634) | 7603 | 222 | 1.48 | 8.92E-13 | 1.49E-10 |
| organelle membrane | (GO:0031090) | 3642 | 105 | 1.46 | 4.7E-05 | 5.49 |
| intracellular non-membrane-bounded organelle | (GO:0043232) | 5304 | 152 | 1.45 | 3.31E-07 | 3.5E-05 |
| non-membrane-bounded organelle | (GO:0043228) | 5307 | 152 | 1.45 | 3.35E-07 | 3.36E-05 |
| intracellular membrane-bounded organelle | (GO:0043231) | 11304 | 308 | 1.38 | 1.02E-17 | 4.09E-15 |
| membrane-bounded organelle | (GO:0043227) | 12791 | 328 | 1.3 | 2.07E-15 | 5.92E-13 |
| intracellular organelle | (GO:0043229) | 13102 | 330 | 1.28 | 3.64E-14 | 8.12E-12 |
| organelle | (GO:0043226) | 13908 | 342 | 1.25 | 1.38E-13 | 2.78E-11 |
| intracellular anatomical structure | (GO:0005622) | 14899 | 366 | 1.25 | 1.87E-17 | 6.24E-15 |
| cytoplasm | (GO:0005737) | 11958 | 293 | 1.24 | 9.95E-09 | 1.17E-06 |
| cellular anatomical entity | (GO:0110165) | 18825 | 402 | 1.08 | 2.78E-09 | 3.99E-07 |
| cellular_component | (GO:0005575) | 18964 | 403 | 1.08 | 7.07E-09 | 9.46E-07 |
